# Screening for breast cancer: A systematic review update to inform the Canadian Task Force on Preventive Health Care guideline

**DOI:** 10.1101/2024.05.29.24308154

**Authors:** Alexandria Bennett, Nicole Shaver, Niyati Vyas, Faris Almoli, Robert Pap, Andrea Douglas, Taddele Kibret, Becky Skidmore, Martin Yaffe, Anna Wilkinson, Jean M. Seely, Julian Little, David Moher

## Abstract

**Objective:** This systematic review update synthesized recent evidence on the benefits and harms of breast cancer screening in women aged ≥ 40 years and aims to inform the Canadian Task Force on Preventive Health Care’s (CTFPHC) guideline update.

**Methods:** We searched Ovid MEDLINE® ALL, Embase Classic+Embase, and Cochrane Central Register of Controlled Trials to update our searches to July 8, 2023. Search results for observational studies were limited to publication dates from 2014 to capture more relevant studies. Screening was performed independently and in duplicate by the review team. To expedite the screening process, machine learning was used to prioritize relevant references. Critical health outcomes, as outlined by the CTFPHC, included breast cancer and all-cause mortality, treatment-related morbidity, and overdiagnosis. Randomized controlled trials (RCTs), non/quasi RCTs, and observational studies were included. Data extraction and quality assessment were performed by one reviewer and verified by another. Risk of bias was assessed using the Cochrane Risk of Bias 2.0 tool for RCTs and the Joanna Brigg’s Institute (JBI) checklists for non-randomized and observational studies. When deemed appropriate, studies were pooled via random-effects models. The overall certainty of the evidence was assessed following GRADE guidance.

**Results:** Three new papers reporting on existing RCT trial data and 26 observational studies were included. No new RCTs were identified in this update. No study reported results by ethnicity, race, proportion of study population with dense breasts, or socioeconomic status.

For breast cancer mortality, RCT data from the prior review reported a significant relative reduction in the risk of breast cancer mortality with screening mammography for a general population of 15% (RR 0.85 95% CI 0.78 to 0.93). In this review update, the breast cancer mortality relative risk reduction based on RCT data remained the same, and absolute effects by age decade over 10 years were 0.27 fewer deaths per 1,000 in those aged 40 to 49; 0.50 fewer deaths per 1,000 in those aged 50 to 59; 0.65 fewer deaths per 1,000 in those aged 60 to 69; and 0.92 fewer deaths per 1,000 in those aged 70 to 74. For observational data, the relative mortality risk reduction ranged from 29% to 62%. Absolute effects from breast cancer mortality over 10 years ranged from 0.79 to 0.94 fewer deaths per 1,000 in those aged 40 to 49; 1.45 to 1.72 fewer deaths per 1,000 in those aged 50 to 59; 1.89 to 2.24 fewer deaths per 1,000 in those aged 60 to 69; and 2.68 to 3.17 fewer deaths per 1,000 in those aged 70 to 74.

For all-cause mortality, RCT data from the prior review reported a non-significant relative reduction in the risk of all-cause mortality of screening mammography for a general population of 1% (RR 0.99, 95% CI 0.98 to 1.00). In this review update, the absolute effects for all-cause mortality over 10 years by age decade were 0.13 fewer deaths per 1,000 in those aged 40 to 49; 0.31 fewer deaths per 1,000 in those aged 50 to 59; 0.71 fewer deaths per 1,000 in those aged 60 to 69; and 1.41 fewer deaths per 1,000 in those aged 70 to 74. No observational data were found for all-cause mortality.

For overdiagnosis, this review update found the absolute effects for RCT data (range of follow-up between 9 and 15 years) to be 1.95 more invasive and in situ cancers per 1,000, or 1 more invasive cancer per 1,000, for those aged 40 to 49 and 1.93 more invasive and in situ cancers per 1,000, or 1.18 more invasive cancers per 1,000, for those aged 50 to 59. A sensitivity analysis removing high risk of bias studies found 1.57 more invasive and in situ cancers, or 0.49 more invasive cancers, per 1,000 for those aged 40 to 49 and 3.95 more invasive and in situ cancers per 1,000, or 2.81 more invasive cancers per 1,000, in those aged 50 to 59. For observational data, one report (follow-up for 13 years) found 0.34 more invasive and in situ cancers per 1,000 in those aged 50 to 69.

Overall, the GRADE certainty of evidence was assessed as low or very low, suggesting that the evidence is very uncertain about the effect of screening for breast cancer on the outcomes evaluated in this review.

**Conclusions:** This systematic review update did not identify any new trials comparing breast cancer screening to no screening. Although 26 new observational studies were identified, the overall quality of evidence remains generally low or very low. Future research initiatives should prioritize studying screening in higher risk populations such as those from different ages, racial or ethnic groups, with dense breasts, or family history.

**Registration:** Protocol available on the Open Science Framework: https://osf.io/xngsu/

## Introduction

Breast cancer remains the most common cancer among those assigned female at birth in Canada, excluding non-melanoma skin cancer.^1^ In 2022, it was projected that for every 100,000 individuals assigned female at birth, there would be approximately 129 new breast cancer cases, and 23 individuals would die from breast cancer.^2^ Among the established risk factors for breast cancer are family history, older age, genetics (e.g., *BRCA1*, *BRCA2* or *PALB2* pathogenic variants), race/ethnicity, breast density, obesity in postmenopausal women, early onset of menarche and lifestyle factors such as delayed childbearing, hormone replacement therapy, and previous chest radiation.^3–6^ Through early detection in asymptomatic women, screening for breast cancer aims to reduce morbidity associated with advanced stages of the disease and breast cancer mortality. The benefits and harms of breast cancer screening have been the subject of intense debate.^7^

Multiple randomized controlled clinical trials (RCTs) conducted between the 1960s and 1990s demonstrated a mortality benefit ranging between 6% and 27%, resulting in the widespread implementation of mammographic screening.^7–10^ In Canada, breast cancer mortality rates have declined steadily following the introduction of organized screening programs and advances in treatment over the same time period.^1,11^ Breast cancer screening detects breast cancers before they are symptomatic, with resulting earlier stage disease at diagnosis, improved mortality and decreased morbidity of treatment.^12^ Despite the demonstrated benefits of screening, harms such as false positives (i.e., additional imaging) and overdiagnosis can be associated with breast cancer screening.^13^ Additional imaging, where subsequent testing reveals no cancer (also referred to as false positives), can cause psychological distress, unnecessary biopsies, and follow-up visits.^14^ Overdiagnosis, where the cancer detected would not have become symptomatic or have led to any harm (including death), may lead to unnecessary invasive treatments.^15,16^

RCTs are considered the gold standard for evaluating the efficacy of interventions and heavily weigh in guideline decision-making due to their rigorous design and methods (e.g., randomization, statistical power). There are limitations to the RCTs evaluating breast cancer screening, as the age of the existing trials does not reflect current screening or treatment practices. The Canadian National Breast Screening Studies (CNBSS) reported an excess of breast cancer mortality in the screening arm in women 40 to 49 years and no benefit of screening in women aged 50 to 59 years, results which were not confirmed in the other RCTs.^17^ Concerns about the CNBSS have been raised about the inclusion of symptomatic patients, potentially biased randomization, as well as the quality of mammography.^18–22^ More recent observational studies provide evidence more reflective of current practice,^23,24^ although have limitations around selection and recall bias.^25^

In 2011 and 2018, the Canadian Task Force on Preventive Health Care (CTFPHC) recommended against routine mammography screening starting at age 40, however, suggested that women may wish to be screened based on their values and preferences; in this circumstance providers should engage in shared decision making.^26^ Currently, nine provinces and territories in Canada have organised screening programs for breast cancer screening which allow for self-referral starting at age 40.^27^ The U.S. Preventive Services Task Force (USPSTF) guidelines recommended screening initiation at age 40 in 2024,^28^ and the UK National Health Services (NHS) offers screening to women starting at age 47.^29^ Australia, Norway,^30^ Finland,^31^ and Denmark^32^ offer mammography screening to women starting at age 50, while Sweden invites women starting at age 40.^33^

Understanding the evidence underpinning screening recommendations in the Canadian healthcare system is critical to updating guidelines and their implementation in public health initiatives. A thorough evaluation of the benefits and harms of breast cancer screening can be used to optimise Canadian breast cancer screening practices and help women and their primary healthcare providers weigh the decision to participate or not to participate in breast cancer screening.

### Objective

This evidence review aims to inform the CTFPHC guideline update with the most recent evidence on the benefits and harms of breast cancer screening. This review will be complimented by an additional evidence review of women’s values and preferences related to screening, and modeling to provide estimates of the dependence of breast cancer outcomes on screening regimens.

We updated the methodology of the 2017 review^34^ by including observational studies and summarizing evidence that focused on screening women at least 40 years of age. This evidence review addressed the following key questions (KQ) as posed by the CTFPHC:

**KQ1.** a) What are the benefits and harms of different mammography-based screening strategies compared to no screening in cisgender women and other adults assigned female at birth aged 40 years and older?
b) Do the benefits and harms of mammography screening differ by population characteristics (e.g., age, breast density, race and ethnicity, socioeconomic status, availability of mammography screening, family history)?

## Methods

We conducted a systematic review to update the evidence review completed in 2017,^34^ broadening the search criteria to capture comparative observational studies. We followed guidance from the Cochrane Handbook,^35^ GRADE working group,^36^ and Chapter 4 from the Task Force Methods manual.^37^ Our review was developed, conducted, and reported using the Preferred Reporting Items for Systematic Reviews and Meta-Analyses (PRISMA) checklist (Appendix 1).^38^ The methods were planned *a priori,* and project materials (e.g., protocol, data extraction forms) are publicly available on Open Science Framework (OSF) (https://osf.io/xngsu/). The full CTFPHC research plan, all KQs posed by the CTFPHC, and deviations from the Task Force Methods Manual are available on OSF.

### Contributors

A full description of the CTFPHC methods for guideline development can be found in the Task Force Methods Manual.^37^ The scope of this systematic review was directed by the CTFPHC. The full research plan was developed collaboratively by the Ottawa Evidence Review and Synthesis Centre and the Canadian Task Force Breast Cancer Working Group (hereafter referred to as the “Working Group”) and approved by the CTFPHC. The Working Group (WG) was composed of five CTFPHC members, the University of Ottawa and Alberta Evidence Review and Synthesis Centres (ERSCs), clinical experts (surgical oncologist, radiation oncologist, radiologist, medical oncologist), patient partners, and members from the Public Health Agency of Canada. The goal of the WG was to direct each step of the overall guideline development process. They assisted with defining the KQs and establishing the eligibility criteria.

The University of Ottawa ERSC was responsible for the conduct of the systematic review (e.g., literature search, study selection, quality assessment, certainty of the evidence evaluation, and the writing of the review). The Ottawa ERSC was supported by an ERSC Advisory Group, consisting of clinical and scientific experts (e.g., a breast radiologist, breast cancer imaging scientist, and family physician/general practitioner oncologist) and a patient partner, to provide expedited guidance related to clinical questions. The patient partner was an individual with lived breast cancer experience and was nominated by one of the clinical experts on the Advisory Group.

The Advisory Group provided interpretation of the systematic review results and the conclusions. The patient partner provided feedback on the comprehensiveness, content and linguistic clarity, and structural cohesiveness of the document. The patient partner was compensated for her contributions (meeting attendance, manuscript drafting and review) following the Canadian Institutes of Health Research guidance.^39^ No compensation for the clinical experts’ time or input was provided.

### Eligibility criteria

The detailed criteria for study inclusion or exclusion are outlined in Table 1, following the PICOT framework with additional information on settings, databases, study designs and languages of interest.^40^ Briefly, the review focused on adults assigned female at birth aged ≥ 40 years and at average or moderately increased risk for breast cancer. For this review, average risk refers to those without factors placing them at higher-than-average risk of cancer (i.e., about 12.5% lifetime risk), whereas women with moderately increased risk (i.e.,12.5 to 20% lifetime risk) will include individuals with an elevated risk of breast cancer (e.g., dense breasts, one first degree relative with history of breast cancer). Studies focusing on only those at high lifetime risk for developing breast cancer (i.e., >20% lifetime risk) were excluded from this review.

**Table 1:** Eligibility criteria on the benefits and harms of screening versus no screening.

|  | Inclusion | Exclusion |
| --- | --- | --- |
| **Population** | Cisgender women and other adults assigned female at birth (including transgender men and nonbinary persons) aged $\geq 40$ years of age and at average or moderately increased risk* for breast cancer<br>Specific populations (using within and between-study data where able, as well different estimations of absolute risks based on assumed baseline risks):<br>• Age (40-44 years, 45-49 years, 50-59 years, 60-69 years, 70-74 years, 75 and older)<br>• Ethnicity and race<br>• Socioeconomic status<br>• Availability of mammography screening<br>• Breast density (e.g., extremely [e.g., BI-RADS category D] vs BI-RADS A-C; other comparisons))<br>• Family history<br>Sensitivity analysis may be conducted with studies enrolling 20% or more participants at high risk for breast cancer. | Studies focusing on adults with high risk of breast cancer (e.g., personal history of breast cancer or high-risk breast lesions; strong family history* of breast or ovarian cancer or significant genetic markers or syndromes [e.g., BRCA1/BRCA2, Li-Fraumeni syndrome]; previously received radiation treatment to the chest [e.g., Hodgkin's]) |
| **Intervention** | Any mammography screening modality (i.e., film or digital mammography [2D mammography], digital breast tomosynthesis [2D synthetic, 3D mammography]) with or without clinical breast examination (CBE)/breast self-examination (BSE):<br>1. Alone<br>2. Digital mammography (2D or 2Ds) with tomosynthesis (3D)<br>3. Digital mammography (2D, 2Ds or 3D) with MRI<br>4. Digital mammography (2D, 2Ds or 3D) with ultrasound<br>5. Digital mammography (2D, 2Ds or 3D) with contrast enhanced mammography<br>Studies need to consider a minimum of 2 rounds of screening. | Breast imaging or clinical examinations conducted for diagnosis or surveillance. Screening strategies that do not include mammography. |
|  | **Subgroups:** screening interval (≤12 months, 13 - 24 months, >24 months); with and without BSE or CBE |  |
| **Comparator** | No screening | N/A |
| **Outcomes** | **Benefits (reductions)**<br>*Critical*<br>1. Breast cancer related mortality<br>2. All-cause mortality<br>3. Treatment-related morbidity, measured by:<br>1. Receipt of radiotherapy (yes/no)<br>2. Receipt of chemotherapy (yes/no)<br>• Subgroup by anthracycline vs no anthracycline<br>3. Type of surgery: complete mastectomy vs partial mastectomy/lumpectomy<br>4. Surgical management of axilla (axial lymph node dissection [ALND] vs sentinel lymph node biopsy [SLNB])<br>4. Stage distribution of breast cancer<br>1. Stage II and higher<br>2. Stage III and higher<br>3. Stage IV/metastatic cancer<br>*Important*<br>5. Breast cancer morbidity (e.g., adverse effects of treatment, physical/functional impairment). Measured using composite scores from different scales<br>**Harms**<br>*Critical*<br>6. Overdiagnoses (We will calculate the number of excess diagnoses from prospective data with at least 10 years of follow up from the time of enrollment over 1,000 persons screened).<br>1. Subgroup by Invasive only vs Including DCIS<br>*Important*<br>7. False-positive rate requiring imaging plus biopsy (cumulative over multiple rounds)<br>8. False-positive rate requiring imaging only or imaging plus biopsy (cumulative over multiple rounds)<br>9. Interval cancers (includes false negatives and clinically detected CAs before next screen or time equivalent)<br>1. Subgroup by Invasive only vs Including DCIS<br>**Benefit or harm**<br>*Critical*<br>10. Health related quality of life (secondary outcome). Measured using composite scores from different scales.<br>*Important*<br>11. Life years gained (or lost) | N/A |
| **Timing** | **Follow-up timing (for mortality outcomes):** subgroup analysis for <10 vs ≥10 years<br>**Publication/exposure date:**<br>Randomized controlled trials (RCTs) s and quasi-randomized trials: No date limit<br>All other nonrandomized studies: Publication date of 2014 and screening initiation date of 2000 to increase their applicability to current technologies and cancer treatments/management. | N/A |
| **Settings** | Primary care or other settings generalizable to primary care, including referrals by primary care providers or organized screening programs.<br>Studies conducted in countries categorized as “Very High” on the Human Development Index (as defined by the United Nations Development Programme).<br>https://hdr.undp.org/data-center/human-development-index#/indicies/HDI | N/A |
| **Databases** | Medline, Embase, Cochrane Library | N/A |
| **Study designs** | **All outcomes**<br>1. RCTs, including cluster RCTs<br>2. Non/quasi-RCTs<br>3. Prospective or retrospective observational studies (cohorts or nested case controls) of large screening cohorts with a concurrent control group (including controlled before-after studies) (i.e., all having exposure data at the individual level and linked with outcomes)<br>4. **If reporting data specific to key demographic groups (i.e., 40-49 and/or 70+ years, race/ethnicity group):** ecological/population-based (e.g., exposure data not at participant level, over multiple years), time trend/series and before-after studies<br>**False-positive outcomes**<br>For false positive rates, we plan to rely on the most recently available Canadian data collated by the Canadian Partnership Against Cancer and from Canadian studies/data sources.<br>Sensitivity analysis for trials: RCT vs quasi, high vs low/unclear risk of bias (ROB), predatory journal or not<br>Subgroups for non-randomized studies: adherence vs intention/offer to screen data | Case series (i.e., all participants have cancer), case reports, diagnostic test accuracy studies, modelling studies, reviews.<br>Outcomes must be evaluated in individuals exposed to screening versus not screening, not comparisons of detection methods that do not capture an individual's longitudinal screening experience (e.g., rates of screen-detected vs. not screen-detected cancer). |
| **Language** | English and French full texts | N/A |

\* For the purposes of this review, average risk refers to those without factors placing them at higher-than-average risk of cancer (i.e. about 12% lifetime risk) risk for breast cancer whereas those with moderately increased risk will include individuals with an elevated risk of breast cancer (e.g., dense breasts, one first degree relative with history of breast cancer). The review will not include studies focusing on those with high risk. Strong family history of breast cancer will be defined as per the CDC.<sup>151</sup>

Eligible screening interventions included film or digital mammography and digital breast tomosynthesis with or without clinical breast examination (CBE). Eligible comparators included a group with no mammography screening offered or groups who did not participate in screening. We included settings associated with or generalizable to primary care, including referrals by primary care providers or organized screening programs. Initially, the eligible screening period for observational studies was after the year 2000 to increase their relevance to current technologies and cancer treatment.

Following discussions with and approval by the WG, we included studies with screening periods overlapping the 2000-year mark to capture the full range of evidence that included screening in the 2000s. All RCTs were included, regardless of date of screening period, as per our eligibility criteria and to build upon our previous 2017 review. We also excluded studies published in languages other than English and French, and where breast imaging or clinical examinations were conducted for diagnosis or surveillance.

Critical outcomes related to the potential benefits of breast cancer screening included reduction in breast cancer-related mortality, all-cause mortality, treatment-related morbidity (receipt of radiotherapy, chemotherapy [subgroup by anthracycline], type of surgery, surgical management of axilla), and stage distribution of breast cancer. Important benefits included a potential reduction in breast cancer morbidity.

Overdiagnosis was considered a critical outcome related to the potential harms of breast cancer screening. Important eligible harms were additional testing (no cancer) (previously called false positives) and interval cancers. Health-related quality of life and life years gained (or lost) were considered as potential benefits or harms.

### Information sources and search strategy

The search strategy was developed and tested through an iterative process by an experienced information specialist (BS) in consultation with the review team (Appendix 2). We adapted our strategy from the 2017 review^41^ and included all study designs in the electronic searches. The MEDLINE strategy was peer-reviewed before execution using the Peer Review of Electronic Search Strategies (PRESS) Checklist (Appendix 3). Using the Ovid platform, we searched Ovid MEDLINE® ALL, Embase Classic+Embase, and Cochrane Central Register of Controlled Trials on July 8, 2023. There were no language restrictions applied to any of the searches, but search results were limited to publication dates from 2014 onwards to capture any observational studies since the last search conducted by the USPSTF in 2016. ^42^ We also searched grey literature sources, submissions by stakeholders, and reference lists. The CTFPHC hosted an online portal during the month of August 2023 open to the public who wished to submit literature that may be relevant to the guideline update (Appendix 4). Grey literature sources are available in Appendix 5.

### Study selection

Literature search results were uploaded to DistillerSR, a reference management software. Prior to screening initiation, a team of reviewers completed a pilot title and abstract screening exercise on a random sample of 50 titles and abstracts. Screening was initiated once reviewer agreement was at least 95% using prespecified inclusion and exclusion criteria for each KQ. Any discrepancies among reviewers were resolved by discussion or consultation with a third reviewer. Any discussed adjustments to the form were tracked. The same pilot process was repeated for full-text screening with 25 randomly selected references.

Screening was performed independently and in duplicate by reviewers using the study eligibility screening forms on DistillerSR (Appendix 6). To expedite the screening process DistillerSR’s machine learning prioritization tool, DAISY, was used to prioritize relevant references based on DistillerSR’s highest remaining score. The remaining unreviewed references (those with a highest remaining score of 0.1 or less) were screened by DistillerSR AI with an additional quality check to ensure accuracy. The selection process is recorded in a PRISMA flow diagram (Figure 1).

**Figure 1:**
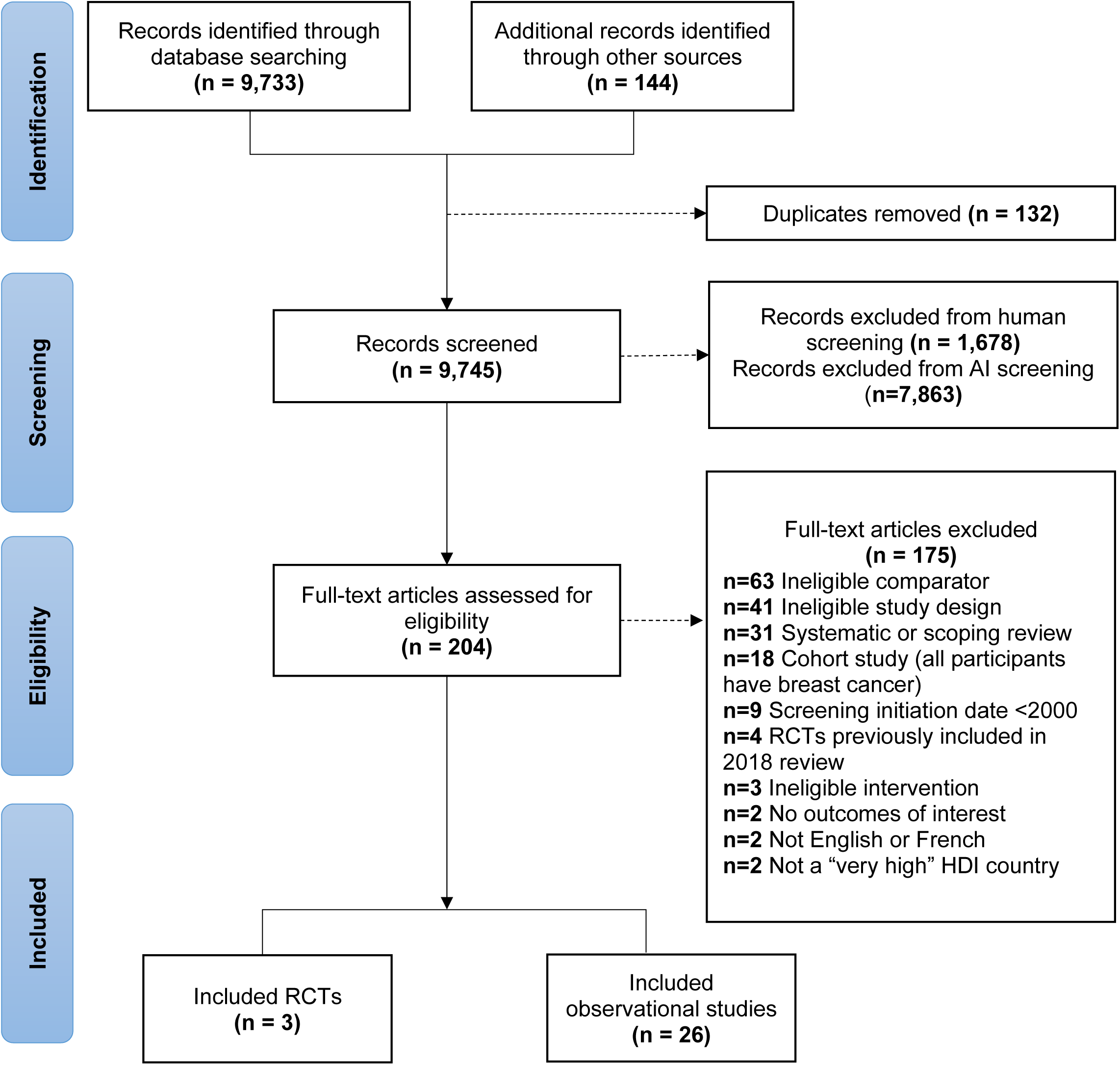
PRISMA flow diagram

### Data extraction

Data extraction was completed in Microsoft Excel by a group of reviewers. Any newly identified studies were assigned to one extractor with a second reviewer to verify. Full extraction forms can be found in Appendix 7 and online on OSF (https://osf.io/xngsu/). Information included details on the study publication, design, and interventions/comparator groups. Relevant outcome data was extracted for each study for both intervention and comparator groups, including sample sizes, adjusted and unadjusted effect measures and reported limitations.

### Risk of bias assessments

Two reviewers completed a risk of bias (RoB) assessment for each included study. Studies were evenly distributed to the two reviewers for assessment followed by validation from the other reviewer. The Cochrane Risk of Bias 2.0 tool was used for RCTs,^43^ and the relevant study design checklist from the JBI tool for each non-randomized and observational studies (e.g., case control, cohort, quasi-experimental).^44^

Consistent with our prior review’s methods, if all Cochrane RoB 2.0 tool domains were ‘low risk’ the overall judgement was low risk, and conversely if at least one important domain was ‘high risk of bias’, then the overall judgement was high risk.

Deficiencies in some domains (e.g., randomization and allocation concealment) were considered to have more serious implications than others (e.g., ‘selective outcome reporting’). We judged trials to be of moderate risk if there were several ‘unclears’ and ‘low risk’, or if a ‘high’ risk was judged in domains which are not considered to have serious implications. For the JBI tools, we evaluated age and hormone replacement therapy use as important variables, in addition to adjustment for self-selection bias. We also evaluated the adjustment for lead time bias and length of follow-up.

Results from the RoB assessments were narratively summarized and presented visually. A scale-based approach was used to tally quality scores for each item (ranging from 0 for unclear/no answers to 1 for items met). JBI does not have a recommended scoring system for their checklists for overall quality. For visualization purposes, observational studies were colour-coded as “high”, “moderate,” or “low” based on tallied scores of 75-100%, 50-75% of items, or below 50% of checklist items sufficiently met.

GRADE risk of bias domain ratings were based not on this coding, but on the specific risk of bias concerns noted for each study. For previously included studies and outcomes, we relied on the prior risk of bias assessments with verification by a single reviewer.^34^ Any changes to the previous risk of bias assessments were documented.

### Data analysis

Study characteristics of included studies are presented in tables and summarized narratively. Risk of bias and methodological quality are also descriptively and visually summarized in tables.

For breast cancer mortality outcomes, the previous systematic review^34^ conducted the main analysis according to both short- and long-case accrual methods. Short-case accrual refers to studies that reported deaths among cases of breast cancer that were diagnosed during the screening intervention period, whereas long-case accrual includes deaths occurring in all cases diagnosed to the end of the follow-up period. In this systematic review update, we present short-case accrual as the primary analysis due to the reduced bias from contamination because women in the control group would not have been screened until the trial was over, while long-case accrual may underestimate the benefits of screening as women in the control group are more likely being screened after the trial. No sensitivity analyses for long-case accrual methods were performed, however results using the long-case accrual are presented in Appendix 8.

When possible, outcomes were presented for a 10-year follow-up period to facilitate decision-making. Some have suggested that 10-15 years after randomization would provide a more reliable estimate of the effect of screening on breast cancer mortality in trials, after which a diluting effect of the control group may occur.^45^ However, the full range of reported follow-up within included studies was reported in the summary of findings tables for each outcome.

#### Summary of Findings

Results were synthesized separately for each study outcome and presented in a GRADE Summary of Findings Table (Appendix 8 and Table 2). Each Summary of Findings Table presents information on the intervention, comparator, number of participants and studies included in the analysis, relative and absolute effects, and the overall certainty of evidence. Following GRADE guidance for indirect calculation of the absolute effect, absolute effects were calculated using the relative effect for each outcome (pooled across included studies, when multiple studies were included for an outcome) and the reported baseline risk in the comparator group (averaged in the case of multiple studies).^46^ Data from The Pan Canadian Study of Mammography Screening (1990-2009)^47^ were used to estimate the baseline risk of breast cancer mortality in an unscreened general Canadian population over 10 years and was 1.8 per 1,000 for 40-49, 3.3 per 1,000 for 50-59, 4.3 per 1,000 for 60-69, and 6.1 per 1,000 for 70-74. We calculated baseline incidence risks from RCT trial data in an unscreened population at 17.7 invasive and in situ cancers (16.7 invasive only) per 1,000 for 40-49, 24.1 invasive and in situ cancers (23.5 invasive only) per 1,000 for 50-59. We did not find any RCT data to determine unscreened incidence for individuals 60-69 or 70-74.

**Table 2.**
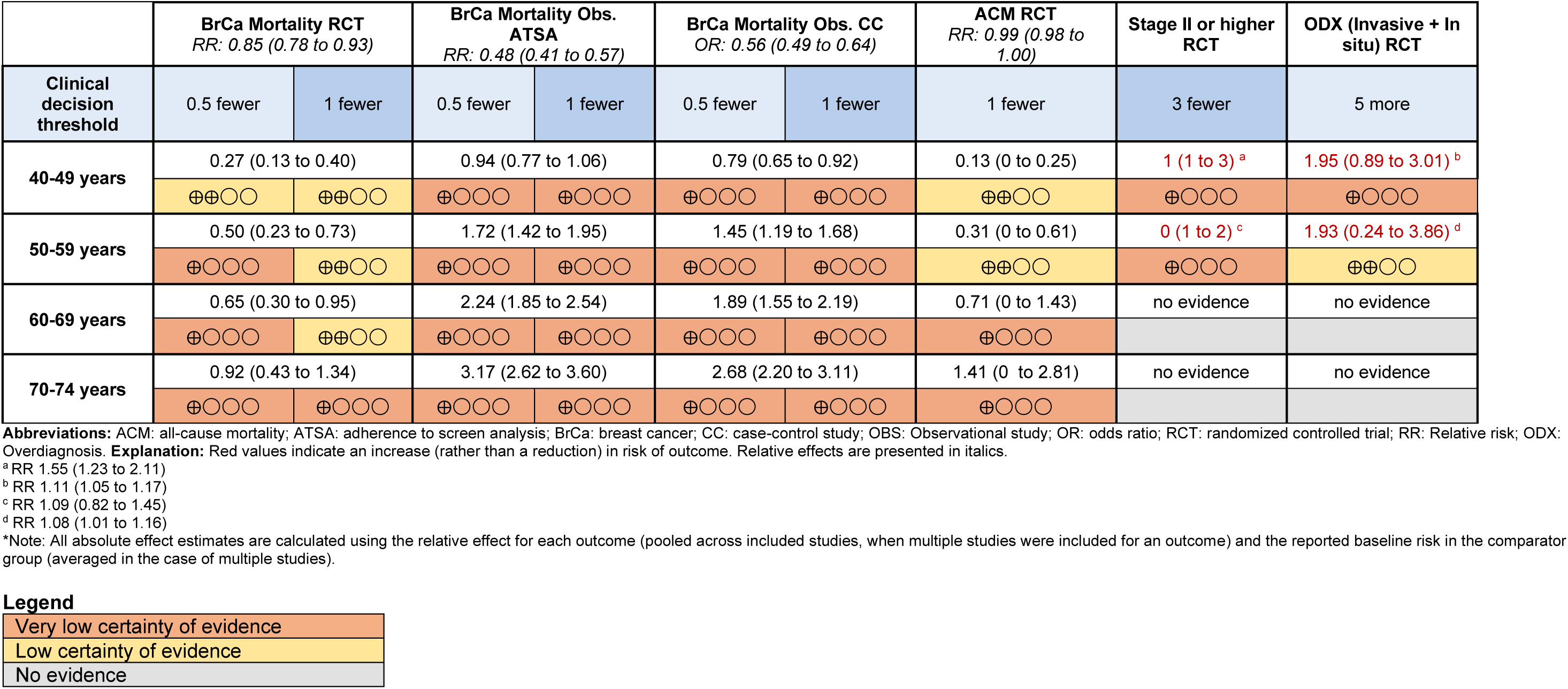
Summary of findings for breast cancer screening by outcome and risk category per 1,000 women over 10 years in a general population.

|  | BrCa Mortality RCT<br>*RR: 0.85 (0.78 to 0.93)* |  | BrCa Mortality Obs.<br>ATSA<br>*RR: 0.48 (0.41 to 0.57)* |  | BrCa Mortality Obs. CC<br>*OR: 0.56 (0.49 to 0.64)* |  | ACM RCT<br>*RR: 0.99 (0.98 to 1.00)* | Stage II or higher<br>RCT | ODX (Invasive + In<br>situ) RCT |
| --- | --- | --- | --- | --- | --- | --- | --- | --- | --- |
| Clinical<br>decision<br>threshold | 0.5 fewer | 1 fewer | 0.5 fewer | 1 fewer | 0.5 fewer | 1 fewer | 1 fewer | 3 fewer | 5 more |
| 40-49 years | 0.27 (0.13 to 0.40) |  | 0.94 (0.77 to 1.06) |  | 0.79 (0.65 to 0.92) |  | 0.13 (0 to 0.25) | 1 (1 to 3) <sup>a</sup> | 1.95 (0.89 to 3.01) <sup>b</sup> |
|  | ⊕⊕○○ | ⊕⊕○○ | ⊕○○○ | ⊕○○○ | ⊕○○○ | ⊕○○○ | ⊕⊕○○ | ⊕○○○ | ⊕○○○ |
| 50-59 years | 0.50 (0.23 to 0.73) |  | 1.72 (1.42 to 1.95) |  | 1.45 (1.19 to 1.68) |  | 0.31 (0 to 0.61) | 0 (1 to 2) <sup>c</sup> | 1.93 (0.24 to 3.86) <sup>d</sup> |
|  | ⊕○○○ | ⊕⊕○○ | ⊕○○○ | ⊕○○○ | ⊕○○○ | ⊕○○○ | ⊕⊕○○ | ⊕○○○ | ⊕⊕○○ |
| 60-69 years | 0.65 (0.30 to 0.95) |  | 2.24 (1.85 to 2.54) |  | 1.89 (1.55 to 2.19) |  | 0.71 (0 to 1.43) | no evidence | no evidence |
|  | ⊕○○○ | ⊕⊕○○ | ⊕○○○ | ⊕○○○ | ⊕○○○ | ⊕○○○ | ⊕○○○ |  |  |
| 70-74 years | 0.92 (0.43 to 1.34) |  | 3.17 (2.62 to 3.60) |  | 2.68 (2.20 to 3.11) |  | 1.41 (0 to 2.81) | no evidence | no evidence |
|  | ⊕○○○ | ⊕○○○ | ⊕○○○ | ⊕○○○ | ⊕○○○ | ⊕○○○ | ⊕○○○ |  |  |

**Abbreviations:** ACM: all-cause mortality; ATSA: adherence to screen analysis; BrCa: breast cancer; CC: case-control study; OBS: Observational study; OR: odds ratio; RCT: randomized controlled trial; RR: Relative risk; ODX: Overdiagnosis. **Explanation:** Red values indicate an increase (rather than a reduction) in risk of outcome. Relative effects are presented in italics.
<sup>a</sup> RR 1.55 (1.23 to 2.11)
<sup>b</sup> RR 1.11 (1.05 to 1.17)
<sup>c</sup> RR 1.09 (0.82 to 1.45)
<sup>d</sup> RR 1.08 (1.01 to 1.16)

Legend
|  |
| --- |
| Very low certainty of evidence |
| Low certainty of evidence |
| No evidence |

#### Pooling of results

It was decided *a priori* to present the results from the RCTs and the observational studies separately due to important differences in study design. Observational results were further stratified by study design (cohort, case-control, ecological and time series) due to differential baseline risk of study bias across the study designs. For exploratory purposes, RCTs and cohort studies were also presented on a single forest plot to visualize relative effects and heterogeneity across studies presenting the same effect measures for mortality outcomes (Appendix 9).

To determine if pooling of studies was appropriate for a single outcome, we first summarized the characteristics of each included study and compared/contrasted each element of the PICOT criteria to explore potential drivers of heterogeneity across study designs. We assessed statistical heterogeneity using the I^2^ statistic. Studies were pooled if (1) studies were similar enough across PICOT criteria elements and (2) there was not substantial statistical heterogeneity (I^2^≤60%). In cases of substantial to considerable heterogeneity (I^2^>60%), pooling was also considered if clinical heterogeneity could be explained via subgroups analyses or meta-regression.^48^

When deemed appropriate, studies were pooled using R software^49^ using the DerSimonian and Laird random-effects method.^50^ Forest plots were used to visually represent data and data were summarized using ranges of included estimates from individual studies (Appendix 8). If new data were identified that could be added to previous meta-analyses conducted in the 2017 review, and it was deemed appropriate using the above criteria, we pooled the estimates of all the studies (existing and new). In cases where data could not be pooled, we adopted a descriptive analysis approach.^51^ . To facilitate the evaluation of certainty of evidence, pooled estimates were used to inform imprecision ratings.

#### Subgroup analyses

Several subgroups were designated to be of interest *a priori*. We planned to explore population subgroups based on age (40-44 years, 45-49 years, 50-59 years, 60-69 years, 70-74 years, 75 and older), ethnicity and race, socioeconomic status, availability of mammography screening, breast density (e.g., extremely [e.g., BI-RADS category D] vs BI-RADS A-C), and family history. Planned mammography screening subgroups were related to screening interval (≤12 months, 13 - 24 months, >24 months) and screening with and without breast self-exam or clinical breast exam. Outcome subgroups include chemotherapy receipt with or without anthracycline, stage at diagnosis, and overdiagnosis and interval cancer outcomes by invasive only vs. including both invasive and DCIS cancers. In non-randomized studies, we also planned subgroups for the type of analysis (i.e., adherence vs intention/offer to screen data). Where available, results are reported for planned subgroups of interest.

#### Sensitivity analyses

We had planned *a priori* to perform sensitivity analyses to explore the robustness and reliability of the review findings. This involved varying parameters within the analysis to evaluate the impact on the overall results. The parameters of interest included studies enrolling 20% or more participants at high risk for breast cancer, type of randomization in RCTs (i.e., randomization vs. quasi randomization), and studies with high vs low/unclear RoB.

#### Analysis of additional imaging data

Using similar methods from our prior review, rates of breast cancer screening mammography requiring additional imaging in those without breast cancer (formerly referred to as “false positives”) were calculated using data from the 2011-2012 CPAC report.^52^ We searched online sources for publicly available quality indicator data for breast cancer screening programs within each province and territory. Eligible data were those that provided recall rates, cancer detection rates and non-malignant biopsy rates stratified by initial versus subsequent screens and by age decade.

CPAC data from 2011-2012 were selected because it provided the most recent publicly available Canadian data for initial versus subsequent screen by age decade. For exploratory purposes, we supplemented these data with more recent provincial data from British Columbia (2019). Any additional identified data from provincial and territorial repositories were also included as supplementary information to help determine the generalizability of the 2011-2012 data to the current Canadian context and observe any changes in rates over time.

Three different additional imaging outcomes were calculated: 1) additional imaging with or without biopsy (no cancer) calculated as the recall rate minus the cancer detection rate, 2) additional imaging with no biopsy (no cancer) calculated as the recall rate minus the sum of the cancer detection rate and non-malignant biopsy rate, and 3) additional imaging and biopsy (no cancer) which is represented as the non-malignant biopsy rate. These outcomes were calculated to approximate rates over a ten-year period. We assumed that women received at least four screens over a 10-year period, if most women would receive a screen every two years (approximating biennial screening for the majority with non-perfect adherence over a 10-year period and noting that some provinces currently offer^53^ or recommend^54^ annual screening in women aged 40-49 or starting at age 45). Two different scenarios were also calculated: 1) assuming individuals started biennial screening in the current age decade (calculated using one initial screen and three subsequent screens over a ten-year period) and 2) assuming individuals started biennial screening in the prior age decade (calculated using four subsequent screens over a 10-year period). For interpretation, we assumed that in each screening round the number of screens was equivalent to the number of women screened.

#### Analysis of overdiagnosis outcomes

For outcomes on overdiagnosis, the previous review did not conduct a quantitative synthesis but rather summarized narratively what previous study authors reported. In this review, we estimated overdiagnosis (sometimes referred to as overdetection; however, for the purposes of this review we use the term overdiagnosis^55^) for trial data using a cumulative-incidence approach. Studies have suggested this to be a robust method to estimate overdiagnosis from RCT data in which there are several years of follow-up after screening stops and the control group is never screened.^56–59^ Cited limitations of this approach include limited external validity, diluted estimates of overdiagnosis, and a dependence on appropriate length of follow-up time.^58,59^ Overdiagnosis estimates from observational studies were reported as calculated by study authors. Estimates derived from observational studies may provide a more contemporary estimate of overdiagnosis, but are also more susceptible to bias due to confounding.^58,59^

### Certainty of the evidence (GRADE)

We assessed the overall certainty of evidence for each outcome with the Grading of Recommendations Assessment, Development and Evaluation (GRADE) framework.^36^ GRADE assessments were performed or updated using the following criteria for all evidence sets. Four domains were graded (imprecision, inconsistency, indirectness, and risk of bias) which were used to inform our overall rating of the certainty of evidence.

While we had planned to assess publication bias using funnel plots, we were unable to do so due to an insufficient number of studies (ten) included for each outcome.^60^ We included footnotes in all summary of findings table detailing the ratings for each domain and our rationale.

Risk of bias ratings were informed by our assessments for each study related to each outcome. Imprecision was assessed using a minimally contextualized approach, wherein evidence was downrated for imprecision if the estimate’s 95% confidence intervals crossed the threshold for an important effect.^61^ Thresholds for each outcome that the WG agreed would be important in a patient’s decision-making were determined *a priori* by the WG (Appendix 10) using surveys followed by a consensus method.

(Appendix 11). For indirectness, we considered factors related to the generalizability of the evidence for a particular outcome to our PICO criteria and a contemporary Canadian context. We downrated for inconsistency if approximately half of the estimates in a single evidence set for an outcome fell on either side of our threshold for an important effect (i.e., half considered to be an important effect and half considered to be a trivial effect).

Following GRADE guidance,^36^ RCTs were graded with evidence starting at “high” certainty of evidence and then downrated, when necessary, based on individual domain ratings. Non-randomized studies, including observational studies and evidence considered to be observational in nature (e.g., additional imaging outcomes), were graded starting at “low” certainty of evidence. We considered uprating evidence if a plausible “large effect” was observed, considered if the relative effect estimate was above 2.0 or below 0.5 based on consistent evidence from at least two studies, with no plausible confounders.^62,63^

## Results

### Protocol deviations

Minor deviations were made from the original protocol. We used the JBI critical appraisal tools^44^ which provided a structured and directed questionnaire based on study design (e.g., case control, cohort, quasi-experimental), rather than the ROBINS-I tool. We found the JBI tool to be more efficient and better accounted for sources of bias in some non-randomized designs. Although observational studies with screening that took place entirely prior to 2000 were excluded, we included studies where the screening period started prior to 2000 and continued after 2000, due to a dearth of studies with screening performed exclusively after 2000. There were no RCTs which included screening performed after 2000. We also changed the language for our target population from an “average risk” population to a “general risk” population, as we found this better reflected the range of potential risk groups that were represented in included studies in an undifferentiated general population. No changes to the inclusion/exclusion criteria were made relating to this change in labelling our target population.

We performed a scaling adjustment for baseline risk for different risk categories (i.e., family history and breast density) based on the estimated calculated incidence rates over 10 years from the Pan Canadian study. This change was requested and approved by the WG to provide an inferred risk for these risk categories given the lack of reported data from included studies. The results of the scaled adjustment can be found in Appendix 8, Tables 1-3 and 5.

We did not perform an evidence review for impacts of screening on intermediate and high-risk groups, rather, we extrapolated the benefit of mammographic screening in a general risk population to the expected increased incidence of breast cancer in intermediate and high-risk groups to attempt to gain an understanding of the benefits of screening in these populations. These baseline risks were extrapolated from published data pertaining to the general population, and therefore have significant limitations, as mortality risks may be different in these groups, and screening may be more or less effective. To estimate a moderately increased baseline risk due to having a family history of cancer, we used the estimate from Engmann et al. that suggested that the odds of developing breast cancer are approximately 1.6 times higher than individuals who do not have a family history of breast cancer.^64^ To estimate a moderately increased baseline risk due to breast density, a review suggested that the risk of dying from breast cancer is approximately 1.9 times higher than individuals who do not have dense breasts.^65^ The baseline risk was calculated by using the incidence of breast cancer mortality in an unscreened Canadian population and multiplying by the relative increase in lifetime risk reported for those with a family history of breast cancer or dense breasts. The calculations for family history were performed under the assumption that cancers diagnosed within the moderately increased risk population would have similar mortality rates as those in the general risk population. The multiplier for breast density was based on differences in mortality rates and would consider differences in incidence and mortality for unscreened women.^66^ As noted above, individuals at high risk for breast cancer (>20% lifetime risk) were not considered in this review. Refer to Appendix 12 for calculations.

Finally, given the paucity of data from included studies reporting on lifetime risk, an additional ad hoc analysis was performed to consider the lifetime mortality risk reduction for women 40-49. For Canadian women, the extrapolated lifetime risk of dying from breast cancer is 27.8 per 1,000 and 17.5% of breast cancer deaths come from cancers that arise in the 40-to-49-year age band.^67^ This gives a lifetime breast cancer mortality rate for unscreened women of 4.86 deaths per 1,000 women. Additional calculations can be found in Appendix 12.

### Results of the search

A total of 9,733 electronic records were identified and an additional 144 records were identified through the online portal. Following de-duplication, 9,745 titles and abstracts were screened, of which 1,678 were excluded from human screening and 7,863 were excluded from AI screening. 204 full-text records were assessed for eligibility, of which 175 were excluded. The PRISMA diagram (Figure 1) and Appendix 13 provides detailed reasons for exclusion.

The three RCTs included were updates on existing RCT trials (one report is on the longer follow-up for the AGE trial,^68^ one report is a health technology assessment (HTA) for the AGE trial,^69^ and one report is an analysis on RCT data reporting on newly added outcome of stage at diagnosis.^70^ No new RCTs were identified in this update.

The new papers reporting on RCT data were added to the ten existing RCTs from the 2017 review. 26 observational studies were included (nine cohort,^57,71–78^ nine case-control,^66,79–86^ five time-trend analyses,^87–91^ and three ecological studies^47,92,93^).

### Study characteristics

Study characteristics of the entire body of evidence are outlined in Appendix 14 and study characteristics of RCTs from previous review are outlined in Appendix 15. Of the 10 RCTs, two were quasi-randomized,^8,94^ two were cluster-randomized trials,^95–97^ and six were parallel-group randomized trials.^10,68,98–103^ The dates of screening for the RCTs ranged from 1963 to 1990, while in the cohort studies, screening dates ranged from 1991 to 2016 (Table 3). For case-control studies, screening dates ranged from 1975 to 2013 and for time series and ecological studies screening dates ranged from 1977 to 2015. For the RCTs, six were conducted in Sweden, two in Canada, one in the United Kingdom, and one in the United States. For the observational studies, four were conducted in Canada, four in the United States, four in The Netherlands, two each from Sweden, Australia, Norway, and United Kingdom, and one each in the Republic of Korea, New Zealand, Italy, Belgium, Germany, and Finland. For the included trials, sample size at randomisation ranged from approximately 18,000 to 160,000. For observational studies, the sample size ranged from approximately 2,000 to over 8,000,000 in the large cohort studies. Mean trial follow-up ranged from 18 years to 30 years, while in cohort studies this range from 7 to 22 years.

**Table 3:** List of included studies and the years of screening.

| **Trial name or author year** | **Country** | **Study design** | **Dates of screening** |
| --- | --- | --- | --- |
| HIP<sup>70,152,153</sup> | United States | RCT | 1963-1966 |
| Malmo I<sup>9,70,154</sup> | Sweden | RCT | 1976-1988 |
| Malmo II<sup>9,70,154</sup> | Sweden | RCT | 1978-1990 |
| Swedish Two County (Ostergotland)<sup>70,154-156</sup> | Sweden | RCT | 1977-1984 |
| Swedish Two County (Kopparberg)<sup>70,155,156</sup> | Sweden | RCT | 1978-1985 |
| CNBSS 1<sup>10,70</sup> | Canada | RCT | 1980-1984 |
| CNBSS2<sup>10,70</sup> | Canada | RCT | 1980-1984 |
| Stockholm<sup>9,70,154</sup> | Sweden | RCT | 1981-1985 |
| Göteborg<sup>9,98</sup> | Sweden | RCT | 1982-1989 |
| AGE<sup>69,157,68,</sup> | United Kingdom | RCT | 1990-1997 |
| Choi 2021<sup>71</sup> | Korea | Cohort | 2002-2015 |
| Duffy 2021<sup>72</sup> | Sweden | Cohort | 1992-2016 |
| Dunn 2021<sup>73</sup> | Australia | Cohort | 2000-2005 |
| Morrell 2017<sup>74</sup> | New Zealand | Cohort | 2000-2011 |
| Lund 2018<sup>75</sup> | Norway | Cohort | 2005-2013 |
| Puliti 2017<sup>57</sup> | Italy | Cohort | 1991-2002 |
| Weedon-Fekjaer 2014<sup>76</sup> | Norway | Cohort | 1995-2009 |
| Richman 2023<sup>77</sup> | United States | Cohort | 2002-2005 |
| Garcia-Albeniz 2020<sup>78</sup> | United States | Cohort | 2000-2008 |
| Blyuss 2023<sup>79</sup> | United Kingdom | Case-control | 1988-2011 |
| De Troeyer 2023<sup>80</sup> | Belgium | Case-control | 2005-2012 |
| Maroni 2021<sup>81</sup> | United Kingdom | Case-control | 1990-2011 |
| van der Waal 2017<sup>66</sup> | Netherlands | Case-control | 1975-2006 |
| Massat 2016<sup>158</sup> | United Kingdom | Case-control | 1990-2009 |
| Beckmann 2015<sup>159</sup> | Australia | Case-control | 1989-2010 |
| Pocobelli 2015<sup>84</sup> | Canada | Case-control | 1995-2008 |
| Paap 2014<sup>85</sup> | Netherlands | Case-control | 1989-2009 |
| Ripping 2016<sup>86</sup> | Netherlands | Case-control | 1975-2013 |
| Katalinic 2020<sup>160</sup> | Germany | Time series | 2013/2014 |
| de Glas 2014<sup>88</sup> | Netherlands | Time series | 1998-2002 |
| Parvinen 2015<sup>161</sup> | Finland | Time series | 1987-2009 |
| Helvie 2014<sup>162</sup> | United States | Time series | 2007-2009 |
| Tabar 2019<sup>91</sup> | Sweden | Time series | 1977-2015 |
| Wilkinson 2023<sup>92</sup> | Canada | Ecological | 2002-2007 |
| Wilkinson 2022<sup>93</sup> | Canada | Ecological | 2002-2007 |
| Coldman 2014<sup>163</sup> | Canada | Ecological | 1990-2009 |

Age was one of the only consistently reported participant characteristics across all studies. Several studies provided outcome data by age; however, the reporting of age interval varied by study (e.g., ten-year age bands vs. five-year age bands). Age of study entry ranged from 39 years to 74 years for RCTs, while observational studies included women from 40 years to 84 years. Limited information regarding other study population characteristics (race/ethnicity, family history, breast density, or socioeconomic status) was reported across trials or observational studies and, therefore, no subgroup analyses were performed for these factors. The ethnicity of participants was reported by one observational study.^74^ A single observational study reported on the proportion of participants with family history of breast cancer.^75^ The proportion of women with dense/fatty breasts was only reported by one case control study.^66^ The Canadian trials and one observational study,^74^ respectively, reported information on socioeconomic status.

For RCTs, film mammography alone was used in seven trials, while three trials screened with film mammography with the addition of clinical breast exam. Five trials used single-view mammography only, and one trial included only two screens, while the rest had four to five screens. The duration of the screening period ranged from three to twelve years. Screening intervals ranged from 12 months to 33 months and the attendance rate ranged from 65% to 88%. The comparator arm for all trials was usual care, and in six RCTs the control group received mammography screening at the end of the screening period.

A single observational study compared those invited to screen and those not invited to screen.^76^ All other cohort and case-control studies provided analyses related to participation/adherence to screening compared to non-participation/adherence to screening. Two ecological studies compared outcomes in five jurisdictions with organized breast cancer screening programs for those aged 40-49 compared to jurisdictions with no screening programs available for that age group.^92,93^ Five ecological studies compared outcomes in periods prior to the implementation of a screening program and after the implementation of such a program.^88–91,104^ The type of mammography provided was not reported in the majority of studies.^47,57,72,73,76,77,79,81,82,84,85,88–93,104^ Two studies specified the use of digital mammography,^75,80^ while two studies used film mammography alone.^66,83^ The remaining studies noted that screening programs included a mix of film and digital mammography, with results not stratified by the type of mammography received.^71,74,86^ No information on the availability of mammography screening or the screening interval was provided for observational studies. When reported, the number of screening rounds that patients underwent varied both within studies and between ranging between at least one mammography screen up to six rounds.^57,66,68,71,74,79–83,85^

### Risk of bias

Overall, three RCTs were rated with an overall risk of bias judgment of moderate risk (UK Age, Malmo I and II), while seven were considered high risk (Gothenburg, Stockholm, Swedish Two County, HIP, and CNBSS I and II). A detailed risk of bias assessment for all RCT outcomes was included in the 2017 report.^34^ All new risk of bias assessments (stage at diagnosis and treatment-related morbidity) and any updated risk of bias assessments can be found in Appendix 16. All prior risk of bias assessments included in the 2017 review^34^ were verified to ensure consistency in ratings between teams. No ratings from the 2017 review were modified, except for those related to the CNBSS (I & II) trials. Following review, this rating was modified from moderate to high risk for the domains of randomization generation and allocation concealment.

There have been concerns expressed about the CNBSS trials with respect to subversion of randomization, causing imbalance between the groups with more symptomatic cancers allocated to the study arm.^18,21,70,105,106^ In addition to introducing bias to mortality estimates this could inflate cancer incidence in the intervention arm which could have been inaccurately coded as overdiagnosis. Others have argued that there are no serious concerns with the trial.^107,108^ We chose to acknowledge the concerns that have been raised with the CBNSS trial and explore the effect of these trials on outcomes of interest (see sensitivity analyses section below).

For the observational studies, overall RoB ratings ranged from high to low, varying by study design (see full summary in Appendix 16, Table 1). All cohort studies except for one were deemed to have recruited similar populations as they were recruited from the same population of women invited to a singular population-based screening program. This domain was downrated in one retrospective cohort study where a portion of individual-level data from never-screened women was missing (i.e., the female population with no recorded screening or breast cancer history).^74^ Person-years from this group were inferred from the median age for ethnic- and age-specific census-derived populations for that year, as provided by Statistics New Zealand.^74^ In relation to the comparability of the cohorts, there was a lack of reporting about adjustment for important confounding factors across studies, including use of hormone replacement therapy, socioeconomic status, or self-selection bias.^72,74^ Lastly, two studies did not report average follow-up length and reasons for loss to follow-up were not reported.^72,74^ Cases and controls were not age-matched in two studies^66,80^ or failed to report adjusting for important confounding factors related to self-selection bias.^66,80–82,84–86^ Three case-control studies did not provide screening details or confirm all women were invited to screening.^82,84,86^ For time-series and ecological studies, there was a lack of reporting on participant characteristics, loss to follow-up and missing data. Five studies also failed to report how outcomes were measured and if this varied over time.^88–91,104^ Ecological studies are more susceptible to bias due to the potential for other changes in context to occur around the time at which the intervention is introduced that also influence the outcome.^109^

### Pooling and Subgroup Analyses

We were unable to perform planned subgroup analyses for most factors of interest due to a lack of data reported for these factors (e.g., race/ethnicity, family history, breast density, socioeconomic status). Where possible, results are presented by age groups, short- vs. long-case accrual, type of screening analysis (e.g., invitation to screen vs. adherence to screen) and screening interval.

A previous subgroup analysis^26^ detected no statistically significant differences in relative risk (RR) of breast cancer mortality associated with screening between age subgroups (age range 40 to 74 years) and concluded that true differences resulting from age were unlikely. Therefore, we used the all-ages RR data rather than focusing on each age decade throughout our GRADE assessments.

We found no direct data on the effect of breast density or family history on outcomes of interest, and so the overall RR for mortality was used to extrapolate the variation in absolute effects across different baseline risk categories (i.e., general population risk, moderately increased risk due to family history, or moderately increased risk due dense breasts). The results applied to a moderately increased risk population due to family history and dense breasts are presented in Appendix 17.

For the observational studies, significant clinical and statistical heterogeneity in the data and an inability to explore this heterogeneity via subgroup analyses precluded us from pooling most outcomes. Sources of clinical heterogeneity likely included both population and study design factors, included differing definitions of “screened” (e.g., serial screeners vs. screened at least once), length of follow-up, different methods of effect estimation, and varying adjustment for confounding factors.

### Sensitivity Analyses

We performed pre-planned sensitivity analyses by removing all high risk of bias trials for outcomes of critical importance (Appendix 18 and 19). A sensitivity analysis including only RCTs at moderate risk of bias for mortality and overdiagnosis outcomes yielded similar results with or without high-risk studies. A sensitivity analysis was also performed for the overdiagnosis outcome by removing the single included study with high risk of bias for outcome (CNBSS). A sensitivity analysis was also performed for pooled observational studies, removing high risk of bias studies, yielding similar results.

We were unable to perform planned sensitivity analyses for other factors of interest due to a lack of data reported for these factors (e.g., studies enrolling 20% or more participants at high risk for breast cancer) or an insufficient number of studies (i.e., type of randomization).

### Findings

#### Breast Cancer Mortality

The short-accrual findings from the RCTs are presented here. Long-case accrual results from the RCTs are available in Appendix 8, Table 2. For the RCTs, the comparison groups were mammography screening (with or without CBE) compared to usual care and all results are applicable to a general population of women. Based on the inferred baseline risk from the Pan Canadian Study, absolute effects were estimated over 10 years, but the length of follow-up observed in RCTs ranged from 13.1 to 30 years. For the observational studies, the comparison groups were adherence to screening with mammography vs. no screening (range of follow-up 8.0 to 38.0 years) except where noted. The overall GRADE ratings were low or very low for all the estimates reported below (Appendix 8, tables 1-6).

##### a) 40 to 49 years

RCTs^10,94,97,99,100^ reported 0.27 fewer breast cancer deaths per 1,000 and observational studies reported 0.79 and 0.94 fewer deaths per 1,000, for case-control^66,80–82,84–86^ and cohort studies^47,71,72,74^, respectively. In studies^89,104^ comparing mortality rates before and after the introduction of BC screening programs, 0.03 fewer deaths per 1,000 person-years were reported post-screening introduction.

##### b) 50 to 59 years

RCTs^10,94,97,99,100^ reported 0.50 fewer breast cancer deaths per 1,000 and observational studies reported 1.45 and 1.72 fewer deaths per 1,000, for case-control^66,80–82,84–86^ and cohort studies^47,71,72,74^, respectively. In studies^89,104^ comparing mortality rates before and after the introduction of BC screening programs values ranged from 0.13 fewer to 0.02 more deaths per 1,000 person-years.

##### c) 60 to 69 years

RCTs^10,94,97,99,100^ reported 0.65 fewer breast cancer deaths per 1,000 and in observational studies reported 1.89 and2.24 fewer deaths per 1,000, for case-control^66,80–82,84–86^ and cohort studies^47,71,72,74^, respectively. In one study^104^ comparing mortality rates before and after the introduction of BC screening programs, 0.17 fewer deaths per 1,000 person-years were reported post-screening introduction, compared to another study^89^ that found 0.21 more per 1,000 person-years post screening introduction for those aged 60 to 74 years.

##### d) 70 to 74 years

RCTs^10,94,97,99,100^ reported 0.92 fewer breast cancer deaths per 1,000 and in observational studies values reported 2.68 and 3.17 fewer deaths per 1,000, for case-control^66,80–82,84–86^ and cohort studies^47,71,72,74^, respectively. For the observational analysis related to stopping vs. continuing screening, one study^78^ reported 0.81 fewer deaths per 1,000 in those who continued screening into their 70s. In one study^104^ comparing mortality rates before and after the introduction of BC screening programs, 0.02 more deaths per 1,000 person-years were reported post-screening introduction.

##### e) 75+ years

For the observational analysis related to stopping vs. continuing screening, one study^78^ reported 0 fewer deaths per 1,000 in those who continued screening into their 70s. In one study^89^ comparing mortality rates before and after the introduction of BC screening programs, 0.12 more deaths per 1,000 person-years were reported post-screening introduction.

##### f) All ages

In one time-trend analysis study^91^ comparing mortality rates before and after the introduction of BC screening programs among women who either did or did not participate in mammography screening, there were 0.30 fewer deaths per 1,000 person-years. The pre-screening period (1958 to 1976) included women who did not have the opportunity to screen compared to those who were invited and participated during the active screening period (1977 to 2015). The same time-trend study presented a comparison within the active screening period between women who were invited and participated in screening versus women who were invited and did not participate and found 0.37 fewer deaths per 1,000 person-years. Results from both periods were provided to enable both contemporaneous and historical comparisons of breast cancer mortality among women before the onset of the screening programs (1958-1976) and starting in 1977 among those who did and did not participate in mammography screening.

#### All-cause mortality

The comparison groups were mammography screening (with or without CBE) compared to usual care in included RCTs.^8,10,99,110^ Based on the inferred baseline risk, absolute effects were estimated over ten years, but the length of follow-up observed in RCTs ranged from 7.9 to 13 years. No observational evidence met inclusion criteria. The overall GRADE ratings were low or very low for all all-cause mortality estimates (Appendix 8, Table 7).

For the following analysis, we did not extract baseline risk estimates from included studies, but rather used data from Statistics Canada to extrapolate these estimates. In those aged 40 to 49 years, RCTs reported 0.13 fewer deaths per 1,000 due to any cause over 10 years. This value was 0.31 fewer deaths per 1,000 in those aged 50 to 59 years, 0.71 in those aged 60 to 69 years, and 1.41 in those aged 70 to 74 years.

#### Breast Cancer Stage at diagnosis

For RCTs, the comparison groups were mammography screening (with or without CBE) compared to usual care. The length of follow-up observed in RCTs ranged from 5 to 10 years. For the observational studies, the comparison groups were adherence to screening with mammography vs. no screening (maximum follow-up 11 to 13 years, minimum NR), except where noted. The overall GRADE ratings were very low for all of the estimates reported below (Appendix 8, Tables 8-10).

##### a) 40 to 49 years

One RCT^70^ reported one more breast cancer case diagnosed at stage II or higher per 1,000. In one observational study^93^ comparing jurisdictions with and without breast cancer screening programs available to those aged 40-49, they reported on the proportion diagnosed by stage: 30 fewer breast cancers diagnosed at stage II per 1,000 breast cancers that occurred, 27 fewer at stage III per 1,000 breast cancers that occurred, and 7 fewer breast cancers diagnosed at stage IV per 1,000 cancers that occurred.

##### b) 50 to 59 years

One RCT^70^ reported 0 fewer breast cancers diagnosed at stage II or higher per 1,000 breast cancers that occur.

##### c) 70-74 years

In ecological studies^57,74^ comparing mortality rates before and after the introduction of BC screening programs, 0.07 to 0.13 fewer cancers per 1,000 person-years were diagnosed at stages III and IV post-screening introduction, depending on the reference period.

##### d) 75+ years

In one ecological study^88^ comparing outcomes before and after the introduction of BC screening programs, 0.01 to 0.03 more cancers per 1,000 person-years were diagnosed at stages III to IV post-screening introduction, depending on the reference period.

##### e) All ages

In RCTs^70^, the screening group reported 3 fewer breast cancers diagnosed at Stage II or higher per 1,000 and one fewer breast cancer diagnosed at Stage III or higher per 1,000. One observational study^57^ found that screening results in 0.51 fewer breast cancers diagnosed at stage II or higher per 1,000. We were unable to estimate the absolute effect (baseline risk NR) for another observational study^74^ that reported a RR of 0.44 in a screening group compared to a non-screening group. In one study^88^ comparing outcomes before and after the introduction of BC screening programs 0.10 fewer cancers were diagnosed at late stage (regional) and 0.01 more were diagnosed at late stage (distant) per 1,000 person-years post-screening introduction.

#### Overdiagnosis

For RCTs, the comparison groups were mammography screening (with or without CBE) compared to usual care over 10 years (range of follow-up 9 to 15 years). For the observational studies, the comparison groups were adherence to screening with mammography vs. no screening over (range of follow-up 8 to 15 years), except where noted. The overall GRADE ratings were either low or very low for all of the estimates reported below (Appendix 8, Tables 11 to 12).

##### a) 40 to 49 years

In RCTs^68,101,102^, the screening group reported 1.95 breast cancers (invasive and in situ) overdiagnosed per 1,000 and 1.0 overdiagnosed invasive cancers per 1,000. The sensitivity analysis removing high risk of bias studies resulted in 1.57 breast cancers (invasive and in situ) overdiagnosed per 1,000 and 0.49 invasive cancers per 1,000. One observational study^75^ reported 1.42 invasive and in situ breast cancers overdiagnosed per 1,000 person-years.

##### b) 50 to 59 years

In RCTs^68,101,102^, the screening group reported 1.93 breast cancers (invasive and in situ) overdiagnosed per 1,000 and 1.18 invasive cancers overdiagnosed per 1,000. The sensitivity analysis removing high risk of bias studies resulted in 3.95 breast cancers (invasive and in situ) overdiagnosed per 1,000 and 2.81 invasive cancers per 1,000. One observational study^75^ reported 0.42 fewer invasive and in situ breast cancers overdiagnosed per 1,000 person-years. Another observational study found^57^ 0.34 invasive and in situ breast cancers overdiagnosed per 1,000 individuals aged 50 to 69 years.

##### c) 60 to 69 years

One observational study^75^ reported 0.15 fewer invasive and in situ diagnosed breast cancers per 1,000 person-years. Another observational study found^57^ 0.34 invasive and in situ breast cancers overdiagnosed per 1,000 individuals aged 50 to 69 years.

##### d) 70 to 74 years

One observational study^77^ reported 20 invasive and in situ breast cancers overdiagnosed per 1,000 individuals.

##### e) 75 + years

One observational study^77^ reported a range between 23 and 57 invasive and in situ breast cancers overdiagnosed per 1,000 individuals.

#### Interval cancers

Interval cancer rates were reported in the intervention arm only of included RCTs (screening with mammography with or without CBE) and were thus treated as descriptive data. Length of follow-up ranged from 4.8 to 7 years. The overall GRADE ratings were low or very low for all of the estimates reported below (Appendix 8, Table 13).

##### a) 40-49 years

In RCTs,^98^ 3.0 interval cancers (2.8 invasive and 0.2 DCIS only), were detected in the mammography arm per 1,000 over the follow-up period of 4.8-7 years (screening interval 18 months).

##### b) 50-59 years

In RCTs,^98^ 1.9 interval cancers (invasive and DCIS), were detected in the mammography arm per 1,000 over the follow-up period of 4.8-7 years (screening interval 18 months).

##### c) All ages

In RCTs,^10,97,98,111,112^ 3.9 interval cancers (invasive and DCIS), were detected per 1,000 over the follow-up period of 5 years (screening interval 12 months). For a screening interval of 13-24 months, 3.1 interval cancers (invasive and DCIS) were detected per 1,000 over a follow-up period of 4.8-7 years and 3.9 interval cancers were detected with a screening interval of >24 months with seven years of follow-up. For an 18-month screening interval, 2.8 invasive cancers and 0.2 DCIS were detected per 1,000 over a follow-up period of 4.8-7 years.

#### Treatment-related morbidity

For RCTs, the comparison groups were mammography screening (with or without CBE) compared to usual care (mean follow-up 7 to 9 years). For the observational studies, the comparison groups were adherence to screening with mammography vs. no screening over (follow-up range 8 to 13 years), except where noted. The overall GRADE ratings were low or very low for all of the estimates reported below (Appendix 8, Tables 14-16). It must be noted that breast conserving surgery was not a treatment option during most of the RCTs and so mastectomy rates may reflect therapy and not morbidity.

##### a) 70-74 years

One observational study^57^ that evaluated those who continued screening into their 70s compared to those who stopped screening found 9 more women who had simple mastectomy, 43 fewer women diagnosed who had radical mastectomy, 111 more women who had radiotherapy, and 59 fewer women having chemotherapy per 1,000.

##### b) 75+ years

One observational study^57^ that evaluated those who continued screening into their 70s compared to those who stopped screening found 7 more women who had simple mastectomy, 28 fewer women diagnosed who had radical mastectomy, 93 more women who had radiotherapy, and 29 fewer women per having chemotherapy per 1,000.

##### c) All ages

In RCTs,^113^ the screening group had 1.84 more mastectomies per 1,000, 2.85 more individuals treated with radiotherapy, and 0.14 fewer treated with chemotherapy.. Observational studies reported 0.9 more women with breast cancer treated with breast conservative surgery as treatment per 1,000 and 0.4 fewer women with breast cancer treated with mastectomy as treatment per 1,000.

#### Additional imaging (no cancer)

Additional imaging (no cancer) rates were estimated for each age subgroup using 2011-2012 CPAC data^114^ (and 2019 data from British Columbia for additional imaging rates with or without biopsy). Rates were estimated for a 10-year period. Our overall certainty of evidence was graded as moderate for all outcomes (Appendix 8, Tables 17a-c). Supplementary data on additional imaging (no cancers) can be found in Appendix 20.

##### a) 40 to 49 years

The number of women requiring additional imaging with or without biopsy (no cancer) in 1,000 women screened every 2-3 years over a 10-year period varied depending on the data source. For CPAC data this rate was estimated at 367.5, while based on BC data the rate was higher at 477.6. Similarly, 312.8 per 1,000 women were estimated to require additional imaging no biopsy (no cancer). We estimated 54.7 women requiring additional imaging and biopsy (no cancer) in 1,000 women screened every 2-3 years over a 10-year period.

##### b) 50 to 59 years

Using CPAC data, we estimated 286.4 and 365.5 women requiring additional imaging with or without biopsy (no cancer) in 1,000 women screened every 2-3 years over a 10-year period depending on if women started screening in their 40s or 50s, respectively. For BC data, we estimated 285.2 and 410.5 women requiring additional imaging with or without biopsy (no cancer) in 1,000 women screened every 2-3 years over a 10-year period depending on if women started screening in their 40s or 50s, respectively.

Using CPAC data, we estimated 252.4 and 319.3 women requiring additional imaging no biopsy (no cancer) for 1,000 women screened every 2-3 years over a 10-year period depending on if women started screening in their 40s or 50s, respectively. We estimated 34 to 46.2 women requiring additional imaging and biopsy (no cancer) in 1,000 women screened every 2-3 years over a 10-year period, depending on if women started screening in their 40s or 50s, respectively.

##### c) 60 to 69 years

We estimated 252.4 (BC data) to 257.2 (CPAC data) women requiring additional imaging with or without biopsy (no cancer) per 1,000 women screened every 2-3 years over a 10-year period, depending on the data source. 224.4 women were estimated to require additional imaging no biopsy (no cancer) per 1,000 women screened every 2-3 years over a 10-year period. We estimated 32.8 women requiring additional imaging and biopsy (no cancer) per 1,000 women screened every 2-3 years over a 10-year period.

##### d) 70 to 74 years

We estimated 220.4 (CPAC data) to 238.4 (BC data) women requiring additional imaging with or without biopsy (no cancer) per 1,000 women screened every 2-3 years over a 10-year period, depending on the data source. 190 women were estimated to require additional imaging no biopsy (no cancer) per 1,000 women screened every 2-3 years over a 10-year period. We estimated 30.4 women requiring additional imaging and biopsy (no cancer) per 1,000 women screened every 2-3 years over a 10-year period.

#### Other Outcomes

We found no eligible studies for inclusion that reported on health-related quality of life or life years gained (or lost).

## Discussion

This review update focused on the benefits and harms of screening compared to no screening and built on the previous 2017 review.^26^ The previous review reported a relative reduction in the risk of breast cancer mortality by 15% (RR 0.85 95% CI 0.78 to 0.93). In this review update, based on RCT data the absolute effects for breast cancer mortality by age decade were 0.27 fewer per 1,000 women screened in those aged 40 to 49; 0.50 fewer per 1,000 in those aged 50 to 59; 0.65 fewer per 1,000 in those aged 60 to 69; and 0.92 fewer per 1,000 in those aged 70 to 74, with the overall body of evidence graded as low or very low, similar to the previous 2017 review. We did not find any new RCTs and screening dates from existing RCTs ranged from 1963 to 1991.

Results from observational studies reported a relative mortality risk reduction that ranged from 29% to 62% and absolute effects from breast cancer mortality that ranged from 0.79 to 0.94 fewer per 1,000 in those aged 40 to 49; 1.45 to 1.72 fewer per 1,000 in those aged 50 to 59; 1.89 to 2.24 fewer per 1,000 in those aged 60 to 69; and 2.68 to 3.17 fewer per 1,000 in those aged 70 to 74, with the overall body of observational evidence graded as very low. Eight of the 26 observational studies included had screening performed exclusively after 2000, with the remainder of studies reflective of screening, and, therefore, treatment, in the 1990s. The magnitude of mortality benefit reduction reported in observational studies ranged from 0.94 to 6.03 fewer per 1,000 across all ages over a range of follow-up between 10 and 22 years; however, combining observational studies resulted in high heterogeneity, and due to the nature of the study designs, a significant increase in risk of bias (see Appendix 21). We found little to no evidence to allow us to assess screening benefits and harms in different ethnic and racial groups, by different mammography technology, or in women with dense breasts. Overall, the evidence quality of the current review was assessed as low or very low, suggesting that the evidence is very uncertain about the effect of screening for breast cancer on the outcomes evaluated in this review.

The rates of overdiagnosis for invasive and in situ cancers was 9% to 11% (1.57 to 1.95 more) and 3% to 6% (0.49 to 1 more) for invasive cancers only in those aged 40 to 49. Overdiagnosis rates for those aged 50 to 59 were 8% to 12% (1.93 to 3.95) for invasive and in situ cancers and 5% to 9% (1.18 to 2.81) for invasive cancers only. The previous CTFPHC guideline relied on the CNBSS trial calculations for estimates of overdiagnosis for invasive cancers at 5 years after screening (32% for 40 to 49, 16% for 50 to 59) and 20 years after screening (48% for 40 to 49, 5% for 50 to 59).^41^ The CNBSS calculated overdiagnosis as the number of cancers in the mammography arm less those in the control arm divided by the screen-detected cancers in the mammography arm.^101^ Our calculations for estimating overdiagnosis, which report a reduced rate of overdiagnosis for women 40-49 (3% compared to 48%), are different in that we used a quantitative cumulative-incidence approach (see Appendix 8, Table 11).

### The importance of the year 2000

The inclusion of eligibility criterion for observational studies with screening performed after the year 2000 was intended to examine current societal factors and breast cancer screening and treatment practices more closely. The incidence of breast cancer in women 40 to 49 has significantly increased by 9.1% from 1984 to 2019, with an increase from 127.8 cases per 100,000 to 139.4 cases per 100,000.^115^ The incidence of breast cancer in women 45-49 is higher, at 167.5 cases per 100,000, compared to 216.2 per 100,000 in women 50-54.^115^ Over the same period, breast cancer mortality has decreased by 46% (from 41.2 deaths per 100,000 in 2984 to 22.4 per 100,000 in 2020).^116^ Improved mortality is likely due to both screening and treatment advances. Improved treatment does not obviate the need for screening as survival remains highest in earlier stage disease and improved treatments and screening synergistically improve outcomes.^117^ Since the year 2000, the Canadian population has become more diverse, with the increasing rates of Canadian women of colour, who have an earlier peak age of diagnosis.^118^ Methods for screening and treatment of breast cancer have evolved greatly over the past 60 years, such that most of the RCTs (conducted prior to 2000) are not representative of current clinical practice. The evolution of breast cancer diagnosis and treatment (see Box 1) has been particularly marked since 2000 with fundamental changes including the introduction of digital mammography, discovery/definition of molecular subtypes, targeted therapies (e.g. trastuzumab), aromatase inhibitors, immunotherapy, and the use of genomic assays informing individual recurrence risk and guiding the need for chemotherapy. None of the RCTs included women diagnosed after 2000, and eight observational studies exclusively included women screened after 2000.^117^

#### Box 1

**Timeline of key breast cancer developments. Blue text represents key randomized trials, red represents key treatments, and purple represents the Pan Canadian study.**

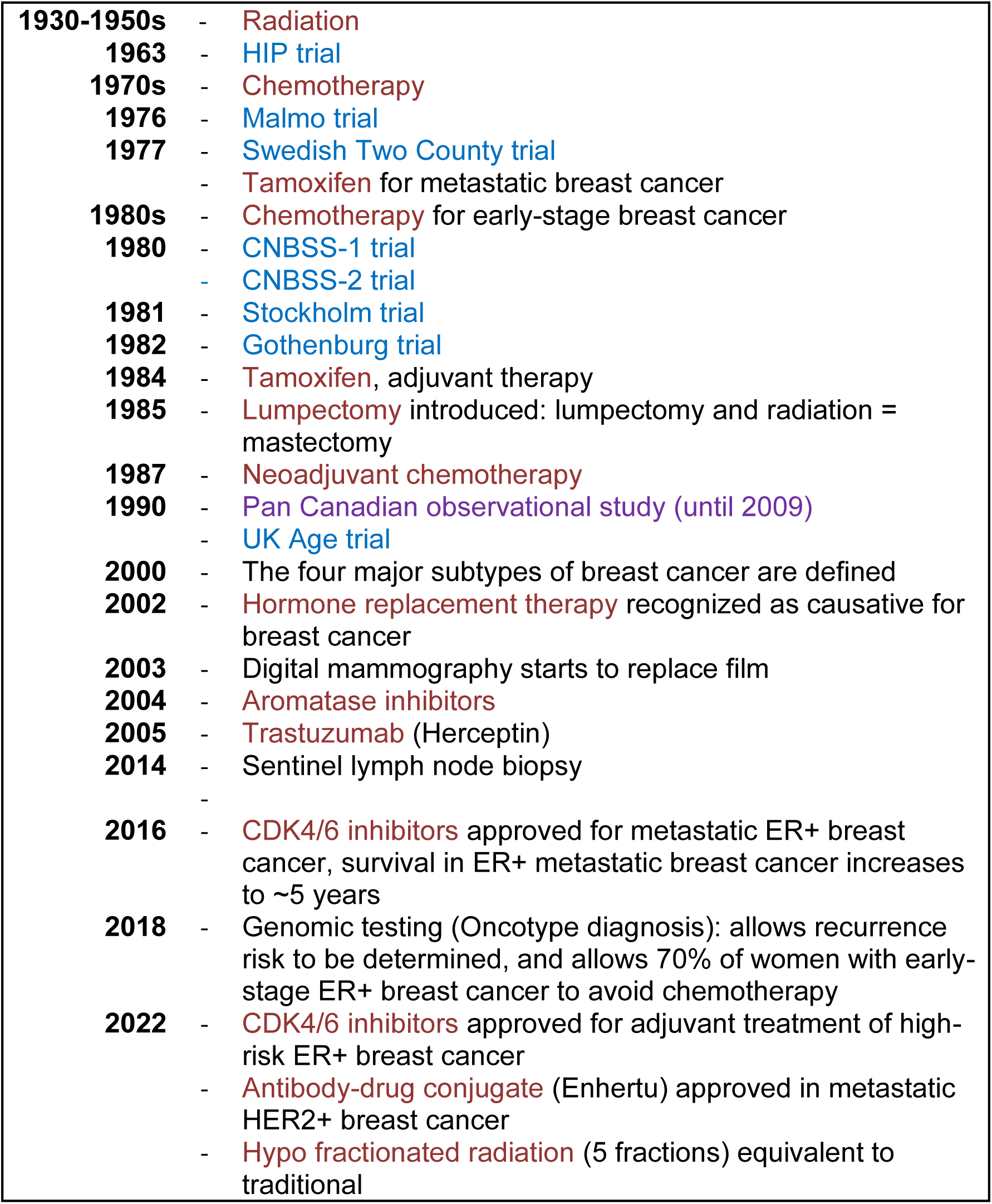

|  |  |
| --- | --- |
| **1930-1950s** | - Radiation |
| **1963** | - HIP trial |
| **1970s** | - Chemotherapy |
| **1976** | - Malmö trial |
| **1977** | - Swedish Two County trial |
|  | - Tamoxifen for metastatic breast cancer |
| **1980s** | - Chemotherapy for early-stage breast cancer |
| **1980** | - CNBSS-1 trial |
|  | - CNBSS-2 trial |
| **1981** | - Stockholm trial |
| **1982** | - Gothenburg trial |
| **1984** | - Tamoxifen, adjuvant therapy |
| **1985** | - Lumpectomy introduced: lumpectomy and radiation = mastectomy |
| **1987** | - Neoadjuvant chemotherapy |
| **1990** | - Pan Canadian observational study (until 2009) |
|  | - UK Age trial |
| **2000** | - The four major subtypes of breast cancer are defined |
| **2002** | - Hormone replacement therapy recognized as causative for breast cancer |
| **2003** | - Digital mammography starts to replace film |
| **2004** | - Aromatase inhibitors |
| **2005** | - Trastuzumab (Herceptin) |
| **2014** | - Sentinel lymph node biopsy |
|  | - |
| **2016** | - CDK4/6 inhibitors approved for metastatic ER+ breast cancer, survival in ER+ metastatic breast cancer increases to ~5 years |
| **2018** | - Genomic testing (Oncotype diagnosis): allows recurrence risk to be determined, and allows 70% of women with early-stage ER+ breast cancer to avoid chemotherapy |
| **2022** | - CDK4/6 inhibitors approved for adjuvant treatment of high-risk ER+ breast cancer |
|  | - Antibody-drug conjugate (Enhertu) approved in metastatic HER2+ breast cancer |
|  | - Hypofractionated radiation (5 fractions) equivalent to traditional |

### RCT vs. observational studies

The difference in the magnitude of benefit of screening noted between the RCTs and observational studies could be due to many factors. RCTs are considered the “gold standard” to evaluate the efficacy of interventions such as screening since they are ideally designed to balance intervention and control arms to prevent selection bias and thereby adjust for recognized and unrecognized confounders. Observational studies are typically limited to screened individuals, compared to the intent-to-screen analysis in RCTs. As an example of a potential bias, individuals who are more health conscious may be more likely to access screening, and so the “healthy user effect” may bias estimates, increasing the mortality benefit in observational studies. Length-time bias may be seen in both types of trials, where more indolent (or low grade) cancer grows more slowly and is therefore more likely to be detected by screening, rather than a more aggressive (or high grade) cancer that is more likely to be clinically detected between screening rounds. Some of the observational studies are more recent than the RCTs and may better reflect the benefits of modern imaging techniques and treatments.

Observational studies, compared to RCTs, may exaggerate estimates of treatment benefit as has been shown;^119,120^ however, one study, which compared observational and RCT results for oncology treatments found that the majority of observational results fell within the confidence intervals of RCT results.^121^

### The complexity of breast cancer

Breast cancer is not a single disease, but rather, a collection of distinct histological and molecular entities, for which screening and treatment have different efficacies.^122^ The definition of molecular subtypes used in current practice was first established in 2000,^123^ after most trials studies in this review were conducted. Modelling has suggested that triple negative breast cancer has the greatest mortality reduction due to screening.^117,124^ Aggressive subtypes of breast cancer are more commonly found in younger women, which may influence the mortality benefit of screening. We did not find any evidence that evaluated mortality reduction relative to molecular or histological subtypes. Evaluating screening for breast cancer without appreciating the heterogeneity of molecular subtypes and distribution of molecular subtypes in different age categories does not give a complete picture of benefits and harms.

### Results reported by the 10-year period

Absolute numbers are presented in terms of impacts on 1,000 women over a 10-year period. A 10-year time frame will view impacts of screening on women 40 to 49 in isolation, and not in terms of benefits and harms which may or may not accrue over a lifetime. Ideally, screening decisions for women 40 to 49 should consider impacts on stage at diagnosis,^93^ mortality, and incidence over the next decade and beyond.^92^ One study in this evidence review showed that the incidence of invasive breast cancer in women 50 to 59 is decreased when women are screened in their 40s,^92^ and another study by the same author reported a stage shift in women 50 to 59 when screening is available.^93^ The lifetime breast cancer mortality rate for unscreened women who develop breast cancer in their 40s is estimated at 5.8 per 1,000, compared to the 10-year estimate at 1.8 deaths per 1,000 in women who did not participate in breast cancer screening, suggesting that the mortality benefit from screening may be greater than what is captured in a ten year time frame.^47,125–127^ The 10-year duration may also contribute to the low estimated impact of screening on all-cause mortality (0.13 fewer deaths per 1,000 women in 10 years). Increasing the duration of follow-up to capture potential later effects of screening on breast cancer deaths over time will inevitably increase the amount of other-cause mortality which naturally occurs with aging.^128,129^

The morbidity associated with the treatment of any stage breast cancer is an important outcome which should be considered in addition to mortality when assessing benefits of screening. Generally, cancers that are detected at an early stage through screening can be treated with less-intensive therapies,.^130–134^ Examples include the ability to employ breast conserving surgery rather than mastectomy^135^, to avoid chemotherapy,^136,137^ or radiation therapy,^138^ and less need to perform axillary lymph node dissection.^133^ The equivalency of lumpectomy vs mastectomy was first established in 1985,^139^ so that prior to this date, mastectomy would have been standard of care. As such, the higher rates of mastectomy noted in the screened populations in the RCTs where screening occurred from 1963 and 1997 may have reflected treatment of cancer, as breast conserving surgery was generally not available. In the later observational trials, increasing rates of conservative surgery and decreasing rates of mastectomy were noted. It is important to recognize that currently in clinical practice, there has been continued de-escalation of treatment with rates of axillary node dissection declining and the use of genomic risk prediction tools that can help decrease the need for chemotherapy in both node-negative and node-positive hormone-receptor positive disease. However, high-risk early-stage cancers, such as HER2+ and triple negative breast cancers, may be treated aggressively given their elevated mortality risk.

The imaging recall rate remains high for breast cancer screening, with 15% of women with a first-time screen requiring follow-up imaging, and 7% of women in subsequent screens returning for further mammogram or ultrasound views.^52^ Recall rates are similar for all age groups.

### Strengths and limitations

Our systematic review adhered to established guidance for conducting and reporting systematic reviews, thus enhancing the credibility and robustness of our findings.^38^ This ensured transparency, consistency, and completeness throughout the review process. We conducted a thorough and systematic search of multiple databases with the help of an information specialist, as well as requesting other literature sources from stakeholders and the general public through an online portal. It is possible that we may have missed some studies after using artificial intelligence throughout the screening process, however our quality checks helped minimize this. Our review also included the input of a patient partner which strengthened the perspectives of our research and underscored the importance of incorporating patient perspectives in breast cancer screening recommendations.^140–142^ It may have been beneficial to have further patient representation from women who have engaged in discussions around breast screening but have not developed breast cancer.

The lack of evidence from screening after the year 2000 is such that the mortality benefits and harms of modern technologies and therapies may not be reflected in our results. Similarly, the 10-year time frame for absolute results may underreport mortality reduction and impacts on all-cause mortality. Additionally, the evidence from this review did not examine life-years saved as one of its outcomes which some may prefer considering when evaluating the benefits of screening.

### Implications for practice and policy

There is a need for more uniform breast cancer screening practices in Canada, and currently there are a variety of approaches with ten provinces/territories moving towards providing screening to women 40 years and older, while two remaining jurisdictions adhere to 2018 CTFPHC guidelines which recommended against routine mammography screening starting at age 40, however, suggested that a woman’s decision to screen may be based on their own values and preferences.^27^

Those developing guidelines or policies related to the evidence in this review should be aware of the limitations of the evidence base. There are no new RCTs available and only eight observational studies with screening that was initiated post 2000. The existing RCTs demonstrate a reduction in breast cancer mortality with screening, and the increased mortality benefit noted in the observational studies, may speak to the inclusion of only women are screened, as opposed to intent to screen, the evolution of technology and therapies over time, as well as the healthy user effect.

RCTs and the observational studies included in this evidence review do not reflect current clinical practices, the differing risks based on race or ethnicity,^4^ nor the benefits for lifetime risk reduction and should be interpreted accordingly. The calculations used to establish a baseline cancer incidence risk in an unscreened population were taken from the Pan-Canadian study (screening between 1990 and 2009)^47^ and recent increases in cancer incidence in women younger than 50 are not reflected in this number.

Additionally, our review did not capture evidence on the benefits or harms of breast cancer screening for those at moderately increased lifetime risk of breast cancer. In an attempt to gain an understanding of screening benefits or harms in these populations, we extrapolated the benefit of mammographic screening in an average risk population to the expected increased incidence of breast cancer in those at moderately increased lifetime risk due to family history or dense breasts. There are limitations with this approach, as screening mammography may be more or less effective in these groups than for an undifferentiated population. Further, the evidence for screening women at above average risk using supplemental screening with MRI or breast ultrasound was not evaluated in this key question, despite emerging evidence showing benefit.^143–146^ These studies show that the sensitivity of mammography decreases and the interval cancer rates increase with increasing breast tissue density^147^ and MRI in combination with mammogram diagnoses an additional 16 cancers per 1,000 women with extremely dense breasts who had previously normal mammograms. Additionally, the relative risk reduction which was applied to these greater risk populations was obtained from RCTs or observational trials of an undifferentiated general population which included individuals of average, intermediate, and high risk. When these trials were performed, no distinction was made for risk factors or breast tissue density.

### Implications for research

Definite high-quality studies are urgently needed to explore the benefits and harms of breast cancer screening with consideration to new screening technologies and with consideration of populations with known varied risks. We did not find any evidence supporting the benefits and harms of screening in individuals as a function of racial or ethnic groups, dense breasts, molecular subtypes, or those with family history. Although there is near consensus that ductal carcinoma in situ (DCIS) is the largest contributor to overdiagnosis,^148,149^ the extent to which this occurs is unclear due to the varying natural history of DCIS and the long follow up duration which would be required for trials to accurately determine this. Current model-based estimates suggest that DCIS develops into invasive breast cancer 30% to 60% of the time.^150^ A more definitive understanding of the biology of DCIS, and ongoing research around de-escalation of therapy for DCIS, is important to minimize overdiagnosis. Future evidence reviews will need to reconsider the premise of “screened vs unscreened”, as with a demonstrated mortality benefit with screening, it is likely unethical to withhold screening in future RCTs, which will limit the availability of new evidence.

### Ottawa ERSC patient partner perspective

The CTFPHC Working Group works with two ERSCs (Ottawa and Edmonton) to develop an analytic framework and key questions to guide its work towards new or updated clinical practice guidelines for cancer screening. When I was invited to work with the Ottawa ERSC as a patient partner, I readily agreed to lend my perspective, along with four clinical and scientific expert partners.

As a breast cancer survivor, peer supporter and advocate I felt I was a perfect candidate to provide an informed patient perspective and was encouraged that the CTFPHC stated: “patients are the ultimate end-user of healthcare findings and the most important stakeholder.”

The inclusion of patient partners is critical to the healthcare decision-making process in the future to ensure that outcomes are considered relevant to the patient. If science is to be presented in a way that is meaningful to patients and that incorporates their perspective as the end users, it needs to be done more transparently and with input across the process, not only within some components of the review.

For five months I attended meetings both with the Ottawa based ERSC and with the Task Force Breast Cancer Working Group, working on Key Question 1 dealing with the harms and benefits of screening. After that time period, my involvement was with the Ottawa ERSC only.

One of the biggest obstacles I faced was the steep learning curve involved in developing fluency not only on the process, but many of the technical presentations and the scientific jargon. Forest plots, data and grade tables were new to me and not readily understood from the outset. In future patient partnerships, an integral key to success for everyone involved would be an introductory workshop for patients-only to better integrate them and make the technical side of the process more accessible and understandable. Clinical experts and scientists arrive already well versed in these things. Patient partners typically do not. Hand in hand with proper pre-consultation process briefing, is the need for fully delineated terms of reference which might better lay out the roles and expectations for the patient partner.

That being said, in my role with the Ottawa ERSC I felt fully integrated and valued. Where some were looking at numbers, I was looking at women and real-life implications of screening. My input was welcomed and indeed heeded. For example, my feedback with respect to language helped us identify as misleading the term “false positives” and subsequently it was changed to “additional imaging”. This speaks to the simple reality that a patient’s interpretation of terminology is important and integral to scientific reviews that in the long run will become applicable to patients in the real world.

As a patient partner in this particular review, I was concerned by the inconsistency of patient involvement and the rather siloed approach of the CTFPHC review in general. I believe that true patient engagement comes only if patients are involved in every aspect of the entire review process. Lived experience can and should go hand-in-hand with research to ensure that study outcomes reflect the knowledge of patients themselves. If this is not done in each aspect of the review, potential gaps from a patient perspective will exist.

Involving patient partners as sought by the ERSC is welcome, and we recognize that this is a new experience. Moving forward, it is critical that patient and public involvement be embedded in such important review processes. Indeed, creating more opportunities for the involvement of patients as partners in all aspects of research will only continue to generate deeper understanding and presumably contribute to important and more well-rounded conclusions for the population as a whole.

## Conclusion

This systematic review update did not identify any new RCT data comparing breast cancer screening to no screening but found 26 observational studies not included in the 2017 review. As such, we present evidence other than RCTs which can contribute to the overall evidence on breast cancer screening. The increased appreciation of the benefits and harms of screening afforded by this review can be used to inform screening guideline development, although the overall low and very low certainty of evidence presents a significant challenge. Our findings highlight the decreasing number of RCTs that compare outcomes in screened versus unscreened women on this topic and reveal the need for further research in specific populations such as different ages, racial or ethnic groups, molecular subtypes, dense breasts, or family history.

## Declarations

### Ethics approval and consent to participate

Not applicable

### Consent for publication

All authors provided consent for publication.

### Availability of data and materials

Any data or materials can be found on the Open Science Framework (https://osf.io/xngsu/).

### Author’s contributions

AB was responsible for project administration, conceptualization, methodology, and contributed to all of aspects of the review. NS contributed to conceptualization, methodology, and participated in all aspects of the review. NV, FA, and RP performed screening, study selection, and risk of bias assessments. AD provided a patient partner perspective throughout the review. TK undertook the statistical analyses. BS developed the search strategy and provided methodological input. MY, AW, and JS provided methodological and clinical input throughout the review. JL contributed to funding acquisition. DM contributed to the conception and design of the review and provided methodological input at all phases of the review. All authors contributed to the critical review and editing of the original manuscript. All authors read and approved the final manuscript.

## Acknowledgements

We would like to acknowledge Dr. Stuart Nicholls for his invaluable contributions to the systematic review team on patient-oriented research as well as Dr. George Wells and Shannon Kelly for their methodological guidance on the statistical analysis. We also thank Kaitryn Campbell (St. Joseph’s Healthcare Hamilton/McMaster University) for peer review of the MEDLINE search strategy. The Canadian Task Force Breast Cancer Working Group assisted with developing the scope of the review and reviewing a draft of the current manuscript.

## References

1. Canadian Cancer Society / Société canadienne du. Breast cancer statistics [Internet]. Canadian Cancer Society. 2023 [cited 2024 Mar 21]. Available from: https://cancer.ca/en/cancer-information/cancer-types/breast/statistics

2. Brenner DR, Poirier A, Woods RR, Ellison LF, Billette JM, Demers AA, et al. Projected estimates of cancer in Canada in 2022. CMAJ [Internet]. 2022 May 2 [cited 2024 Jan 14];194(17):E601–7. Available from: https://www.cmaj.ca/content/194/17/e601

3. Nelson HD, Zakher B, Cantor A, Fu R, Griffin J, O’Meara ES, et al. Risk factors for breast cancer for women aged 40 to 49 years: a systematic review and meta-analysis. Ann Intern Med. 2012 May 1;156(9):635–48.

4. Stapleton SM, Oseni TO, Bababekov YJ, Hung YC, Chang DC. Race/ethnicity and age distribution of breast cancer diagnosis in the United States. JAMA surgery. 2018;153(6):594–5.

5. Lee CI, Chen LE, Elmore JG. Risk-based breast cancer screening: implications of breast density. Medical Clinics. 2017;101(4):725–41.

6. Boyd NF, Guo H, Martin LJ, Sun L, Stone J, Fishell E, et al. Mammographic Density and the Risk and Detection of Breast Cancer. N Engl J Med. 2007 Jan 18;356(3):227–36.

7. Lauby-Secretan Béatrice, Scoccianti Chiara, Loomis Dana, Benbrahim-Tallaa Lamia, Bouvard Véronique, Bianchini Franca, et al. Breast-Cancer Screening — Viewpoint of the IARC Working Group. New England Journal of Medicine. 2015;372(24):2353–8.

8. Nyström L, Andersson I, Bjurstam N, Frisell J, Nordenskjöld B, Rutqvist LE. Long-term effects of mammography screening: updated overview of the Swedish randomised trials. The Lancet. 2002;359(9310):909–19.

9. Nyström L, Bjurstam N, Jonsson H, Zackrisson S, Frisell J. Reduced breast cancer mortality after 20+ years of follow-up in the Swedish randomized controlled mammography trials in Malmö, Stockholm, and Göteborg. J Med Screen. 2017 Mar;24(1):34–42.

10. Miller AB, Wall C, Baines CJ, Sun P, To T, Narod SA. Twenty five year follow-up for breast cancer incidence and mortality of the Canadian National Breast Screening Study: randomised screening trial. BMJ. 2014 Feb 11;348(feb11 9):g366–g366.

11. Seely JM, Alhassan T. Screening for breast cancer in 2018—what should we be doing today? Curr Oncol [Internet]. 2018 Jun [cited 2024 Jan 14];25(Suppl 1):S115–24. Available from: https://www.ncbi.nlm.nih.gov/pmc/articles/PMC6001765/

12. IARC Working Group on the Evaluation of Cancer-Preventive. Breast Cancer Screening. IARC Handbooks of Cancer Prevention, Volume 15. 2016;

13. Nelson HD, Fu R, Cantor A, Pappas M, Daeges M, Humphrey L. Effectiveness of Breast Cancer Screening: Systematic Review and Meta-analysis to Update the 2009 U.S. Preventive Services Task Force Recommendation. 2016;164(4).

14. Salz T, Richman AR, Brewer NT. Meta-analyses of the effect of false-positive mammograms on generic and specific psychosocial outcomes. Psychooncology. 2010 Oct;19(10):1026–34.

15. Srivastava S, Koay EJ, Borowsky AD, De Marzo AM, Ghosh S, Wagner PD, et al. Cancer overdiagnosis: a biological challenge and clinical dilemma. Nat Rev Cancer [Internet]. 2019 Jun [cited 2024 Jan 14];19(6):349–58. Available from: https://www.ncbi.nlm.nih.gov/pmc/articles/PMC8819710/

16. Sardar P, Kundu A, Chatterjee S, Nohria A, Nairooz R, Bangalore S, et al. Long-term cardiovascular mortality after radiotherapy for breast cancer: A systematic review and meta-analysis. Clin Cardiol. 2017 Feb;40(2):73–81.

17. Miller AB, Baines CJ, To T, Wall C. Canadian National Breast Screening Study: 2. Breast cancer detection and death rates among women aged 50 to 59 years. CMAJ. 1992 Nov 15;147(10):1477–88.

18. Seely JM, Eby PR, Yaffe MJ. The Fundamental Flaws of the CNBSS Trials: A Scientific Review. Journal of Breast Imaging. 2022 Mar 1;4(2):108–19.

19. Yaffe MJ, Seely JM, Gordon PB, Appavoo S, Kopans DB. The randomized trial of mammography screening that was not—A cautionary tale. J Med Screen. 2022 Mar;29(1):7–11.

20. Duffy SW. Problems with the Canadian national breast screening studies [Internet]. Vol. 4, Journal of Breast Imaging. Oxford University Press US; 2022 [cited 2024 Apr 2]. p. 120–1. Available from: https://academic.oup.com/jbi/article-abstract/4/2/120/6555325

21. Boyd NF. The review of randomization in the Canadian National Breast Screening Study: Is the debate over? CMAJ. 1997 Jan 15;156(2):207–9.

22. Kopans DB. The Canadian National Breast Screening Studies are compromised and their results are unreliable. They should not factor into decisions about breast cancer screening. Breast Cancer Res Treat. 2017 Aug 1;165(1):9–15.

23. Keelan S, Flanagan M, Hill ADK. Evolving Trends in Surgical Management of Breast Cancer: An Analysis of 30 Years of Practice Changing Papers. Front Oncol. 2021;11:622621.

24. Hortobagyi GN. Breast Cancer: 45 Years of Research and Progress. JCO. 2020 Jul 20;38(21):2454–62.

25. Beau AB, Andersen PK, Vejborg I, Lynge E. Limitations in the Effect of Screening on Breast Cancer Mortality. JCO. 2018 Oct 20;36(30):2988–94.

26. Klarenbach S, Sims-Jones N, Lewin G, Singh H, Thériault G, Tonelli M, et al. Recommendations on screening for breast cancer in women aged 40–74 years who are not at increased risk for breast cancer. CMAJ. 2018 Dec 10;190(49):E1441–51.

27. My breast screening. [cited 2024 Apr 24]. Available from: https://mybreastscreening.ca/about/

28. U.S. Preventive Services Task Force. Draft Recommendation: Breast Cancer: Screening | United States Preventive Services Taskforce [Internet]. 2023 [cited 2023 Jun 22]. Available from: https://www.uspreventiveservicestaskforce.org/uspstf/draft-recommendation/breast-cancer-screening-adults

29. Marmot MG, Altman DG, Cameron DA, Dewar JA, Thompson SG, Wilcox M. The benefits and harms of breast cancer screening: an independent review. Br J Cancer. 2013 Jun 11;108(11):2205–40.

30. Cancer Registry of Norway. BreastScreen Norway [Internet]. 2023 [cited 2024 Apr 2]. Available from: https://www.kreftregisteret.no/en/screening/BreastScreen_Norway/breastscreen-norway/

31. Finish Cancer Registry. Breast cancer screening [Internet]. Syöpärekisteri. 2023 [cited 2024 Apr 2]. Available from: https://cancerregistry.fi/screening/breast-cancer-screening/

32. Danish Cancer Society. Kræftens Bekæmpelse [Internet]. 2024 [cited 2024 Apr 2]. Available from: https://www.cancer.dk/forebyg-kraeft/screening/brystkraeft/in-other-languages/

33. Lind H, Svane G, Kemetli L, Törnberg S. Breast Cancer Screening Program in Stockholm County, Sweden – Aspects of Organization and Quality Assurance. Breast Care (Basel). 2010 Oct;5(5):353–7.

34. Ottawa Knowledge Synthesis Group. Breast cancer screening: Part A. An evidence report to inform an update of the Canadian Task Force on Preventive Health Care 2011 Guideline [Internet]. 2018 [cited 2023 Jun 22]. Available from: https://canadiantaskforce.ca/wp-content/uploads/2019/02/Systematic-Review-Evidence-Report_v2_FINAL.pdf

35. Higgins J, Green S. Cochrane Handbook for Systematic Reviews of Interventions Version 5.1.0. [Internet] http://handbook.cochrane.org/.2011.

36. GRADE Working Group. Handbook for grading the quality of evidence and the strength of recommendations using the GRADE approach. 2013 Oct; Available from: https://gdt.gradepro.org/app/handbook/handbook.html

37. Canadian Task Force on Preventive Health Care. Canadian Task Force on Preventive Health Care Procedure Manual [Internet]. 2014. Available from: https://canadiantaskforce.ca/wp-content/uploads/2016/12/procedural-manual-en_2014_Archived.pdf

38. Page MJ, McKenzie JE, Bossuyt PM, Boutron I, Hoffmann TC, Mulrow CD, et al. The PRISMA 2020 statement: an updated guideline for reporting systematic reviews. BMJ. 2021;372.

39. Government of Canada CI of HR. Patient Partner Compensation Guidelines - CIHR [Internet]. 2022 [cited 2024 Feb 23]. Available from: https://cihr-irsc.gc.ca/e/53261.html

40. Riva JJ, Malik KMP, Burnie SJ, Endicott AR, Busse JW. Commentary What is your research question? An introduction to the PICOT format for clinicians.

41. Klarenbach S, Sims-Jones N, Lewin G, Singh H, Thériault G, Tonelli M, et al. Recommendations on screening for breast cancer in women aged 40–74 years who are not at increased risk for breast cancer. CMAJ [Internet]. 2018 Dec 10 [cited 2024 Jan 14];190(49):E1441–51. Available from: https://www.ncbi.nlm.nih.gov/pmc/articles/PMC6279444/

42. Siu AL, on behalf of the U.S. Preventive services task force. Screening for depression in children and adolescents: US Preventive Services Task Force recommendation statement. Ann Intern Med. 2016;164(5):360.

43. Sterne JAC, Savović J, Page MJ, Elbers RG, Blencowe NS, Boutron I, et al. RoB 2: a revised tool for assessing risk of bias in randomised trials. BMJ. 2019 Aug 28;366:l4898.

44. Joanna Briggs Institute. The Joanna Briggs Institute Critical Appraisal tools for use in JBI Systematic Reviews Checklist for Prevalence Studies [Internet]. Joanna Briggs Institute. 2017. Available from: https://jbi.global/sites/default/files/2019-05/JBI_Critical_Appraisal-Checklist_for_Prevalence_Studies2017_0.pdf

45. The Independent UK Panel on Breast Cancer Screening, Marmot MG, Altman DG, Cameron DA, Dewar JA, Thompson SG, et al. The benefits and harms of breast cancer screening: an independent review: A report jointly commissioned by Cancer Research UK and the Department of Health (England) October 2012. Br J Cancer. 2013 Jun;108(11):2205–40.

46. Skoetz N, Goldkuhle M, van Dalen EC, Akl EA, Trivella M, Mustafa RA, et al. GRADE guidelines 27: how to calculate absolute effects for time-to-event outcomes in summary of findings tables and Evidence Profiles. Journal of Clinical Epidemiology. 2020 Feb;118:124–31.

47. Coldman A, Phillips N, Wilson C, Decker K, Chiarelli AM, Brisson J, et al. Pan-Canadian study of mammography screening and mortality from breast cancer. 2014;106(11).

48. Deeks JJ, Higgins JPT, Altman DG. Chapter 10: Analysing data and undertaking meta-analyses. In: Cochrane Handbook for Systematic Reviews of Interventions version 62 (updated February 2021) [Internet]. 2021 [cited 2021 Mar 9]. Available from: https://training.cochrane.org/handbook/current/chapter-10

49. R Core Team. R: A language and environment for statistical computing. R Foundation for Statistical Computing, Vienna, Austria. [Internet]. 2021. Available from: https://www.R-project.org/

50. DerSimonian R, Laird N. Meta-analysis in clinical trials. Controlled clinical trials. 1986;7(3):177–88.

51. McKenzie JE, Brennan SE. Chapter 12: Synthesizing and presenting findings using other methods. In: Cochrane Handbook for Systematic Reviews of Interventions. 2023.

52. Canadian Partnership Against Cancer (CPAC). Breast Cancer Screening in Canada: Monitoring and Evaluation of Quality Indicators - Results Report, January 2011 to December 2012. Toronto, ON: Canadian Partnership Against Cancer; 2016.

53. Nova Scotia Breast Screening Program. Imaging Guidelines [Internet]. Nova Scotia Breast Cancer Screening Program. Available from: https://nsbreastscreening.ca/program-information/imaging-guidelines

54. Health PEI. PEI Breast Screening Program [Internet]. Government of Prince Edward Island. 2023. Available from: https://www.princeedwardisland.ca/en/information/health-pei/pei-breast-screening-program

55. Brodersen J, Schwartz LM, Heneghan C, O’Sullivan JW, Aronson JK, Woloshin S. Overdiagnosis: what it is and what it isn’t. BMJ EBM. 2018 Feb;23(1):1–3.

56. Biesheuvel C, Barratt A, Howard K, Houssami N, Irwig L. Effects of study methods and biases on estimates of invasive breast cancer overdetection with mammography screening: a systematic review. The lancet oncology. 2007;8(12):1129–38.

57. Puliti D, Bucchi L, Mancini S, Paci E, Baracco S, Campari C, et al. Advanced breast cancer rates in the epoch of service screening: The 400,000 women cohort study from Italy. European Journal of Cancer. 2017 Apr;75:109–16.

58. Carter JL, Coletti RJ, Harris RP. Quantifying and monitoring overdiagnosis in cancer screening: a systematic review of methods. BMJ. 2015 Jan 7;350(jan07 5):g7773–g7773.

59. Ripping TM, ten Haaf K, Verbeek ALM, van Ravesteyn NT, Broeders MJM. Quantifying Overdiagnosis in Cancer Screening: A Systematic Review to Evaluate the Methodology. JNCI: Journal of the National Cancer Institute [Internet]. 2017 Oct 1 [cited 2024 Apr 3];109(10). Available from: https://academic.oup.com/jnci/article/doi/10.1093/jnci/djx060/3845953

60. Egger M, Davey Smith G, Schneider M, Minder C. Bias in meta-analysis detected by a simple, graphical test. BMJ. 1997 Sep;315(7109):629–34.

61. Zeng L, Brignardello-Petersen R, Hultcrantz M, Mustafa RA, Murad MH, Iorio A, et al. GRADE Guidance 34: update on rating imprecision using a minimally contextualized approach. Journal of Clinical Epidemiology. 2022 Oct;150:216–24.

62. Schünemann HJ, Higgins JP, Vist GE, Glasziou P, Akl EA, Skoetz N, et al. Chapter 14: Completing ‘Summary of findings’ tables and grading the certainty of the evidence. In: Cochrane Handbook for Systematic Reviews of Interventions [Internet]. 2023. Available from: https://training.cochrane.org/handbook/current/chapter-14

63. Guyatt GH, Oxman AD, Sultan S, Glasziou P, Akl EA, Alonso-Coello P, et al. GRADE guidelines: 9. Rating up the quality of evidence. J Clin Epidemiol. 2011 Dec;64(12):1311–6.

64. Engmann NJ, Golmakani MK, Miglioretti DL, Sprague BL, Kerlikowske K. Population-Attributable Risk Proportion of Clinical Risk Factors for Breast Cancer. JAMA Oncol. 2017 Sep 1;3(9):1228–36.

65. Chiu SYH, Duffy S, Yen AMF, Tabár L, Smith RA, Chen HH. Effect of Baseline Breast Density on Breast Cancer Incidence, Stage, Mortality, and Screening Parameters: 25-Year Follow-up of a Swedish Mammographic Screening. Cancer Epidemiology, Biomarkers & Prevention. 2010 May 1;19(5):1219–28.

66. van der Waal D, Ripping TM, Verbeek ALM, Broeders MJM. Breast cancer screening effect across breast density strata: A case-control study: Screening effect across breast density strata. Int J Cancer. 2017 Jan 1;140(1):41–9.

67. Liu JL, Zhang SX, Billette JM, Demers AA. Lifetime probability of developing cancer and dying from cancer in Canada, 1997 to 2020. Health Rep. 2023 Sep 20;34(9):14–21.

68. Duffy SW, Vulkan D, Cuckle H, Parmar D, Sheikh S, Smith RA, et al. Effect of mammographic screening from age 40 years on breast cancer mortality (UK Age trial): final results of a randomised, controlled trial. The Lancet Oncology. 2020 Sep;21(9):1165–72.

69. Duffy S, Vulkan D, Cuckle H, Parmar D, Sheikh S, Smith R, et al. Annual mammographic screening to reduce breast cancer mortality in women from age 40 years: long-term follow-up of the UK Age RCT. Health Technol Assess. 2020 Oct;24(55):1–24.

70. Tarone RE. The excess of patients with advanced breast cancer in young women screened with mammography in the Canadian National Breast Screening Study. Cancer. 1995;75(4):997–1003.

71. Choi E, Jun JK, Suh M, Jung KW, Park B, Lee K, et al. Effectiveness of the Korean National Cancer Screening Program in reducing breast cancer mortality. NPJ breast cancer. 2021;7(1):83.

72. Duffy SW, Tabar L, Yen AMF, Dean PB, Smith RA, Jonsson H, et al. Beneficial Effect of Consecutive Screening Mammography Examinations on Mortality from Breast Cancer: A Prospective Study. Radiology. 2021;299(3):541–7.

73. Dunn N, Youl P, Moore J, Harden H, Walpole E, Evans E, et al. Breast-cancer mortality in screened versus unscreened women: Long-term results from a population-based study in Queensland, Australia. Journal of medical screening. 2021;28(2):193–9.

74. Morrell S, Taylor R, Roder D, Robson B, Gregory M, Craig K. Mammography service screening and breast cancer mortality in New Zealand: a National Cohort Study 1999–2011. Br J Cancer. 2017 Mar;116(6):828–39.

75. Lund E, Nakamura A, Thalabard JC. No overdiagnosis in the Norwegian Breast Cancer Screening Program estimated by combining record linkage and questionnaire information in the Norwegian Women and Cancer study. European Journal of Cancer. 2018 Jan;89:102–12.

76. Weedon-Fekjaer H, Romundstad PR, Vatten LJ. Modern mammography screening and breast cancer mortality: population study. BMJ (Clinical research ed). 2014;348:g3701.

77. Richman IB, Long JB, Soulos PR, Wang SY, Gross CP. Estimating Breast Cancer Overdiagnosis After Screening Mammography Among Older Women in the United States. Ann Intern Med. 2023 Sep 19;176(9):1172–80.

78. Garcia-Albeniz X, Hernan MA, Logan RW, Price M, Armstrong K, Hsu J. Continuation of Annual Screening Mammography and Breast Cancer Mortality in Women Older Than 70 Years. Annals of internal medicine. 2020;172(6):381–9.

79. Blyuss O, Dibden A, Massat NJ, Parmar D, Cuzick J, Duffy SW, et al. A case-control study to evaluate the impact of the breast screening programme on breast cancer incidence in England. Cancer medicine. 2023;12(2):1878–87.

80. De Troeyer K, Silversmit G, Rosskamp M, Truyen I, Van Herck K, Goossens MM, et al. The effect of the Flemish breast cancer screening program on breast cancer-specific mortality: A case-referent study. Cancer epidemiology. 2023;82:102320.

81. Maroni R, Massat NJ, Parmar D, Dibden A, Cuzick J, Sasieni PD, et al. A case-control study to evaluate the impact of the breast screening programme on mortality in England. Br J Cancer. 2021 Feb 16;124(4):736–43.

82. Massat NJ, Dibden A, Parmar D, Cuzick J, Sasieni PD, Duffy SW. Impact of Screening on Breast Cancer Mortality: The UK Program 20 Years On. 2016;25(3).

83. Beckmann KR, Lynch JW, Hiller JE, Farshid G, Houssami N, Duffy SW, et al. A novel case-control design to estimate the extent of over-diagnosis of breast cancer due to organised population-based mammography screening. 2015;136(6).

84. Pocobelli G, Weiss NS. Breast cancer mortality in relation to receipt of screening mammography: a case–control study in Saskatchewan, Canada. Cancer Causes Control. 2015 Feb;26(2):231–7.

85. Paap E, Verbeek ALM, Botterweck AAM, van Doorne-Nagtegaal HJ, Imhof-Tas M, e Koning HJ, et al. Breast cancer screening halves the risk of breast cancer death: a case-referent study. Breast (Edinburgh, Scotland). 2014;23(4):439–44.

86. Ripping TM, van der Waal D, Verbeek ALM, Broeders MJM. The relative effect of mammographic screening on breast cancer mortality by socioeconomic status. Medicine. 2016 Aug;95(31):e4335.

87. Hübner J, Katalinic A, Waldmann A, Kraywinkel K. Long-term Incidence and Mortality Trends for Breast Cancer in Germany. Geburtshilfe Frauenheilkd. 2020 Jun;80(06):611–8.

88. de Glas NA, e Craen AJM, Bastiaannet E, Op ’t Land EG, Kiderlen M, van de Water W, et al. Effect of implementation of the mass breast cancer screening programme in older women in the Netherlands: population based study. BMJ (Clinical research ed). 2014;349:g5410.

89. Parvinen I, Heinävaara S, Anttila A, Helenius H, Klemi P, Pylkkänen L. Mammography screening in three Finnish residential areas: comprehensive population-based study of breast cancer incidence and incidence-based mortality 1976–2009. British journal of cancer. 2015;112(5):918–24.

90. Helvie MA, Chang JT, Hendrick RE, Banerjee M. Reduction in late-stage breast cancer incidence in the mammography era: Implications for overdiagnosis of invasive cancer. Cancer. 2014 Sep;120(17):2649–56.

91. Tabár L, Dean PB, Chen TH, Yen AM, Chen SL, Fann JC, et al. The incidence of fatal breast cancer measures the increased effectiveness of therapy in women participating in mammography screening. Cancer. 2019 Feb 15;125(4):515–23.

92. Wilkinson AN, Ellison LF, Billette JM, Seely JM. Impact of Breast Cancer Screening on 10-Year Net Survival in Canadian Women Age 40-49 Years. J Clin Oncol. 2023 Oct 10;41(29):4669–77.

93. Wilkinson AN, Billette JM, Ellison LF, Killip MA, Islam N, Seely JM. The Impact of Organised Screening Programs on Breast Cancer Stage at Diagnosis for Canadian Women Aged 40–49 and 50–59. Current Oncology. 2022;29(8):5627–43.

94. Nystrom L, Bjurstam N, Jonsson H, Zackrisson S, Frisell J. Reduced breast cancer mortality after 20+ years of follow-up in the Swedish randomized controlled mammography trials in Malmo, Stockholm, and Goteborg. 2016;

95. Yen AM, Duffy SW, Chen TH, Chen L, Chiu SY, Fann JC, et al. Long-term incidence of breast cancer by trial arm in one county of the Swedish Two-County Trial of mammographic screening. Cancer. 2012 Dec;118(23):5728–32.

96. Tabar L, Chen THH, Hsu CY, Wu WYY, Yen AMF, Chen SLS, et al. Evaluation issues in the Swedish Two-County Trial of breast cancer screening: An historical review. 2016;

97. Tabár L, Vitak B, Chen THH, Yen AMF, Cohen A, Tot T, et al. Swedish Two-County Trial: Impact of Mammographic Screening on Breast Cancer Mortality during 3 Decades. Radiology. 2011 Sep;260(3):658–63.

98. Bjurstam N, Björneld L, Warwick J, Sala E, Duffy SW, Nyström L, et al. The Gothenburg Breast Screening Trial: The Gothenburg Breast Screening Trial. Cancer. 2003 May 15;97(10):2387–96.

99. Moss SM, Wale C, Smith R, Evans A, Cuckle H, Duffy SW. Effect of mammographic screening from age 40 years on breast cancer mortality in the UK Age trial at 17 years’ follow-up: a randomised controlled trial. 2015;16(9).

100. Shapiro S. Current results of the breast cancer screening randomized trial. The Health Insurance Plan (HIP) of Greater New York Study. Screening for breast cancer. 1988;3–15.

101. Baines CJ, To T, Miller AB. Revised estimates of overdiagnosis from the Canadian National Breast Screening Study. 2016;90.

102. Zackrisson S, Andersson I, Janzon L, Manjer J, Garne JP. Rate of over-diagnosis of breast cancer 15 years after end of Malmö mammographic screening trial: follow-up study. Bmj. 2006;332(7543):689–92.

103. Habbema JDF, Oortmarssen GJ van, van Putten DJ, Lubbe JT, Maas PJ van der. Age-specific reduction in breast cancer mortality by screening: an analysis of the results of the Health Insurance Plan of Greater New York study. Journal of the National Cancer Institute. 1986;77(2):317–20.

104. Katalinic A, Eisemann N, Kraywinkel K, Noftz MR, Hübner J. Breast cancer incidence and mortality before and after implementation of the German mammography screening program. Intl Journal of Cancer. 2020 Aug;147(3):709–18.

105. Goel V, Cohen MM, Kaufert P, MacWilliam L. Assessing the extent of contamination in the Canadian National Breast Screening Study. American Journal of Preventive Medicine. 1998 Oct;15(3):206–11.

106. Cohen MM, Kaufert PA, Macwilliam L, Tate RB. Using an alternative data source to examine randomization in the Canadian National Breast Screening Study. Journal of Clinical Epidemiology. 1996 Sep;49(9):1039–44.

107. Bailar JC, MacMahon B. Randomization in the Canadian National Breast Screening Study: a review for evidence of subversion. CAN MED ASSOC J.

108. Baines CJ. The Canadian National Breast Screening Study. Why? What next? And so what? Cancer. 1995 Nov 15;76(10 Suppl):2107–12.

109. Sterne, Hernán, McAleenan, Reeves, Higgins. Chapter 25: Assessing risk of bias in a non-randomized study. In: Cochrane Handbook for Systematic Reviews of Interventions version 64 [Internet]. Cochrane; 2023. Available from: www.training.cochrane.org/handbook.

110. Aron JL, Prorok PC. An analysis of the mortality effect in a breast cancer screening study. International journal of epidemiology. 1986;15(1):36–43.

111. Frisell J, Lidbrink E, Hellström L, Rutqvist LE. Followup after 11 years – update of mortality results in the Stockholm mammographic screening trial. Breast Cancer Res Treat. 1997 Sep;45(3):263–70.

112. Bjurstam NG, Bjorneld LM, Duffy SW. Updated results of the Gothenburg Trial of Mammographic Screening. 2016;122(12).

113. Gøtzsche PC, Jørgensen KJ. Screening for breast cancer with mammography. Cochrane database of systematic reviews [Internet]. 2013 [cited 2024 Feb 21];(6). Available from: https://www.cochranelibrary.com/cdsr/doi/10.1002/14651858.CD001877.pub5/abstract

114. Canadian Partnership Against Cancer. Breast Cancer Screening in Canada: Monitoring and Evaluation of Quality Indicators - Results Report, January 2011 to December 2012. Toronto: Canadian Partnership Against Cancer; 2016.

115. Seely JM, Ellison LF, Billette JM, Zhang SX, Wilkinson AN. Incidence of Breast Cancer in Younger Women: A Canadian Trend Analysis. Can Assoc Radiol J. 2024 Apr 25;08465371241246422.

116. Canadian Cancer Statistics Advisory Committee in collaboration with the Canadian Cancer Society, Statistics Canada and the Public Health Agency of Canada. Canadian Cancer Statistics 2023 [Internet]. 2023 [cited 2023 Nov 19]. Available from: cancer.ca/Canadian-Cancer-Statistics-2023-EN

117. Caswell-Jin JL, Sun LP, Munoz D, Lu Y, Li Y, Huang H, et al. Analysis of Breast Cancer Mortality in the US—1975 to 2019. JAMA. 2024;331(3):233–41.

118. Lofters AK, McBride ML, Li D, Whitehead M, Moineddin R, Jiang L, et al. Disparities in breast cancer diagnosis for immigrant women in Ontario and BC: results from the CanIMPACT study. BMC Cancer. 2019 Jan 9;19(1):42.

119. Schulz KF, Altman DG, Moher D, CONSORT Group. CONSORT 2010 statement: updated guidelines for reporting parallel group randomized trials. Ann Intern Med. 2010 Jun 1;152(11):726–32.

120. Altman DG. The Revised CONSORT Statement for Reporting Randomized Trials: Explanation and Elaboration. Ann Intern Med. 2001 Apr 17;134(8):663.

121. Soni PD, Hartman HE, Dess RT, Abugharib A, Allen SG, Feng FY, et al. Comparison of Population-Based Observational Studies With Randomized Trials in Oncology. JCO. 2019 May 10;37(14):1209–16.

122. Ma T, Semsarian CR, Barratt A, Parker L, Pathmanathan N, Nickel B, et al. Should low-risk DCIS lose the cancer label? An evidence review. Breast Cancer Res Treat. 2023 Jun;199(3):415–33.

123. Perou CM, Sørlie T, Eisen MB, Van De Rijn M, Jeffrey SS, Rees CA, et al. Molecular portraits of human breast tumours. nature. 2000;406(6797):747–52.

124. Plevritis SK, Munoz D, Kurian AW, Stout NK, Alagoz O, Near AM, et al. Association of screening and treatment with breast cancer mortality by molecular subtype in US women, 2000-2012. Jama. 2018;319(2):154–64.

125. Yaffe MJ, Mittmann N, Alagoz O, Trentham-Dietz A, Tosteson AN, Stout NK. The effect of mammography screening regimen on incidence-based breast cancer mortality. J Med Screen. 2018 Dec;25(4):197–204.

126. Oeffinger KC, Fontham ET, Etzioni R, Herzig A, Michaelson JS, Shih YCT, et al. Breast cancer screening for women at average risk: 2015 guideline update from the American Cancer Society. Jama. 2015;314(15):1599–614.

127. Government of Canada SC. Deaths and age-specific mortality rates, by selected grouped causes [Internet]. 2021 [cited 2024 Feb 29]. Available from: https://www150.statcan.gc.ca/t1/tbl1/en/tv.action?pid=1310039201

128. Yaffe MJ, Mainprize JG. The value of all-cause mortality as a metric for assessing breast cancer screening. JNCI: Journal of the National Cancer Institute. 2020;112(10):989–93.

129. Heijnsdijk EAM, Csanádi M, Gini A, Ten Haaf K, Bendes R, Anttila A, et al. All-cause mortality versus cancer-specific mortality as outcome in cancer screening trials: A review and modeling study. Cancer Medicine. 2019 Oct;8(13):6127–38.

130. Wilkinson AN, Seely JM, Rushton M, Williams P, Cordeiro E, Allard-Coutu A, et al. Capturing the true cost of breast cancer treatment: Molecular subtype and stage-specific per-case activity-based costing. Current Oncology. 2023;30(9):7860–73.

131. Barth RJ, Gibson GR, Carney PA, Mott LA, Becher RD, Poplack SP. Detection of Breast Cancer on Screening Mammography Allows Patients to Be Treated with Less-Toxic Therapy. American Journal of Roentgenology. 2005 Jan;184(1):324–9.

132. Ahn S, Wooster M, Valente C, Moshier E, Meng R, Pisapati K, et al. Impact of Screening Mammography on Treatment in Women Diagnosed with Breast Cancer. Ann Surg Oncol. 2018 Oct;25(10):2979–86.

133. Leung JWT. Screening Mammography Reduces Morbidity of Breast Cancer Treatment. American Journal of Roentgenology. 2005 May;184(5):1508–9.

134. Elder K, Nickson C, Pattanasri M, Cooke S, Machalek D, Rose A, et al. Treatment Intensity Differences After Early-Stage Breast Cancer (ESBC) Diagnosis Depending on Participation in a Screening Program. Ann Surg Oncol. 2018 Sep;25(9):2563–72.

135. Freedman GM, Anderson PR, Goldstein LJ, Hanlon AL, Cianfrocca ME, Millenson MM, et al. Routine mammography is associated with earlier stage disease and greater eligibility for breast conservation in breast carcinoma patients age 40 years and older. Cancer. 2003 Sep;98(5):918–25.

136. Coldman AJ, Phillips N, Speers C. A retrospective study of the effect of participation in screening mammography on the use of chemotherapy and breast conserving surgery. Intl Journal of Cancer. 2007 May 15;120(10):2185–90.

137. Sparano JA, Gray RJ, Makower DF, Pritchard KI, Albain KS, Hayes DF, et al. Adjuvant Chemotherapy Guided by a 21-Gene Expression Assay in Breast Cancer. N Engl J Med. 2018 Jul 12;379(2):111–21.

138. Whelan TJ, Smith S, Parpia S, Fyles AW, Bane A, Liu FF, et al. Omitting Radiotherapy after Breast-Conserving Surgery in Luminal A Breast Cancer. N Engl J Med. 2023 Aug 17;389(7):612–9.

139. Fisher B, Bauer M, Margolese R, Poisson R, Pilch Y, Redmond C, et al. Five-Year Results of a Randomized Clinical Trial Comparing Total Mastectomy and Segmental Mastectomy with or without Radiation in the Treatment of Breast Cancer. N Engl J Med. 1985 Mar 14;312(11):665–73.

140. Concannon TW, Fuster M, Saunders T, Patel K, Wong JB, Leslie LK, et al. A Systematic Review of Stakeholder Engagement in Comparative Effectiveness and Patient-Centered Outcomes Research. J GEN INTERN MED. 2014 Dec 1;29(12):1692–701.

141. Brett J, Staniszewska S, Mockford C, Herron-Marx S, Hughes J, Tysall C, et al. Mapping the impact of patient and public involvement on health and social care research: a systematic review. Health Expectations. 2014;17(5):637–50.

142. Forsythe LP, Carman KL, Szydlowski V, Fayish L, Davidson L, Hickam DH, et al. Patient Engagement In Research: Early Findings From The Patient-Centered Outcomes Research Institute. Health Affairs. 2019 Mar;38(3):359–67.

143. Kuhl CK, Strobel K, Bieling H, Leutner C, Schild HH, Schrading S. Supplemental Breast MR Imaging Screening of Women with Average Risk of Breast Cancer. Radiology. 2017 May 1;283(2):361–70.

144. Bakker MF, De Lange SV, Pijnappel RM, Mann RM, Peeters PHM, Monninkhof EM, et al. Supplemental MRI Screening for Women with Extremely Dense Breast Tissue. N Engl J Med. 2019 Nov 28;381(22):2091–102.

145. Veenhuizen SGA, De Lange SV, Bakker MF, Pijnappel RM, Mann RM, Monninkhof EM, et al. Supplemental Breast MRI for Women with Extremely Dense Breasts: Results of the Second Screening Round of the DENSE Trial. Radiology. 2021 May;299(2):278–86.

146. Ohuchi N, Suzuki A, Sobue T, Kawai M, Yamamoto S, Zheng YF, et al. Sensitivity and specificity of mammography and adjunctive ultrasonography to screen for breast cancer in the Japan Strategic Anti-cancer Randomized Trial (J-START): a randomised controlled trial. 2016;387(10016).

147. Kerlikowske K, Zhu W, Tosteson ANA, Sprague BL, Tice JA, Lehman CD, et al. Identifying Women With Dense Breasts at High Risk for Interval Cancer: A Cohort Study. Ann Intern Med. 2015 May 19;162(10):673–81.

148. Poelhekken K, Lin Y, Greuter MJ, van der Vegt B, Dorrius M, de Bock GH. The natural history of ductal carcinoma in situ (DCIS) in simulation models: A systematic review. The Breast [Internet]. 2023 [cited 2024 Feb 29]; Available from: https://www.sciencedirect.com/science/article/pii/S0960977623005350

149. Van Luijt PA, Heijnsdijk EAM, Fracheboud J, Overbeek LIH, Broeders MJM, Wesseling J, et al. The distribution of ductal carcinoma in situ (DCIS) grade in 4232 women and its impact on overdiagnosis in breast cancer screening. Breast Cancer Res. 2016 Dec;18(1):47.

150. Chootipongchaivat S, Van Ravesteyn NT, Li X, Huang H, Weedon-Fekjær H, Ryser MD, et al. Modeling the natural history of ductal carcinoma in situ based on population data. Breast Cancer Res. 2020 Dec;22(1):53.

151. Breast and Ovarian Cancer and Family History Risk Categories | CDC [Internet]. 2023 [cited 2023 Jul 18]. Available from: https://www.cdc.gov/genomics/disease/breast_ovarian_cancer/risk_categories.htm

152. Shapiro S, Venet W, Strax P. Current results of the breast cancer screening randomized trial: the health insurance plan (HIP) of greater New York study. Hans Huber. 1988;3–15.

153. Habbema J, Oortmarssen G, Putten D van, Lubbe J, Maas P. Age-specific reduction in breast cancer mortality by screening: an analysis of the results of the Health Insurance Plan of Greater New York study. JNCI: Journal of the National Cancer Institute. 1986;77(2):317–20.

154. Nyström L, Andersson I, Bjurstam N, Frisell J, Nordenskjöld B, Rutqvist LE. Long-term effects of mammography screening: updated overview of the Swedish randomised trials. The Lancet. 2002 Mar;359(9310):909–19.

155. Tabar L, Fagerberg G, Chen H, Duffy S, Smart C, Gad A. Efficacy of breast cancer screening by age. Cancer. 1995;75(10):2507–17.

156. Tabár L, Vitak B, Chen THH, Yen AMF, Cohen A, Tot T, et al. Swedish Two-County Trial: Impact of Mammographic Screening on Breast Cancer Mortality during 3 Decades. Radiology. 2011 Sep;260(3):658–63.

157. Moss SM, Wale C, Smith R, Evans A, Cuckle H, Duffy SW. Effect of mammographic screening from age 40 years on breast cancer mortality in the UK Age trial at 17 years’ follow-up: a randomised controlled trial. The Lancet Oncology. 2015;16(9):1123–32.

158. Massat NJ, Dibden A, Parmar D, Cuzick J, Sasieni PD, Duffy SW. Impact of Screening on Breast Cancer Mortality: The UK Program 20 Years On. Cancer epidemiology, biomarkers & prevention : a publication of the American Association for Cancer Research, cosponsored by the American Society of Preventive Oncology. 2016;25(3):455–62.

159. Beckmann KR, Lynch JW, Hiller JE, Farshid G, Houssami N, Duffy SW, et al. A novel case-control design to estimate the extent of over-diagnosis of breast cancer due to organised population-based mammography screening. International journal of cancer. 2015;136(6):1411–21.

160. Katalinic A, Eisemann N, Kraywinkel K, Noftz MR, Hubner J. Breast cancer incidence and mortality before and after implementation of the German mammography screening program. International journal of cancer. 2020;147(3):709–18.

161. Parvinen I, Heinavaara S, Anttila A, Helenius H, Klemi P, Pylkkanen L. Mammography screening in three Finnish residential areas: Comprehensive population-based study of breast cancer incidence and incidence-based mortality 1976-2009. British Journal of Cancer. 2015;112(5):918–24.

162. Helvie MA, Chang JT, Hendrick RE, Banerjee M. Reduction in late-stage breast cancer incidence in the mammography era: Implications for overdiagnosis of invasive cancer. Cancer. 2014;120(17):2649–56.

163. Coldman A, Phillips N, Wilson C, Decker K, Chiarelli AM, Brisson J, et al. Pan-Canadian Study of Mammography Screening and Mortality from Breast Cancer. JNCI: Journal of the National Cancer Institute [Internet]. 2014 Nov [cited 2023 Sep 11];106(11). Available from: https://academic.oup.com/jnci/article-lookup/doi/10.1093/jnci/dju261

