## Appendices for "Screening for breast cancer: A systematic review update to inform the Canadian Task Force on Preventive Health Care guideline"

#### Appendix 1 – PRISMA checklist

| Section and Topic | Item # | Checklist item | Location where item is reported |
|---|---|---|---|
| **TITLE** | | | |
| Title | 1 | Identify the report as a systematic review. | 1 |
| **ABSTRACT** | | | |
| Abstract | 2 | See the PRISMA 2020 for Abstracts checklist. | 3 |
| **INTRODUCTION** | | | |
| Rationale | 3 | Describe the rationale for the review in the context of existing knowledge. | 7 |
| Objectives | 4 | Provide an explicit statement of the objective(s) or question(s) the review addresses. | 7 |
| **METHODS** | | | |
| Eligibility criteria | 5 | Specify the inclusion and exclusion criteria for the review and how studies were grouped for the syntheses. | 9 |
| Information sources | 6 | Specify all databases, registers, websites, organisations, reference lists and other sources searched or consulted to identify studies. Specify the date when each source was last searched or consulted. | 11 |
| Search strategy | 7 | Present the full search strategies for all databases, registers, and websites, including any filters and limits used. | Appendix |
| Selection process | 8 | Specify the methods used to decide whether a study met the inclusion criteria of the review, including how many reviewers screened each record and each report retrieved, whether they worked independently, and if applicable, details of automation tools used in the process. | 11 |
| Data collection process | 9 | Specify the methods used to collect data from reports, including how many reviewers collected data from each report, whether they worked independently, any processes for obtaining or confirming data from study investigators, and if applicable, details of automation tools used in the process. | 12 |
| Data items | 10a | List and define all outcomes for which data were sought. Specify whether all results that were compatible with each outcome domain in each study were sought (e.g. for all measures, time points, analyses), and if not, the methods used to decide which results to collect. | 13 |
| | 10b | List and define all other variables for which data were sought (e.g. participant and intervention characteristics, funding sources). Describe any assumptions made about any missing or unclear information. | 13 |
| Study risk of bias assessment | 11 | Specify the methods used to assess risk of bias in the included studies, including details of the tool(s) used, how many reviewers assessed each study and whether they worked independently, and if applicable, details of automation tools used in the process. | 12 |
| Effect measures | 12 | Specify for each outcome the effect measure(s) (e.g. risk ratio, mean difference) used in the synthesis or presentation of results. | 13 |
| Synthesis methods | 13a | Describe the processes used to decide which studies were eligible for each synthesis (e.g. tabulating the study intervention characteristics and comparing against the planned groups for each synthesis (item #5)). | 14 |
| | 13b | Describe any methods required to prepare the data for presentation or synthesis, such as handling of missing summary statistics, or data conversions. | 14 |
| | 13c | Describe any methods used to tabulate or visually display results of individual studies and syntheses. | 14 |
| | 13d | Describe any methods used to synthesize results and provide a rationale for the choice(s). If meta-analysis was performed, describe the model(s), method(s) to identify the presence and extent of statistical heterogeneity, and software package(s) used. | 14 |

| Section and Topic | Item # | Checklist item | Location where item is reported |
|---|---|---|---|
| | 13e | Describe any methods used to explore possible causes of heterogeneity among study results (e.g. subgroup analysis, meta-regression). | 15 |
| | 13f | Describe any sensitivity analyses conducted to assess robustness of the synthesized results. | 15 |
| Reporting bias assessment | 14 | Describe any methods used to assess risk of bias due to missing results in a synthesis (arising from reporting biases). | 15 |
| Certainty assessment | 15 | Describe any methods used to assess certainty (or confidence) in the body of evidence for an outcome. | 13 |
| **RESULTS** | | | |
| Study selection | 16a | Describe the results of the search and selection process, from the number of records identified in the search to the number of studies included in the review, ideally using a flow diagram. | 20 |
| | 16b | Cite studies that might appear to meet the inclusion criteria, but which were excluded, and explain why they were excluded. | Appendix |
| Study characteristics | 17 | Cite each included study and present its characteristics. | 20 |
| Risk of bias in studies | 18 | Present assessments of risk of bias for each included study. | 22 |
| Results of individual studies | 19 | For all outcomes, present, for each study: (a) summary statistics for each group (where appropriate) and (b) an effect estimate and its precision (e.g. confidence/credible interval), ideally using structured tables or plots. | Appendix |
| Results of syntheses | 20a | For each synthesis, briefly summarise the characteristics and risk of bias among contributing studies. | 25-34 |
| | 20b | Present results of all statistical syntheses conducted. If meta-analysis was done, present for each the summary estimate and its precision (e.g. confidence/credible interval) and measures of statistical heterogeneity. If comparing groups, describe the direction of the effect. | 25-34 |
| | 20c | Present results of all investigations of possible causes of heterogeneity among study results. | 24 |
| | 20d | Present results of all sensitivity analyses conducted to assess the robustness of the synthesized results. | 24 |
| Reporting biases | 21 | Present assessments of risk of bias due to missing results (arising from reporting biases) for each synthesis assessed. | 22 |
| Certainty of evidence | 22 | Present assessments of certainty (or confidence) in the body of evidence for each outcome assessed. | 25-34 |
| **DISCUSSION** | | | |
| Discussion | 23a | Provide a general interpretation of the results in the context of other evidence. | 34 |
| | 23b | Discuss any limitations of the evidence included in the review. | 39 |
| | 23c | Discuss any limitations of the review processes used. | 39 |
| | 23d | Discuss implications of the results for practice, policy, and future research. | 39 |
| **OTHER INFORMATION** | | | |
| Registration and protocol | 24a | Provide registration information for the review, including register name and registration number, or state that the review was not registered. | 5 |
| | 24b | Indicate where the review protocol can be accessed, or state that a protocol was not prepared. | 5 |

| Section and Topic | Item # | Checklist item | Location where item is reported |
|---|---|---|---|
| | 24c | Describe and explain any amendments to information provided at registration or in the protocol. | 18 |
| Support | 25 | Describe sources of financial or non-financial support for the review, and the role of the funders or sponsors in the review. | 2 |
| Competing interests | 26 | Declare any competing interests of review authors. | 2 |
| Availability of data, code and other materials | 27 | Report which of the following are publicly available and where they can be found: template data collection forms; data extracted from included studies; data used for all analyses; analytic code; any other materials used in the review. | 2 |

*From:* Page MJ, McKenzie JE, Bossuyt PM, Boutron I, Hoffmann TC, Mulrow CD, et al. The PRISMA 2020 statement: an updated guideline for reporting systematic reviews. BMJ 2021;372:n71. doi: 10.1136/bmj.n71

For more information, visit: http://www.prisma-statement.org/

#### Appendix 2 - Database search strategy

CTFPHC – Breast Cancer – Harms & Benefits
Final Strategy
2023 Jul 8

Ovid Multifile

Database: Embase Classic+Embase <1947 to 2023 July 07>, Ovid MEDLINE(R) ALL <1946 to July 06, 2023>, EBM Reviews - Cochrane Central Register of Controlled Trials <June 2023>
Search Strategy:
--------------------------------------------------------------------------------
1 exp Breast Neoplasms/ (1031502)
2 ((breast* or mamma or mammar*) adj3 (cancer* or carcinoid* or carcinoma* or carcinogen* or adenocarcinoma* or adeno-carcinoma* or malignan* or neoplasia* or neoplasm* or sarcoma* or tumour* or tumor*)).tw,kw,kf. (1093944)
3 exp Carcinoma, Intraductal, Noninfiltrating/ (13607)
4 intraductal carcinoma*.tw,kw,kf. (3402)
5 (ductal carcinoma in situ or DCIS).tw,kw,kf. (26073)
6 or/1-5 [BREAST CANCER] (1309330)
7 exp Breast Neoplasms/di, pc (142043)
8 exp Mass Screening/ (464018)
9 screen*.tw,kw,kf. (2475388)
10 "Early Detection of Cancer"/ (51154)
11 ((early or earlier or earliest) adj3 (detect* or diagnos* or identif* or recogni*)).tw,kw,kf. (857194)
12 exp Breast Neoplasms/dg [diagnostic imaging] (28712)
13 exp Mammography/ (103519)
14 (mammograph* or mammogram*).tw,kw,kf. (90372)
15 exp Magnetic Resonance Imaging/ (1784668)
16 (fMRI or fMRIs or MRI or MRIs or NMRI or NMRIs or MR imaging or NMR imaging or magnetic resonance imag* or magnetic resonance tomograph* or MR tomograph*).tw,kw,kf. (1437177)
17 (chemical shift imaging or proton spin tomograph* or zeugmatograph*).tw,kw,kf. (2731)
18 (ultrasound* or ultrason* or echograph* or echotomograph* or echo-tomograph* or sonograph*).tw,kw,kf. (1304950)
19 (echomammogra* or echo-mammogra*).tw,kw,kf. (95)
20 Imaging, Three-Dimensional/ (183178)
21 ((3D or "3-D") adj3 imag*).tw,kw,kf. (70567)
22 (("3" or three) adj dimension* adj3 imag*).tw,kw,kf. (46838)
23 tomosynthes*.tw,kw,kf. (5567)
24 or/7-23 [SCREENING] (6686083)
25 6 and 24 [BREAST CANCER SCREENING] (327407)
26 exp Infant/ not (exp Adult/ and exp Infant/) (1984574)
27 exp Child/ not (exp Adult/ and exp Child/) (4012787)
28 Adolescent/ not (exp Adult/ and Adolescent/) (1432274)

29 or/26-28 (5072306)
30 25 not 29 [CHILD-ONLY REMOVED] (325404)
31 exp Animals/ not (exp Animals/ and Humans/) (17644214)
32 30 not 31 [ANIMAL-ONLY REMOVED] (272302)
33 (comment or editorial or news or newspaper article).pt. (2472356)
34 (letter not (letter and randomized controlled trial)).pt. (2543284)
35 32 not (33 or 34) [OPINION PIECES REMOVED] (257499)
36 (Case Reports not (Case Reports and Randomized Controlled Trial)).pt. (2343931)
37 (case adj (series or study or studies or report or reports)).ti. (955591)
38 35 not (36 or 37) [CASE STUDIES/SERIES REMOVED] (243638)
39 limit 38 to yr="2014-current" (114748)
40 (controlled clinical trial or randomized controlled trial or pragmatic clinical trial).pt. (687633)
41 clinical trials as topic.sh. (238023)
42 (randomi#ed or randomly or RCT$1 or placebo*).tw. (4183310)
43 ((singl* or doubl* or trebl* or tripl*) adj (mask* or blind* or dumm*)).tw. (806676)
44 trial.ti. (1110226)
45 or/40-44 (4818073)
46 39 and 45 [RCTs] (10185)
47 controlled clinical trial.pt. (95362)
48 Controlled Clinical Trial/ or Controlled Clinical Trials as Topic/ (600612)
49 (control* adj2 trial).tw,kw,kf. (1180360)
50 Non-Randomized Controlled Trials as Topic/ (14974)
51 (nonrandom* or non-random* or quasi-random* or quasi-experiment*).tw,kw,kf. (186551)
52 (nRCT or non-RCT).tw,kw,kf. (1327)
53 Controlled Before-After Studies/ (252993)
54 (control* adj3 ("before and after" or "before after")).tw,kw,kf. (14017)
55 Interrupted Time Series Analysis/ (246419)
56 time series.tw,kw,kf. (100272)
57 (pre- adj5 post-).tw,kw,kf. (408710)
58 ((pretest adj5 posttest) or (pre-test adj5 post-test)).tw,kw,kf. (39051)
59 Historically Controlled Study/ (263364)
60 (control* adj2 study).tw,kw,kf. (1072731)
61 Control Groups/ (126139)
62 (control* adj2 group?).tw,kw,kf. (1818179)
63 trial.ti. (1110226)
64 or/47-63 (4979231)
65 39 and 64 [nRCTs] (11142)
66 exp Cohort Studies/ (3738436)
67 cohort?.tw,kw,kf. (2424497)
68 Retrospective Studies/ (2360506)
69 (longitudinal or prospective or retrospective).tw,kw,kf. (4620303)
70 ((followup or follow-up) adj (study or studies)).tw,kw,kf. (151254)

71 Observational study.pt. (143681)
72 (observation$2 adj (study or studies)).tw,kw,kf. (441671)
73 ((population or population-based) adj (study or studies or analys#s)).tw,kw,kf. (60358)
74 ((multidimensional or multi-dimensional) adj (study or studies)).tw,kw,kf. (354)
75 Comparative Study.pt. (1912764)
76 ((comparative or comparison) adj (study or studies)).tw,kw,kf. (360187)
77 exp Case-Control Studies/ (1677569)
78 ((case-control* or case-based or case-comparison or case-compeer or case-referrent or case-referent) adj3 (study or studies)).tw,kw,kf. (337507)
79 Multicenter Study.pt. (335633)
80 ((multicenter or multi-center or multicentre or multi-centre) adj (study or studies)).tw,kw,kf. (229129)
81 or/66-80 (10499257)
82 39 and 81 [OBSERVATIONAL STUDIES] (39698)
83 46 or 65 or 82 [RCTs, nRCTs, OBSERVATIONAL STUDIES, 2014-PRESENT] (47568)
84 exp Mass Screening/ae [Adverse Effects] (1118)
85 exp Mass Screening/mo [Mortality] (85)
86 "Early Detection of Cancer"/ae [Adverse Effects] (361)
87 "Early Detection of Cancer"/mo [Mortality] (99)
88 exp Mammography/ae [Adverse Effects] (965)
89 exp Mammography/mo [Mortality] (26)
90 exp Diagnostic Errors/ (256455)
91 exp Neoplasms, Radiation-Induced/ (22415)
92 Mortality/ (1016691)
93 ((avoid* or declin* or decreas* or lessen* or lower* or prevent* or reduc*) adj5 (death* or fatal* or morbidit* or mortalit*)).tw,kw,kf. (748202)
94 ((avoid* or declin* or decreas* or lessen* or lower* or prevent* or reduc*) adj5 (advanced stage? or (advanc* adj3 cancer?) or biopsy or biopsies or chemotherap* or chemo-therap* or disfigur* or exacerbat* or incidental finding? or progress* stage? or progression* or late? stage? or stage? III or stage? 3 or stage? IV or stage? 4)).tw,kw,kf. (282238)
95 ((avoid* or declin* or decreas* or lessen* or lower* or prevent* or reduc*) adj5 (adverse* or harm* or impair* or injur* or invasiv* or side-effect* or sideeffect* or undesirabl* or un-desirabl*)).tw,kw,kf. (718080)
96 ((avoid* or declin* or decreas* or lessen* or lower* or prevent* or reduc*) adj5 (alarm* or anxiet* or anxious* or distress* or emotion* or feeling* or psycholog* or uncertain* or un-certain*)).tw,kw,kf. (242773)
97 (misdiagnos* or mis-diagnos* or misdetect* or mis-detect* or misidentif* or mis-identif*).tw,kw,kf. (124301)
98 (miss$3 adj3 (detect* or diagnos* or identif*)).tw,kw,kf. (34711)
99 ((undetected or un-detected or ("not" adj3 detect*) or undiagnos* or un-diagnos* or ("not" adj3 diagnos*) or unidentif* or un-identif* or ("not" adj3 identif*)) adj3 (cancer* or carcinoid* or carcinoma* or carcinogen* or adenocarcinoma* or adeno-carcinoma* or lump or lumps or malignan* or neoplasia* or neoplasm* or sarcoma* or tumour* or tumor*)).tw,kw,kf. (21197)
100 (overdiagnos* or over diagnos*).tw,kw,kf. (17948)
101 (false adj (negative* or positive*)).tw,kw,kf. (223512)
102 ((error* or false$2 or wrong$3) adj3 (alarm* or detect* or diagnos*)).tw,kw,kf. (71561)
103 exp Medical Overuse/ (23414)
104 overtreat*.tw,kw,kf. (17805)
105 ((medical or health service? or procedur* or therap* or treatment*) adj3 (overuse? or overusing or overutilis* or overutiliz*)).tw,kw,kf. (2405)

106 ((inappropriate* or unnecessar*) adj3 (followup or follow-up or health care or healthcare or procedur* or therap* or treatment*)).tw,kw,kf. (44277)
107 (inappropriate* or unnecessar* or safe or adverse or adversely or undesirabl* or un-desirabl* or unintend* or un-intend* or unintent* or un-intent* or unsafe* or un-safe* or unwanted or un-wanted or harm* or injurious* or risk or risks or side-effect* or sideeffect* or reaction* or complication*).ti,kw,kf. (3614192)
108 ((adverse* or undesirabl* or un-desirabl* or unintend* or un-intend* or unintent* or un-intent* or unwanted or un-wanted or harm* or toxic or injurious* or serious* or fatal) adj5 (affect or affected or affecting or affects or consequence* or effect* or react or reacts or reacted or reacting or reaction* or side-effect* or sideeffect* or event* or outcome* or incident*)).tw,kw,kf. (2394363)
109 ((adverse* or inappropriat* or unnecessar* or undesirabl* or un-desirabl* or unintend* or un-intend* or unintent* or un-intent* or unwanted or un-wanted or injurious* or serious*) adj5 (alarm* or anxiet* or anxious* or distress* or emotion* or feeling* or psycholog* or uncertain* or un-certain*)).tw,kw,kf. (34213)
110 exp Neoplasm Metastasis/ and (avoid* or declin* or decreas* or lessen* or lower* or prevent* or reduc*).ti. (16969)
111 (benefit* or beneficial*).ti,kw,kf. (244174)
112 or/84-111 [BENEFITS/HARMS] (8596924)
113 83 and 112 [nRCTs, OBSERVATIONAL STUDIES, 2014-PRESENT - BENEFITS AND HARMS] (15099)
114 113 use medall [MEDLINE RECORDS] (5656)
115 exp breast cancer/ (942993)
116 ((breast* or mamma or mammar*) adj3 (cancer* or carcinoid* or carcinoma* or carcinogen* or adenocarcinoma* or adeno-carcinoma* or malignan* or neoplasia* or neoplasm* or sarcoma* or tumour* or tumor*)).ti,kw,kf. (753136)
117 intraductal carcinoma*.ti,kw,kf. (1516)
118 (ductal carcinoma in situ or DCIS).ti,kw,kf. (10454)
119 or/115-118 [BREAST CANCER] (1108734)
120 exp breast cancer/di, pc [diagnosis, prevention] (123365)
121 mass screening/ or cancer screening/ (311545)
122 screen*.ti,kw,kf. (621693)
123 early cancer diagnosis/ (13353)
124 ((early or earlier or earliest) adj3 (detect* or diagnos* or identif* or recogni*)).ti,kw,kf. (116845)
125 exp mammography/ (103519)
126 (mammograph* or mammogram*).ti,kw,kf. (46531)
127 breast magnetic resonance imaging/ (614)
128 (fMRI or fMRIs or MRI or MRIs or NMRI or NMRIs or MR imaging or NMR imaging or magnetic resonance imag* or magnetic resonance tomograph* or MR tomograph*).ti,kw,kf. (630153)
129 (chemical shift imaging or proton spin tomograph* or zeugmatograph*).ti,kw,kf. (1194)
130 (ultrasound* or ultrason* or echograph* or echotomograph* or echo-tomograph* or sonograph*).ti,kw,kf. (603624)
131 (echomammogra* or echo-mammogra*).ti,kw,kf. (73)
132 three-dimensional imaging/ (197470)
133 ((3D or "3-D") adj3 imag*).ti,kw,kf. (14230)
134 (("3" or three) adj dimension* adj3 imag*).ti,kw,kf. (13339)
135 tomosynthes*.ti,kw,kf. (4357)
136 or/120-135 [SCREENING] (2375020)
137 119 and 136 [BREAST CANCER SCREENING] (204028)
138 male/ not female/ (6570826)

139 137 not 138 [MALE-ONLY REMOVED] (201333)
140 exp adolescent/ not exp adult/ (1432463)
141 exp child/ not exp adult/ (4012787)
142 or/140-141 (4551929)
143 139 not 142 [CHILD-ONLY REMOVED] (200552)
144 (exp animal/ or exp animal experimentation/ or exp animal model/ or exp animal experiment/ or nonhuman/ or exp vertebrate/) not (exp human/ or exp human experimentation/ or exp human experiment/) (13235119)
145 143 not 144 [ANIMAL-ONLY REMOVED] (197599)
146 editorial.pt. or (letter.pt. not randomized controlled trial/) (3978550)
147 conference abstract.pt. (4813001)
148 145 not (146 or 147) [OPINION PIECES, CONFERENCE ABSTRACTS REMOVED] (175534)
149 (case report/ or exp case study/) not randomized controlled trial/ (5444300)
150 (case adj (series or study or studies or report or reports)).ti. (955591)
151 148 not (149 or 150) [CASE STUDIES/SERIES REMOVED] (160868)
152 limit 151 to yr="2014-current" [DATE LIMITS APPLICABLE TO OBSERVATIONAL STUDY SEARCH] (67507)
153 exp randomized controlled trial/ or controlled clinical trial/ (1716372)
154 clinical trial/ (1656994)
155 exp "controlled clinical trial (topic)"/ (274165)
156 (randomi#ed or randomi#ation? or randomly or RCT? or placebo*).ti,kw,kf. (1511469)
157 ((singl* or doubl* or trebl* or tripl*) adj (mask* or blind* or dumm*)).ti,kw,kf. (323173)
158 trial.ti. (1110226)
159 or/153-158 (4044688)
160 152 and 159 [RCTs] (5131)
161 controlled clinical trial/ (582481)
162 "controlled clinical trial (topic)"/ (14151)
163 (control* adj2 trial).ti,kw,kf. (875614)
164 (nonrandom* or non-random* or quasi-random* or quasi-experiment*).ti,kw,kf. (25241)
165 (nRCT or non-RCT).ti,kw,kf. (23)
166 (control* adj3 ("before and after" or "before after")).ti,kw,kf. (872)
167 time series analysis/ (38799)
168 time series.ti,kw,kf. (26963)
169 pretest posttest control group design/ (707)
170 (pre- adj5 post-).ti,kw,kf. (19926)
171 ((pretest adj5 posttest) or (pre-test adj5 post-test)).ti,kw,kf. (7574)
172 controlled study/ (10136653)
173 (control* adj2 study).ti,kw,kf. (688045)
174 control group/ (130398)
175 (control* adj2 group?).ti,kw,kf. (27359)
176 trial.ti. (1110226)
177 or/155-170 (4151332)
178 152 and 177 [nRCTs] (5316)
179 cohort analysis/ (1389236)

180 cohort?.ti,kw,kf. (472993)
181 retrospective study/ (2627466)
182 longitudinal study/ (363170)
183 prospective study/ (1559366)
184 (longitudinal or prospective or retrospective).ti,kw,kf. (1067953)
185 follow up/ (2132289)
186 ((followup or follow-up) adj (study or studies)).ti,kw,kf. (62193)
187 observational study/ (480181)
188 (observation$2 adj (study or studies)).ti,kw,kf. (102330)
189 population research/ (134718)
190 ((population or population-based) adj (study or studies or analys#s)).ti,kw,kf. (22545)
191 ((multidimensional or multi-dimensional) adj (study or studies)).ti,kw,kf. (139)
192 exp comparative study/ (3627289)
193 ((comparative or comparison) adj (study or studies)).ti,kw,kf. (223839)
194 exp case control study/ (1677569)
195 ((case-control* or case-based or case-comparison or case-compeer or case-referrent or case-referent) adj3 (study or studies)).ti,kw,kf. (107167)
196 major clinical study/ (5118942)
197 multicenter study/ (725450)
198 ((multicenter or multi-center or multicentre or multi-centre) adj (study or studies)).ti,kw,kf. (127846)
199 or/179-198 (13854212)
200 152 and 199 [OBSERVATIONAL STUDIES] (32240)
201 160 or 178 or 200 [RCTs, nRCTs, OBSERVATIONAL STUDIES, 2014-PRESENT] (34489)
202 mass screening/ae [adverse drug reaction] (971)
203 exp mammography/ae [adverse drug reaction] (965)
204 exp diagnostic error/ (256455)
205 mortality/ (1016691)
206 cancer mortality/ (108053)
207 exp radiation induced neoplasm/ (22415)
208 ((avoid* or declin* or decreas* or lessen* or lower* or prevent* or reduc*) adj5 (death* or fatal* or morbidit* or mortalit*)).ti,kw,kf. (51730)
209 ((avoid* or declin* or decreas* or lessen* or lower* or prevent* or reduc*) adj5 (advanced stage? or (advanc* adj3 cancer?) or biopsy or biopsies or chemotherap* or chemo-therap* or disfigur* or exacerbat* or incidental finding? or progress* stage? or progression* or late? stage? or stage? III or stage? 3 or stage? IV or stage? 4)).ti,kw,kf. (24097)
210 ((avoid* or declin* or decreas* or lessen* or lower* or prevent* or reduc*) adj5 (adverse* or harm* or impair* or injur* or invasiv* or side-effect* or sideeffect* or undesirabl* or un-desirabl*)).ti,kw,kf. (89663)
211 ((avoid* or declin* or decreas* or lessen* or lower* or prevent* or reduc*) adj5 (alarm* or anxiet* or anxious* or distress* or emotion* or feeling* or psycholog* or uncertain* or un-certain*)).ti,kw,kf. (22890)
212 (misdiagnos* or mis-diagnos* or misdetect* or mis-detect* or misidentif* or mis-identif*).ti,kw,kf. (18034)
213 (miss$3 adj3 (detect* or diagnos* or identif*)).ti,kw,kf. (4039)
214 ((undetected or un-detected or ("not" adj3 detect*) or undiagnos* or un-diagnos* or ("not" adj3 diagnos*) or unidentif* or un-identif* or ("not" adj3 identif*)) adj3 (cancer* or carcinoid* or carcinoma* or carcinogen* or adenocarcinoma* or adeno-carcinoma* or lump or lumps or malignan* or neoplasia* or neoplasm* or sarcoma* or tumour* or tumor*)).ti,kw,kf. (1185)

215 (overdiagnos* or over diagnos*).ti,kw,kf. (4106)
216 (false adj (negative* or positive*)).ti,kw,kf. (21412)
217 ((error* or false$2 or wrong$3) adj3 (alarm* or detect* or diagnos*)).ti,kw,kf. (13968)
218 exp medical overuse/ (23414)
219 overtreat*.ti,kw,kf. (2587)
220 ((medical or health service? or procedur* or therap* or treatment*) adj3 (overuse? or overusing or overutilis* or overutiliz*)).ti,kw,kf. (815)
221 ((inappropriate* or unnecessar*) adj3 (followup or follow-up or health care or healthcare or procedur* or therap* or treatment*)).ti,kw,kf. (2472)
222 (inappropriate* or unnecessar* or safe or adverse or adversely or undesirabl* or un-desirabl* or unintend* or un-intend* or unintent* or un-intent* or unsafe* or un-safe* or unwanted or un-wanted or harm* or injurious* or risk or risks or side-effect* or sideeffect* or reaction* or complication*).ti,kw,kf. (3614192)
223 ((adverse* or undesirabl* or un-desirabl* or unintend* or un-intend* or unintent* or un-intent* or unwanted or un-wanted or harm* or toxic or injurious* or serious* or fatal) adj5 (affect or affected or affecting or affects or consequence* or effect* or react or reacts or reacted or reacting or reaction* or side-effect* or sideeffect* or event* or outcome* or incident*)).ti,kw,kf. (300180)
224 ((adverse* or inappropriat* or unnecessar* or undesirabl* or un-desirabl* or unintend* or un-intend* or unintent* or un-intent* or unwanted or un-wanted or injurious* or serious*) adj5 (alarm* or anxiet* or anxious* or distress* or emotion* or feeling* or psycholog* or uncertain* or un-certain*)).ti,kw,kf. (3394)
225 exp metastasis/ and (avoid* or declin* or decreas* or lessen* or lower* or prevent* or reduc*).ti. (16969)
226 (benefit* or beneficial*).ti,kw,kf. (244174)
227 or/202-226 [BENEFITS/HARMS] (5283049)
228 201 and 227 [RCTs, nRCTs, OBSERVATIONAL STUDIES, 2014-PRESENT - BENEFITS AND HARMS] (7917)
229 228 use emczd [EMBASE RECORDS] (5434)
230 exp Breast Neoplasms/ (1031502)
231 ((breast* or mamma or mammar*) adj3 (cancer* or carcinoid* or carcinoma* or carcinogen* or adenocarcinoma* or adeno-carcinoma* or malignan* or neoplasia* or neoplasm* or sarcoma* or tumour* or tumor*)).ti,ab,kw. (1079601)
232 exp Carcinoma, Intraductal, Noninfiltrating/ (13607)
233 intraductal carcinoma*.ti,ab,kw. (3376)
234 (ductal carcinoma in situ or DCIS).ti,ab,kw. (26006)
235 or/230-234 [BREAST CANCER] (1303998)
236 exp Breast Neoplasms/di, pc (142043)
237 exp Mass Screening/ (464018)
238 screen*.ti,ab,kw. (2457803)
239 "Early Detection of Cancer"/ (51154)
240 ((early or earlier or earliest) adj3 (detect* or diagnos* or identif* or recogni*)).ti,ab,kw. (849461)
241 exp Breast Neoplasms/dg [diagnostic imaging] (28712)
242 exp Mammography/ (103519)
243 (mammograph* or mammogram*).ti,ab,kw. (90191)
244 exp Magnetic Resonance Imaging/ (1784668)
245 (fMRI or fMRIs or MRI or MRIs or NMRI or NMRIs or MR imaging or NMR imaging or magnetic resonance imag* or magnetic resonance tomograph* or MR tomograph*).ti,ab,kw. (1421280)
246 (chemical shift imaging or proton spin tomograph* or zeugmatograph*).ti,ab,kw. (2696)
247 (ultrasound* or ultrason* or echograph* or echotomograph* or echo-tomograph* or sonograph*).ti,ab,kw. (1295110)

248 (echomammogra* or echo-mammogra*).ti,ab,kw. (95)
249 Imaging, Three-Dimensional/ (183178)
250 ((3D or "3-D") adj3 imag*).ti,ab,kw. (68464)
251 (("3" or three) adj dimension* adj3 imag*).ti,ab,kw. (44442)
252 tomosynthes*.ti,ab,kw. (5513)
253 or/236-252 [SCREENING] (6654665)
254 235 and 253 [BREAST CANCER SCREENING] (325365)
255 exp Infant/ not (exp Adult/ and exp Infant/) (1984574)
256 exp Child/ not (exp Adult/ and exp Child/) (4012787)
257 Adolescent/ not (exp Adult/ and Adolescent/) (1432274)
258 or/255-257 (5072306)
259 254 not 258 [CHILD-ONLY REMOVED] (323374)
260 conference proceeding.pt. (222693)
261 259 not 260 [CONFERENCE ABSTRACTS REMOVED] (322140)
262 limit 261 to yr="2014-current" (148543)
263 exp Mass Screening/ae [Adverse Effects] (1118)
264 exp Mass Screening/mo [Mortality] (85)
265 "Early Detection of Cancer"/ae [Adverse Effects] (361)
266 "Early Detection of Cancer"/mo [Mortality] (99)
267 exp Mammography/ae [Adverse Effects] (965)
268 exp Mammography/mo [Mortality] (26)
269 exp Diagnostic Errors/ (256455)
270 exp Neoplasms, Radiation-Induced/ (22415)
271 Mortality/ (1016691)
272 ((avoid* or declin* or decreas* or lessen* or lower* or prevent* or reduc*) adj5 (death* or fatal* or morbidit* or mortalit*)).ti,ab,kw. (747649)
273 ((avoid* or declin* or decreas* or lessen* or lower* or prevent* or reduc*) adj5 (advanced stage? or (advanc* adj3 cancer?) or biopsy or biopsies or chemotherap* or chemo-therap* or disfigur* or exacerbat* or incidental finding? or progress* stage? or progression* or late? stage? or stage? III or stage? 3 or stage? IV or stage? 4)).ti,ab,kw. (282054)
274 ((avoid* or declin* or decreas* or lessen* or lower* or prevent* or reduc*) adj5 (adverse* or harm* or impair* or injur* or invasiv* or side-effect* or sideeffect* or undesirabl* or un-desirabl*)).ti,ab,kw. (712155)
275 ((avoid* or declin* or decreas* or lessen* or lower* or prevent* or reduc*) adj5 (alarm* or anxiet* or anxious* or distress* or emotion* or feeling* or psycholog* or uncertain* or un-certain*)).ti,ab,kw. (242462)
276 (misdiagnos* or mis-diagnos* or misdetect* or mis-detect* or misidentif* or mis-identif*).ti,ab,kw. (124214)
277 (miss$3 adj3 (detect* or diagnos* or identif*)).ti,ab,kw. (34526)
278 ((undetected or un-detected or ("not" adj3 detect*) or undiagnos* or un-diagnos* or ("not" adj3 diagnos*) or unidentif* or un-identif* or ("not" adj3 identif*)) adj3 (cancer* or carcinoid* or carcinoma* or carcinogen* or adenocarcinoma* or adeno-carcinoma* or lump or lumps or malignan* or neoplasia* or neoplasm* or sarcoma* or tumour* or tumor*)).ti,ab,kw. (21184)
279 (overdiagnos* or over diagnos*).ti,ab,kw. (17939)
280 (false adj (negative* or positive*)).ti,ab,kw. (222230)
281 ((error* or false$2 or wrong$3) adj3 (alarm* or detect* or diagnos*)).ti,ab,kw. (69859)
282 exp Medical Overuse/ (23414)
283 overtreat*.tw,kw,kf. (17805)

284 ((medical or health service? or procedur* or therap* or treatment*) adj3 (overuse? or overusing or overutilis* or overutiliz*)).ti,ab,kw. (2132)
285 ((inappropriate* or unnecessar*) adj3 (followup or follow-up or health care or healthcare or procedur* or therap* or treatment*)).ti,ab,kw. (44061)
286 (inappropriate* or unnecessar* or safe or adverse or adversely or undesirabl* or un-desirabl* or unintend* or un-intend* or unintent* or un-intent* or unsafe* or un-safe* or unwanted or un-wanted or harm* or injurious* or risk or risks or side-effect* or sideeffect* or reaction* or complication*).ti. (2809297)
287 ((adverse* or undesirabl* or un-desirabl* or unintend* or un-intend* or unintent* or un-intent* or unwanted or un-wanted or harm* or toxic or injurious* or serious* or fatal) adj5 (affect or affected or affecting or affects or consequence* or effect* or react or reacts or reacted or reacting or reaction* or side-effect* or sideeffect* or event* or outcome* or incident*)).ti,ab,kw. (2352653)
288 ((adverse* or inappropriat* or unnecessar* or undesirabl* or un-desirabl* or unintend* or un-intend* or unintent* or un-intent* or unwanted or un-wanted or injurious* or serious*) adj5 (alarm* or anxiet* or anxious* or distress* or emotion* or feeling* or psycholog* or uncertain* or un-certain*)).ti,ab,kw. (34102)
289 exp Neoplasm Metastasis/ and (avoid* or declin* or decreas* or lessen* or lower* or prevent* or reduc*).ti. (16969)
290 (benefit* or beneficial*).ti,kw,kf. (244174)
291 or/263-290 [BENEFITS/HARMS] (7968699)
292 262 and 291 [BREAST CANCER SCREENING - BENEFITS/HARMS - 2014-PRESENT] (33482)
293 292 use cctr [CENTRAL RECORDS] (973)
294 114 or 229 or 293 [ALL DATABASES] (12063)
295 limit 294 to yr="2020-current" (5322)
296 remove duplicates from 295 (4277)
297 limit 294 to yr="2016-2019" (4658)
298 remove duplicates from 297 (3772)
299 294 not (295 or 297) (2083)
300 remove duplicates from 299 (1684)
301 296 or 298 or 300 [TOTAL UNIQUE RECORDS] (9733)
302 301 use medall [MEDLINE UNIQUE RECORDS] (5635)
303 301 use emczd [EMBASE UNIQUE REORDS] (3503)
304 301 use cctr [CENTRAL UNIQUE REORDS] (595)

**************************

#### Appendix 3 – Completed *PRESS* assessment

Reference: McGowan J, Sampson M, Salzwedel DM, Cogo E, Foerster V, Lefebvre C. PRESS Peer Review of Electronic Search Strategies: 2015 guideline statement. *J Clin Epidemiol* 2016;75:40-6. Available: http://www.jclinepi.com/article/S0895-4356(16)00058-5/pdf.

**Search submission: This section to be filled in by the searcher**

Searcher: Becky Skidmore

Date submitted: 2023 Jul 5 Date requested by: 2023 Jul 7

**1. Systematic Review Title**

Screening for breast cancer: An evidence review to inform the update the Canadian Task Force on Preventive Health Care guideline

**2. This search strategy is …**

| | |
|---|---|
| X | My PRIMARY (core) database strategy — First time submitting a strategy for search question and database |
| | My PRIMARY (core) strategy — Follow-up review NOT the first time submitting a strategy for search question and database. If this is a response to peer review, itemize the changes made to the review suggestions |
| | SECONDARY search strategy— First time submitting a strategy for search question and database |
| | SECONDARY search strategy — NOT the first time submitting a strategy for search question and database. If this is a response to peer review, itemize the changes made to the review suggestions |

**3. Database** (e.g., MEDLINE, CINAHL)

MEDLINE

4. **Interface** (e.g., Ovid, EbscoHost…)

Ovid

#### 5. Research Question (Describe the purpose of the search) *[mandatory]*

This evidence review aims to address the following three key questions (KQs):

1. a) What are the benefits and harms of different mammography-based screening strategies compared to no screening in adults greater than 40 years of age and not at high risk for breast cancer?

   b) Do the benefits and harms differ by population characteristics and risk markers (e.g., age, breast density, race/ethnicity, family history)?

2. a) What are the comparative benefits and harms of different mammography-based breast cancer screening strategies in adults greater than 40 years of age and not at high risk for breast cancer?

   b) Do the comparative benefits and harms differ by population characteristics and risk markers (e.g., age, breast density, race/ethnicity, family history)?

3. How do patients weigh the benefits and harms of breast cancer screening?

#### 6. PICO Format Outline the PICOs for your question — i.e., Patient, Intervention, Comparison, Outcome, and Study Design — as applicable

| | |
|---|---|
| **P** | Adults with female sex-specific breast tissue aged ≥40 years of age and not at high-risk for breast cancer<br><br>Specific populations (using within and between-study data where able):<br>• Age (40-44 years, 45-49 years, 50-69 years, 70-74 years, 75 and older)<br>• Ethnicity, especially Indigenous and Black populations<br>• Socioeconomic status<br>• Geographical location (rural vs. urban settings)<br>• Breast density (i.e., extremely [e.g., BIRADS category D] vs not extremely dense breasts; other comparisons. |
| **I / Exposure** | Adults with female sex-specific breast tissue aged ≥40 years of age and not at high-risk for breast cancer<br><br>Specific populations (using within and between-study data where able):<br>• Age (40-44 years, 45-49 years, 50-69 years, 70-74 years, 75 and older)<br>• Ethnicity, especially Indigenous and Black populations<br>• Socioeconomic status |

| | |
|---|---|
| | • Geographical location (rural vs. urban settings)<br>• Breast density (i.e., extremely [e.g., BIRADS category D] vs not extremely dense breasts; other comparisons. |
| **C** | |
| **O** | **Benefits (reductions)**<br>1. Breast cancer mortality<br>2. All-cause mortality<br>3. Advanced-stage disease (stage III/IV) (including one of (hierarchy): stage III + IV disease; size ≥50 mm, 4+ positive lymph nodes)<br>4. Stage II or higher (including one of (hierarchy): Stage II+, size ≥20 mm, 1+ positive lymph node)<br>5. Treatment-related morbidity (i.e. invasiveness of treatment e.g., less invasive surgeries, need for chemotherapy [early vs. advanced disease])<br>6. Breast cancer morbidity (e.g., adverse effects of treatment, physical/functional impairment)<br>7. Detection of invasive cancer<br><br>**Harms** (7-14 are proportions/# with 1+)<br>8. Overdiagnoses[c]<br>9. Detection of DCIS (cumulative)<br>10. False-positive screens (single round)<br>11. False-positive screens (cumulative over multiple rounds)<br>12. Biopsies on false-positives[d] (single round<br>13. Biopsies on false positives (cumulative over multiple rounds)<br>14. Interval cancers (includes FNs and clinically detected CAs before next screen or time equivalent)<br>15. False negatives<br>16. Incidental findings (if using MRI or ultrasound) |
| **S** | **All outcomes**<br>1. Randomized controlled trials, including cluster<br>2. Non/quasi-randomized controlled trials<br>3. Prospective or retrospective observational studies of large screening cohorts with a concurrent control group (including controlled before-after studies) (i.e., all having exposure data at the individual level and linked with outcomes)<br>4. **If reporting data specific to key demographic groups (i.e., 40-49 and/or 70+ years, race/ethnicity group):** ecological/population-based (e.g., exposure data not at participant level, over multiple years), time trend/series and before-after studies |

#### 7. Inclusion Criteria (List criteria such as age groups, study designs, etc., to be included) *[optional]*

2014 – present

**9. Exclusion Criteria** (List criteria such as study designs, date limits, etc., to be excluded)

**10. Was a search filter applied?** Yes

**If YES, which one(s) (e.g., Cochrane RCT filter, PubMed Clinical Queries filter)? Provide the source if this is a published filter.**
*[mandatory if YES to previous question — textbox]*

Cochrane HSSS, 2008 – sensitivity and specificity-maximizing version with slight adjustments
Other design filters derived from CADTH's

**11. Notes or comments you feel would be useful for the peer reviewer** *[optional]*

Limited time available for this review so important to contain volume where possible. We are including accepted design filters and overlaying this with the outcomes/benefits/harms of interest.
Team has noted they do not want male-only population removed.
Not interested in breast self-examination or physical examination unless included as part of mammography screening.

**12. Please copy and paste your search strategy here, exactly as run, including the number of hits per line. [mandatory]**

Database: Ovid MEDLINE(R) ALL <1946 to July 03, 2023>
Search Strategy:
--------------------------------------------------------------------------------
1 exp Breast Neoplasms/ (341966)
2 ((breast* or mamma or mammar*) adj3 (cancer* or carcinoid* or carcinoma* or carcinogen* or adenocarcinoma* or adeno-carcinoma* or malignan* or neoplasia* or neoplasm* or sarcoma* or tumour* or tumor*)).tw,kw,kf. (424125)
3 exp Carcinoma, Intraductal, Noninfiltrating/ (11207)
4 intraductal carcinoma*.tw,kw,kf. (1194)
5 (ductal carcinoma in situ or DCIS).tw,kw,kf. (9201)
6 or/1-5 [**BREAST CANCER**] (488694)
7 exp Breast Neoplasms/di, pc (52815)
8 exp Mass Screening/ (143920)
9 screen*.tw,kw,kf. (972801)
10 "Early Detection of Cancer"/ (37565)
11 ((early or earlier or earliest) adj3 (detect* or diagnos* or identif* or recogni*)).tw,kw,kf. (333269)
12 exp Breast Neoplasms/dg [diagnostic imaging] (28678)
13 exp Mammography/ (33286)
14 (mammograph* or mammogram*).tw,kw,kf. (36841)
15 exp Magnetic Resonance Imaging/ (533234)
16 (fMRI or fMRIs or MRI or MRIs or NMRI or NMRIs or MR imaging or NMR imaging or magnetic resonance imag* or magnetic resonance tomograph* or MR tomograph*).tw,kw,kf. (543225)

17 (chemical shift imaging or proton spin tomograph* or zeugmatograph*).tw,kw,kf. (1175)
18 (ultrasound* or ultrason* or echograph* or echotomograph* or echo-tomograph* or sonograph*).tw,kw,kf. (498623)
19 (echomammogra* or echo-mammogra*).tw,kw,kf. (11)
20 Imaging, Three-Dimensional/ (81241)
21 ((3D or "3-D") adj3 imag*).tw,kw,kf. (28379)
22 (("3" or three) adj dimension* adj3 imag*).tw,kw,kf. (21005)
23 tomosynthes*.tw,kw,kf. (2309)
24 or/7-23 [**SCREENING**] (2536319)
25 6 and 24 [**BREAST CANCER SCREENING**] (122336)
26 exp Infant/ not (exp Adult/ and exp Infant/) (920674)
27 exp Child/ not (exp Adult/ and exp Child/) (1389724)
28 Adolescent/ not (exp Adult/ and Adolescent/) (685754)
29 or/26-28 (2138455)
30 25 not 29 [**CHILD-ONLY REMOVED**] (121693)
31 exp Animals/ not (exp Animals/ and Humans/) (5135938)
32 30 not 31 [**ANIMAL-ONLY REMOVED**] (120206)
33 (comment or editorial or news or newspaper article).pt. (1682535)
34 (letter not (letter and randomized controlled trial)).pt. (1215441)
35 32 not (33 or 34) [**OPINION PIECES REMOVED**] (112557)
36 Case Reports.pt. (2344054)
37 (case adj (series or study or studies or report or reports)).ti. (413284)
38 35 not (36 or 37) [**CASE STUDIES/SERIES REMOVED**] (102565)
39 **limit 38 to yr="2014-current"** (45629)
40 (controlled clinical trial or randomized controlled trial or pragmatic clinical trial).pt. (687391)
41 clinical trials as topic.sh. (201056)
42 (randomi#ed or randomly or RCT$1 or placebo*).tw. (1203771)
43 ((singl* or doubl* or trebl* or tripl*) adj (mask* or blind* or dumm*)).tw. (197998)
44 trial.ti. (288296)
45 or/40-44 (1610578)
46 39 and 45 [**RCTs**] (3124)
47 controlled clinical trial.pt. (95352)
48 Controlled Clinical Trial/ or Controlled Clinical Trials as Topic/ (100965)
49 (control* adj2 trial).tw,kw,kf. (211810)
50 Non-Randomized Controlled Trials as Topic/ (1062)
51 (nonrandom* or non-random* or quasi-random* or quasi-experiment*).tw,kw,kf. (73381)
52 (nRCT or non-RCT).tw,kw,kf. (526)
53 Controlled Before-After Studies/ (727)
54 (control* adj3 ("before and after" or "before after")).tw,kw,kf. (5290)
55 Interrupted Time Series Analysis/ (1857)
56 time series.tw,kw,kf. (45941)
57 (pre- adj5 post-).tw,kw,kf. (130195)
58 ((pretest adj5 posttest) or (pre-test adj5 post-test)).tw,kw,kf. (11719)

59 Historically Controlled Study/ (227)
60 (control* adj2 study).tw,kw,kf. (210670)
61 Control Groups/ (1970)
62 (control* adj2 group?).tw,kw,kf. (623533)
63 trial.ti. (288296)
64 or/47-63 (1385115)
65 39 and 64 [**nRCTs**] (3280)
66 exp Cohort Studies/ (2497201)
67 cohort?.tw,kw,kf. (859912)
68 Retrospective Studies/ (1128310)
69 (longitudinal or prospective or retrospective).tw,kw,kf. (1686220)
70 ((followup or follow-up) adj (study or studies)).tw,kw,kf. (58993)
71 Observational study.pt. (143540)
72 (observation$2 adj (study or studies)).tw,kw,kf. (164751)
73 ((population or population-based) adj (study or studies or analys#s)).tw,kw,kf. (27804)
74 ((multidimensional or multi-dimensional) adj (study or studies)).tw,kw,kf. (151)
75 Comparative Study.pt. (1912761)
76 ((comparative or comparison) adj (study or studies)).tw,kw,kf. (134460)
77 exp Case-Control Studies/ (1427076)
78 ((case-control* or case-based or case-comparison or case-compeer or case-referrent or case-referent) adj3 (study or studies)).tw,kw,kf. (141417)
79 Multicenter Study.pt. (335485)
80 ((multicenter or multi-center or multicentre or multi-centre) adj (study or studies)).tw,kw,kf. (55453)
81 or/66-80 (5424225)
82 39 and 81 [**OBSERVATIONAL STUDIES**] (16409)
83 46 or 65 or 82 [**RCTs, nRCTs, OBSERVATIONAL STUDIES, 2014-PRESENT**] (18655)
84 exp Mass Screening/ae [Adverse Effects] (928)
85 exp Mass Screening/mo [Mortality] (85)
86 "Early Detection of Cancer"/ae [Adverse Effects] (361)
87 "Early Detection of Cancer"/mo [Mortality] (99)
88 exp Mammography/ae [Adverse Effects] (737)
89 exp Mammography/mo [Mortality] (26)
90 exp Diagnostic Errors/ (122510)
91 exp Neoplasms, Radiation-Induced/ (19503)
92 Mortality/ (49412)
93 ((avoid* or declin* or decreas* or lessen* or lower* or reduc*) adj5 (death* or fatal* or morbidit* or mortalit*)).tw,kw,kf. (246706)
94 ((avoid* or declin* or decreas* or lessen* or lower* or reduc*) adj5 (advanced stage? or (advanc* adj3 cancer?) or biopsy or biopsies or chemotherap* or chemo-therap* or disfigur* or exacerbat* or incidental finding? or progress* stage? or progression* or late? stage? or stage? III or stage? 3 or stage? IV or stage? 4)).tw,kw,kf. (70395)
95 ((avoid* or declin* or decreas* or lessen* or lower* or reduc*) adj5 (adverse* or harm* or impair* or injur* or invasiv* or undesirabl*)).tw,kw,kf. (193250)

96 ((avoid* or declin* or decreas* or lessen* or lower* or reduc*) adj5 (alarm* or anxiet* or anxious* or distress* or emotion* or feeling* or psycholog* or uncertaint*)).tw,kw,kf. (82382)
97 (misdiagnos* or mis-diagnos* or misdetect* or mis-detection or misidentif* or mis-identif*).tw,kw,kf. (50432)
98 (miss$3 adj3 (detect* or diagnos* or identif*)).tw,kw,kf. (13278)
99 ((undetected or ("not" adj3 detect*) or undiagnos* or ("not" adj3 diagnos*) or unidentif* or ("not" adj3 identif*)) adj3 (cancer* or carcinoid* or carcinoma* or carcinogen* or adenocarcinoma* or adeno-carcinoma* or lump or lumps or malignan* or neoplasia* or neoplasm* or sarcoma* or tumour* or tumor*)).tw,kw,kf. (8200)
100 (overdiagnos* or over diagnos*).tw,kw,kf. (6837)
101 (false adj (negative* or positive*)).tw,kw,kf. (89775)
102 ((error* or false$2 or wrong$3) adj3 (alarm* or detect* or diagnos*)).tw,kw,kf. (28787)
103 exp Medical Overuse/ (14991)
104 overtreat*.tw,kw,kf. (6645)
105 ((medical or health service? or procedur* or therap* or treatment*) adj3 (overuse? or overusing or overutilis* or overutiliz*)).tw,kw,kf. (926)
106 ((inappropriate* or unnecessar*) adj3 (followup or follow-up or health care or healthcare or procedur* or therap* or treatment*)).tw,kw. (16431)
107 (inappropriate* or unnecessar* or safe or adverse or adversely or undesirabl* or unintend* or unintent* or unsafe* or unwanted or harm* or injurious* or risk or risks or reaction* or complication*).ti,kw,kf. (1398305)
108 ((adverse* or undesirabl* or unintend* or unintent* or unwanted or harm* or toxic or injurious* or serious* or fatal) adj5 (affect or affected or affecting or affects or consequence* or effect* or react or reacts or reacted or reacting or reaction* or event* or outcome* or incident*)).tw,kw,kf. (850371)
109 ((adverse* or inappropriat* or unnecessar* or undesirabl* or unintend* or unintent* or unwanted or injurious* or serious*) adj5 (alarm* or anxiet* or anxious* or distress* or emotion* or feeling* or psycholog* or uncertaint*)).tw,kw,kf. (12271)
110 (benefit* or beneficial*).ti,kw,kf. (95225)
111 or/84-110 [**BENEFITS/HARMS**] (2972927)
112 83 and 111 [**nRCTs, OBSERVATIONAL STUDIES, 2014-PRESENT - BENEFITS AND HARMS**] (5569)

***************************

### Peer review assessment: this section to be filled in by the reviewer

| | | |
|---|---|---|
| iewer: Kaitryn Campbell | | Date completed: 6 Jul 2023 |

Do you wish to be acknowledged? (If yes, the review team will be advised to add an acknowledgement to any publications related to this work).
Yes please.

The suggested acknowledgement is “We thank Kaitryn Campbell, MLIS, MSc for peer review of the Medline search strategy.”

#### 1. TRANSLATION

| | |
|---|---|
| A ---No revisions | X |
| B --- Revision(s) suggested | |
| C --- Revision(s) required | |

If “B” or “C,” please provide an explanation or example:

**2. BOOLEAN AND PROXIMITY OPERATORS**

| | |
|---|---|
| A ---No revisions | X |
| B --- Revision(s) suggested | |
| C --- Revision(s) required | |

If “B” or “C,” please provide an explanation or example:

**3. SUBJECT HEADINGS**

| | |
|---|---|
| A ---No revisions | |
| B --- Revision(s) suggested | X |
| C --- Revision(s) required | |

If “B” or “C,” please provide an explanation or example:

Line 37, consider doing same thing as you did with Line 34: (case reports not (case reports and randomized controlled trial)).pt., because Case Reports + RCT as publication types exist

For Benefits/Harms Concept, consider adding exp Neoplasm Metastasis/ with appropriate free text terms.

**4. TEXT WORD SEARCHING**

| | |
|---|---|
| A ---No revisions | |
| B --- Revision(s)suggested | X |
| C --- Revision(s) required | |

If “B” or “C,” please provide an explanation or example:

As a synonym everywhere “harm” etc. appear, consider adding: side-effect* OR sideeffect*

Line 93-96, consider adding “prevent*” to (avoid* or declin* or decreas* or lessen* or lower* or reduc*)

Line 97, should “mis-detection” be “mis-detect*”?

Lin 99, consider hyphenating all the following terms: undetected or undiagnos* or unidentif*

Line 104, consider hyphenating: overtreat*

Line 105, consider hyphenating all the following terms: overuse? or overusing or overutilis* or overutiliz*

Line 107, consider hyphenating all the following terms: undesirabl* or unintend* or unintent* or unsafe* or unwanted –also see lines 108 & 109 for all terms mentioned above and consider hyphenating

#### 5. SPELLING, SYNTAX, AND LINE NUMBERS

| | |
|---|---|
| A ---No revisions | X |
| B --- Revision(s)suggested | |
| C --- Revision(s) required | |

If “B” or “C,” please provide an explanation or example:

#### 6. LIMITS AND FILTERS

| | |
|---|---|
| A ---No revisions | X |
| B --- Revision(s) suggested | |
| C --- Revision(s) required | |

If “B” or “C,” please provide an explanation or example:

#### OVERALL EVALUATION (Note: If one or more “revision required” is noted above, the response below must be “revisions required”.)

| | |
|---|---|
| A ---No revisions | |
| B --- Revision(s) suggested | X |
| C --- Revision(s) required | |

Additional comments:

Nicely done. I’ve made a number of minor comments for your consideration.

#### Appendix 4 – Portal submission

| Date of submission | Link | Document name | KQ1 Decision | Notes *e.g., reason for exclusion,* |
|---|---|---|---|---|
| 31-Jul-23 | https://www.mdpi.com/1718-7729/29/5/286 | Marrying Story with Science: The Impact of Outdated and Inconsistent Breast Cancer Screening Practices in Canada | exclude | case report |
| 31-Jul-23 | https://pubmed.ncbi.nlm.nih.gov/25274578/ | Pan-Canadian study of mammography screening and mortality from breast cancer | include (duplicate) | NA |
| 31-Jul-23 | https://pubmed.ncbi.nlm.nih.gov/34279132/ | Breast Density and Risk of Interval Cancers: The Effect of Annual Versus Biennial Screening Mammography Policies in Canada | Exclude (not included in KQ2) | ineligible comparator, comparing annual versus biennial screening (KQ2: Biased selection into study groups) |
| 31-Jul-23 | https://pubmed.ncbi.nlm.nih.gov/34134531/ | The Added Value of Supplemental Breast Ultrasound Screening for Women With Dense Breasts: A Single Center Canadian Experience | Exclude (not included in KQ2) | retrospective review of screening program in women with dense breasts, no comparator group (KQ2 not in excluded study list but within-person comparison of US vs DM) |
| 31-Jul-23 | https://pubmed.ncbi.nlm.nih.gov/26501844/ | Breast Tumor Prognostic Characteristics and Biennial vs Annual Mammography, Age, and Menopausal Status | Exclude (included in KQ2) | ineligible comparator (comparing annual vs biennial) |
| 31-Jul-23 | pubmed.ncbi.nlm.nih.gov/34134531/ | The Added Value of Supplemental Breast Ultrasound Screening for Women With Dense Breasts: A Single Center Canadian Experience | duplicate | NA |
| 31-Jul-23 | https://pubmed.ncbi.nlm.nih.gov/35258677/ | Breast cancer screening in women with extremely dense breasts recommendations of the European Society of Breast Imaging (EUSOBI) | exclude | ineligible study design (not primary research) |
| 31-Jul-23 | https://pubmed.ncbi.nlm.nih.gov/34812692/ | The randomized trial of mammography screening that was not-A cautionary tale | exclude | ineligible study design, commentary |
| 31-Jul-23 | https://academic.oup.com/jbi/article/4/2/108/6555324?login=false | The Fundamental Flaws of the CNBSS Trials: A Scientific Review | exclude | ineligible study design, scientific review |
| 31-Jul-23 | academic.oup.com/jbi/article/4/2/135/6555326?login=false | Errors in Conduct of the CNBSS Trials of Breast Cancer Screening Observed by Research Personnel | exclude | investigation of CNBSS trials, ineligible study design |
| 31-Jul-23 | academic.oup.com/jbi/article/4/2/135/6555326?login=false | Errors in Conduct of the CNBSS Trials of Breast Cancer Screening Observed by Research Personnel | duplicate | NA |
| 31-Jul-23 | https://pubmed.ncbi.nlm.nih.gov/22972810/ | Overdiagnosis in mammographic screening for breast cancer in Europe: a literature review | exclude | ineligible study design, literature review |
| 31-Jul-23 | https://pubmed.ncbi.nlm.nih.gov/12518005/#:~:text=A%20significant%2019%25%20reduction%20in,0.97%3B%20p%3D0.01) | All-cause mortality among breast cancer patients in a screening trial: support for breast cancer mortality as an end point | exclude | Results from Tabar study on the Swedish Two-County study on all-cause mortality already included (Nystrom 2002, Tabar 1989) |
| 31-Jul-23 | https://www.mdpi.com/1718-7729/29Con/8/444 | The Impact of Organised Screening Programs on Breast Cancer Stage at Diagnosis for Canadian Women Aged 40–49 and 50–59 | New include | |
| 31-Jul-23 | https://pubmed.ncbi.nlm.nih.gov/26676234/ | Clinical outcomes of modelling mammography screening strategies | exclude | ineligible comparator |

| Date of submission | Link | Document name | KQ1 Decision | Notes *e.g., reason for exclusion,* |
|---|---|---|---|---|
| 1-Aug-23 | https://pubmed.ncbi.nlm.nih.gov/29987612/ | The Impact of Screening Mammography on Treatment in Women Diagnosed with Breast Cancer | exclude | ineligible comparator |
| 1-Aug-23 | https://academic.oup.com/jbi/article/1/3/161/5553855 | Breast Cancer Screening: Beyond Mortality | exclude | ineligible study design |
| 1-Aug-23 | https://pubmed.ncbi.nlm.nih.gov/30411328/ | The incidence of fatal breast cancer measures the increased effectiveness of therapy in women participating in mammography screening | exclude | ineligible comparator |
| 1-Aug-23 | https://pubmed.ncbi.nlm.nih.gov/30411328/ | The incidence of fatal breast cancer measures the increased effectiveness of therapy in women participating in mammography screening | duplicate | NA |
| 1-Aug-23 | https://pubmed.ncbi.nlm.nih.gov/29267146/ | Obligate Overdiagnosis Due to Mammographic Screening: A Direct Estimate for U.S. Women | exclude | ineligible study design (overdiagnosis rates in the US, may be useful) |
| 1-Aug-23 | https://pubmed.ncbi.nlm.nih.gov/36048585/ | Addressing Misinformation About the Canadian Breast Screening Guidelines | exclude | ineligible study design (could be useful background) |
| 1-Aug-23 | https://pubmed.ncbi.nlm.nih.gov/8234686/ | A critical appraisal of the Canadian National Breast Cancer Screening Study | exclude | ineligible study design |
| 1-Aug-23 | https://pubmed.ncbi.nlm.nih.gov/7842421/ | The excess of patients with advanced breast cancer in young women screened with mammography in the Canadian National Breast Screening Study | New include | Quantitative evaluation of distribution of patients with BC with four or more positive lymph nodes in the CNBSS study, could provide additional info for CNBSS study for 40–49-year-olds. Percentages of patients with breast cancer who were at an advanced state at diagnosis in the NBSS and in previous randomized screening trials were compared. |
| 1-Aug-23 | https://pubmed.ncbi.nlm.nih.gov/8055437/ | Statistical power in breast cancer screening trials and mortality reduction among women 40-49 years of age with particular emphasis on the National Breast Screening Study of Canada | exclude | ineligible study design - commentary |
| 1-Aug-23 | https://pubmed.ncbi.nlm.nih.gov/8372753/ | The Canadian National Breast Screening Study: a Canadian critique | exclude | ineligible study design - commentary |
| 1-Aug-23 | https://pubmed.ncbi.nlm.nih.gov/8234686/ | A critical appraisal of the Canadian National Breast Cancer Screening Study | duplicate | NA |
| 1-Aug-23 | https://pubmed.ncbi.nlm.nih.gov/8372752/ | The Canadian National Breast Screening Study: a critical review | exclude | ineligible study design - microsimulation model |
| 1-Aug-23 | https://europepmc.org/article/med/32058543 | The Value of All-Cause Mortality as a Metric for Assessing Breast Cancer Screening | exclude | ineligible study design |
| 1-Aug-23 | https://academic.oup.com/jbi/article/4/2/105/6539318 | Randomized Controlled Mammography Screening Trials Revisited | exclude | ineligible study design - letter to the editor |
| 1-Aug-23 | https://pubmed.ncbi.nlm.nih.gov/32602400/ | Women's Acceptance of Overdetection in Breast Cancer Screening: Can We Assess Harm-Benefit Tradeoffs? | exclude | ineligible study design, commentary |
| 1-Aug-23 | https://www.ncbi.nlm.nih.gov/pmc/articles/PMC9221595/ | How Did CNBSS Influence Guidelines for So Long and What Can That Teach Us? | exclude | ineligible study design, commentary |

| Date of submission | Link | Document name | KQ1 Decision | Notes *e.g., reason for exclusion,* |
|---|---|---|---|---|
| 1-Aug-23 | https://pubmed.ncbi.nlm.nih.gov/20882563/ | Effectiveness of population-based service screening with mammography for women ages 40 to 49 years: evaluation of the Swedish Mammography Screening in Young Women (SCRY) cohort | exclude | ineligible comparator, compared invited and attended screening |
| 1-Aug-23 | https://pubmed.ncbi.nlm.nih.gov/25413699/ | Insights from the breast cancer screening trials: how screening affects the natural history of breast cancer and implications for evaluating service screening programs | exclude | ineligible study design - systematic review (could use for advanced breast cancer outcomes) |
| 1-Aug-23 | https://www.ncbi.nlm.nih.gov/pmc/articles/PMC6406216/?fbclid=IwAR08LiKJCIGMmkDngQFx92zloaOr2MzRJjt7us5h2CKdAfjny3vIkkVbKbo | Molecular breast imaging detected invasive lobular carcinoma in dense breasts: A case report | exclude | ineligible study design - case report |
| 1-Aug-23 | https://www.facingourrisk.org/uploads/assets/press_releases/nccn-newptgls-bcscreen-final-embargoed-62e1b7f6876c1.pdf?fbclid=IwAR38bvw_1aeFfx16QRMcF9OOVxSvO6j18q7Hmi1rxXmAy-yF1KSFlpNuvg | NCCN Publishes New Patient Guidelines for Breast Cancer Screening and Diagnosis Emphasizing Annual Mammograms for All Average-Risk Women Over 40 | exclude | ineligible study design - patient guideline publication announcement |
| 1-Aug-23 | https://ebm.bmj.com/content/27/5/253 | Adapt or die - How the pandemic made the shift from EBM to EBM+ more urgent | exclude | ineligible study design - narrative review |
| 1-Aug-23 | https://pubmed.ncbi.nlm.nih.gov/34134531/ | The Added Value of Supplemental Breast Ultrasound Screening for Women With Dense Breasts: A Single Center Canadian Experience | duplicate | |
| 2-Aug-23 | https://www.nejm.org/doi/full/10.1056/nejmsb1002538 | Lessons from the Mammography Wars | exclude | ineligible study design - commentary/editorial |
| 2-Aug-23 | https://www.nejm.org/doi/full/10.1056/nejmsb1002538 | Lessons from the Mammography Wars | duplicate | |
| 2-Aug-23 | https://cancer-rose.fr/2023/06/26/quest-ce-que-lhistoire-naturelle-du-cancer | Qu'est ce que l'histoire naturelle du cancer | exclude | ineligible study design - narrative review |
| 2-Aug-23 | https://cancer-rose.fr/2022/05/23/congres-preventing-overdiagnosis-calgary-8-12-juin-2022/ | Documents presented at the Preventing Overdiagnosis congress in Calgary - From balanced information request to censorship. Situation in France | exclude | ineligible study design - narrative review |
| 2-Aug-23 | https://cancer-rose.fr/2021/10/23/quest-ce-quun-surdiagnostic/ | Le surdiagnostic et ses conséquences | exclude | ineligible study design - narrative review |

| Date of submission | Link | Document name | KQ1 Decision | Notes *e.g., reason for exclusion,* |
|---|---|---|---|---|
| 8-Aug-23 | no link | Breast cancer screening in women with extremely dense breasts recommendations of the European Society of Breast Imaging (EUSOBI) | duplicate | NA |
| 8-Aug-23 | no link | Breast Cancer screening in New Brunswick in 2023 | unclear, no link provided | Unclear on what this refers to; title not found |
| 8-Aug-23 | No link | Terri Lynn Mills Story | exclude | Not found - likely personal story |
| 9-Aug-23 | No link | Assurance of Timely Access to Breast Cancer Diagnosis and Treatment by a Regional Breast Health Clinic Serving Both Urban and Rural-Remote Communities | exclude | Case-only cohort study |
| 11-Aug-23 | No link | Update on ongoing breast cancer research with Stats Can | unclear, no link provided | Unclear on what this refers to; title not found |
| 16-Aug-23 | No link | Marrying Story with Science: The Impact of Outdated and Inconsistent Breast Cancer Screening Practices in Canada | duplicate | NA |
| 16-Aug-23 | https://pubmed.ncbi.nlm.nih.gov/27632020/ | Breast cancer screening effect across breast density strata: A case-control study | include (duplicate) | NA |
| 16-Aug-23 | https://pubmed.ncbi.nlm.nih.gov/37150275/ | Breast Cancer Screening for Women at Higher-Than-Average Risk: Updated Recommendations From the ACR | exclude | ineligible study design - other (guideline) |
| 16-Aug-23 | https://pubmed.ncbi.nlm.nih.gov/25984843/ | Identifying women with dense breasts at high risk for interval cancer: a cohort study | exclude | ineligible comparator (all women screened with DM [no comparator)) |
| 16-Aug-23 | https://pubmed.ncbi.nlm.nih.gov/27824483/ | Using Volumetric Breast Density to Quantify the Potential Masking Risk of Mammographic Density | exclude | ineligible comparator - screen detected vs non screen detected cancers |
| 16-Aug-23 | https://pubmed.ncbi.nlm.nih.gov/19155400/ | Tailored supplemental screening for breast cancer: what now and what next? | exclude | ineligible study design (other - narrative review) |
| 16-Aug-23 | https://pubmed.ncbi.nlm.nih.gov/36626696/ | Prospective Multicenter Diagnostic Performance of Technologist-Performed Screening Breast Ultrasound After Tomosynthesis in Women With Dense Breasts (the DBTUST) | exclude | Exclude (not included in KQ2 draft [likely later date than search but no DM comparator]) |
| 16-Aug-23 | https://pubmed.ncbi.nlm.nih.gov/35699706/ | Association of Screening With Digital Breast Tomosynthesis vs Digital Mammography With Risk of Interval Invasive and Advanced Breast Cancer | exclude | Exclude (included in KQ2) |
| 16-Aug-23 | https://pubmed.ncbi.nlm.nih.gov/14735182/ | Breast density as a determinant of interval cancer at mammographic screening | exclude | prior to 2014 |
| 16-Aug-23 | https://pubmed.ncbi.nlm.nih.gov/17229950/ | Mammographic density and the risk and detection of breast cancer | exclude | prior to 2014 |
| 16-Aug-23 | https://pubmed.ncbi.nlm.nih.gov/15337416/ | The randomized trials of breast cancer screening: what have we learned? | exclude | ineligible study design (other - narrative review) |
| 16-Aug-23 | https://pubmed.ncbi.nlm.nih.gov/34003218/ | Screening for Colorectal Cancer US Preventive Services Task Force Recommendation Statement | exclude | ineligible intervention/comparator - does not evaluate BC screening |

| Date of submission | Link | Document name | KQ1 Decision | Notes *e.g., reason for exclusion,* |
|---|---|---|---|---|
| 16-Aug-23 | https://academic.oup.com/jbi/article/1/4/283/5610410 | Screening Breast Ultrasound Using Handheld or Automated Technique in Women with Dense Breasts | exclude | ineligible study design (other - narrative review) |
| 16-Aug-23 | https://pubmed.ncbi.nlm.nih.gov/34134531/ | The Added Value of Supplemental Breast Ultrasound Screening for Women With Dense Breasts: A Single Center Canadian Experience | duplicate | NA |
| 16-Aug-23 | https://pubmed.ncbi.nlm.nih.gov/26547101/ | Sensitivity and specificity of mammography and adjunctive ultrasonography to screen for breast cancer in the Japan Strategic Anti-cancer Randomized Trial (J-START): a randomised controlled trial | Exclude (included in KQ2) | Ineligible comparator (both groups screened - comparator had mammography) |
| 16-Aug-23 | https://pubmed.ncbi.nlm.nih.gov/33724062/ | Supplemental Breast MRI for Women with Extremely Dense Breasts: Results of the Second Screening Round of the DENSE Trial | Exclude (included in KQ2) | ineligible comparator (all women screened) |
| 16-Aug-23 | https://pubmed.ncbi.nlm.nih.gov/32055796/ | Survival Outcomes of Screening with Breast MRI in Women at Elevated Risk of Breast Cancer | Exclude (Not Included in KQ2) | Ineligible comparator (KQ2: all at high risk) |
| 16-Aug-23 | https://pubmed.ncbi.nlm.nih.gov/32096852/ | Comparison of Abbreviated Breast MRI vs Digital Breast Tomosynthesis for Breast Cancer Detection Among Women With Dense Breasts Undergoing Screening | Exclude (Not Included in KQ2) | ineligible comparator (all women screened) (KQ2: Studies using paired designs (i.e., within-person comparison)) |
| 16-Aug-23 | https://pubmed.ncbi.nlm.nih.gov/33724062/ | Supplemental Breast MRI for Women with Extremely Dense Breasts: Results of the Second Screening Round of the DENSE Trial | duplicate | NA |
| 16-Aug-23 | https://pubmed.ncbi.nlm.nih.gov/28221097/ | Supplemental Breast MR Imaging Screening of Women with Average Risk of Breast Cancer | Exclude (Not included in KQ2) | ineligible comparator (all women screened) (KQ2: Studies using paired designs (i.e., within-person comparison)) |
| 16-Aug-23 | https://pubmed.ncbi.nlm.nih.gov/34279132/ | Breast Density and Risk of Interval Cancers: The Effect of Annual Versus Biennial Screening Mammography Policies in Canada | duplicate | NA |
| 16-Aug-23 | https://pubmed.ncbi.nlm.nih.gov/25984843/ | Identifying women with dense breasts at high risk for interval cancer: a cohort study | duplicate | NA |
| 17-Aug-23 | https://www2.gnb.ca/content/gnb/en/departments/health/NewBrunswickCancerNetwork/content/assessment_tool_breast_cancer.html | Healthcare Provider- Stepwise Approach to Breast Cancer Risk Assessment | exclude | ineligible study design (other - narrative review) |
| 17-Aug-23 | https://www2.gnb.ca/content/gnb/fr/ministeres/sante/reseau_du_cancer_du_nouveau-brunswick/content/outil_evaluation_cancer_du_sein.html | Fournisseur de soins de santé: Approche par étapes de l'évaluation du risque du cancer du sein | exclude | ineligible study design (other - narrative review) |
| 19-Aug-23 | https://pubmed.ncbi.nlm.nih.gov/22357883/ | Impact of Mammography Detection on the Course of Breast Cancer in Women Aged 40–49 Years | exclude | prior to 2014 |
| 19-Aug-23 | https://www.acpjournals.org/doi/epdf/10.7326/M21-3577 | Estimation of Breast Cancer Overdiagnosis in a U.S. Breast Screening Cohort | exclude | Ineligible comparator |

| Date of submission | Link | Document name | KQ1 Decision | Notes *e.g., reason for exclusion,* |
|---|---|---|---|---|
| 20-Aug-23 | https://bmchealthservres.biomedcentral.com/articles/10.1186/s12913-023-09738-4 | The aggregate value of cancer screenings in the United States: full potential value and value considering adherence | exclude | ineligible study design (modelling) |
| 20-Aug-23 | https://www.health.harvard.edu/cancer/early-breast-cancer-survival-rates-increasing | Early breast cancer survival rates increasing | exclude | ineligible study design (other - narrative review) |
| 20-Aug-23 | https://ascopubs.org/doi/pdf/10.1200/JCO.23.00348?role=tab | Impact of Breast Cancer Screening on 10-Year Net Survival in Canadian Women Age 40-49 Years | New include | Recent 2023 paper that is relevant - ecological study design with data for 40-49; |
| 20-Aug-23 | https://www.mdpi.com/1718-7729/29/6/311 | Overdetection of Breast Cancer | exclude | ineligible study design - Modelling study |
| 20-Aug-23 | https://www.mdpi.com/1718-7729/29/5/291 | The Impact of Dense Breasts on the Stage of Breast Cancer at Diagnosis: A Review and Options for Supplemental Screening | exclude | ineligible study design - review |
| 20-Aug-23 | https://pubmed.ncbi.nlm.nih.gov/29150482/ | Effect of Mammography Screening on Mortality by Histological Grade | include (duplicate) | NA |
| 20-Aug-23 | https://www.mdpi.com/1718-7729/29/8/445 | Misinformation and Facts about Breast Cancer Screening | exclude | ineligible study design |
| 20-Aug-23 | https://www.mdpi.com/1718-7729/29/6/313 | How Did CNBSS Influence Guidelines for So Long and What Can That Teach Us? | duplicate | NA |
| 20-Aug-23 | https://pubmed.ncbi.nlm.nih.gov/21257850/ | United States Preventive Services Task Force Screening Mammography Recommendations: Science Ignored | exclude | modelling study, pre-2014 |
| 20-Aug-23 | https://onlinelibrary.wiley.com/doi/abs/10.1046/j.1524-4741.1998.430139.x | Biasing the Interpretation of Mammography Screening Data by Age Grouping: Nothing Changes Abruptly at Age 50 | exclude | Ineligible comparator and observational study prior to 2014 |
| 20-Aug-23 | https://jamanetwork.com/journals/jama/fullarticle/2463262 | Breast Cancer Screening for Women at Average Risk 2015 Guideline Update From the American Cancer Society | exclude | Systematic review; reviewed included references |
| 20-Aug-23 | https://pubmed.ncbi.nlm.nih.gov/29064760/ | Screening Mammography in Women 40-49 Years Old Current Evidence | exclude | ineligible study design (other - narrative review) |
| 20-Aug-23 | https://pubmed.ncbi.nlm.nih.gov/26442924/ | Influence of tumour stage at breast cancer detection on survival in modern times: population based study in 173,797 patients | exclude | Ineligible comparator |
| 20-Aug-23 | https://jamanetwork.com/journals/jamasurgery/fullarticle/2673936 | Race/Ethnicity and Age Distribution of Breast Cancer Diagnosis in the United States | exclude | ineligible intervention/comparator - does not evaluate BC screening |
| 20-Aug-23 | https://www.cancer.org/content/dam/cancer-org/research/cancer-facts-and-statistics/breast-cancer-facts-and-figures/breast-cancer-facts-and-figures-2019-2020.pdf | Breast Cancer Facts & Figures 2019-2020 | exclude | ineligible study design (other - narrative review) |
| 20-Aug-23 | https://academic.oup.com/jbi/article/1/3/161/5553855?login=true | Breast Cancer Screening: Beyond Mortality | duplicate | NA |

| Date of submission | Link | Document name | KQ1 Decision | Notes *e.g., reason for exclusion,* |
|---|---|---|---|---|
| 20-Aug-23 | https://pubmed.ncbi.nlm.nih.gov/16517548/ | Rate of over-diagnosis of breast cancer 15 years after end of Malmö mammographic screening trial: follow-up study | include (duplicate from 2018 review) | NA |
| 20-Aug-23 | https://pubmed.ncbi.nlm.nih.gov/22972810/ | Overdiagnosis in mammographic screening for breast cancer in Europe: a literature review | duplicate | NA |
| 20-Aug-23 | https://pubmed.ncbi.nlm.nih.gov/6802546/ | The National Study of Breast Cancer Screening Protocol for a Canadian Randomized Controlled trial of screening for breast cancer in women | exclude | Protocol for Miller RCT |
| 20-Aug-23 | https://pubmed.ncbi.nlm.nih.gov/25274578/ | Pan-Canadian study of mammography screening and mortality from breast cancer | duplicate | NA |
| 20-Aug-23 | https://journals.sagepub.com/doi/full/10.1177/09691413211059461 | The randomized trial of mammography screening that was not—A cautionary tale | duplicate | NA |
| 20-Aug-23 | https://jamanetwork.com/journals/jamainternalmedicine/fullarticle/1861037 | Consequences of False-Positive Screening Mammograms | Exclude (included in KQ3) | Values and preferences outcomes |
| 20-Aug-23 | https://pubmed.ncbi.nlm.nih.gov/25274578/ | Pan-Canadian study of mammography screening and mortality from breast cancer | duplicate | NA |
| 20-Aug-23 | https://pubmed.ncbi.nlm.nih.gov/30411328/ | The incidence of fatal breast cancer measures the increased effectiveness of therapy in women participating in mammography screening | duplicate | NA |
| 20-Aug-23 | https://www.thelancet.com/pdfs/journals/lanonc/PIIS1470-2045(20)30398-3.pdf | Effect of mammographic screening from age 40 years on breast cancer mortality (UK Age trial): final results of a randomised, controlled trial | include (duplicate) | NA |
| 20-Aug-23 | https://www.thelancet.com/journals/lanonc/article/PIIS1470-2045(15)00057-1/fulltext | The UK Age Trial: screening women in their forties | exclude | ineligible study design (other - commentary): note that UK AGE trial RCT itself is included |
| 20-Aug-23 | https://pubmed.ncbi.nlm.nih.gov/24018987/ | A failure analysis of invasive breast cancer: most deaths from disease occur in women not regularly screened | exclude | Ineligible study design - case only cohort study |
| 20-Aug-23 | https://pubmed.ncbi.nlm.nih.gov/21712474/ | Swedish two-county trial: impact of mammographic screening on breast cancer mortality during 3 decades | include (duplicate from 2018 review) | NOTE: Not picked up by searches, but RCT previously included in 2018 review |
| 20-Aug-23 | https://academic.oup.com/jbi/article/1/3/161/5553855 | Breast Cancer Screening: Beyond Mortality | duplicate | NA |
| 20-Aug-23 | https://bmccancer.biomedcentral.com/articles/10.1186/s12885-021-07917-2 | Screening is associated with lower mastectomy rates in eastern Switzerland beyond stage effects | exclude | Ineligible study design - case only cohort study |
| 20-Aug-23 | https://pubmed.ncbi.nlm.nih.gov/17290404/ | A retrospective study of the effect of participation in screening mammography on the use of chemotherapy and breast conserving surgery | exclude | Ineligible study design - case only cohort study |
| 20-Aug-23 | https://pubmed.ncbi.nlm.nih.gov/29987612/ | Impact of Screening Mammography on Treatment in Women Diagnosed with Breast Cancer | duplicate | NA |

| Date of submission | Link | Document name | KQ1 Decision | Notes<br>*e.g., reason for exclusion,* |
|---|---|---|---|---|
| 20-Aug-23 | https://pubmed.ncbi.nlm.nih.gov/34427912/ | Looking at breast cancer through the ethnic and racial lens One size definitely does not fit all | exclude | ineligible study design (other - commentary) |
| 20-Aug-23 | https://pubmed.ncbi.nlm.nih.gov/34427920/ | Age distributions of breast cancer diagnosis and mortality by race and ethnicity in US women | exclude | ineligible comparator |
| 20-Aug-23 | https://academic.oup.com/jbi/article/2/5/416/5901429 | Breast Cancer Screening Recommendations: African American Women Are at a Disadvantage | exclude | ineligible study design (other - narrative review) |
| 20-Aug-23 | https://www.ajronline.org/doi/full/10.2214/ajr.184.1.01840324 | Detection of Breast Cancer on Screening Mammography Allows Patients to Be Treated with Less-Toxic Therapy | exclude | ineligible comparator - Case-only cohort study |
| 20-Aug-23 | https://pubmed.ncbi.nlm.nih.gov/22972810/ | Overdiagnosis in mammographic screening for breast cancer in Europe: a literature review | duplicate | NA |
| 20-Aug-23 | https://pubmed.ncbi.nlm.nih.gov/16505392/ | Screening mammography: do women prefer a higher recall rate given the possibility of earlier detection of cancer? | Exclude (included in KQ3) | outcomes not of interest for KQ1 (patient perspectives) |
| 20-Aug-23 | https://www.ncbi.nlm.nih.gov/pmc/articles/PMC6001765/ | Screening for breast cancer in 2018—what should we be doing today? | exclude | ineligible study design (other - narrative review) |
| 20-Aug-23 | https://academic.oup.com/jbi/article/1/3/161/5553855?login=true | Breast Cancer Screening: Beyond Mortality | duplicate | NA |
| 20-Aug-23 | https://pubmed.ncbi.nlm.nih.gov/34158298/ | Estimations of overdiagnosis in breast cancer screening vary between 0% and over 50%: why? | exclude | ineligible study design - commentary |
| 20-Aug-23 | https://pubmed.ncbi.nlm.nih.gov/26976857/ | Overdiagnosis in Mammographic Screening because of Competing Risk of Death | exclude | ineligible study design - modelling study |
| 20-Aug-23 | https://academic.oup.com/jbi/article/1/4/278/5584369 | Perspectives on the Overdiagnosis of Breast Cancer Associated with Mammographic Screening | exclude | ineligible study design (other - narrative review) |
| 20-Aug-23 | https://pubs.rsna.org/doi/abs/10.1148/radiol.11110716?journalCode=radiology | Mammographic Screening and “Overdiagnosis” | exclude | ineligible study design (other - commentary) |
| 20-Aug-23 | https://academic.oup.com/jbi/article/3/3/273/6260880 | Breast Cancer Screening and Anxiety | exclude | ineligible study design (other - commentary) |
| 20-Aug-23 | https://journals.sagepub.com/doi/full/10.1177/08465371211021996 | Imaging, Paternalism and the Worried Patient: Rethinking Our Approach | exclude | ineligible study design (other - commentary) |
| 20-Aug-23 | https://pubmed.ncbi.nlm.nih.gov/34134531/ | The Added Value of Supplemental Breast Ultrasound Screening for Women With Dense Breasts: A Single Center Canadian Experience | duplicate | NA |
| 20-Aug-23 | https://pubmed.ncbi.nlm.nih.gov/34279132/ | Breast Density and Risk of Interval Cancers: The Effect of Annual Versus Biennial Screening Mammography Policies in Canada | duplicate | NA |
| 20-Aug-23 | https://pubmed.ncbi.nlm.nih.gov/34482760/ | Breast Density and Risk of Interval Cancers | exclude | ineligible study design (other - commentary) |
| 20-Aug-23 | https://www.ajronline.org/doi/epdf/10.2214/ajr.161.4.8372752 | The Canadian National Breast Screening Study : A critical review | exclude | ineligible study design (other - critical review) |

| Date of submission | Link | Document name | KQ1 Decision | Notes *e.g., reason for exclusion,* |
|---|---|---|---|---|
| 20-Aug-23 | https://acsjournals.onlinelibrary.wiley.com/doi/epdf/10.1002/1097-0142%2819950215%2975%3A4%3C997%3A%3AAID-CNCR2820750415%3E3.0.CO%3B2-M | The excess of patients with advanced breast cancer in young women screened with mammography in the Canadian National Breast Screening Study | duplicate | NA |
| 20-Aug-23 | https://pubmed.ncbi.nlm.nih.gov/9012723/ | The review of randomization in the Canadian National Breast Screening Study Is the debate over | exclude | ineligible study design (other - commentary) |
| 22-Aug-23 | No link | Statistical Supplement Quality Indicators by Age Group 2019 small cells suppressed | unclear, no link provided | Can't find study |
| 22-Aug-23 | No link | Statistical Supplement Quality Indicators by Age Group 2020 Small Cell Suppressed | duplicate | Can't find study |
| 23-Aug-23 | https://www.mdpi.com/1718-7729/29/5/286 | Marrying Story with Science: The Impact of Outdated and Inconsistent Breast Cancer Screening Practices in Canada | duplicate | NA |
| 25-Aug-23 | https://www.mdpi.com/1718-7729/29/8/444 | The Impact of Organised Screening Programs on Breast Cancer Stage at Diagnosis for Canadian Women Aged 40–49 and 50–59 | duplicate | NA |
| 25-Aug-23 | https://pubmed.ncbi.nlm.nih.gov/24159387/ | Screening prior to Breast Cancer Diagnosis: The More Things Change, the More They Stay the Same | exclude | Observational study published prior to 2014 and no eligible comparator |
| 25-Aug-23 | https://jamanetwork.com/journals/jamanetworkopen/fullarticle/2808381 | Patterns in Cancer Incidence Among People Younger Than 50 Years in the US, 2010 to 2019 | exclude | ineligible comparator (does not examine BC screening) |
| 27-Aug-23 | https://www.rsna.org/news/2023/january/mri-detects-cancer-in-dense-breasts | Breast MRI Effective at Detecting Cancer in Dense Breasts | Exclude | ineligible comparator (no unscreened participants); note: source submitted was a news article, evaluation was done on associated SR: https://pubs.rsna.org/doi/10.1148/radiol.221785 |
| 27-Aug-23 | https://www.breastcancer.org/research-news/dense-breasts-mri-supplemental | Breast MRI Best Supplemental Screening for Dense Breasts | Exclude | ineligible comparator (no unscreened participants); note: source submitted was a news article, evaluation was done on associated SR: https://pubs.rsna.org/doi/10.1148/radiol.221785 |
| 27-Aug-23 | https://www.ncbi.nlm.nih.gov/pmc/articles/PMC9122856/ | Breast cancer screening in women with extremely dense breasts recommendations of the European Society of Breast Imaging (EUSOBI) | duplicate | NA |
| 28-Aug-23 | https://www.birpublications.org/doi/10.1259/bjr.20211388 | Breast cancer risk predictions by birth cohort and ethnicity in a population-based screening mammography program | include (duplicate) | NA |
| 29-Aug-23 | https://www.mdpi.com/1718-7729/29/8/444 | The Impact of Organised Screening Programs on Breast Cancer Stage at Diagnosis for Canadian Women Aged 40–49 and 50–59 | duplicate | NA |

| Date of submission | Link | Document name | KQ1 Decision | Notes *e.g., reason for exclusion,* |
|---|---|---|---|---|
| 29-Aug-23 | https://ascopubs.org/doi/full/10.1200/JCO.23.00348 | Impact of Breast Cancer Screening on 10-Year Net Survival in Canadian Women Age 40-49 Years | duplicate | NA |
| 29-Aug-23 | https://pubs.rsna.org/doi/10.1148/radiol.2021203935?url_ver=Z39.88-2003&rfr_id=ori:rid:crossref.org&rfr_dat=cr_pub%20%200pubmed | Beneficial Effect of Consecutive Screening Mammography Examinations on Mortality from Breast Cancer: A Prospective Study | include (duplicate) | NA |
| 29-Aug-23 | https://journals.sagepub.com/doi/full/10.1177/09691413211060680 | All-cause mortality in multi-cancer screening trials | exclude | ineligible study design (other - commentary) |
| 29-Aug-23 | https://acsjournals.onlinelibrary.wiley.com/doi/10.1002/cncr.32859 | Mammography screening reduces rates of advanced and fatal breast cancers: Results in 549,091 women | exclude | ineligible study design |
| 29-Aug-23 | https://academic.oup.com/jnci/article/106/11/dju261/1496367?login=false | Pan-Canadian Study of Mammography Screening and Mortality from Breast Cancer | duplicate | NA |
| 29-Aug-23 | https://acsjournals.onlinelibrary.wiley.com/doi/full/10.1002/cncr.31840 | The incidence of fatal breast cancer measures the increased effectiveness of therapy in women participating in mammography screening | duplicate | NA |
| 29-Aug-23 | https://pubmed.ncbi.nlm.nih.gov/34279132/ | Breast Density and Risk of Interval Cancers: The Effect of Annual Versus Biennial Screening Mammography Policies in Canada | duplicate | NA |
| 29-Aug-23 | https://www.ncbi.nlm.nih.gov/pmc/articles/PMC7491203/#:~:text=The%20UK%20Age%20trial%20reported,randomisation%2C%20and%20was%20attenuated%20thereafter. | Effect of mammographic screening from age 40 years on breast cancer mortality (UK Age trial): final results of a randomised, controlled trial | include (duplicate) | NA |
| 1-Sep-23 | https://www.mdpi.com/1718-7729/30/9/571 | Capturing the True Cost of Breast Cancer Treatment: Molecular Subtype and Stage-Specific per-Case Activity-Based Costing | exclude | ineligible comparator (does not examine BC screening) |

#### Appendix 5 – Grey literature search

| Organization | Date searched | Website Link | Articles found |
|---|---|---|---|
| **Relevant websites** | | | |
| Canadian Cancer Trials | 15-Aug-23 | https://clinicaltrials.gov/ | none found |
| ClinicalTrials.gov | 15-Aug-23 | https://clinicaltrials.gov/ | none found |
| WHO International Clinical Trials Registry Platform | 15-Aug-23 | https://trialsearch.who.int/Default.aspx | none found |
| ISRCTN | 15-Aug-23 | https://www.isrctn.com/search?q=breast+cancer+screening | none found |
| CenterWatch | 15-Aug-23 | https://www.centerwatch.com | none found |
| British Columbia Cancer Agency | 15-Aug-23 | http://www.bccancer.bc.ca | none found |
| Cancer Care Ontario | 15-Aug-23 | https://www.cancercareontario.ca/en | none found |
| Canadian Cancer Society | 15-Aug-23 | https://cancer.ca/en/ | none found |
| World Conference on Breast Cancer | 15-Aug-23 | https://www.cancerscience.scientexconference.com/ | none found |
| **Clinical Practice Guidelines** | | | |
| Canadian Medical Association (CMA) | 16-Aug-23 | https://joulecma.ca/cpg/homepage | none found |
| Canadian Partnership Against Cancer | 16-Aug-23 | https://www.partnershipagainstcancer.ca/tools/cancer-guidelines-database/ | none found |
| Canadian Standards Association (CSA) | 16-Aug-23 | https://store.csagroup.org/?cclcl=en_US | none found |
| The College of Physicians and Surgeons of Ontario (CPSO) | 16-Aug-23 | https://www.cpso.on.ca/ | none found |
| **Internet Search** | | | |
| Google (first 5 pages) | 16-Aug-23 | http://www.google.com | none found |
| Google Scholar (first 5 pages) | 16-Aug-23 | https://scholar.google.com/ | none found |
| CMA | 16-Aug-23 | https://www.cma.ca/about-cma | none found |

#### Appendix 6 – DistillerSR screening forms

##### Level 1 – Title and abstract screening

1. After reviewing the PICO criteria, is this study eligible for inclusion?
   - Yes
   - No

| **Instructions in Distiller for exclusions:** |
|---|
| **At this stage please exclude:**<br>• The following populations:<br> ◦ All participants are considered high-risk (e.g., selected on the basis that they all have BRCA1 gene or family history of cancer)<br>• The following study designs:<br> ◦ Cross-sectional studies<br> ◦ Modelling studies (e.g., prediction models, nomograms, **simulation studies**)<br> ◦ Diagnostic accuracy studies (e.g., measuring sensitivity, specificity of a tool)<br> ◦ Studies focusing on behaviours, attitudes, or preferences of screening.<br> ◦ Narrative reviews, editorials, commentaries, case reports<br> ◦ Protocols<br>• The following interventions:<br> ◦ Mammography for diagnosis or imaging<br> ◦ Screening with clinical breast examination or breast self-examination alone<br> ◦ Screening with MRI or ultrasound alone<br>**Note:**<br>Ensure that the participants enrolled into the study that are included based on a breast cancer diagnosis have been linked back to being screen- or not screen-detected cancer.<br>For references without abstracts, we will exclude, unless you can infer from the title that it is clearly a breast cancer screening study. |

##### Level 2 – Full-text screening

1. Is the language of publication English or French?
   - Yes **(include)**
   - No **(exclude)**
2. Please select the study design (*drop down menu*)
   - Randomized or quasi-randomized controlled trial **(include)**
   - Non-randomized controlled trial (published ≥2014) **(include)**
   - Cohort study (published ≥2014) **(include)**
   - Case-control study (published ≥2014) **(include)**
   - Controlled before and after study (published ≥2014) **(include)**

- Time trend/series (published ≥2014) **(include)**
- Ecological (population-based) study (published ≥2014) **(include)**
- Review (including systematic, scoping, and narrative reviews) **(exclude)**
- Observational study (published < 2014) **(exclude)**
- Abstract or conference proceeding **(exclude)**
- Protocol **(exclude)**
- Other **(exclude)**

3. Do any of the following criteria below apply to the study?
    - Participants are younger than 40 years of age **(exclude)**
    - Participants are at high risk of breast cancer (personal or family history of breast or ovarian cancer, significant genetic markers such as BRCA1/BRCA2 or Li-Fraumeni syndrome) **(exclude)**
    - Study uses a screening strategy other than mammography **(exclude)**
    - Ineligible comparator (must be a "no screened group") **(exclude)**
    - Breast imaging or clinical examinations were conducted for diagnosis or surveillance **(exclude)**
    - Not a primary care setting **(exclude)**
    - Screening initiation date <2000 **(exclude)**
    - Does not include any outcomes of interest **(exclude)**
    - None of the above **(include)**
4. Please select which country the study was conducted from the list below *(drop down menu)*
   *Countries that were considered "Very High" on the HDI were included; all others were excluded at this stage.*

Typically, these questions are nested. If an answer is an include (as indicated), the form allows us to proceed to the next question. If an answer was an exclude, the form would end, and that reference would be excluded for that reason.

#### Appendix 7 – Data extraction form

**Level 3 – Data Extraction**
First Tab – Summary Characteristics

---

**Green items can be pulled from Distiller Datarama report and responses to level 2 responses*

*Study identification*

1. First author
2. Year of publication
3. Name of journal/source document
4. Country

*Study characteristics*

5. Study aim
6. Type of study/study design
    a. If cohort study, specify
        i. Screened vs unscreened [SU]
        ii. Offered screening vs not [ON]
        iii. Adhered to screening vs offered but not attended [OA]
        iv. Not applicable [NA]
7. Trial name or database (e.g., CNBSS, SEER)
8. Total study period
9. Dates of screening
10. Screening initiation date prior to and after 2000? (*yes/no*)
11. Duration of follow-up

*Population details*

12. Population details (*Provide brief details on the population studied, copy and pasted from the text is acceptable (e.g., age of participants enrolled, existing comorbidities) otherwise type NR*)
13. Total number of participants
14. Mean or median age at entry
15. Recruitment method (*Details on how the population was selected (e.g., Registry, Claims database, etc.*)
16. Eligibility criteria
17. Other populations studied (e.g., ethnic groups, LGBTQ+, disability)
18. Population health status (*Details on any comorbidities, family history of BC, history of other cancers*)

*Control group (not exposed)*

19. Name of the control/not exposed group
20. Concurrent or historical control?
21. Control group details

22. Total number of participants in the control group
23. Total number in the control group received/not exposed (*Number of participants who received the control (after exclusions, for example)*
24. Number lost to follow-up (if applicable)
25. Number excluded (if applicable)
26. Reasons for exclusions or lost to follow-up

*Intervention group (exposed)*

27. Name of the intervention/exposed group (e.g., type of screening test)
28. Type of mammography received
29. Number of screening intervals
30. Duration of screening interval
31. Intervention/exposed group details as reported (e.g., screening interval, number of screening rounds)
32. Total number of participants in the intervention/exposed group
33. Number lost to follow-up (if applicable)
34. Number excluded (if applicable)
35. Reasons for exclusions or lost to follow-up

**Level 3 – Data Extraction**
Second Tab – Outcome Data

*Outcome data*

1. Relevant outcomes evaluated*

**We will extract absolute values (total numbers and percentages, n/%), odds ratios (OR), relative risks (RR), hazard ratios (HR), standard deviations (SD) and confidence intervals (CI). We will also report on any adjusted variables.*

Outcomes of interest:

**Benefits (reductions)**

i. Breast cancer related mortality
ii. All-cause mortality
iii. Treatment-related morbidity, measured by:
    a. Receipt of radiotherapy (yes/no)
    b. Receipt of chemotherapy (yes/no)
        i. Subgroup by anthracycline vs no anthracycline
    c. Type of surgery: complete mastectomy vs partial mastectomy/lumpectomy
    d. Surgical management of axilla (axial lymph node dissection [ALND] vs sentinel lymph node biopsy [SLNB])
iv. Stage distribution of breast cancer
    a. Stage II and higher
    b. Stage III and higher
    c. Stage IV

v. Breast cancer morbidity (e.g., adverse effects of treatment, physical/functional impairment). Measured using composite scores from different scales

**Harms**

vi. Overdiagnoses (We will calculate the number of excess diagnoses from prospective data with at least 10 years of follow up from the time of enrollment over 1,000 persons screened).
vii. Additional imaging +/- biopsy (cumulative over multiple rounds)
viii. Additional imaging+ biopsy (cumulative over multiple rounds)
ix. Interval cancers (includes false negatives and clinically detected cancers before next screen or time equivalent)
    a. Subgroup by Invasive vs DCIS

**Benefit or harm**

x. Health related quality of life (secondary outcome)
xi. Life years gained (or lost)

*Study findings*

2. Overall study conclusions
3. Reported limitations

#### Appendix 8 – GRADE summary of findings tables and forest plots

##### Table 1: Breast cancer mortality (RCTs, short-case accrual, stratified by age, over 10 years)

**Mammography +/- CBE compared to Usual Care**

| Outcomes | Absolute effects: Risk with Usual Care (Assumed Risk) ‡ | Absolute effects: Absolute effect (95% CI) | Relative effect (95% CI) § | № of participants (studies) * | Quality of the evidence (GRADE) Clinical threshold of 0.5 | Quality of the evidence (GRADE) Clinical threshold of 1 | Comments |
|---|---|---|---|---|---|---|---|
| Sub-Group: Breast-Cancer Mortality (40-49 years)<br><br># Randomised: Unclear<br># Analyzed: Unclear<br>Range of follow-up (yrs): 17.7 to 25.7 | **General population** | | **RR 0.85**<br>(0.78 to 0.93) | Unavailable<br>(8 [1-3, 5-7, 9-10] RCTs) [a] | ⊕⊕◯◯<br>LOW [b,c,d,e,f] | ⊕⊕◯◯<br>LOW [b,c,d,e,f] | Using a threshold of 0.5 or 1 fewer deaths per 1,000, screening may make little to no difference in reducing breast cancer mortality over 10 years for individuals aged 40 to 49 years in a general population. |
| | 1.8 per 1,000 | **0.27 fewer per 1,000**<br>(from 0.13 fewer to 0.40 fewer) | | | | | |
| | **Moderately increased risk**<br>Based on family history (scaled adjustment) | | | | ⊕◯◯◯<br>VERY LOW [b,c,d,f,g] | ⊕⊕◯◯<br>LOW [b,c,d,e,f] | Using a threshold of 0.5 fewer deaths per 1,000, we are very uncertain whether screening decreases breast cancer mortality over 10 years for individuals aged 40 to 49 at moderately increased risk for breast cancer.<br><br>Using a threshold of 1 fewer death per 1,000, screening may make little to no difference in reducing breast cancer mortality over 10 years for individuals aged 40 to 49 years at moderately increased risk for breast cancer. |
| | 2.9 per 1,000 | **0.44 fewer per 1,000**<br>(0.20 fewer to 0.64 fewer) | | | | | |
| | **Moderately increased risk**<br>Based on dense breasts (scaled adjustment) | | | | ⊕◯◯◯<br>VERY LOW [b,c,d,f,g] | ⊕⊕◯◯<br>LOW [b,c,d,e,f] | Using a threshold of 0.5 fewer deaths per 1,000, we are very uncertain whether screening decreases breast cancer mortality over 10 years for individuals aged 40 to 49 at moderately increased risk for breast cancer.<br><br>Using a threshold of 1 fewer death per 1,000, screening may make little to no difference in reducing breast cancer mortality over 10 years for individuals aged 40 to 49 years at moderately increased risk for breast |
| | 3.5 per 1,000 | **0.53 fewer per 1,000**<br>(0.25 fewer to 0.77 fewer) | | | | | |
| Sub-Group: Breast-Cancer Mortality (50-59 years)<br><br># Randomised: Unclear<br># Analyzed: Unclear<br>Range of follow-up (yrs): 18.0 to 30.0 | **General population** | | **RR 0.85**<br>(0.78 to 0.93) | Unavailable<br>(6 [1,3,4,8-10] RCTs) [a] | ⊕◯◯◯<br>VERY LOW [b,c-d,f,g] | ⊕⊕◯◯<br>LOW [b,c,d,e,f] | Using a threshold of 0.5 fewer deaths per 1,000, we are very uncertain whether screening decreases breast cancer mortality over 10 years for individuals aged 50 to 59 a general population risk for breast cancer.<br><br>Using a threshold of 1 fewer death per 1,000, screening may make little to no difference in reducing breast cancer mortality over 10 years for individuals aged 50 to 59 years in a general population. |
| | 3.3 per 1,000 | **0.50 fewer per 1,000**<br>(0.23 fewer to 0.73 fewer) | | | | | |
| | **Moderately increased risk**<br>Based on family history (scaled adjustment) | | | | ⊕◯◯◯<br>VERY LOW [b,d,f,g,h] | ⊕◯◯◯<br>VERY LOW [b,c-d,f,g] | Using a threshold of 0.5 or 1 fewer deaths per 1,000, we are very uncertain whether screening decreases breast cancer mortality over 10 years for individuals aged 50 to 59 years at moderately increased risk for breast cancer. |
| | 5.3 per 1,000 | **0.79 fewer per 1,000**<br>(0.37 fewer to 1.16 fewer) | | | | | |

#### Mammography +/- CBE compared to Usual Care

| Outcomes | Absolute effects | | Relative effect (95% CI) § | № of participants (studies) * | Quality of the evidence (GRADE) Clinical threshold of 0.5 | Quality of the evidence (GRADE) Clinical threshold of 1 | Comments |
|---|---|---|---|---|---|---|---|
| | **Risk with Usual Care (Assumed Risk) ‡** | **Absolute effect (95% CI)** | | | | | |
| | **Moderately increased risk** Based on dense breasts (scaled adjustment) | | | | ⊕◯◯◯ VERY LOW [b,d,f,g,h] | ⊕◯◯◯ VERY LOW [b,c-d,f,g] | Using a threshold of 0.5 or 1 fewer deaths per 1,000, we are very uncertain whether screening decreases breast cancer mortality over 10 years for individuals aged 50 to 59 years at moderately increased risk for breast cancer. |
| | 6.3 per 1,000 | **0.95 fewer per 1,000** (0.44 fewer to 1.39 fewer) | | | | | |
| Sub-Group: Breast-Cancer Mortality (60-69 years)<br><br># Randomised: Unclear<br># Analyzed: Unclear<br>Range of follow-up (yrs): 13.1 to 30.0 | **General population** | | **RR 0.85** (0.78 to 0.93) | Unavailable (4 [3,4,9,11] RCTs) [h] | ⊕◯◯◯ VERY LOW [b,c,d,f,g] | ⊕⊕◯◯ LOW [b,c,d,e,f] | Using a threshold of 0.5 fewer deaths per 1,000, we are very uncertain whether screening decreases breast cancer mortality over 10 years for individuals aged 60 to 69 in a general population.<br><br>Using a threshold of 1 fewer death per 1,000, screening may make little to no difference in reducing breast cancer mortality over 10 years for individuals aged 60 to 69 years in a general population. |
| | 4.3 per 1,000 | **0.65 fewer per 1,000** (0.30 fewer to 0.95 fewer) | | | | | |
| | **Moderately increased risk** Based on family history (scaled adjustment) | | | | ⊕◯◯◯ VERY LOW [b,d,e,f,h] | ⊕◯◯◯ VERY LOW [b,c,d,f,g] | Using a threshold of 0.5 or 1 fewer deaths per 1,000, we are very uncertain whether screening decreases breast cancer mortality over 10 years for individuals aged 60 to 69 in at moderately increased risk. |
| | 6.9 per 1,000 | **1.04 fewer per 1,000** (0.48 fewer to 1.52 fewer) | | | | | |
| | **Moderately increased risk** Based on dense breasts (scaled adjustment) | | | | ⊕◯◯◯ VERY LOW [b,d,e,f,h] | ⊕◯◯◯ VERY LOW [b,c,d,f,g] | Using a threshold of 0.5 or 1 fewer deaths per 1,000, we are very uncertain whether screening decreases breast cancer mortality over 10 years for individuals aged 60 to 69 in at moderately increased risk. |
| | 8.2 per 1,000 | **1.23 fewer per 1,000** (0.57 fewer to 1.80 fewer) | | | | | |
| Sub-Group: Breast-Cancer Mortality (70-74 years)<br><br># Randomised: 18,233<br># Analyzed: Unclear<br>Range of follow-up (yrs): 13.2 to 13.6 | **General population** | | **RR 0.85** (0.78 to 0.93) | Unavailable (2 [3,5] RCTs) | ⊕◯◯◯ VERY LOW [b,d,f,g,h] | ⊕◯◯◯ VERY LOW [b,c,d,f,g] | Using a threshold of 0.5 or 1 fewer deaths per 1,000, we are very uncertain whether screening decreases breast cancer mortality over 10 years for individuals aged 70 to 74 years in a general population. |
| | 6.1 per 1,000 | **0.92 fewer per 1,000** (0.43 fewer to 1.34 fewer) | | | | | |
| | **Moderately increased risk** Based on family history (scaled adjustment) | | | | ⊕◯◯◯ VERY LOW [b,d,e,f,h] | ⊕◯◯◯ VERY LOW [b,c,d,f,g] | Using a threshold of 0.5 or 1 fewer deaths per 1,000, we are very uncertain whether screening decreases breast cancer mortality over 10 years for individuals aged 70 to 74 years at moderately increased risk for breast cancer. |
| | 9.8 per 1,000 | **1.47 fewer per 1,000** (0.69 fewer to 2.16 fewer) | | | | | |

#### Mammography +/- CBE compared to Usual Care

| Outcomes | Absolute effects | | Relative effect (95% CI) § | № of participants (studies) * | Quality of the evidence (GRADE) Clinical threshold of 0.5 | Quality of the evidence (GRADE) Clinical threshold of 1 | Comments |
|---|---|---|---|---|---|---|---|
| | **Risk with Usual Care (Assumed Risk) ‡** | **Absolute effect (95% CI)** | | | | | |
| | **Moderately increased risk** Based on dense breasts (scaled adjustment) | | | | ⊕◯◯◯ VERY LOW [b,d,e,f,h] | ⊕◯◯◯ VERY LOW [b,d,f,g,h] | Using a threshold of 0.5 o 1 fewer deaths per 1,000, we are very uncertain whether screening decreases breast cancer mortality over 10 years for individuals aged 70 to 74 years at moderately increased risk for breast cancer. |
| | 11.6 per 1,000 | **1.74 fewer per 1,000** (0.81 fewer to 2.55 fewer) | | | | | |

‡The baseline risk represents the breast cancer mortality rate over 10 years in an unscreened group based on observational data reported by Coldman et al.[1] To calculate a moderately increased risk group due to family history, we used an estimate from Engmann et al.[2] suggesting that having a first degree relative increases the lifetime risk by 1.6 times and multiplied the general population risk estimate by 1.6. To calculate a moderately increased risk group due to dense breasts, we used an estimate from the Swedish mammography trial which suggested those with high breast density have a relative increased lifetime risk of 1.9.[3]
§ The relative effect is based on our previous systematic review and guideline[4] where a subgroup analysis of relative risk by age was assessed and no difference in RR among subgroups was detected and true differences resulting from age were deemed unlikely. Therefore, we used the RR for all ages rather than focusing on each decade of age as we had previously done in our 2018 guideline.
*The number of participants and studies reflect the previous analysis for each age decade, rather than the number of studies that are included in the relative effect estimate for all ages.
**CI:** Confidence interval; **RR:** Risk ratio

**Bibliography:** 1: Gothenburg (Nystrom 2016[5]), 2: Age (Moss 2015[6]), 3. Swedish Two County (Kopparberg & Ostergotland) (Tabar 2011[7]), 4: Malmo I (Nystrom 2016[5]), 5: Malmo I (Nystrom 2002[8]), 6: Malmo II (Nystrom 2016[5]), 7: CNBSS 1 (Miller 2014[9]), 8: CNBSS 2 (Miller 2014[9]), 9: HIP (Shapiro 1988[10]), 10: Stockholm (Nystrom 2016[5]), 11: Stockholm (Nystrom 2002[8])

**GRADE Working Group grades of evidence**
**High quality:** We are very confident that the true effect lies close to that of the estimate of the effect
**Moderate quality:** We are moderately confident in the effect estimate: The true effect is likely to be close to the estimate of the effect, but there is a possibility that it is substantially different
**Low quality:** Our confidence in the effect estimate is limited: The true effect may be substantially different from the estimate of the effect
**Very low quality:** We have very little confidence in the effect estimate: The true effect is likely to be substantially different from the estimate of effect

#### Explanations

a. Two studies considered quasi-randomised (Stockholm & Gothenburg).
b. Randomisation and allocation concealment were either not reported or there were serious deficiencies in these areas, therefore we rated down once for risk of bias. A sensitivity analysis by risk of bias is presented in Supplemental Material, Appendix 1 and no differences in relative risk were detected between high risk and moderate risk of bias papers. True differences resulting from risk of bias were deemed unlikely, however we still rated down once due to concerns with risk of bias impacting the overall estimate.
c. All point estimates in our pooled analysis lie to one side of our threshold. We did not rate down for inconsistency.
d. Breast density was not addressed. Studies reported in Nystrom 2002 and Nystrom 2016 included one round of screening in the control group as part of the short-case accrual calculation. Therefore, the study estimates may be underestimated (a larger benefit from the intervention may be possible). Data are from trials initiated in the 1960s-1990s and the intervention groups were primarily screened with film mammography. Due to advances in mammography technology and treatment practices, we expect that the magnitude of screening effect may differ if applied to today's Canadian screening context. There are no high-quality clinical trials examining the impact of screening on breast cancer screening deaths using contemporary screening methods. We downrated once for indirectness.
e. Given the large sample size; an optimal sample size calculation was not warranted. The 95% CI does not cross the clinical decision threshold; therefore, we did not rate down for imprecision.
f. According to Egger et al.[11], 10 trials are needed to asses publication bias. We cannot assess publication bias due to insufficient number of trials, therefore, we did not rate down for publication bias.
g. Given the large sample sizes; an optimal sample size calculation was not warranted. The 95% CI crosses the clinical decision threshold; therefore, we rated down once for imprecision.
h. Approximately half of the point estimates in our pooled analysis lie on either side of our threshold. We rated down once for inconsistency.

#### Table 1 forest plot

| Reference | Study | Age (at entry) | Mean Follow-up (yrs) | log [RR] | SE | Weight | Risk Ratio [95%CI] |
|---|---|---|---|---|---|---|---|
| Miller 2014 | CNBSS 1 & 2 | 40-69 | 21.9 | 0.05 | 0.11 | 15.3% | 1.05 [0.85, 1.30] |
| Moss 2015 | AGE | 39-41 | 17.7[a] | -0.13 | 0.09 | 22.2% | 0.88 [0.74, 1.04] |
| Nystrom 2016 | Gothenburg | 40-59 | 24.0 | -0.30 | 0.14 | 9.3% | 0.74 [0.56, 0.98] |
| Nystrom 2016 | Malmo I | 45-70 | 30.0 | -0.13 | 0.12 | 13.7% | 0.88 [0.70, 1.10] |
| Nystrom 2016 | Malmo II | 43-49 | 22.0 | -0.16 | 0.22 | 4.0% | 0.85 [0.55, 1.32]* |
| Nystrom 2016 | Stockholm | 40-65 | 25.0 | -0.06 | 0.17 | 6.5% | 0.94 [0.67, 1.32] |
| Shapiro 1988 | HIP | 40-64 | 18.0 | -0.24 | 0.12 | 12.9% | 0.79 [0.63, 1.00] |
| Tabar 2011 | Swedish Two County | 40-74 | 29.0[b] | -0.31 | 0.10 | 16.2% | 0.73 [0.59, 0.90]** |
| **Total (95% CI)** | | | | | | **100.0%** | **0.85 [0.78, 0.93]** |

Heterogeneity: $Tau^2 = 0.00$; $Chi^2 = 7.80$, df = 7 (P = 0.35); $I^2 = 10\%$

Test for overall effect: Z = 3.49 (P = 0.0005)

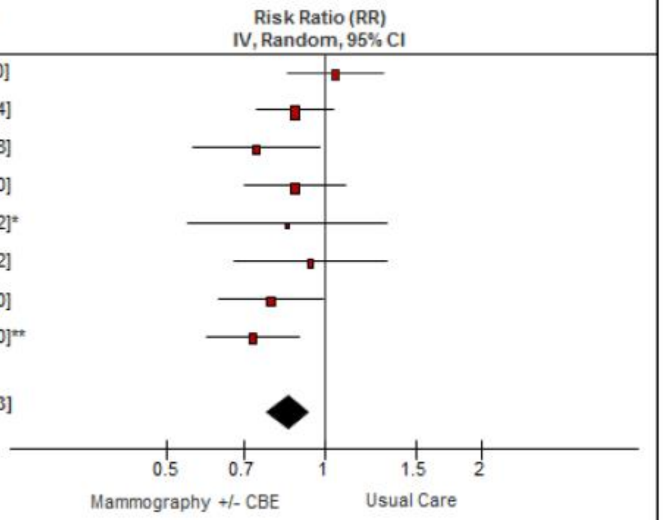

[a]Median; [b]Time since randomization; *Adjusted for Age; ** Adjusted for clustering

#### Table 2: Breast cancer mortality (RCTs, long-case accrual, stratified by age, over 10 years)

##### Mammography +/- CBE compared to Usual Care

| Outcomes | Absolute effects: Risk with Usual Care (Assumed Risk) ‡ | Absolute effects: Absolute effect (95% CI) | Relative effect (95% CI) § | № of participants (studies)* | Quality of the evidence (GRADE) Clinical threshold of 0.5 | Quality of the evidence (GRADE) Clinical threshold of 1 | Comments |
|---|---|---|---|---|---|---|---|
| Sub-Group: Breast-Cancer Mortality (40-49 years)<br><br># Randomised: Unclear<br># Analyzed: Unclear<br>Range of follow-up (yrs): 17.7 to 25.7 | **General population** | | **RR 0.82**<br>(0.71 to 0.94) | Unavailable<br>(6 RCTs [1-5, 7]) [a] | ⊕⊕◯◯<br>LOW [b,c,d,e,f] | ⊕⊕◯◯<br>LOW [b,c,d,e,f] | Using a threshold of 0.5 or 1 fewer deaths per 1,000, screening may make little to no difference in reducing breast cancer mortality over 10 years for individuals aged 40 to 49 years in a general population. |
| | 1.8 per 1,000 | **0.32 fewer per 1,000**<br>(from 0.11 fewer to 0.52 fewer) | | | | | |
| | **Moderately increased risk**<br>Based on family history (scaled adjustment) | | | | ⊕◯◯◯<br>VERY LOW [b,c,d,f,g] | ⊕⊕◯◯<br>LOW [b,c,d,e,f] | Using a threshold of 0.5 fewer deaths per 1,000, we are very uncertain whether screening decreases breast cancer mortality over 10 years for individuals aged 40 to 49 at moderately increased risk for breast cancer.<br><br>Using a threshold of 1 fewer death per 1,000, screening may make little to no difference in reducing breast cancer mortality over 10 years for individuals aged 40 to 49 years at moderately increased risk for breast cancer. |
| | 2.9 per 1,000 | **0.52 fewer per 1,000**<br>(0.17 fewer to 0.84 fewer) | | | | | |
| | **Moderately increased risk**<br>Based on dense breasts (scaled adjustment) | | | | ⊕◯◯◯<br>VERY LOW [b,c,d,f,g] | ⊕⊕◯◯<br>LOW [b,c,d,e,f] | Using a threshold of 0.5 fewer deaths per 1,000, we are very uncertain whether screening decreases breast cancer mortality over 10 years for individuals aged 40 to 49 at moderately increased risk for breast cancer.<br><br>Using a threshold of 1 fewer death per 1,000, screening may make little to no difference in reducing breast cancer mortality over 10 years for individuals aged 40 to 49 years at moderately increased risk for breast cancer. |
| | 3.5 per 1,000 | **0.63 fewer per 1,000**<br>(0.21 fewer to 1.02 fewer) | | | | | |
| Sub-Group: Breast-Cancer Mortality (50-59 years)<br><br># Randomised: Unclear<br># Analyzed: Unclear<br>Range of follow-up (yrs): 18.0 to 30.0 | **General population** | | **RR 0.82**<br>(0.71 to 0.94) | Unavailable<br>(5 RCTs [1, 3, 4, 6, 7]) [a] | ⊕◯◯◯<br>VERY LOW [b,d,f,g,h] | ⊕⊕◯◯<br>LOW [b,d,e,f,h] | Using a threshold of 0.5 fewer deaths per 1,000, we are very uncertain whether screening decreases breast cancer mortality over 10 years for individuals aged 50 to 59 a general population risk for breast cancer.<br><br>Using a threshold of 1 fewer death per 1,000, screening may make little to no difference in reducing breast cancer mortality over 10 years for individuals aged 50 to 59 years in a general population. |
| | 3.3 per 1,000 | **0.59 fewer per 1,000**<br>(0.20 fewer to 0.96 fewer) | | | | | |
| | **Moderately increased risk**<br>Based on family history (scaled adjustment) | | | | ⊕◯◯◯<br>VERY LOW [b,d,f,g,h] | ⊕◯◯◯<br>VERY LOW [b,d,g,f,h] | Using a threshold of 0.5 or 1 fewer deaths per 1,000, we are very uncertain whether screening decreases breast cancer mortality over 10 years for individuals aged 50 to 59 years at moderately increased risk for breast cancer. |
| | 5.3 per 1,000 | **0.95 fewer per 1,000**<br>(0.32 fewer to 1.54 fewer) | | | | | |
| | **Moderately increased risk**<br>Based on dense breasts (scaled adjustment) | | | | ⊕◯◯◯<br>VERY LOW [b,d,f,g,h] | ⊕◯◯◯<br>VERY LOW [b,d,f,g,h] | Using a threshold of 0.5 or 1 fewer deaths per 1,000, we are very uncertain whether screening decreases breast cancer mortality over 10 years for individuals aged 50 to 59 years at moderately increased risk for breast cancer. |
| | 6.3 per 1,000 | **1.13 fewer per 1,000**<br>(0.38 fewer to 1.83 fewer) | | | | | |

#### Mammography +/- CBE compared to Usual Care

| Outcomes | Absolute effects: Risk with Usual Care (Assumed Risk) ‡ | Absolute effects: Absolute effect (95% CI) | Relative effect (95% CI) § | № of participants (studies)* | Quality of the evidence (GRADE) Clinical threshold of 0.5 | Quality of the evidence (GRADE) Clinical threshold of 1 | Comments |
|---|---|---|---|---|---|---|---|
| Sub-Group: Breast-Cancer Mortality (60-69 years)<br><br># Randomised: Unclear<br># Analyzed: Unclear<br>Range of follow-up (yrs): 13.1 to 30.0 | **General population** | | **RR 0.82**<br>(0.71 to 0.94) | Unavailable<br>(3 RCTs[3,4,7]) | ⊕◯◯◯<br>VERY LOW[b,d,f,g,h] | ⊕◯◯◯<br>VERY LOW[b,d,f,g,h] | Using a threshold of 0.5 or 1 fewer deaths per 1,000, we are very uncertain whether screening decreases breast cancer mortality over 10 years for individuals aged 60 to 69 years in a general population. |
| | 4.3 per 1,000 | **0.77 fewer per 1,000**<br>(0.26 fewer to 1.25 fewer) | | | | | |
| | **Moderately increased risk**<br>Based on family history (scaled adjustment) | | | | ⊕◯◯◯<br>VERY LOW[b,d,f,g,h] | ⊕◯◯◯<br>VERY LOW[b,d,f,g,h] | Using a threshold of 0.5 or 1 fewer deaths per 1,000, we are very uncertain whether screening decreases breast cancer mortality over 10 years for individuals aged 60 to 69 years at moderately increased risk for breast cancer. |
| | 6.9 per 1,000 | **1.24 fewer per 1,000**<br>(0.41 fewer to 2 fewer) | | | | | |
| | **Moderately increased risk**<br>Based on dense breasts (scaled adjustment) | | | | ⊕⊕◯◯<br>LOW[b,d,e,f,h] | ⊕◯◯◯<br>VERY LOW[b,d,f,g,h] | Using a threshold of 0.5 fewer deaths per 1,000, screening may make little to no difference in reducing breast cancer mortality over 10 years for individuals aged 60 to 69 years at moderately increased risk.<br><br>Using a threshold of 1 fewer death per 1,000, we are very uncertain whether screening decreases breast cancer mortality over 10 years for individuals aged 60 to 69 years at moderately increased risk for breast cancer. |
| | 8.2 per 1,000 | **1.48 fewer per 1,000**<br>(0.49 fewer to 2.38 fewer) | | | | | |
| Sub-Group: Breast-Cancer Mortality (70-74 years)<br><br># Randomised: 18,233<br># Analyzed: Unclear<br>Range of follow-up (yrs): 13.2 to 13.6 | **General population** | | **RR 0.82**<br>(0.71 to 0.94) | Unavailable<br>(2 RCTs[3,4]) | ⊕◯◯◯<br>VERY LOW[b,d,f,g,h] | ⊕◯◯◯<br>VERY LOW[b,d,f,g,h] | Using a threshold of 0.5 or 1 fewer deaths per 1,000, we are very uncertain whether screening decreases breast cancer mortality over 10 years for individuals aged 70 to 74 years in a general population. |
| | 6.1 per 1,000 | **1.10 fewer per 1,000**<br>(0.37 fewer to 1.77 fewer) | | | | | |
| | **Moderately increased risk**<br>Based on family history (scaled adjustment) | | | | ⊕⊕◯◯<br>LOW[b,d,e,f,h] | ⊕◯◯◯<br>VERY LOW[b,d,f,g,h] | Using a threshold of 0.5 fewer deaths per 1,000, screening may make little to no difference in reducing breast cancer mortality over 10 years for individuals aged 70 to 74 at moderately increased risk for breast cancer.<br><br>Using a threshold of 1 fewer death per 1,000, we are very uncertain whether screening decreases breast cancer mortality over 10 years for individuals aged 70 to 74 years at moderately increased risk for breast cancer. |
| | 9.8 per 1,000 | **1.76 fewer per 1,000**<br>(0.59 fewer to 2.84 fewer) | | | | | |
| | **Moderately increased risk**<br>Based on dense breasts (scaled adjustment) | | | | ⊕⊕◯◯<br>LOW[b,d,e,f,h] | ⊕◯◯◯<br>VERY LOW[b,d,f,g,h] | Using a threshold of 0.5 fewer deaths per 1,000, screening may make little to no difference in reducing breast cancer mortality over 10 years for individuals aged 70 to 74 at moderately increased risk for breast cancer.<br><br>Using a threshold of 1 fewer death per 1,000, we are very uncertain whether screening decreases breast cancer mortality over 10 years for individuals aged 70 to 74 years at moderately increased risk for breast cancer. |
| | 11.6 per 1,000 | **2.09 fewer per 1,000**<br>(0.70 fewer to 3.36 fewer) | | | | | |

| Mammography +/- CBE compared to Usual Care | | | | | | | |
|---|---|---|---|---|---|---|---|
| Outcomes | **Absolute effects** | | Relative effect (95% CI) § | № of participants (studies)* | Quality of the evidence (GRADE) Clinical threshold of 0.5 | Quality of the evidence (GRADE) Clinical threshold of 1 | Comments |
| | **Risk with Usual Care (Assumed Risk) ‡** | **Absolute effect (95% CI)** | | | | | |

‡The baseline risk represents the breast cancer mortality rate over 10 years in an unscreened group based on Canadian observational data reported by Coldman et al.[1] To calculate moderately increased risk group, we used an estimate from Engmann et al.[2] suggesting that having a first degree relative increases the lifetime risk by 1.6 times and multiplied the general population risk estimate by 1.6.
§ The relative effect is based on our previous systematic review and guideline[4] where a subgroup analysis of relative risk by age was assessed and no difference in RR among subgroups was detected and true differences resulting from age were deemed unlikely. Therefore, we used the RR for all ages rather than focusing on each decade of age as we had previously done in our 2018 guideline.
*The number of participants and studies reflect the previous analysis for each age decade, rather than the number of studies that are included in the relative effect estimate for all ages.
**CI:** Confidence interval; **RR:** Risk ratio

**Bibliography:**
1: Gothenburg (Bjurstam 2003[12]), 2: Age (Moss 2015[6]), 3. Swedish Two County (Kopparberg) (Tabar 1995[13]), 4: Swedish Two County (Ostergotland) (Tabar 1995[13]), 5: CNBSS 1 (Miller 2014[9]), 6: CNBSS 2 (Miller 2014[9]), 7: HIP (Habbema 1986[14])
*Note: Long-case accrual unavailable for the following studies: Malmo I, Malmo II, and Stockholm*

**GRADE Working Group grades of evidence**
**High quality:** We are very confident that the true effect lies close to that of the estimate of the effect
**Moderate quality:** We are moderately confident in the effect estimate: The true effect is likely to be close to the estimate of the effect, but there is a possibility that it is substantially different
**Low quality:** Our confidence in the effect estimate is limited: The true effect may be substantially different from the estimate of the effect
**Very low quality:** We have very little confidence in the effect estimate: The true effect is likely to be substantially different from the estimate of effect

#### Explanations

a. One study considered quasi-randomised (Gothenburg)
b. Randomisation and allocation concealment were either not reported or there were serious deficiencies in these areas, therefore we rated down once for risk of bias.
c. All point estimates in our pooled analysis lie to one side of our threshold. We did not rate down for inconsistency .
d. Breast density was not addressed. For some studies, the control group received screening after the screening period. Studies reported in Nystrom 2002 and Nystrom 2016 included one round of screening in the control group as part of the short-case accrual calculation. Therefore, the study estimates may be underestimated (a larger benefit from the intervention may be possible). Downrated once for indirectness. Data are from trials initiated in the 1960s-1990s and the intervention groups were primarily screened with film mammography. Due to advances in mammography technology and treatment practices, we expect that the magnitude of screening effect may differ if applied to today's Canadian screening context. There are no high-quality clinical trials examining the impact of screening on breast cancer screening deaths using contemporary screening methods.
e. Given the large sample sizes; an optimal sample size calculation was not warranted. The 95% CI does not cross the clinical decision threshold; therefore, we did not rate down for imprecision.
f. According to Egger et al.[11], 10 trials are needed to asses publication bias. We cannot assess publication bias due to insufficient number of trials, therefore, we did not rate down for publication bias.
g. Given the large sample sizes; an optimal sample size calculation was not warranted. The 95% CI crosses the clinical decision threshold; therefore, we rated down once for imprecision.
h. Approximately half of the point estimates in our pooled analysis lie on either side of our threshold. We rated down once for inconsistency.

#### Table 2 forest plot

| Reference | Study | Age (at entry) | Mean Follow-up (yrs) | log [RR] | SE | Weight | Risk Ratio [95%CI] | Risk Ratio (RR) IV, Random, 95% CI |
|---|---|---|---|---|---|---|---|---|
| Bjurstam 2003 | Gothenburg | 40-59 | 13.8 | -0.29 | 0.13 | 14.0% | 0.75 [0.58, 0.97] | |
| Habbema 1986 | HIP | 40-64 | 14.0 | -0.25 | 0.10 | 16.9% | 0.78 [0.64, 0.96] | |
| Miller 2014 | CNBSS 1 & 2 | 40-69 | 21.9 | -0.01 | 0.06 | 21.9% | 0.99 [0.88, 1.12] | |
| Moss 2015 | AGE | 39-41 | 17.7[a] | -0.07 | 0.08 | 19.9% | 0.93 [0.80, 1.09] | |
| Tabar 1995 | Kopparberg | 40-74 | 12.5 | -0.51 | 0.14 | 13.4% | 0.60 [0.46, 0.79]* | |
| Tabar 1995 | Ostergotland | 40-74 | 12.5 | -0.25 | 0.13 | 13.9% | 0.78 [0.60, 1.01]* | |
| **Total (95% CI)** | | | | | | **100.0%** | **0.82 [0.71, 0.94]** | |

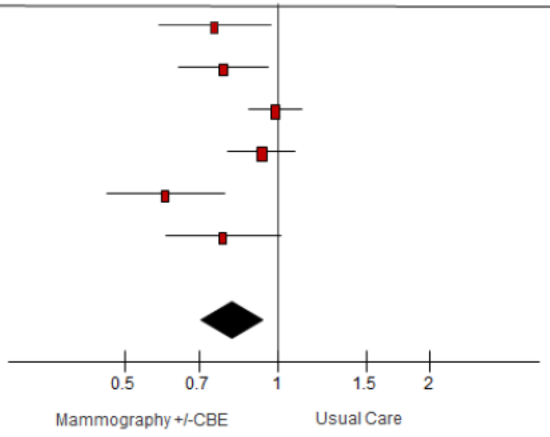

Heterogeneity: Tau² = 0.02; Chi² = 15.42, df = 5 (P = 0.009); I² = 68%
Test for overall effect: Z = 2.79 (P = 0.005)

[a]Median; *Adjusted for age and clustering

**Table 3: Breast cancer mortality (Observational studies, stratified by age, adherence to screen analysis over 10 years)**

#### Screening with mammography* compared to no screening

| Outcomes | Absolute effects | | Risk ratio (95% CI) | № of participants (studies) | Quality of the evidence (GRADE) Clinical threshold of 0.5 | Quality of the evidence (GRADE) Clinical threshold of 1 | Comments |
|---|---|---|---|---|---|---|---|
| | **Risk with Usual Care (Assumed Risk) ‡** | **Absolute effect (95% CI)** | | | | | |
| Sub-Group: Breast-Cancer Mortality (40-49 years)<br><br>Range of follow-up (yrs): 10.0 to 22.0 | **General population** | | **RR 0.48****<br>(0.41 to 0.57) | Unavailable<br>(4 studies[1-4]) | ⊕◯◯◯<br>VERY LOW [a,b,c,d,f,g] | ⊕◯◯◯<br>VERY LOW [a,b,c,e,f,g] | Using a threshold of 0.5 or 1 fewer deaths per 1,000, we are very uncertain whether screening decreases breast cancer mortality over 10 years for individuals aged 40 to 49 years in a general population. |
| | 1.8 per 1,000 | **0.94 fewer per 1,000**<br>(from 0.77 fewer to 1.06 fewer) | | | | | |
| | **Moderately increased risk**<br>Based on family history (scaled adjustment) | | | | ⊕◯◯◯<br>VERY LOW [a,b,c,d,f,g] | ⊕◯◯◯<br>VERY LOW [a,b,c,d,f,g] | Using a threshold of 0.5 or 1 fewer deaths per 1,000, we are very uncertain whether screening decreases breast cancer mortality over 10 years for individuals aged 40 to 49 years at a moderately increased risk for breast cancer. |
| | 2.9 per 1,000 | **1.51 fewer per 1,000**<br>(from 1.25 fewer to 1.71 fewer) | | | | | |
| | **Moderately increased risk**<br>Based on dense breasts (scaled adjustment) | | | | ⊕◯◯◯<br>VERY LOW [a,b,c,d,f,g] | ⊕◯◯◯<br>VERY LOW [a,b,c,d,f,g] | Using a threshold of 0.5 or 1 fewer deaths per 1,000, we are very uncertain whether screening decreases breast cancer mortality over 10 years for individuals aged 40 to 49 years at a moderately increased risk for breast cancer. |
| | 3.5 per 1,000 | **1.82 fewer per 1,000**<br>(1.51 fewer to 2.07 fewer) | | | | | |
| Sub-Group: Breast-Cancer Mortality (50-59 years)<br><br>Range of follow-up (yrs): 10.0 to 22.0 | **General population** | | **RR 0.48****<br>(0.41 to 0.57) | Unavailable<br>(4 studies[1-4]) | ⊕◯◯◯<br>VERY LOW [a,b,c,d,f,g] | ⊕◯◯◯<br>VERY LOW [a,b,c,d,f,g] | Using a threshold of 0.5 or 1 fewer deaths per 1,000, we are very uncertain whether screening decreases breast cancer mortality over 10 years for individuals aged 50 to 59 years in a general population. |
| | 3.3 per 1,000 | **1.72 fewer per 1,000**<br>(from 1.42 to 1.95) | | | | | |
| | **Moderately increased risk**<br>Based on family history (scaled adjustment) | | | | ⊕◯◯◯<br>VERY LOW [a,b,c,d,f,g] | ⊕◯◯◯<br>VERY LOW [a,b,c,d,f,g] | Using a threshold of 0.5 or 1 fewer deaths per 1,000, we are very uncertain whether screening decreases breast cancer mortality over 10 years for individuals aged 50 to 59 years at a moderately increased risk for breast cancer. |
| | 5.3 per 1,000 | **2.76 fewer per 1,000**<br>(from 2.28 to 3.13) | | | | | |
| | **Moderately increased risk**<br>Based on dense breasts (scaled adjustment) | | | | ⊕◯◯◯<br>VERY LOW [a,b,c,d,f,g] | ⊕◯◯◯<br>VERY LOW [a,b,c,d,f,g] | Using a threshold of 0.5 or 1 fewer deaths per 1,000, we are very uncertain whether screening decreases breast cancer mortality over 10 years for individuals aged 50 to 59 years at a moderately increased risk for breast cancer. |
| | 6.3 per 1,000 | **3.28 fewer per 1,000**<br>(2.71 fewer to 3.72 fewer) | | | | | |
| Sub-Group: Breast-Cancer Mortality (60-69 years)<br><br>Range of follow-up (yrs): 10.0 to 22.0 | **General population** | | **RR 0.48****<br>(0.41 to 0.57) | Unavailable<br>(4 studies[1-4]) | ⊕◯◯◯<br>VERY LOW [a,b,c,d,f,g] | ⊕◯◯◯<br>VERY LOW [a,b,c,d,f,g] | Using a threshold of 0.5 or 1 fewer deaths per 1,000, we are very uncertain whether screening decreases breast cancer mortality over 10 years for individuals aged 60 to 69 years in a general population. |
| | 4.3 per 1,000 | **2.24 fewer per 1,000**<br>(from 1.85 to 2.54) | | | | | |
| | **Moderately increased risk**<br>Based on family history (scaled adjustment) | | | | | | Using a threshold of 0.5 or 1 fewer deaths per 1,000, we are very uncertain whether screening decreases breast cancer |

#### Screening with mammography* compared to no screening

| Outcomes | Absolute effects: Risk with Usual Care (Assumed Risk) ‡ | Absolute effects: Absolute effect (95% CI) | Risk ratio (95% CI) | № of participants (studies) | Quality of the evidence (GRADE) Clinical threshold of 0.5 | Quality of the evidence (GRADE) Clinical threshold of 1 | Comments |
|---|---|---|---|---|---|---|---|
| | 6.9 per 1,000 | **3.59 fewer per 1,000** (from 2.97 to 4.07) | | | ⊕◯◯◯ VERY LOW [a,b,c,d,f,g] | ⊕◯◯◯ VERY LOW [a,b,c,d,f,g] | mortality over 10 years for individuals aged 60 to 69 years at a moderately increased risk for breast cancer. |
| | **Moderately increased risk** Based on dense breasts (scaled adjustment) | | | | ⊕◯◯◯ VERY LOW [a,b,c,d,f,g] | ⊕◯◯◯ VERY LOW [a,b,c,d,f,g] | Using a threshold of 0.5 or 1 fewer deaths per 1,000, we are very uncertain whether screening decreases breast cancer mortality over 10 years for individuals aged 60 to 69 years at a moderately increased risk for breast cancer. |
| | 8.2 per 1,000 | **4.26 fewer per 1,000** (3.53 fewer to 4.84 fewer) | | | | | |
| Sub-Group: Breast-Cancer Mortality (70-74 years)<br><br>Range of follow-up (yrs): 10.0 to 22.0 | **General population** | | **RR 0.48**** (0.41 to 0.57) | Unavailable (4 studies[1-4]) | ⊕◯◯◯ VERY LOW [a,b,c,d,f,g] | ⊕◯◯◯ VERY LOW [a,b,c,d,f,g] | Using a threshold of 0.5 or 1 fewer deaths per 1,000, we are very uncertain whether screening decreases breast cancer mortality over 10 years for individuals aged 70 to 74 years in a general population. |
| | 6.1 per 1,000 | **3.17 fewer per 1,000** (from 2.62 to 3.60) | | | | | |
| | **Moderately increased risk** Based on family history (scaled adjustment) | | | | ⊕◯◯◯ VERY LOW [a,b,c,d,f,g] | ⊕◯◯◯ VERY LOW [a,b,c,d,f,g] | Using a threshold of 0.5 or 1 fewer deaths per 1,000, we are very uncertain whether screening decreases breast cancer mortality over 10 years for individuals aged 70 to 74 years at a moderately increased risk for breast cancer. |
| | 9.8 per 1,000 | **5.10 fewer per 1,000** (from 4.21 to 5.78) | | | | | |
| | **Moderately increased risk** Based on dense breasts (scaled adjustment) | | | | ⊕◯◯◯ VERY LOW [a,b,c,d,f,g] | ⊕◯◯◯ VERY LOW [a,b,c,d,f,g] | Using a threshold of 0.5 or 1 fewer deaths per 1,000, we are very uncertain whether screening decreases breast cancer mortality over 10 years for individuals aged 70 to 74 years at a moderately increased risk for breast cancer. |
| | 11.6 per 1,000 | **6.03 fewer per 1,000** (4.99 fewer to 6.84 fewer) | | | | | |

‡The baseline risk (in the control group) was not representative of all included studies. Numerators and/or denominators were either unclear or not reported for some studies. For the age subgroup calculations, the baseline risk for each age group was taken from the Coldman cohort study.
*Studies varied between film and digital mammography.
**Pooling was performed for a screening adherence analysis. To note that Coldman reported a standardized mortality ratio (SMR) however, it has been noted in the literature that an SMR can approximate a RR when the mortality rate in the control group is less than 10 per 1000 for a one-year period in a 10-year age band (Symons and Taulbee, 1981). The statistical heterogeneity of this estimate is high ($I^2$=94%). Other sensitivity analyses for combining these four studies are provided in Supplemental KQ1 GRADE Material, Appendix 3.
**CI:** Confidence interval; **RR:** Risk ratio

**Bibliography:** 1: Choi 2021[15], 2: Coldman 2014[16], 3. Duffy 2021[17], 4: Morrell 2017[18]

**GRADE Working Group grades of evidence**
**High quality:** We are very confident that the true effect lies close to that of the estimate of the effect
**Moderate quality:** We are moderately confident in the effect estimate: The true effect is likely to be close to the estimate of the effect, but there is a possibility that it is substantially different
**Low quality:** Our confidence in the effect estimate is limited: The true effect may be substantially different from the estimate of the effect
**Very low quality:** We have very little confidence in the effect estimate: The true effect is likely to be substantially different from the estimate of effect

#### Explanations

a. We rated down once for the lack of adjustment for important confounding factors across studies, including use of hormone replacement therapy, socioeconomic status, or other adjustment for self-selection bias. Lack of reporting or measurement of population at increased risk of breast cancer (Duffy, Morell). Studies did not report average follow-up length and reasons for loss to follow-up are not reported (Duffy, Morell).
b. Heterogeneity is very high across studies (I2=94%); (p-value<0.0001). Estimates from studies included rate ratios, risk ratios and standardized mortality ratios, with varying degrees of adjustment for confounding factors. We are unable to explain the high statistical heterogeneity through sensitivity analyses (Supplemental KQ1 GRADE Material, Appendix 3), however, all individual estimates point to a reduction in BC mortality. Similarly, all point estimates in our pooled analysis lie to one side of our threshold, therefore we did not rate down for inconsistency.
c. We did not rate for indirectness as both the studies (Duffy and Coldman) are population-based studies representing general population..

d. The 95% CI does not cross the clinical decision threshold; therefore, we did not rate down for imprecision.
e. The 95% CI crosses the clinical decision threshold; therefore, we rated down once for imprecision.
f. We did not rate up for the magnitude of effect because not all plausible confounders (e.g., age, hormone replacement therapy, breast density, elevated risk), were adjusted for, decreasing our confidence in the estimated effect. Following GRADE guidance, the RR is on the threshold of being considered a large effect (i.e., RR either >2.0 or <0.5 based on consistent evidence from at least 2 studies, with no plausible confounders).
g. According to Egger et al.[11], 10 studies are needed to asses publication bias. We cannot assess publication bias due to insufficient number of trials, therefore, we did not rate down for publication bias.

#### Table 3 forest plot

| Study or Subgroup | log[Risk Ratio] | SE | Weight | Risk Ratio IV, Random, 95% CI | Year |
|---|---|---|---|---|---|
| Coldman 2014 | -0.5108 | 0.0563 | 25.9% | 0.60 [0.54, 0.67] | 2014 |
| Morrell 2017 | -0.9676 | 0.1297 | 17.4% | 0.38 [0.29, 0.49] | 2017 |
| Duffy 2021 | -0.6733 | 0.0385 | 27.6% | 0.51 [0.47, 0.55] | 2021 |
| Choi 2021 | -0.844 | 0.0117 | 29.1% | 0.43 [0.42, 0.44] | 2021 |
| **Total (95% CI)** | | | **100.0%** | **0.48 [0.41, 0.57]** | |

Heterogeneity: Tau² = 0.02; Chi² = 50.17, df = 3 (P < 0.00001); I² = 94%
Test for overall effect: Z = 8.60 (P < 0.00001)

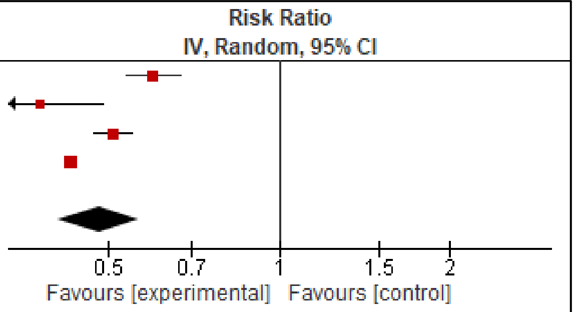

All adherence to screen papers: Coldman, Morrel, Duffy, Choi. The cohort adherence to screen: Morrel reported BC mortality for ever screened cohort (control group is never screened).

#### Table 4: Breast cancer mortality (Observational studies, stratified by age, stop screening analysis)

##### "Continue Screening" After Baseline Examination compared to "Stop Screening" After Baseline Examination

| Outcomes | Absolute effects: Baseline risk with stopping screening | Absolute effects: Absolute effect (95% CI) | Hazard ratio (95% CI) | № of participants (studies) | Quality of the evidence (GRADE) Clinical threshold of 0.5 | Quality of the evidence (GRADE) Clinical threshold of 1 | Comments |
|---|---|---|---|---|---|---|---|
| Sub-Group: Breast-Cancer Mortality (70-74 years)<br><br># of follow-up (yrs): 8.0 | **General population**<br>3.7 per 1,000 | **0.81 fewer per 1,000**<br>(from 0.19 to 1.37 fewer) | **HR 0.78**<br>(0.63 to 0.95) | 1235459<br>(1 study[1]) | ⨁◯◯◯<br>VERY LOW [a,b,c,d,e] | ⨁◯◯◯<br>VERY LOW [a,b,c,d,e,g] | Using a threshold of 0.5 or 1 fewer deaths per 1,000, we are very uncertain whether continuing screening decreases breast cancer mortality over 10 years for individuals aged 70 to 74 years in a general population. |
| Sub-Group: Breast-Cancer Mortality (75-84 years)<br><br># of follow-up (yrs): 8.0 | **General population**<br>3.7 per 1,000 | **0.0 fewer per 1,000**<br>(from 0.63 fewer to 0.70 more) | **HR 1.00**<br>(0.83 to 1.19) | 1403735<br>(1 study[1]) | ⨁◯◯◯<br>VERY LOW [a,b,c,d,e] | ⨁⨁◯◯<br>LOW [a,b,c,f,e,g] | Using a threshold of 0.5 fewer deaths per 1,000, we are very uncertain whether continuing screening decreases breast cancer mortality over 10 years for individuals aged 75 to 84 years in a general population.<br><br>Using a threshold of 1 fewer death per 1,000, continuing screening may make little to no difference in reducing breast cancer mortality over 10 years for individuals aged 75 to 84 years in a general population. |

**CI:** Confidence interval; **HR:** Hazard ratio

**Bibliography:** 1: Garcia-Albeniz 2020[19]

**GRADE Working Group grades of evidence**
**High quality:** We are very confident that the true effect lies close to that of the estimate of the effect
**Moderate quality:** We are moderately confident in the effect estimate: The true effect is likely to be close to the estimate of the effect, but there is a possibility that it is substantially different
**Low quality:** Our confidence in the effect estimate is limited: The true effect may be substantially different from the estimate of the effect
**Very low quality:** We have very little confidence in the effect estimate: The true effect is likely to be substantially different from the estimate of effect

#### Explanations

a. We did not downrate for risk of bias. Study was judged to be of moderate quality using the JBI critical appraisal tool for cohort studies.
b. We did not downrate for inconsistency (only one study included).
c. We did not downrate for indirectness. The study answers the question of stopping versus continuing screening and all patients have received at least one baseline mammography.
d. The 95% CI crosses the clinical decision threshold; therefore, we rated down once for imprecision.
e. We did not rate up for the magnitude of effect because the effect size did not meet the threshold for uprating. Following GRADE guidance, the RR is on the threshold of being considered a large effect (i.e., RR either >2.0 or <0.5 based on consistent evidence from at least 2 studies, with no plausible confounders).
f. The 95% CI does not cross the clinical decision threshold and we did not rate down for imprecision.
g. According to Egger et al.[11], 10 studies are needed to asses publication bias. We cannot assess publication bias due to insufficient number of trials, therefore, we did not rate down for publication bias.

#### Table 5: Breast cancer mortality (Observational case-control studies, stratified by age)

##### Screening with mammography* compared to no screening

| Outcomes | Absolute effects | | Range of relative effects (95% CI)** | № of participants (studies) | Quality of the evidence (GRADE) Clinical threshold of 0.5 | Quality of the evidence (GRADE) Clinical threshold of 1 | Comments |
|---|---|---|---|---|---|---|---|
| | **Risk with Usual Care (Assumed Risk) ‡** | **Absolute effect (95% CI)** | | | | | |
| Sub-Group: Breast-Cancer Mortality (40-49 years)<br><br>Range of follow-up (yrs): 11.0 to 38.0 | **General population** | | **OR 0.56** (0.49 to 0.64) | Unavailable (7 studies[1-7]) | ⊕◯◯◯ VERY LOW [a,b,c,d,e,f] | ⊕◯◯◯ VERY LOW [a,b,c,d,e,f] | Using a threshold of 0.5 or 1 fewer deaths per 1,000, we are very uncertain whether screening decreases breast cancer mortality over 10 years for individuals aged 40 to 49 years in a general population. |
| | 1.8 per 1,000 | **0.79 fewer per 1,000** (from 0.65 fewer to 0.92 fewer) | | | | | |
| | **Moderately increased risk** Based on family history (scaled adjustment) | | | | ⊕◯◯◯ VERY LOW [a,b,c,d,e,f] | ⊕◯◯◯ VERY LOW [a,b,c,d,e,f] | Using a threshold of 0.5 or 1 fewer deaths per 1,000, we are very uncertain whether screening decreases breast cancer mortality over 10 years for individuals aged 40 to 49 years at a moderately increased risk for breast cancer. |
| | 2.9 per 1,000 | **1.28 fewer per 1,000** (from 1.04 fewer to 1.48 fewer) | | | | | |
| | **Moderately increased risk** Based on dense breasts (scaled adjustment) | | | | ⊕◯◯◯ VERY LOW [a,b,c,d,e,f] | ⊕◯◯◯ VERY LOW [a,b,c,d,e,f] | Using a threshold of 0.5 or 1 fewer deaths per 1,000, we are very uncertain whether screening decreases breast cancer mortality over 10 years for individuals aged 40 to 49 years at a moderately increased risk for breast cancer. |
| | 3.5 per 1,000 | **1.54 fewer per 1,000** (1.26 fewer to 1.79 fewer) | | | | | |
| Sub-Group: Breast-Cancer Mortality (50-59 years)<br><br>Range of follow-up (yrs): 11.0 to 38.0 | **General population** | | **OR 0.56** (0.49 to 0.64) | Unavailable (7 studies[1-7]) | ⊕◯◯◯ VERY LOW [a,b,c,d,e,f] | ⊕◯◯◯ VERY LOW [a,b,c,d,e,f] | Using a threshold of 0.5 or 1 fewer deaths per 1,000, we are very uncertain whether screening decreases breast cancer mortality over 10 years for individuals aged 50 to 59 years in a general population. |
| | 3.3 per 1,000 | **1.45 fewer per 1,000** (from 1.19 fewer to 1.68 fewer) | | | | | |
| | **Moderately increased risk** Based on family history (scaled adjustment) | | | | ⊕◯◯◯ VERY LOW [a,b,c,d,e,f] | ⊕◯◯◯ VERY LOW [a,b,c,d,e,f] | Using a threshold of 0.5 or 1 fewer deaths per 1,000, we are very uncertain whether screening decreases breast cancer mortality over 10 years for individuals aged 50 to 59 years at a moderately increased risk for breast cancer. |
| | 5.3 per 1,000 | **2.33 fewer per 1,000** (from 1.91 fewer to 2.70 fewer) | | | | | |
| | **Moderately increased risk** Based on dense breasts (scaled adjustment) | | | | ⊕◯◯◯ VERY LOW [a,b,c,d,e,f] | ⊕◯◯◯ VERY LOW [a,b,c,d,e,f] | Using a threshold of 0.5 or 1 fewer deaths per 1,000, we are very uncertain whether screening decreases breast cancer mortality over 10 years for individuals aged 50 to 59 years at a moderately increased risk for breast cancer. |
| | 6.3 per 1,000 | **2.77 fewer per 1,000** (2.27 fewer to 3.21 fewer) | | | | | |
| Sub-Group: Breast-Cancer Mortality (60-69 years)<br><br>Range of follow-up (yrs): 11.0 to 38.0 | **General population** | | **OR 0.56** (0.49 to 0.64) | Unavailable (7 studies[1-7]) | ⊕◯◯◯ VERY LOW [a,b,c,d,e,f] | ⊕◯◯◯ VERY LOW [a,b,c,d,e,f] | Using a threshold of 0.5 or 1 fewer deaths per 1,000, we are very uncertain whether screening decreases breast cancer mortality over 10 years for individuals aged 60 to 69 years in a general population. |
| | 4.3 per 1,000 | **1.89 fewer per 1,000** (from 1.55 fewer to 2.19 fewer) | | | | | |
| | **Moderately increased risk** Based on family history (scaled adjustment) | | | | | | Using a threshold of 0.5 or 1 fewer deaths per 1,000, we are very uncertain whether screening decreases breast |

#### Screening with mammography* compared to no screening

| Outcomes | Absolute effects: Risk with Usual Care (Assumed Risk) ‡ | Absolute effects: Absolute effect (95% CI) | Range of relative effects (95% CI)** | № of participants (studies) | Quality of the evidence (GRADE) Clinical threshold of 0.5 | Quality of the evidence (GRADE) Clinical threshold of 1 | Comments |
|---|---|---|---|---|---|---|---|
| | 6.9 per 1,000 | **3.04 fewer per 1,000** (from 2.48 fewer to 3.52 fewer) | | | ⊕◯◯◯ VERY LOW [a,b,c,d,e,f] | ⊕◯◯◯ VERY LOW [a,b,c,d,e,f] | cancer mortality over 10 years for individuals aged 60 to 69 years at a moderately increased risk for breast cancer. |
| | **Moderately increased risk** Based on dense breasts (scaled adjustment) | | | | ⊕◯◯◯ VERY LOW [a,b,c,d,e,f] | ⊕◯◯◯ VERY LOW [a,b,c,d,e,f] | Using a threshold of 0.5 or 1 fewer deaths per 1,000, we are very uncertain whether screening decreases breast cancer mortality over 10 years for individuals aged 60 to 69 years at a moderately increased risk for breast cancer. |
| | 8.2 per 1,000 | **3.61 fewer per 1,000** (2.95 fewer to 4.18 fewer) | | | | | |
| Sub-Group: Breast-Cancer Mortality (70-74 years)<br><br>Range of follow-up (yrs): 11.0 to 38.0 | **General population** | | **OR 0.56** (0.49 to 0.64) | Unavailable (7 studies[1-7]) | ⊕◯◯◯ VERY LOW [a,b,c,d,e,f] | ⊕◯◯◯ VERY LOW [a,b,c,d,e,f] | Using a threshold of 0.5 or 1 fewer deaths per 1,000, we are very uncertain whether screening decreases breast cancer mortality over 10 years for individuals aged 70 to 74 years in a general population. |
| | 6.1 per 1,000 | **2.68 fewer per 1,000** (from 2.20 fewer to 3.11 fewer) | | | | | |
| | **Moderately increased risk** Based on family history (scaled adjustment) | | | | ⊕◯◯◯ VERY LOW [a,b,c,d,e,f] | ⊕◯◯◯ VERY LOW [a,b,c,d,e,f] | Using a threshold of 0.5 or 1 fewer deaths per 1,000, we are very uncertain whether screening decreases breast cancer mortality over 10 years for individuals aged 70 to 74 years at a moderately increased risk for breast cancer. |
| | 9.8 per 1,000 | **4.31 fewer per 1,000** (from 3.53 fewer to 5.0 fewer) | | | | | |
| | **Moderately increased risk** Based on dense breasts (scaled adjustment) | | | | ⊕◯◯◯ VERY LOW [a,b,c,d,e,f] | ⊕◯◯◯ VERY LOW [a,b,c,d,e,f] | Using a threshold of 0.5 or 1 fewer deaths per 1,000, we are very uncertain whether screening decreases breast cancer mortality over 10 years for individuals aged 70 to 74 years at a moderately increased risk for breast cancer. |
| | 11.6 per 1,000 | **5.10 fewer per 1,000** (4.18 fewer to 5.92 fewer) | | | | | |

‡The baseline risk represents the breast cancer mortality rate over 10 years in an unscreened group based on Canadian observational data reported by Coldman et al.[1] To calculate a moderately increased group risk, we used an estimate from Engmann et al.[2] suggesting that having a first degree relative increases the lifetime risk by 1.6 times and multiplied the general population baseline risk estimate by 1.6.
*Studies varied between film and digital mammography.
**Absolute risks were calculated using odds ratios (all adherence to screen exposure).
**CI:** Confidence interval; **OR:** Odds ratio

**Bibliography:** 1:Paap 2014[20], 2: Pocobelli 2015[21], 3. Massat 2016[22], 4: Ripping 2017[23], 5. van der Waal 2017[24], 6. Maroni 2021[25], 7. De Troeyer 2023[26]

**GRADE Working Group grades of evidence**
**High quality:** We are very confident that the true effect lies close to that of the estimate of the effect
**Moderate quality:** We are moderately confident in the effect estimate: The true effect is likely to be close to the estimate of the effect, but there is a possibility that it is substantially different
**Low quality:** Our confidence in the effect estimate is limited: The true effect may be substantially different from the estimate of the effect
**Very low quality:** We have very little confidence in the effect estimate: The true effect is likely to be substantially different from the estimate of effect

#### Explanations

a. We rated down once for risk of bias. Cases and controls were not age matched (De Troeyer and van der Waal) or failed to adjust for important confounding factors related to self-selection bias (De Troeyer, Maroni, Van der Waal, Massat, Pocobelli, Paap and Ripping). Several studies did not provide screening details or confirm all women were invited to screening (Massat, Pocobelli, Paap, Ripping). Average follow-up length not clearly reported across studies.
b. All individual estimates point to a reduction in BC mortality, so we did not downrate for inconsistency.
c. We did not downrate for indirectness since the studies used population-based approach and are reflective of general population.

d. Given the large sample sizes; an optimal sample size calculation was not warranted. The 95% CI does not cross the clinical decision threshold; therefore, we did not rate down for imprecision.
e. We did not rate up for the magnitude of effect because not all plausible confounders (e.g., age, hormone replacement therapy, breast density), were adjusted for, decreasing our confidence in the estimated effect. Following GRADE guidance, the RR is not considered a large effect (i.e., RR either >2.0 or <0.5 based on consistent evidence from at least 2 studies, with no plausible confounders).
f. According to Egger et al.[11], 10 studies are needed to asses publication bias. We cannot assess publication bias due to insufficient number of trials, therefore, we did not rate down for publication bias.

#### Table 5 forest plot

| Study or Subgroup | log[Odds Ratio] | SE | Weight | Odds Ratio IV, Random, 95% CI | Year |
|---|---|---|---|---|---|
| Paap 2014 | -0.8675 | 0.1187 | 12.6% | 0.42 [0.33, 0.53] | 2014 |
| Pocobelli 2015 | -0.7133 | 0.0589 | 17.8% | 0.49 [0.44, 0.55] | 2015 |
| Massat 2016 | -0.4943 | 0.1693 | 9.0% | 0.61 [0.44, 0.85] | 2016 |
| Ripping 2017 | -0.3425 | 0.1153 | 12.9% | 0.71 [0.57, 0.89] | 2017 |
| van der Waal 2017 | -0.3711 | 0.1299 | 11.7% | 0.69 [0.53, 0.89] | 2017 |
| Maroni 2021 | -0.478 | 0.0546 | 18.2% | 0.62 [0.56, 0.69] | 2021 |
| De Troeyer 2023 | -0.7133 | 0.0589 | 17.8% | 0.49 [0.44, 0.55] | 2023 |
| **Total (95% CI)** | | | **100.0%** | **0.56 [0.49, 0.64]** | |

Heterogeneity: $Tau^2 = 0.02$; $Chi^2 = 25.91$, df = 6 (P = 0.0002); $I^2 = 77\%$
Test for overall effect: Z = 8.61 (P < 0.00001)

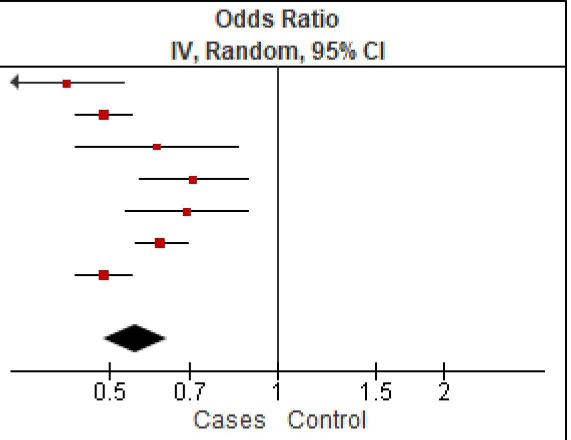

#### Table 6: Breast cancer mortality (Quasi-experimental, sub-groups)

##### Before-and-after BC screening program / Jurisdictions with or without BC screening program in 40-49 years

| Outcomes | Before BC screening implementation (N) | After BC screening implementation (N) | Absolute Effect | Relative Effects | № of participants (studies) | Certainty of evidence (GRADE) |
|---|---|---|---|---|---|---|
| Breast Cancer Mortality<br>Sub-group: 40-49 (Age)<br>Follow-up (yrs.): Unavailable**Error! Bookmark not defined.** | 0.2 per 1,000 person-years | 0.17 per 1,000 person-years | 0.03 fewer per 1,000 person-years | Unavailable | N=323719 (1 Study [A]) | ⊕◯◯◯<br>VERY LOW[1,5,6,7] |
| Breast Cancer Mortality<br>Sub-group: 40-49 (Age)<br>Follow-up (yrs.): 11 years | 0.15 per 1,000 person-years | 0.12 per 1,000 person-years | 0.03 fewer per 1,000 person-years | Unavailable | N= 40.7 million women-years (1 Study [B]) | ⊕◯◯◯<br>VERY LOW[2,5,6,7] |
| Breast Cancer Mortality<br>Sub-group: 50-59 (Age)<br>Follow-up (yrs.): Unavailable**Error! Bookmark not defined.** | 0.49 per 1,000 person-years | 0.36 per 1,000 person-years | 0.13 fewer per 1,000 person-years | Unavailable | N=323719 (1 Study [A]) | ⊕◯◯◯<br>VERY LOW[1,5,6,7] |
| Breast Cancer Mortality<br>Sub-group: 50-59 (Age)<br>Follow-up (yrs.): 11 years | 0.32 per 1,000 person-years | 0.34 per 1,000 person-years | 0.02 more per 1,000 person-years | Unavailable | N= 40.7 million women-years (1 Study [B]) | ⊕◯◯◯<br>VERY LOW[2,5,6,7] |
| Breast Cancer Mortality<br>Sub-group: 60-69 (Age)<br>Follow-up (yrs.): Unavailable**Error! Bookmark not defined.** | 0.80 per 1,000 person-years | 0.63 per 1,000 person-years | 0.17 fewer per 1,000 person-years | Unavailable | N=323719 (1 Study [A]) | ⊕◯◯◯<br>VERY LOW[1,5,6,7] |
| Breast Cancer Mortality<br>Sub-group: 70-79 (Age)<br>Follow-up (yrs.): Unavailable**Error! Bookmark not defined.** | 112/100,000 person-years<br>1.12 per 1,000 person-years | 114/100,000 person-years<br>1.14 per 1,000 person-years | 0.02 more per 1,000 person-years | Unavailable | N=323719 (1 Study [A]) | ⊕◯◯◯<br>VERY LOW[1,5,6,7] |
| Breast Cancer Mortality<br>Sub-group: 60-74 (Age)<br>Follow-up (yrs.): 11 years | 58/153,905 person-years<br>0.38 per 1,000 person-years | 98/166,317 person-years<br>0.59 per 1,000 person-years | 0.21 more per 1,000 person-years | Unavailable | N= 40.7 million women-years (1 Study [B]) | ⊕◯◯◯<br>VERY LOW[2,5,6,7] |
| Breast Cancer Mortality<br>Sub-group: 75-84 (Age)<br>Follow-up (yrs.): 11 years | 0.72 per 1,000 person-years | 0.84 per 1,000 person-years | 0.12 more per 1,000 person-years | Unavailable | N= 40.7 million women-years (1 Study) [B] | ⊕◯◯◯<br>VERY LOW[2,5,6,7] |

| Outcomes | | | Absolute Effect | Relative Effects | № of participants (studies) | Certainty of evidence (GRADE) |
|---|---|---|---|---|---|---|
| | **Before BC screening implementation (N)** | **After BC screening implementation (N)** | | | | |
| Incidence of fatal breast cancer within 10 years of diagnosis<br><br>Sub-group: Comparison made during the active screening period (1977 to 2015)<br><br>Follow-up (yrs.): 10 years | Women who were invited and did not participate in screening during the screening period:<br><br>0.62 per 1,000 person-years | Women who were invited and participated in screening during the screening period:<br><br>0.25 per 1,000 person-years | 0.37 fewer per 1,000 person-years | Relative Risk: **0.40** (0.34 to 0.48) | N=52,438 (Mean no. of women aged 40 to 69 years); (1 study) [C] | ⊕◯◯◯ VERY LOW[3,5,6,7] |
| Incidence of fatal breast cancer within 10 years of diagnosis<br><br>Time-trend analysis looking at those who did not have the opportunity to screen in the pre-screening period (1958 to 1976) compared to those who were invited and participated during the active screening period (1977 to 2015)<br><br>Follow-up (yrs.): 10 years | 0.55 per 1,000 person-years | 0.25 per 1,000 person-years | 0.30 fewer per 1,000 person-years | Relative Risk: **0.46** (0.39 to 0.53) | N=52,438 (Mean no. of women aged 40 to 69 years); (1 study) [C] | ⊕◯◯◯ VERY LOW[3,5,6,7] |
| Incidence-based BC mortality rate ratio<br><br>Subgroup: 40-49 years<br><br>Follow-up (yrs.): 10 years | Provincial/territorial mammography screening programs not including women aged 40-49 years:<br><br>NR | Provincial/territorial mammography screening programs including women aged 40-49 years:<br><br>NR | NR | Rate Ratio: 0.92; 95% CI, 0.85 to 0.99 | N=21,103 [D] | ⊕◯◯◯ VERY LOW[4,5,6,8] |

**Bibliography:**

A: Katalinic 2020[27]    B: Parvinen 2015[28]    C: Tabar 2019[29]    D. Wilkinson 2023[30]

### Explanations

1. We did not downrate for RoB. Study assessed at low risk of bias (RoB score for JBI Quasi-experimental tool=7/9). Study reported no data on reliability of outcomes and average follow-up period.
2. We downrated once for RoB. Study at moderate risk of bias (RoB score for JBI Quasi-experimental tool=5/9). Different number of participants across comparative groups. No information on lost to follow-up participants. No data on reliability of outcome measures.
3. We downrated once for RoB. Study at moderate risk of bias (RoB score for JBI Quasi-experimental tool=6/9). No information on control group, loss-to follow-up patients and reliability of outcome measures
4. We did not downrate for RoB. Study at low risk of bias (RoB score for JBI Quasi-experimental tool=7/9). Noted that there may be differences in the participants and access to care/treatment across screening and non-screening jurisdictions beyond screening that could impact survival differences.
5. Unable to evaluate imprecision using thresholds, as the baseline rates were not available to allow the calculation of absolute effects. Therefore, a minimally contextualized approach was used. The total population is large (>2000) and there is a large event rate (>300). We did not downrate for imprecision.
6. Not downrated for inconsistency. Single study evaluated outcome (unable to evaluate heterogeneity).
7. Downrated once for indirectness. Pre-screening periods ranged across studies between 1958 and 2004. There are population-level differences that may affect mortality beyond the introduction of mammography screening between the pre-screening period and the post-screening period.
8. Downrated once for indirectness. Study assessed the effect of screening programs on outcomes of interest, rather than the effect of individual-level mammography screening. Not all women in screening jurisdictions participated in screening and it is unknown if BCs were diagnosed by screening or through other means (e.g., interval cancers, symptoms).

#### Table 7: All-cause mortality (RCTs, stratified by age, over 10 years)

##### Mammography +/- CBE compared to Usual Care

| Outcomes | Absolute effects | | Relative effect § (95% CI) | № of participants* (studies) | Quality of the evidence (GRADE) | Comments |
|---|---|---|---|---|---|---|
| | **Risk with Usual Care (Assumed Risk) ‡** | **Absolute effect (95% CI)** | | | | |
| Sub-Group: All-Cause Mortality (40-49 years)<br><br># Randomised: 311,066<br># Analyzed: Unclear<br><br>Range of follow-up (yrs): 7.9 to 17.7 | 12.7 per 1,000 | **0.13 fewer per 1,000** (from 0 fewer to 0.25 fewer) | **RR 0.99** (0.98 to 1.00) | Unavailable (7 RCTs[1-6, 8]) [a] | ⊕⊕◯◯ LOW [b,c,d,e,f] | Using a threshold of 1 fewer death per 1,000, screening may make little to no difference in reducing mortality from any cause over 10 years for individuals aged 40 to 49 years. |
| Sub-Group: All-Cause Mortality (50-59 years)<br><br># Randomised: 79,749<br># Analyzed: 79,695<br><br>Range of follow-up (yrs): 7.9 to 13.0 | 30.6 per 1,000 | **0.31 fewer per 1,000** (from 0 fewer to 0.61 fewer) | **RR 0.99** (0.98 to 1.00) | 79,695 (3 RCTs [3,4,7]) | ⊕⊕◯◯ LOW [b,c,d,e,f] | Using a threshold of 1 fewer death per 1,000, screening may make little to no difference in reducing mortality from any cause over 10 years for individuals aged 50 to 59 years. |
| Sub-Group: All-Cause Mortality (60-69 years)<br><br># Randomised: 39,681<br># Analyzed: 39,681<br><br>Range of follow-up (yrs): 7.9 | 71.3 per 1,000 | **0.71 fewer per 1,000** (from 0 fewer to 1.43 fewer) | **RR 0.99** (0.98 to 1.00) | 39,681 (2 RCTs [3,4]) | ⊕◯◯◯ VERY LOW [b,c,d,f,g] | Using a threshold of 1 fewer death per 1,000, we are very uncertain whether screening decreases mortality from any cause over 10 years for individuals aged 60 to 69 years. |
| Sub-Group: All-Cause Mortality (70-74 years)<br><br># Randomised: 17,646<br># Analyzed: 17,646<br><br>Range of follow-up (yrs): 7.9 | 140.6 per 1,000 | **1.41 fewer per 1,000** (from 0 fewer to 2.81 fewer) | **RR 0.99** (0.98 to 1.00) | 17,646 (2 RCTs [3,4]) | ⊕◯◯◯ VERY LOW [b,c,d,f,g] | Using a threshold of 1 fewer death per 1,000, we are very uncertain whether screening decreases mortality from any cause over 10 years for individuals aged 70 to 74 years. |

‡The baseline risk has been calculated using deaths and age-specific mortality rates data from Statistics Canada and estimated over a 10-year period. (https://www150.statcan.gc.ca/t1/tbl1/en/tv.action?pid=1310039201&pickMembers%5B0%5D=2.11&pickMembers%5B1%5D=3.3&cubeTimeFrame.startYear=2017&cubeTimeFrame.endYear=2021&referencePeriods=20170101%2C20210101)
§ Following the same logic as breast cancer mortality, the relative effect is based on our previous systematic review and guideline[4] where a subgroup analysis of relative risk by age was assessed and no difference in RR among subgroups was detected and true differences resulting from age were deemed unlikely. Therefore, we used the RR for all ages rather than focusing on each decade of age as we had previously done in our 2018 guideline.
*The number of participants and studies reflect the previous analysis for each age decade, rather than the number of studies that are included in the relative effect estimate for all ages.
**CI:** Confidence interval; **RR:** Risk ratio

#### Mammography +/- CBE compared to Usual Care

| Outcomes | Absolute effects | | Relative effect § (95% CI) | № of participants* (studies) | Quality of the evidence (GRADE) | Comments |
|---|---|---|---|---|---|---|
| | **Risk with Usual Care (Assumed Risk) ‡** | **Absolute effect (95% CI)** | | | | |

**GRADE Working Group grades of evidence**
**High quality:** We are very confident that the true effect lies close to that of the estimate of the effect
**Moderate quality:** We are moderately confident in the effect estimate: The true effect is likely to be close to the estimate of the effect, but there is a possibility that it is substantially different
**Low quality:** Our confidence in the effect estimate is limited: The true effect may be substantially different from the estimate of the effect
**Very low quality:** We have very little confidence in the effect estimate: The true effect is likely to be substantially different from the estimate of effect

**Bibliography:**

1: Age (Moss 2015[6])
2. Malmo II (Nystrom 2002[8])
3: Swedish Two County (Kopparberg) (Tabar 1989[31])
4: Swedish Two County (Ostergotland) (Tabar 1989[31])
5: Stockholm (Frisell 1997[32])
6: CNBSS 1 (Miller 2002[33])
7: CNBSS 2 (Miller 2000[34])
8: Gothenburg (Bjurstam 1997[35])

#### Explanations

a. Two studies considered quasi-randomised (Stockholm & Gothenburg)
b. Randomisation and allocation concealment were either not reported or there were serious deficiencies in these areas, we downrated once for risk of bias.
c. All point estimates in our pooled analysis lie to one side of our threshold. We did not rate down for inconsistency.
d. Breast density was not addressed. For some studies, the control group received screening after the screening period. Studies reported in Nystrom 2002 and Nystrom 2016 included one round of screening in the control group as part of the short-case accrual calculation. Therefore, the study estimates may be underestimated (a larger benefit from the intervention may be possible). Downrated once for indirectness. Data are from trials initiated in the 1960s-1990s and the intervention groups were primarily screened with film mammography. Due to advances in mammography technology and treatment practices, we expect that the magnitude of screening effect may differ if applied to today's Canadian screening context. There are no high-quality clinical trials examining the impact of screening on breast cancer screening deaths using contemporary screening methods.
e. Not downrated for imprecision i) The number of events and total population are large (>300 threshold for events); and (ii) the 95%CIs include the null, but do not cross clinical decision threshold (1 fewer or 1 more). Given the large sample sizes, an optimal sample size calculation was not warranted.
f. According to Egger et al.[11], 10 trials are needed to asses publication bias. We cannot assess publication bias due to insufficient number of trials, therefore, we did not rate down for publication bias.
g. Downrated once for imprecision. i) The number of events and total population are large (>300 threshold for events); and (ii) the 95%CIs include the null and cross the clinical decision threshold (1 fewer or 1 more). Given the large sample sizes, an optimal sample size calculation was not warranted.

#### Table 7 forest plot

| Reference | Study | Age (at entry) | Mean Follow-up (yrs) | log [RR] | SE | Weight | Risk Ratio [95%CI] |
|---|---|---|---|---|---|---|---|
| Aron and Prorok 1986 | HIP | 40-59 | 10.0 | -0.01 | 0.03 | 4.7% | 0.99 [0.93, 1.05] |
| Miller 2014 | CNBSS 1& 2 | 40-59 | 25.0 | 0.02 | 0.02 | 11.4% | 1.02 [0.98, 1.06] |
| Moss 2015 | AGE | 39-41 | 17.7[a] | -0.02 | 0.03 | 6.7% | 0.98 [0.93, 1.03] |
| Nystrom 2002 | Gothenburg | 40-59 | 13.2 | -0.06 | 0.03 | 4.3% | 0.94 [0.88, 1.00] |
| Nystrom 2002 | Malmo I | 45-70 | 19.2 | -0.01 | 0.01 | 42.8% | 0.99 [0.97, 1.01] |
| Nystrom 2002 | Malmo II | 43-49 | 9.1 | 0.03 | 0.08 | 0.8% | 1.03 [0.89, 1.20] |
| Nystrom 2002 | Ostergotland | 40-74 | 17.2 | -0.02 | 0.02 | 18.6% | 0.98 [0.95, 1.01] |
| Nystrom 2002 | Stockholm | 40-64 | 14.7 | -0.01 | 0.02 | 10.7% | 0.99 [0.95, 1.03] |
| **Total (95% CI)** | | | | | | **100.0%** | **0.99 [0.98, 1.00]** |

Heterogeneity: $Tau^2 = 0.00$; $Chi^2 = 5.57$, df = 7 (P = 0.59); $I^2 = 0\%$

Test for overall effect: Z = 1.65 (P = 0.10)

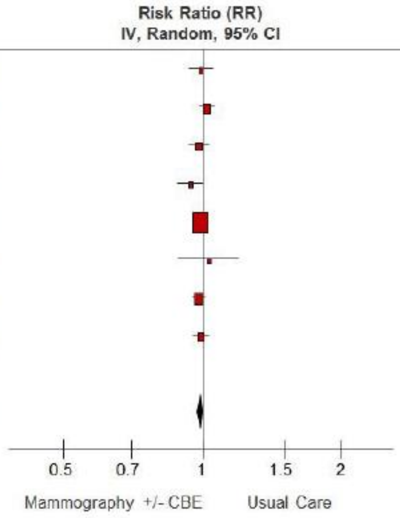

[a]Median

#### Table 8: Stage at diagnosis (RCTs, all ages)

##### Screening with film mammography (with or without CBE) compared to usual care

| Outcomes | Absolute Effects | | Relative effect (95% CI) | № of participants (studies) | Quality of the evidence (GRADE) | Comments |
|---|---|---|---|---|---|---|
| | **Risk with Usual Care (Assumed Risk)** | **Absolute effect (95% CI)** | | | | |
| Invasive Breast Cancer Diagnosed at Stage II or higher (all ages)*<br><br>Range of follow-up (yrs): 5.0 to 10.0 | 9.1 per 1000 | **3 fewer per 1,000** (from 5 fewer to 1 more) | **RR 0.72**<br>(0.49 to 1.06) | Unclear<br>(5 RCTs[1-5]) | ⊕◯◯◯<br>VERY LOW [a,b,c,d,e,l] | Using a threshold of 3 fewer breast cancers diagnosed at stage II or higher per 1,000, we are very uncertain whether screening decreases the number of individuals with stage II+ at diagnosis in those at general population risk for breast cancer (all ages). |
| Invasive Breast Cancer Diagnosed at Stage II or higher (Ages 40-49 years)*<br><br>Follow-up (yrs): 7.0 | 2.6 per 1000 | **1 more per 1,000** (from 1 more to 3 more) | **RR 1.55**<br>(1.23 to 2.11) | Unclear<br>(1 RCT[3]) | ⊕◯◯◯<br>VERY LOW [f,g,d,h,l] | Using a threshold of 3 fewer breast cancers diagnosed at stage II or higher per 1,000, we are very uncertain whether screening makes little to no difference on the number of individuals with stage II+ at diagnosis in those at general population risk for breast cancer (40-49 years). |
| Invasive Breast Cancer Diagnosed at Stage II or higher (Ages 50-59 years)*<br><br>Follow-up (yrs): 7.0 | 4.6 per 1000 | **0 fewer per 1,000** (from 1 fewer to 2 more) | **RR 1.09**<br>(0.82 to 1.45) | Unclear<br>(1 RCT[3]) | ⊕◯◯◯<br>VERY LOW [f,g,d,h,l] | Using a threshold of 3 fewer breast cancers diagnosed at stage II or higher per 1,000, we are very uncertain whether screening makes little to no difference on the number of individuals with stage II+ at diagnosis in those at general population risk for breast cancer (50-59 years). |
| Invasive Breast Cancer Diagnosed at Stage III or higher (all ages)*<br><br>Range of follow-up (yrs): 5.0 to 10.0 | 2.2 per 1000 | **1 fewer per 1,000** (from 1 fewer to 0 fewer) | **RR 0.64**<br>(0.47 to 0.88) | Unclear<br>(3 RCTs[2,4,5]) | ⊕◯◯◯<br>VERY LOW [i,j,d,k,l] | Using a threshold of 2 fewer breast cancers being diagnosed at stage III or higher per 1,000, we are very uncertain whether screening makes little to no difference on the number of individuals with stage III+ at diagnosis in those at general population risk for breast cancer (all ages). |

*Rates calculated using number of participants with stage II+ or stage III+ reported in Tarone 1995 for included trials and the number of participants randomized in each trial.
**CI:** Confidence interval; **RR:** Risk ratio

**Bibliography:**
1: Swedish Two County (Kopparberg & Ostergotland) (Tarone 1995[36]) 2: Malmo I (Tarone 1995[36]) 3: CNBSS 1 (Tarone 1995[36]) 4: HIP (Tarone 1995[36]) 5: Stockholm (Tarone 1995[36])

**GRADE Working Group grades of evidence**
**High quality:** We are very confident that the true effect lies close to that of the estimate of the effect
**Moderate quality:** We are moderately confident in the effect estimate: The true effect is likely to be close to the estimate of the effect, but there is a possibility that it is substantially different
**Low quality:** Our confidence in the effect estimate is limited: The true effect may be substantially different from the estimate of the effect
**Very low quality:** We have very little confidence in the effect estimate: The true effect is likely to be substantially different from the estimate of effect

Explanations

a. One study considered quasi-randomised (Stockholm)
b. Downrated once for risk of bias. Randomisation and allocation concealment were either not reported sufficiently (Malmo I, HIP) or there were serious deficiencies in these areas (CNBSS-I, Stockholm).
c. Approximately half of the point estimates in our pooled analysis lie on either side of our threshold. We rated down once for inconsistency.
d. Data are from trials initiated in the 1960s-1990s and the intervention groups were primarily screened with film mammography. Due to advances in mammography technology and treatment practices, we expect that the magnitude of screening effect may differ if applied to today's Canadian screening context. There are no high-quality clinical trials examining the impact of screening on breast cancer screening deaths using contemporary screening methods.
e. Downrated once for imprecision. CI crosses threshold for benefit of breast cancer screening for proportion of patients diagnosed at stage II or higher.
f. Downrated once for risk of bias. High risk of bias due to concerns with randomisation method and allocation concealment (CNBSS I).
g. Not downrated for inconsistency. Single study evaluated outcome.
h. Downrated once for imprecision. Low number of events (fewer than 300) and confidence interval crosses threshold for harm.
i. Downrated once for risk of bias. High risk of bias due to risk of bias in randomization and allocation concealment (Stockholm) and use of local endpoint committee for blinding of outcomes (HIP).
j. All point estimates in our pooled analysis lie to one side of our threshold. We did not rate down for inconsistency.
k. Did not downrate for imprecision. Large population and CI does not cross below the threshold for benefit of breast cancer screening for proportion of population diagnosed at stage III.
l. According to Egger et al.[11], 10 trials are needed to asses publication bias. We cannot assess publication bias due to insufficient number of trials, therefore, we did not rate down for publication bias.

#### Table 8 forest plots

Breast Cancer Diagnosis at stage II or higher

| Study or Subgroup | Mammography Events | Mammography Total | Usual Care Events | Usual Care Total | Weight | Risk Ratio M-H, Random, 95% CI |
|---|---|---|---|---|---|---|
| Tarone (CNBSS I& II) 1995 | 200 | 44925 | 156 | 44910 | 19.8% | 1.28 [1.04, 1.58] |
| Tarone (HIP) 1995 | 162 | 30239 | 190 | 30765 | 19.8% | 0.87 [0.70, 1.07] |
| Tarone (Malmo I) 1995 | 190 | 21088 | 231 | 21195 | 20.0% | 0.83 [0.68, 1.00] |
| Tarone (Stockholm) 1995 | 173 | 39139 | 210 | 20978 | 19.9% | 0.44 [0.36, 0.54] |
| Tarone (Swedish Two (Kopparbeg/Ostergotland)) 1995 | 460 | 38589 | 453 | 18582 | 20.5% | 0.49 [0.43, 0.56] |
| **Total (95% CI)** | | **173980** | | **136430** | **100.0%** | **0.72 [0.49, 1.06]** |
| Total events | 1185 | | 1240 | | | |

Heterogeneity: Tau² = 0.18; Chi² = 87.11, df = 4 (P < 0.00001); I² = 95%
Test for overall effect: Z = 1.68 (P = 0.09)

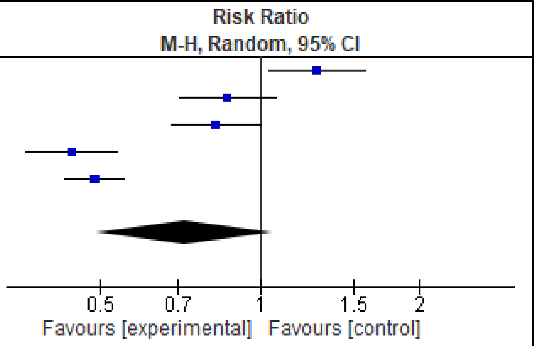

Breast Cancer Diagnosis at stage III or higher

| Study or Subgroup | Mammography Events | Mammography Total | Usual Care Events | Usual Care Total | Weight | Risk Ratio M-H, Random, 95% CI |
|---|---|---|---|---|---|---|
| Tarone (HIP) 1995 | 42 | 30239 | 65 | 30765 | 35.3% | 0.66 [0.45, 0.97] |
| Tarone (Malmo I) 1995 | 48 | 21088 | 59 | 21195 | 36.0% | 0.82 [0.56, 1.20] |
| Tarone (Stockholm) 1995 | 34 | 39139 | 39 | 20978 | 28.7% | 0.47 [0.30, 0.74] |
| **Total (95% CI)** | | **90466** | | **72938** | **100.0%** | **0.64 [0.47, 0.88]** |
| Total events | 124 | | 163 | | | |

Heterogeneity: Tau² = 0.03; Chi² = 3.38, df = 2 (P = 0.18); I² = 41%
Test for overall effect: Z = 2.81 (P = 0.005)

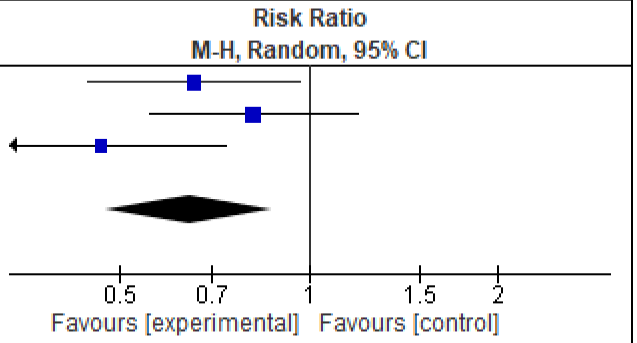

#### Table 9: Stage at diagnosis (Observational studies, all ages)

##### Screening with mammography* compared to no screening

| Outcomes | Absolute effects | | Relative effect (95% CI) | № of participants (studies) | Quality of the evidence (GRADE) | Comments |
|---|---|---|---|---|---|---|
| | **Risk with Usual Care (Assumed Risk)** | **Absolute effect (95% CI)** | | | | |
| Distant degree of spread at diagnosis | NR | **Not estimable**** | **RR 0.44 (0.37 to 0.52)** | 869,857 (1 study[1]) | ⊕◯◯◯ VERY LOW [a,b,c,d,e] | We are very uncertain about if screening with mammography compared to no screening reduces the proportion of individuals with distant degree of breast cancer spread at diagnosis |
| Stage II+ at diagnosis | 1.81 per 1000 | 0.51 fewer per 1000 (0.43 fewer to 0.58 fewer) | **Incidence Rate ratio 0.72 (0.68 to 0.76)** | 413,447 (1 study[2]) | ⊕◯◯◯ VERY LOW [b,c,e f,g,] | Using a threshold of 3 fewer breast cancers being diagnosed at stage II or higher per 1,000, we are very uncertain whether screening makes little to no difference in the number of individuals with stage II+ at diagnosis in those at general population risk for breast cancer. |

***The corresponding risk** (and its 95% confidence interval) is based on **the assumed risk** in the comparison group and the **relative effect** of the intervention (and its 95% CI).
**Study did not provide baseline risk values for usual care or breast cancer screening groups to calculate absolute risk.
**CI:** Confidence interval; **RR:** Risk ratio

**Bibliography:**
1: Morrell 2017[18]
2: Puliti 2017[37]

**GRADE Working Group grades of evidence**
**High quality:** We are very confident that the true effect lies close to that of the estimate of the effect
**Moderate quality:** We are moderately confident in the effect estimate: The true effect is likely to be close to the estimate of the effect, but there is a possibility that it is substantially different
**Low quality:** Our confidence in the effect estimate is limited: The true effect may be substantially different from the estimate of the effect
**Very low quality:** We have very little confidence in the effect estimate: The true effect is likely to be substantially different from the estimate of effect

#### Explanations

a. Downrated twice for risk of bias. Study at high risk of bias (RoB score for JBI cohort tool=4/11). Non-screening population inferred from census-derived population data rather than individual data and lack of reporting on outcome measurement. Lack of adjustment for important confounding factors (use of HRT, breast density). Unclear report of average follow-up time for population and no description of number of women lost to follow-up.
b. Not downrated for inconsistency. Single study evaluated outcome (unable to evaluate heterogeneity).
c. Not downrated for indirectness. Studies used population-based approach which was reflective of general population
d. Not downrated for imprecision. Unable to calculate absolute effects to determine if benefit for threshold is crossed, so a minimally contextualized approach was used. The total population is large (>2000) and there is a large event rate (>300). Given the large sample sizes and that the confidence interval does not include the null value, an optimal sample size calculation is not warranted.
e. According to Egger et al.[11], 10 studies are needed to asses publication bias. We cannot assess publication bias due to insufficient number of trials, therefore, we did not rate down for publication bias.
f. Downrated once for risk of bias. Study at moderate risk of bias (RoB score for JBI cohort tool=7/11). Lack of adjustment for important confounding factors (use of HRT, breast density). No description of number of women lost to follow-up.
g. Not downrated for imprecision. Large population and CI does not cross threshold for breast cancer screening benefit for stage III at diagnosis.

#### Table 10: Stage distribution of Breast Cancer (Quasi-experimental, Sub-groups)

##### Before-and-after BC screening program implementation/ Jurisdictions with or without BC screening program in 40-49 years

| Outcomes | Rates | | Absolute Effect | Relative effect (95% CI) | № of participants (studies) | Risk of bias (Score) |
|---|---|---|---|---|---|---|
| | **Before BC screening implementation (N)** | **After BC screening implementation (N)** | | | | |
| Advanced stage defined as stages III and IV as per the TNM classification<br><br>Sub-group: 70-75 years (Screening uptake period; 1998-2002)[2]<br><br>Follow-up (yrs.): Unavailable | 0.59 per 1,000 person-years | 0.46 per 1,000 person-years | 0.13 fewer per 1,000 person-years | Incidence Rate Ratio: **0.79**[1] (0.71 to 0.87) | N= 38442 (1 study)[A] | ⊕◯◯◯ VERY LOW [3,6,7,8] |
| Advanced stage defined as stages III and IV as per the TNM classification<br><br>Sub-group: 76-80 years (Screening uptake period; 1998-2002)[2]<br><br>Follow-up (yrs.): Unavailable | 0.66 per 1,000 person-years | 0.69 per 1,000 person-years | 0.03 more per 1,000 person-years | Incidence Rate Ratio: **1.04**[1] (0.94 to 1.17) | N= 38442 (1 study)[A] | ⊕◯◯◯ VERY LOW [3,6,7,8] |
| Advanced stage defined as stages III and IV as per the TNM classification<br><br>Sub-group: 70-75 years (Screening uptake period; 2003-2011)[2]<br><br>Follow-up (yrs.): Unavailable | 0.59 per 1,000 person-years | 0.52 per 1,000 person-years | 0.07 fewer per 1,000 person-years | Incidence Rate Ratio: **0.88**[1] (0.81 to 0.97)[1] | N= 38442 (1 study)[A] | ⊕◯◯◯ VERY LOW [3,6,7,8] |
| Advanced stage defined as stages III and IV as per the TNM classification<br><br>Sub-group: 76-80 years (Screening uptake period; 2003-2011)[2]<br><br>Follow-up (yrs.): Unavailable | 0.66 per 1,000 person-years | 0.67 per 1,000 person-years | 0.01 more per 1,000 person-years | Incidence Rate Ratio: **1.02**[1] (0.92 to 1.13) | N= 38442 (1 study)[A] | ⊕◯◯◯ VERY LOW [3,6,7,8] |
| Sub-group: Late stage (Regional)<br>Age group: Women aged ≥40 years (all ages)<br>Follow-up (yrs.): Unavailable | 0.87 per 1,000 person-years | 0.77 per 1,000 person-years | 0.10 fewer per 1,000 person-years | Unavailable | Unavailable**Error! Bookmark not defined.** (1 Study)[B] | ⊕◯◯◯ VERY LOW [4,6,7,8] |

| Outcomes | Rates | | Absolute Effect | Relative effect (95% CI) | № of participants (studies) | Risk of bias (Score) |
|---|---|---|---|---|---|---|
| | **Before BC screening implementation (N)** | **After BC screening implementation (N)** | | | | |
| Sub-group: Late stage (Distant)<br>Age group: Women aged ≥40 years (all ages)<br>Follow-up (yrs.): Unavailable | 0.17 per 1,000 person-years | 0.18 per 1,000 person-years | 0.01 more per 1,000 person-years | Unavailable | Unavailable**Error! Bookmark not defined.** (1 Study) [B] | ⊕◯◯◯<br>VERY LOW [4,6,7,8] |
| Proportion of BC diagnosed at Stage II<br><br>Subgroup: 40-49 years | Jurisdictions without organised screening programs for women 40–49 with annual recall:<br><br>437 per 1,000 | Jurisdictions with organised screening programs for women 40–49 with annual recall:<br><br>407 per 1,000 | 30 fewer per 1,000 | Unavailable<br>p < 0.001 | Unavailable (1 Study) [C] | ⊕◯◯◯<br>VERY LOW [5,6,7,9] |
| Proportion of BC diagnosed at Stage III<br><br>Subgroup: 40-49 years | Jurisdictions without organised screening programs for women 40–49 with annual recall:<br><br>183 per 1,000 | Jurisdictions with organised screening programs for women 40–49 with annual recall:<br><br>156 per 1,000 | 27 fewer per 1,000 | Unavailable<br>p < 0.001 | Unavailable (1 Study) [C] | ⊕◯◯◯<br>VERY LOW [5,6,7,9] |
| Proportion of BC diagnosed at Stage IV<br><br>Subgroup: 40-49 years | Jurisdictions without organised screening programs for women 40–49 with annual recall:<br><br>46 per 1,000 | Jurisdictions with organised screening programs for women 40–49 with annual recall:<br><br>39 per 1,000 | 7 fewer per 1,000 | Unavailable<br>p = 0.001 | Unavailable (1 Study) [C] | ⊕◯◯◯<br>VERY LOW [5,6,7,9] |
| Proportion of BC diagnosed at Stage II<br><br>Subgroup: 50-59 years | Jurisdictions without organised screening programs for women 40–49 with annual recall:<br><br>372 per 1,000 | Jurisdictions with organised screening programs for women 40–49 with annual recall:<br><br>360 per 1,000 | 12 fewer per 1,000 | Unavailable<br>p = 0.003 | Unavailable (1 Study) [C] | ⊕◯◯◯<br>VERY LOW [5,6,7,9] |

| Outcomes | Rates | | Absolute Effect | Relative effect (95% CI) | № of participants (studies) | Risk of bias (Score) |
|---|---|---|---|---|---|---|
| | **Before BC screening implementation (N)** | **After BC screening implementation (N)** | | | | |
| Proportion of BC diagnosed at Stage III<br><br>Subgroup: 50-59 years | Jurisdictions without organised screening programs for women 40–49 with annual recall:<br><br>136 per 1,000 | Jurisdictions with organised screening programs for women 40–49 with annual recall:<br>123 per 1,000 | 13 fewer per 1,000 | Unavailable $p < 0.001$ | Unavailable (1 Study) [C] | ⨁◯◯◯<br>VERY LOW [5,6,7,9] |
| Proportion of BC diagnosed at Stage IV<br><br>Subgroup: 50-59 years | Jurisdictions without organised screening programs for women 40–49 with annual recall:<br>NR | Jurisdictions with organised screening programs for women 40–49 with annual recall:<br>NR | NR | Unavailable | Unavailable (1 Study) [C] | ⨁◯◯◯<br>VERY LOW [5,6,7,9] |

**Bibliography:**

A: de Glas 2014[38]

B: Helvie 2014[39]

C. Wilkinson 2022[40]

#### Explanations

1. Unadjusted incidence rate ratio (IRR)
2. Comparison of screening period to pre-screening period of 1995-1997.
3. Study at Low risk of bias (RoB score for JBI Quasi-experimental tool=7/9). No information on loss-to follow-up patients and reliability of outcome measures.
4. Study at moderate risk of bias (RoB score for JBI Quasi-experimental tool=6/9). No information on control group, loss-to follow-up patients and reliability of outcome measures
5. Study at low risk of bias (RoB score for JBI Quasi-experimental tool=8/9). Noted that there may be differences in access to care across screening and non-screening jurisdictions beyond screening that could impact the stage of BC diagnosis.
6. Unable to evaluate imprecision using thresholds, as the baseline rates were not available to allow the calculation of absolute effects. Therefore, a minimally contextualized approach was used. The total population is large (>2000) and there is a large event rate (>300). We did not downrate for imprecision.
7. Not downrated for inconsistency. Single study evaluated outcome (unable to evaluate heterogeneity).
8. Downrated once for indirectness. Pre-screening periods ranged across studies between 1958 and 2004. There are population-level differences that may affect mortality beyond the introduction of mammography screening between the pre-screening period and the post-screening period.
9. Downrated once for indirectness. Study assessed the effect of screening programs on outcomes of interest, rather than the effect of individual-level mammography screening. Not all women in screening jurisdictions participated in screening and it is unknown if BCs were diagnosed by screening or through other means (e.g., interval cancers, symptoms).

#### Table 11: Overdiagnosis over 10 years (RCTs, stratified by age)

##### Mammography +/- CBE compared to Usual Care

| Outcomes | Absolute effects | | Relative effect (95% CI) | № of participants (studies) | Quality of the evidence (GRADE) | Comments |
|---|---|---|---|---|---|---|
| | Incident rates with usual care (Assumed rate) ‡ | Absolute risk (95% CI) | | | | |
| **Main analysis:** Overdiagnosis invasive + in situ cancers (40-49 years)<br><br>Range of follow-up (yrs): 9 to 15 | 17.7 per 1,000 | **1.95 more per 1,000** (from 0.89 more to 3.01 more) | **RR 1.11** (1.05 to 1.17) | 293,152 (3 [1-3] RCTs) [a] | ⊕◯◯◯ VERY LOW [a,b,c,d,e] | Using a threshold of 5, we are very uncertain whether screening leads to at least 5 overdiagnosed cancers in individuals aged 40 to 49 years. |
| Other analysis: Overdiagnosis invasive cancers only (40-49 years)<br><br>Range of follow-up (yrs): 9 to 15 | 16.7 per 1,000 | **1 more per 1,000** (from 0 to 2 more) | **RR 1.06** (1.00 to 1.12) | 293,152 (3 [1-3] RCTs) [a] | ⊕⊕◯◯ LOW [a,c,d,e,f] | Using a threshold of 5, screening may lead to little to no difference in overdiagnosed invasive cancers in individuals aged 40 to 49 years. |
| **Main analysis:** Overdiagnosis invasive + in situ cancers (50-59 years)<br><br>Range of follow-up (yrs): 10 to 15 | 24.1 per 1,000 | **1.93 more per 1,000** (from 0.24 more to 3.86 more) | **RR 1.08** (1.01 to 1.16) | 132,231 (2 [1,2] RCTs) [a] | ⊕⊕◯◯ LOW [a,c,d,e,f] | Using a threshold of 5, screening may lead to little to no difference in overdiagnosed cancers in individuals aged 50 to 59 years. |
| Other analysis: Overdiagnosis invasive cancers only (50-59 years)<br><br>Range of follow-up (yrs): 10 to 15 | 23.5 per 1,000 | **1.18 more per 1,000** (from 0.71 fewer to 3.06 more) | **RR 1.05** (0.97 to 1.13) | 132,231 (2 [1,2] RCTs) [a] | ⊕⊕◯◯ LOW [a,c,d,e,f] | Using a threshold of 5, screening may lead to little to no difference in overdiagnosed invasive cancers in individuals aged 50 to 59 years. |

**Mammography +/- CBE compared to Usual Care**

| Outcomes | Absolute effects | | Relative effect (95% CI) | № of participants (studies) | Quality of the evidence (GRADE) | Comments |
|---|---|---|---|---|---|---|
| | **Incident rates with usual care (Assumed rate) ‡** | **Absolute risk (95% CI)** | | | | |

‡The assumed rate was calculated using the control event rates across included studies.
**CI:** Confidence interval

**Bibliography:** 1: Malmo I (Zackrisson 2006[41]); 2: CNBSS 1 & 2 (Baines 2016[42]); 3: AGE (Duffy 2020[43])

**GRADE Working Group grades of evidence**
**High quality:** We are very confident that the true effect lies close to that of the estimate of the effect
**Moderate quality:** We are moderately confident in the effect estimate: The true effect is likely to be close to the estimate of the effect, but there is a possibility that it is substantially different
**Low quality:** Our confidence in the effect estimate is limited: The true effect may be substantially different from the estimate of the effect
**Very low quality:** We have very little confidence in the effect estimate: The true effect is likely to be substantially different from the estimate of effect

#### Explanations

a. We downrated once for risk of bias. Randomisation and allocation concealment were either not reported or there were serious deficiencies in these areas.
b. Approximately half point estimates in our pooled estimate cross our threshold, we downrated once for inconsistency.
c. Downrated once for indirectness. Data are from trials initiated in the 1960s-1990s and some trial estimates included participants outside the previously defined age decades (e.g., in the 40-49 age decade, one study included some individuals in their 50s).
d. Not rated down for imprecision. Clinical decision threshold set at 5.
e. Not downrated for publication bias. Cannot assess publication bias (insufficient number of trials).
f. Not downrated for inconsistency; all point estimates in our pooled analysis lie to one side of our threshold.

#### Table 11 forest plots

Age 40-49 (invasive + in situ)

| Study or Subgroup | | Intervention Events | Intervention Total | Control Events | Control Total | Weight | Risk Ratio M-H, Random, 95% CI |
|---|---|---|---|---|---|---|---|
| AGE | Age:[39-41] | 953 | 53914 | 1731 | 107007 | 46.4% | 1.09 [1.01, 1.18] |
| CNBSS 1 & 2 | Age:[40-59] | 958 | 44981 | 828 | 44967 | 33.6% | 1.16 [1.05, 1.27] |
| Malmo I | Age:[45-69] | 540 | 21088 | 507 | 21195 | 19.9% | 1.07 [0.95, 1.21] |
| **Total (95% CI)** | | | **119983** | | **173169** | **100.0%** | **1.11 [1.05, 1.17]** |
| Total events | | 2451 | | 3066 | | | |

Heterogeneity: Tau² = 0.00; Chi² = 1.27, df = 2 (P = 0.53); I² = 0%
Test for overall effect: Z = 3.81 (P = 0.0001)

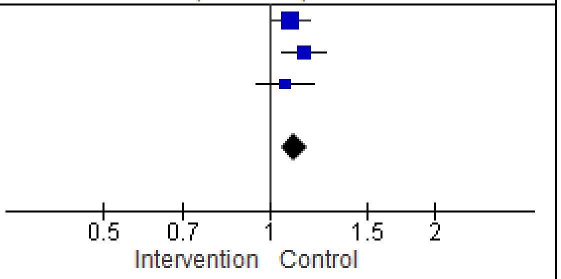

Age 40-49 (invasive only)

| Study or Subgroup | | Intervention Events | Intervention Total | Control Events | Control Total | Weight | Risk Ratio M-H, Random, 95% CI | Year |
|---|---|---|---|---|---|---|---|---|
| Malmo I | Age:[45-54] | 481 | 21088 | 454 | 21195 | 19.7% | 1.06 [0.94, 1.21] | 2006 |
| CNBSS I | Age:[40-49] | 912 | 44981 | 817 | 44967 | 35.5% | 1.12 [1.02, 1.23] | 2016 |
| AGE | Age:[39-41] | 835 | 53914 | 1628 | 107007 | 44.8% | 1.02 [0.94, 1.11] | 2020 |
| **Total (95% CI)** | | | **119983** | | **173169** | **100.0%** | **1.06 [1.00, 1.12]** | |
| Total events | | 2228 | | 2899 | | | | |

Heterogeneity: Tau² = 0.00; Chi² = 2.08, df = 2 (P = 0.35); I² = 4%
Test for overall effect: Z = 2.04 (P = 0.04)

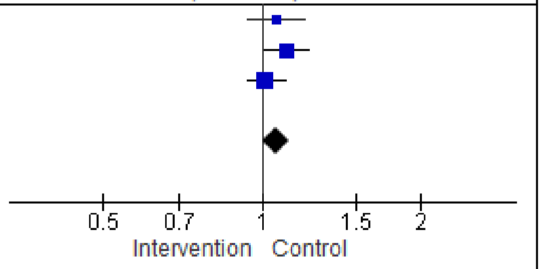

Age 50-59 (invasive + in situ)

| Study or Subgroup | | Intervention Events | Intervention Total | Control Events | Control Total | Weight | Risk Ratio M-H, Random, 95% CI |
|---|---|---|---|---|---|---|---|
| CNBSS 1 & 2 | Age:[50-59] | 942 | 44981 | 898 | 44967 | 55.2% | 1.05 [0.96, 1.15] |
| Malmo I | Age:[55-69] | 780 | 21088 | 698 | 21195 | 44.8% | 1.12 [1.02, 1.24] |
| **Total (95% CI)** | | | **66069** | | **66162** | **100.0%** | **1.08 [1.01, 1.16]** |
| Total events | | 1722 | | 1596 | | | |

Heterogeneity: Tau² = 0.00; Chi² = 0.99, df = 1 (P = 0.32); I² = 0%
Test for overall effect: Z = 2.28 (P = 0.02)

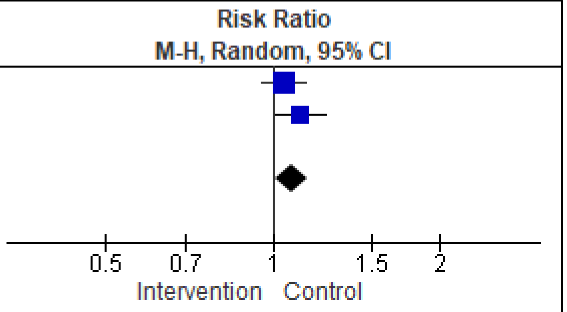

Age 50-59 (invasive only)

| Study or Subgroup | | Intervention Events | Intervention Total | Control Events | Control Total | Weight | Risk Ratio M-H, Random, 95% CI |
|---|---|---|---|---|---|---|---|
| CNBSS 1 & 2 | Age:[50-59] | 899 | 44981 | 891 | 44967 | 54.9% | 1.01 [0.92, 1.11] |
| Malmo I | Age:[55-69] | 719 | 21088 | 662 | 21195 | 45.1% | 1.09 [0.98, 1.21] |
| **Total (95% CI)** | | | **66069** | | **66162** | **100.0%** | **1.05 [0.97, 1.13]** |
| Total events | | 1618 | | 1553 | | | |

Heterogeneity: Tau² = 0.00; Chi² = 1.25, df = 1 (P = 0.26); I² = 20%
Test for overall effect: Z = 1.13 (P = 0.26)

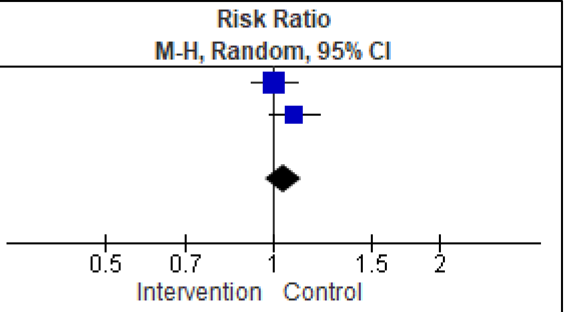

#### Table 12: Overdiagnosis (Observational studies, stratified by age)

##### Mammography +/- CBE compared to Usual Care

| Outcomes | **Summary‡** | № of participants (studies) | Quality of the evidence (GRADE) | Comments |
|---|---|---|---|---|
| Overdiagnosis invasive + in situ cancers (40-49 years)<br><br>Range of follow-up (yrs): 8 years<br>Screening interval: biennial | One study reported the number of invasive and in situ breast cancers among women 49 to 52 years and found a rate of 3.87 per 1,000 person years in the screened group and 2.45 per 1,000 person years in the unscreened group [RR 1.49 (95% CI 1.18 to 1.88)]. | Unclear<br>(1 study)[1] | ⊕◯◯◯<br>VERY LOW [a-e] | We are very uncertain whether screening leads to at least 5 overdiagnosed cancers in individuals aged 40 to 49 years. |
| Overdiagnosis invasive + in situ cancers (50-59 years)<br><br>Range of follow-up (yrs): 8 to 13 years<br>Screening interval: biennial | Two studies reported the number of invasive and in situ breast cancers. One study reported among 53- to 59-year-olds and found a rate of 2.77 per 1,000 person years in the screened group and 3.19 per 1,000 person years in the unscreened group. The second study found a rate of 3.74 per 1,000 individuals in the screening group and 3.40 per 1,000 individuals in the control group among 50- to 69-year-olds. | Unclear<br>(2 studies)[1,2] | ⊕◯◯◯<br>VERY LOW [a,d-g] | We are very uncertain whether screening leads to at least 5 overdiagnosed cancers in individuals aged 50 to 59 years. |
| Overdiagnosis invasive + in situ cancers (60-69 years)<br><br>Range of follow-up (yrs): 8 to 13 years<br>Screening interval: biennial | Two studies reported the number of invasive and in situ breast cancers. One study reported among 60- to 69-year-olds and found a rate of 3.59 per 1,000 person years in the screened group and 3.44 per 1,000 person years in the unscreened group. The second study found a rate of 3.74 per 1,000 individuals in the screening group and 3.40 per 1,000 individuals in the control group among 50- to 69-year-olds. | Unclear<br>(2 studies)[1,2] | ⊕◯◯◯<br>VERY LOW [a,d-g] | We are very uncertain whether screening leads to at least 5 overdiagnosed cancers in individuals aged 60 to 69 years. |
| Overdiagnosis invasive + in situ cancers (70-74 years)<br><br>Range of follow-up (yrs): 15 years<br>Screening interval: 3 years | One study reported the number of invasive and in situ breast cancers among women 70 to 74 years and found a rate of 61 per 1,000 individuals in the screened group and 41 per 1,000 individuals in the unscreened group [HR 1.47 (95% CI 1.19 to 1.81)]. | 19,925<br>(1 study)[3] | ⊕⊕◯◯<br>LOW [b,d,e,h,i] | Screening may lead to at least 5 overdiagnosed cancers in 1,000 individuals aged 70 to 74 years. |
| Overdiagnosis invasive + in situ cancers (75 years and older)<br><br>Range of follow-up (yrs): 15 years<br>Screening interval: 3 years | One study reported the number of invasive and in situ breast cancers among women 75 to 84 years and found a rate of 49 per 1,000 individuals in the screened group and 26 per 1,000 individuals in the unscreened group (HR 1.92 (95% CI 1.60 to 2.30)]. The same study reported for those aged 85 years or older and found a rate of 28 per 1,000 individuals in the screened group and 13 per 1,000 individuals in the unscreened group [HR 2.20 (95% CI 1.43 to 3.40)]. | 34,710<br>(1 study)[3] | ⊕⊕◯◯<br>LOW [b,d,e,h,i] | Screening may lead to at least 5 overdiagnosed cancers in 1,000 individuals aged 75 years or older. |

#### Mammography +/- CBE compared to Usual Care

| Outcomes | Summary‡ | № of participants (studies) | Quality of the evidence (GRADE) | Comments |
|---|---|---|---|---|

‡ We did not pool due to variations in reporting (e.g., different denominators such as person years). Results are narratively summarized.
**CI:** Confidence interval, **RR:** relative risk, **HR:** hazard ratio

**Bibliography:** 1 Lund 2018[44]; 2: Puliti 2017[37]; 3: Richman 2023[45]

**GRADE Working Group grades of evidence**
**High quality:** We are very confident that the true effect lies close to that of the estimate of the effect
**Moderate quality:** We are moderately confident in the effect estimate: The true effect is likely to be close to the estimate of the effect, but there is a possibility that it is substantially different
**Low quality:** Our confidence in the effect estimate is limited: The true effect may be substantially different from the estimate of the effect
**Very low quality:** We have very little confidence in the effect estimate: The true effect is likely to be substantially different from the estimate of effect

#### Explanations

a. Downrated once for risk of bias. One study had a follow up time deemed not sufficient to be long enough for outcomes to occur (at least 10-15 years) and there are concerns with adjusting for lead time or self selection bias.
b. Not downrated for inconsistency. One study assessed.
c. Downrated once for indirectness. We reported what study authors report and some study subjects in their 50s were included in the estimate, it's not truly a reflection of the desired age group.
d. Not downrated for imprecision. Narrative analysis. Clinical decision threshold of 5 more or fewer. Unable to assess OIS.
e. According to Egger et al.[11], 10 studies are needed to asses publication bias. We cannot assess publication bias due to insufficient number of trials, therefore, we did not rate down for publication bias.
f. Downrated once for inconsistency. Reporting of estimates varied between studies. One cannot be confident that the same methodological approach was used.
g. Downrated once for indirectness. We reported what study authors report and some study subjects in their 50s or 60s were missing in the estimate, it's not truly a reflection of the desired age group.
h. Not downrated for risk of bias.
i. Not downrated for indirectness.

#### Table 13: Interval cancers (Intervention arm only - descriptive data of RCTs)

| Screening with mammography (with or without CBE) compared to usual care | | | | |
|---|---|---|---|---|
| Outcomes | Impact | № of participants (studies) | Quality of the evidence (GRADE) | Comments |
| Invasive and DCIS (All ages) – screening interval **<=12 months**<br><br># R: 44,925<br># A: Unclear<br>Range of follow-up (yrs): 5.0 | 3.9 (95% CI 3.4 to 4.5) interval cancers (Invasive and DCIS) were detected in the mammography arm per 1000 women (176/44,925 randomized) over the follow-up period of five years (screening interval 12 months). | Unclear (1 RCT)[1] | ⊕⊕◯◯ LOW [a,b,c,d,j] | Using a threshold of 6 interval cancers over 10 years, screening may lead to little to no difference in interval cancers (invasive and DCIS).<br><br>We cannot comment on the comparative effectiveness of breast cancer screening for interval cancers, as interval cancers detected by screening cannot be measured in a non-screening comparator group. Interpretation of this estimate should be informed by additional data that is reflective of the current Canadian context. |
| Invasive and DCIS (All ages) – screening interval **13-24 months**<br><br># R: 62,222<br># A: Unclear<br>Range of follow-up (yrs): 4.8-7.0 | 3.1 (95% CI 2.6 to 3.7) interval cancers (Invasive and DCIS) were detected in the mammography arm per 1000 women over the follow-up period of 4.8-7 years (screening interval 18 months). | Unclear (2 RCTs)[2,3] | ⊕◯◯◯ VERY LOW [e,f,c,d,j] | Using a threshold of 6 interval cancers over 10 years, we are very uncertain if screening leads to little to no difference in interval cancers (invasive and DCIS).<br><br>We cannot comment on the comparative effectiveness of breast cancer screening for interval cancers, as interval cancers detected by screening cannot be measured in a non-screening comparator group. Interpretation of this estimate should be informed by additional data that is reflective of the current Canadian context. |

#### Screening with mammography (with or without CBE) compared to usual care

| Outcomes | Impact | № of participants (studies) | Quality of the evidence (GRADE) | Comments |
|---|---|---|---|---|
| Invasive and DCIS (All ages) – screening interval **>24 months**<br><br># R: 77,080<br># A: Unclear<br>Range of follow-up (yrs): 7.0 | 3.9 (95% CI 3.4 to 4.3) interval cancers (Invasive and DCIS) were detected in the mammography arm per 1000 women (298/77,080 randomised) over the follow-up period of 7 years (screening interval 23-33 months). | Unclear (1 RCT)[4] | ⊕⊕◯◯<br>LOW [a,b,c,d,j] | Using a threshold of 6 interval cancers over 10 years, screening may lead to little to no difference in interval cancers (invasive and DCIS).<br><br>We cannot comment on the comparative effectiveness of breast cancer screening for interval cancers, as interval cancers detected by screening cannot be measured in a non-screening comparator group. Interpretation of this estimate should be informed by additional data that is reflective of the current Canadian context. |
| Invasive Only (All ages) – 18-month screening interval<br><br># R: 61,968<br># A: Unclear<br>Mean follow-up (yrs): 4.8-7.0 | 2.8 (95% CI 2.4 to 3.3) interval cancers (Invasive cancers) were detected in the mammography arm per 1000 women over the follow-up period of 4.8-7 years (screening interval 18 months). | Unclear (2 RCTs)[2,5] | ⊕◯◯◯<br>VERY LOW [e,g,c,d,j] | Using a threshold of 6 interval cancers over 10 years, we are very uncertain if screening leads to little to no difference in interval cancers (invasive only).<br><br>We cannot comment on the comparative effectiveness of breast cancer screening for interval cancers, as interval cancers detected by screening cannot be measured in a non-screening comparator group. Interpretation of this estimate should be informed by additional data that is reflective of the current Canadian context. |

| Screening with mammography (with or without CBE) compared to usual care | | | | |
|---|---|---|---|---|
| Outcomes | Impact | № of participants (studies) | Quality of the evidence (GRADE) | Comments |
| DCIS Only (All ages) – 18-month screening interval<br><br># R: 61,968<br># A: Unclear<br>Mean follow-up (yrs): 4.8-7.0 | 0.2 (95% CI 0.1 to 0.5) interval cancers (DCIS cancers) were detected in the mammography arm per 1000 women over the follow-up period of 4.8-7 years (screening interval 18 months). | Unclear (2 RCTs)[2,5] | ⊕◯◯◯<br>VERY LOW [e,h,c,d,j] | Using a threshold of 6 interval cancers over 10 years, we are very uncertain if screening leads to little to no difference in interval cancers (DCIS).<br><br>We cannot comment on the comparative effectiveness of breast cancer screening for interval cancers, as interval cancers detected by screening cannot be measured in a non-screening comparator group. Interpretation of this estimate should be informed by additional data that is reflective of the current Canadian context. |
| Age group 39-49 years (Invasive and DCIS) – 18-month screening interval<br><br># R: 11,724<br># A: Unclear<br>Mean follow-up (yrs): 4.8-7.0 | 3.0 (95% CI 2.1 to 4.2) interval cancers (Invasive and DCIS) were detected in the mammography arm per 1000 women (35/11,724 randomised) over the follow-up period of 4.8-7 years (screening interval 18 months). | Unclear (1 RCT)[5] | ⊕⊕◯◯<br>LOW [i,b,c,d,j] | Using a threshold of 6 interval cancers over 10 years, screening may lead to little to no difference in interval cancers (invasive and DCIS).<br><br>We cannot comment on the comparative effectiveness of breast cancer screening for interval cancers, as interval cancers detected by screening cannot be measured in a non-screening comparator group. Interpretation of this estimate should be informed by additional data that is reflective of the current Canadian context. |

| Screening with mammography (with or without CBE) compared to usual care | | | | |
|---|---|---|---|---|
| Outcomes | Impact | № of participants (studies) | Quality of the evidence (GRADE) | Comments |
| Age group 39-49 years (Invasive Only) – 18-month screening interval<br><br># R: 11,724<br># A: Unclear<br>Mean follow-up (yrs): 4.8-7.0 | 2.8 (95% CI 1.9 to 3.9) interval cancers (Invasive) were detected in the mammography arm per 1000 women (33/11,724 randomised) over the follow-up period of 4.8-7 years (screening interval 18 months). | Unclear (1 RCT)[5] | ⊕⊕◯◯<br>LOW [i,b,c,d,j] | Using a threshold of 6 interval cancers over 10 years, screening may lead to little to no difference in interval cancers (invasive).<br><br>We cannot comment on the comparative effectiveness of breast cancer screening for interval cancers, as interval cancers detected by screening cannot be measured in a non-screening comparator group. Interpretation of this estimate should be informed by additional data that is reflective of the current Canadian context. |
| Age group 39-49 years (DCIS Only) – 18-month screening interval<br><br># R: 11,724<br># A: Unclear<br>Mean follow-up (yrs): 4.8-7.0 | 0.2 (95% CI 0.02 to 0.6) interval cancers (DCIS) were detected in the mammography arm per 1000 women (2/11,724 randomised) over the follow-up period of 4.8-7 years (screening interval 18 months). | Unclear (1 RCT)[5] | ⊕⊕◯◯<br>LOW [i,b,c,d,j] | Using a threshold of 6 interval cancers over 10 years, screening may lead to little to no difference in interval cancers (DCIS).<br><br>We cannot comment on the comparative effectiveness of breast cancer screening for interval cancers, as interval cancers detected by screening cannot be measured in a non-screening comparator group. Interpretation of this estimate should be informed by additional data that is reflective of the current Canadian context. |

#### Screening with mammography (with or without CBE) compared to usual care

| Outcomes | Impact | № of participants (studies) | Quality of the evidence (GRADE) | Comments |
|---|---|---|---|---|
| Age group 50-59 years (Invasive and DCIS) – 18-month screening interval<br><br># R: 9,926<br># A: Unclear<br>Mean follow-up (yrs): 4.8-7.0 | 1.9 (95% CI 1.2 to 3.0) interval cancers (Invasive; no DCIS detected) were detected in the mammography arm per 1000 women (19/9,926 randomised) over the follow-up period of 4.8-7 years (screening interval 18 months). | Unclear (1 RCT)[5] | ⊕⊕◯◯<br>LOW [i,b,c,d,j] | Using a threshold of 6 interval cancers over 10 years, screening may lead to little to no difference in interval cancers (invasive and DCIS).<br><br>We cannot comment on the comparative effectiveness of breast cancer screening for interval cancers, as interval cancers detected by screening cannot be measured in a non-screening comparator group. Interpretation of this estimate should be informed by additional data that is reflective of the current Canadian context. |

**Bibliography:**
1: CNBSS 1 & 2 (Miller 2014[9])
2: Stockholm (Frisell 1997[32])
3. Gothenburg (Bjurstam 2016[46])
4: Swedish Two County (Tabar 2011[7])
5: Gothenburg (Bjurstam 2003[12])

#### Explanations

a. Downrated once for risk of bias due to a lack of reporting for how interval cancers were detected and unclear reporting on who was used in the analysis.
b. Not downrated for inconsistency. Single study evaluated for outcome.
c. Downrated once for indirectness. Data are from trials initiated in the 1960s-1990s and the intervention groups were primarily screened with film mammography. Due to advances in mammography technology and treatment practices, we expect that the magnitude of screening effect may differ if applied to today's Canadian screening context. There are no high-quality clinical trials examining the impact of screening on interval cancers using contemporary screening methods.
d. The 95% CI does not cross the clinical decision threshold; therefore, we did not rate down for imprecision.
e. Downrated once for risk of bias. Studies ranged from moderate to high risk of bias. Lack of reporting for how interval cancers were detected and unclear reporting on who was used in the analysis.
f. Inconsistency is moderately high (I^2 = 61%). Rated down once.
g. Inconsistency is moderately high (I^2 = 52%). Rated down once.
h. Inconsistency is moderately high (I^2 = 57%). Rated down once.
i. Downrated once for risk of bias. Lack of reporting for how interval cancers were detected and missing important demographic details in intervention group.
j. According to Egger et al.[11], 10 trials are needed to asses publication bias. We cannot assess publication bias due to insufficient number of trials, therefore, we did not rate down for publication bias.

#### Table 13 forest plots

Forest plot for Invasive and DCIS (All ages):

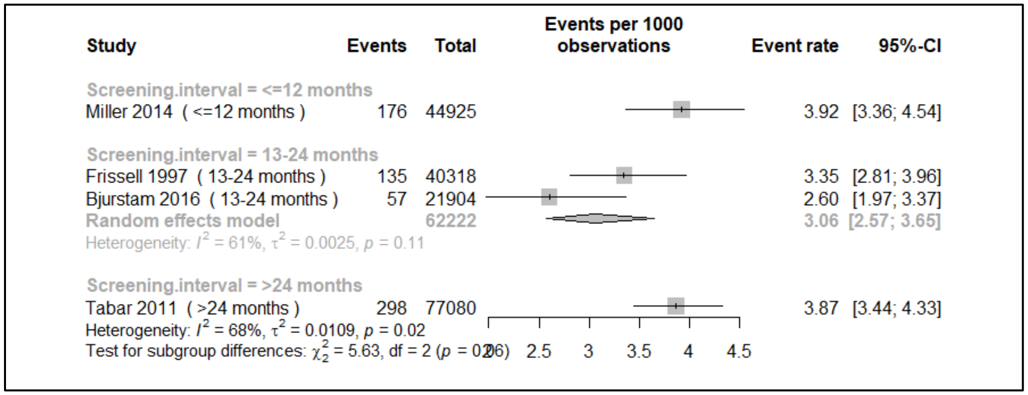

- Notes: Rates presented per 1000 women
- Follow-up rates varied between studies: Miller 5.0 years, Frissell and Bjurstam 4.8-7.0 years, Tabar 7.0 years

Forest plot for Invasive-only interval cancers (All ages):

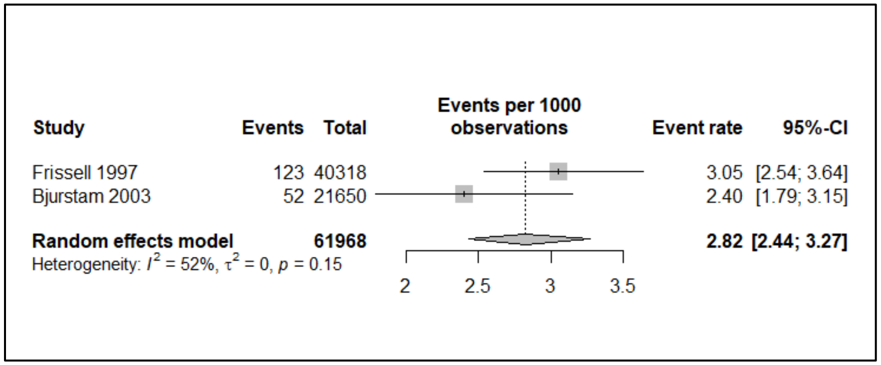

- Notes: Rates presented per 1000 women
- Follow-up rates varied between studies: 4.8-7.0 years

#### Forest plot for DCIS-only interval cancer (All ages):

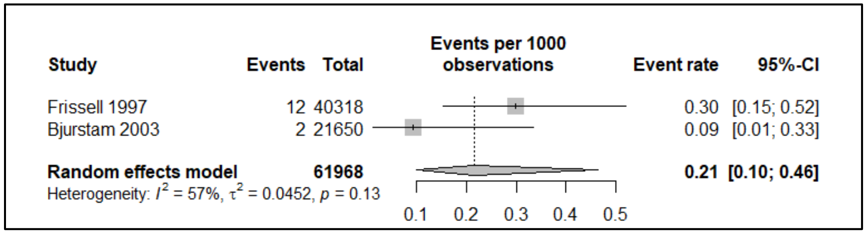

- Notes: Rates presented per 1000 women
- Follow-up rates varied between studies: 4.8-7.0 years

#### Interval Cancers – Age Subgroups

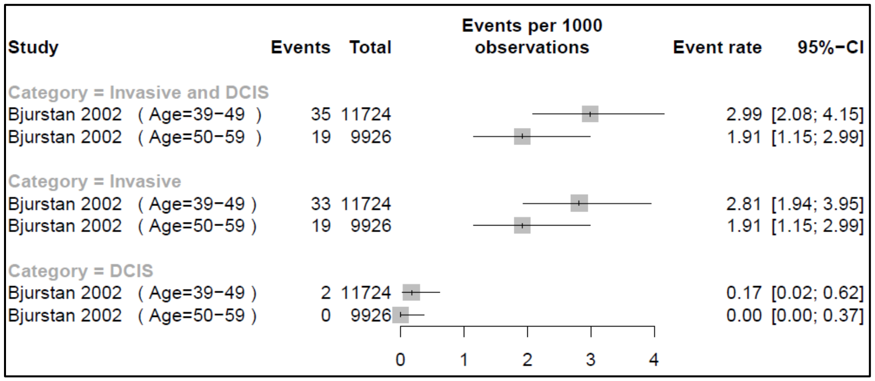

- Notes: Rates presented per 1000 women
- Follow-up rate ranged between 4.8-7.0 years

#### Table 14: Treatment-related morbidity (RCTs, all ages)

##### Screening with mammography (with or without CBE) compared to usual care

| Outcomes | Absolute effects | | Relative effect (95% CI) | № of participants (studies) | Quality of the evidence (GRADE) | Comments |
|---|---|---|---|---|---|---|
| | Risk with Usual Care (Assumed Risk)** | Absolute effect (95% CI) | | | | |
| Number of mastectomies<br><br>Mean follow up: 7-9 years | 9.2 per 1000 | **1.84 more per 1000** (from 1.01 more to 2.76 more) | **RR 1.20 (1.11 to 1.30)** | 250479 (5 RCTs[1-5]) | ⊕◯◯◯ VERY LOW [a,b,c,d,i] | Using a threshold of 2 fewer breast cancers requiring a full mastectomy per 1,000, we are very uncertain whether screening makes little to no difference in the number of mastectomies over 7-9 years in those in a general population. |
| Number treated with radiotherapy<br><br>Mean follow up: 7-9 years | 8.9 per 1000 | **2.85 more per 1000** (from 1.42 more to 4.45 more) | **RR 1.32 (1.16 to 1.50)** | 100383 (2 RCTs[3,4]) | ⊕⊕◯◯ LOW [b,e,f,g,i] | Using a threshold of 5 fewer breast cancers requiring radiotherapy per 1,000, screening may make little to no difference in the number treated with radiotherapy over 7-9 years in those in a general population. |
| Number treated with chemotherapy<br><br>Mean follow up: 7-9 years | 3.6 per 1000 | **0.14 fewer per 1000** (from 0.79 fewer to 0.68 more) | **RR 0.96 (0.78 to 1.19)** | 100383 (2 RCTs[3,4]) | ⊕⊕◯◯ LOW [b,e,f,h,i] | Using a threshold of 2 fewer breast cancers requiring chemotherapy per 1,000, screening may make little to no difference in the number treated with chemotherapy over 7-9 years in those at a general population risk of breast cancer. |

***The corresponding risk** (and its 95% confidence interval) is based on **the assumed risk** in the comparison group and the **relative effect** of the intervention (and its 95% CI).
**The overall event rate in the usual care group across included trials was used for the baseline risk.
**CI:** Confidence interval; **RR:** Risk ratio

**Bibliography:**
1: CNBSS 1 (Gøtzsche 2013[47]) 2: CNBSS 2 (Gøtzsche 2013[47]) 3: Malmo I (Gøtzsche 2013[47]) 4: Swedish Two County (Kopparberg) (Gøtzsche 2013[47]) 5: Stockholm (Gøtzsche 2013[47])

**GRADE Working Group grades of evidence**
**High quality:** We are very confident that the true effect lies close to that of the estimate of the effect
**Moderate quality:** We are moderately confident in the effect estimate: The true effect is likely to be close to the estimate of the effect, but there is a possibility that it is substantially different
**Low quality:** Our confidence in the effect estimate is limited: The true effect may be substantially different from the estimate of the effect
**Very low quality:** We have very little confidence in the effect estimate: The true effect is likely to be substantially different from the estimate of effect

#### Explanations

a. Downrated once for risk of bias. Randomisation and allocation concealment were either not reported (Malmo I, Swedish two county [Kopparberg]) or there were serious deficiencies in these areas (CNBSS 1&2, Stockholm).
b. All point estimates in our pooled analysis lie to one side of our threshold. We did not rate down for inconsistency.
c. Downrated once for indirectness. Data are from trials initiated in the 1960s-1990s and the intervention groups were primarily screened with film mammography. Due to advances in mammography technology and treatment practices, we expect that the magnitude of screening effect may differ if applied to today's Canadian screening context. There are no high-quality clinical trials examining the impact of screening on breast cancer treatment-related morbidity using contemporary screening methods. For some studies, the control group received screening after the screening period (Stockholm and Swedish Two County [Kopparberg]).
d. Downrated once for imprecision. CI crosses threshold for breast cancer screening harm for breast cancers requiring a full mastectomy (versus lumpectomy).
e. Downrated once for risk of bias. Randomisation and allocation concealment were not reported (Malmo I, Swedish two county [Kopparberg]).
f. Downrated once for indirectness. Trial data is from trials mainly initiated in the 1970s-1980s and the intervention groups were primarily screened with film mammography. Due to advances in mammography technology and treatment practices, we expect that the magnitude of screening effect would differ if applied to today's Canadian screening context. In one of the studies, the control group received screening after the screening period (Swedish Two County [Kopparberg]).
g. Not downrated for imprecision. The CI does not cross the threshold for breast cancer screening harm for breast cancers requiring radiotherapy.
h. Not downrated for imprecision. The CI does not cross the threshold for breast cancer screening benefit or harm for breast cancers requiring chemotherapy.
i. According to Egger et al.[11], 10 trials are needed to asses publication bias. We cannot assess publication bias due to insufficient number of trials, therefore, we did not rate down for publication bias.

#### Table 15: Treatment-related morbidity (Observational studies, all ages, adherence to screen analysis)

**Screening with mammography* compared to no screening**

| Outcomes | Absolute effects | | Relative effect (95% CI) | № of participants (studies) | Quality of the evidence (GRADE) | Comments |
|---|---|---|---|---|---|---|
| | **Risk with Usual Care (Assumed Risk)** | **Absolute effect (95% CI)** | | | | |
| Breast cancers with conservative surgery as treatment<br><br>Range of follow-up (yrs): Median 11, IQR 9-13 | 1.83/1000 | **0.9 more per 1,000** (from 0.7 more to 1.1 more) | **Rate ratio 1.5 (1.4 to 1.6)** | 413,447 (1 study[1]) | ⨁◯◯◯ VERY LOW [a,b,c,d,e] | Using a threshold of 2 fewer breast cancers requiring a full mastectomy per 1,000, screening may make little to no difference in the number of breast cancers with conservative surgery as treatment in individuals in a general population. |
| Breast cancers with mastectomy as treatment<br><br>Range of follow-up (yrs): Median 11, IQR 9-13 | 1.24/1000 | **0.4 fewer per 1,000** (from 0.35 fewer to 0.46 fewer) | **Rate ratio 0.68 (0.63 to 0.72)** | 413,447 (1 study[1]) | ⨁◯◯◯ VERY LOW [a,b,c,d,e] | Using a threshold of 2 fewer breast cancers requiring a full mastectomy per 1,000, screening may make little to no difference in the number of breast cancers with mastectomy as treatment in individuals in a general population. |

***The corresponding risk** (and its 95% confidence interval) is based on **the assumed risk** in the comparison group and the **relative effect** of the intervention (and its 95% CI).
**CI:** Confidence interval; **RR:** Risk ratio

**Bibliography:**
1: Puliti 2017[37]

**GRADE Working Group grades of evidence**
**High quality:** We are very confident that the true effect lies close to that of the estimate of the effect
**Moderate quality:** We are moderately confident in the effect estimate: The true effect is likely to be close to the estimate of the effect, but there is a possibility that it is substantially different
**Low quality:** Our confidence in the effect estimate is limited: The true effect may be substantially different from the estimate of the effect
**Very low quality:** We have very little confidence in the effect estimate: The true effect is likely to be substantially different from the estimate of effect

#### Explanations

a. Study downrated once for moderate risk of bias (RoB score for JBI cohort tool=7/11). Lack of adjustment for important confounding factors (use of HRT, breast density). No description of number of women lost to follow-up.
b. Not downrated for inconsistency. Single study evaluated outcome (unable to evaluate heterogeneity).
c. Not downrated for indirectness. Studies used population-based approach which was reflective of general population.
d. Not downrated for imprecision. Large population and CI does not cross threshold for breast cancer screening benefit for breast cancers requiring a full mastectomy (2 less breast cancers requiring mastectomy).
e. According to Egger et al.[11], 10 studies are needed to asses publication bias. We cannot assess publication bias due to insufficient number of trials, therefore, we did not rate down for publication bias.

#### Table 16: Treatment-related morbidity (Observational studies, by age subgroup, stop screening analysis)

**"Continue Screening" After Baseline Examination compared to "Stop Screening" After Baseline Examination**

| Outcomes | 70-74 Age Subgroup | 75-84 Age Subgroup | № of participants (studies) | Quality of the evidence (GRADE) | Comments |
|---|---|---|---|---|---|
| Simple mastectomy<br><br># of follow-up (yrs): 8.0 | The proportion of women (aged 70-74) diagnosed with breast cancer who received simple mastectomy in the continue screening strategy was 11.3 (10.8–11.8) and 10.4 (9.5–11.3) in the stop screening strategy group (absolute difference 9 more per 1,000). | The proportion of women (aged 75-84) diagnosed with breast cancer who received simple mastectomy in the continue screening strategy was 10.8 (10.3–11.2) and 10.1 (9.4–10.9) in the stop screening strategy group (absolute difference 7 more per 1,000) | 2,639,194 (1 study)[1] | ⊕⊕◯◯ LOW [a,b,c,d,e] | We found low certainty evidence comparing the proportions of women requiring simple mastectomy among those who continued mammography screening and those who stopped screening in their 70s (70-74 and 75-84 age groups). |
| Radical mastectomy<br><br># of follow-up (yrs): 8.0 | The proportion of women (aged 70-74) diagnosed with breast cancer who received radical mastectomy in the continue screening strategy was 13.9 (13.4–14.5) and 18.2 (17.0–19.4) in the stop screening strategy group (absolute difference 43 fewer per 1,000) | The proportion of women (aged 75-84) diagnosed with breast cancer who received radical mastectomy in the continue screening strategy was 14.2 (13.7–14.6) and 17.0 (16.0–17.9) in the stop screening strategy group (absolute difference 28 fewer per 1,000). | 2,639,194 (1 study)[1] | ⊕⊕◯◯ LOW [a,b,c,d,e] | We found low certainty evidence comparing the proportions of women requiring radical mastectomy among those who continued mammography screening and those who stopped screening in their 70s (70-74 and 75-84 age groups). |
| Radiotherapy<br><br># of follow-up (yrs): 8.0 | The proportion of women (aged 70-74) diagnosed with breast cancer who received radiotherapy in the continue screening strategy was 51.0 (50.3–51.8) and 39.9 (38.6–41.3) in the stop screening strategy group (absolute difference 111 more per 1,000) | The proportion of women (aged 75-84) diagnosed with breast cancer who received radiotherapy in the continue screening strategy was 41.2 (40.4–41.9) and 31.9 (30.7–33.1) in the stop screening strategy group (absolute difference 93 more per 1,000). | 2,639,194 (1 study)[1] | ⊕⊕◯◯ LOW [a,b,c,d,e] | We found low certainty evidence comparing the proportions of women requiring radiotherapy among those who continued mammography screening and those who stopped screening in their 70s (70-74 and 75-84 age groups). |
| Chemotherapy<br><br># of follow-up (yrs): 8.0 | The proportion of women (aged 70-74) diagnosed with breast cancer who received chemotherapy in the continue screening strategy was 15.2 (14.7–15.8) and 21.1 (20.0–22.1) in the stop screening strategy group (absolute difference 59 fewer per 1,000) | The proportion of women (aged 75-84) diagnosed with breast cancer who received chemotherapy in the continue screening strategy was 8.6 (8.3–9.1) and 11.5 (10.6–12.3) in the stop screening strategy group (absolute difference 29 fewer per 1,000). | 2,639,194 (1 study)[1] | ⊕⊕◯◯ LOW [a,b,c,d,e] | We found low certainty evidence comparing the proportions of women requiring chemotherapy among those who continued mammography screening and those who stopped screening in their 70s (70-74 and 75-84 age groups). |

#### “Continue Screening” After Baseline Examination compared to “Stop Screening” After Baseline Examination

| Outcomes | 70-74 Age Subgroup | 75-84 Age Subgroup | № of participants (studies) | Quality of the evidence (GRADE) | Comments |
|---|---|---|---|---|---|

**Bibliography:**
1: Garcia-Albeniz 2020[19]

a. We did not downrate for risk of bias. Study was judged to be of moderate quality using the JBI critical appraisal tool for cohort studies.
b. We did not downrate for inconsistency (only one study included).
c. We did not downrate for indirectness. The study answers the question of stopping versus continuing screening and all patients have received at least one baseline mammography.
d. Unable to evaluate imprecision using thresholds, as the baseline rate of treatment is not provided in the “stop screening” group to allow the calculation of absolute effects. Therefore, a minimally contextualized approach was used. The total population is large (>2000) and there is a large event rate (>300). We did not downrate for imprecision.
e. According to Egger et al.[11], 10 studies are needed to asses publication bias. We cannot assess publication bias due to insufficient number of trials, therefore, we did not rate down for publication bias.

### Table 16 forest plots*

**from Gøtzsche, P. C., & Jørgensen, K. J. (2013). Screening for breast cancer with mammography. Cochrane database of systematic reviews, (6).*

**Analysis 1.15:** Comparison 1 Screening with mammography versus no screening, Outcome 15 Number of mastectomies.

| Study or subgroup | Screening n/N | No screening n/N | Risk Ratio M-H, Fixed, 95% CI | Weight | Risk Ratio M-H, Fixed, 95% CI |
|---|---|---|---|---|---|
| **1.15.1 Adequately randomised trials** | | | | | |
| Canada 1980a | 183/25214 | 157/25216 | | 14.65% | 1.17[0.94,1.44] |
| Canada 1980b | 197/19711 | 176/19694 | | 16.43% | 1.12[0.91,1.37] |
| Malmö 1976 | 424/21242 | 339/21244 | | 31.63% | 1.25[1.09,1.44] |
| **Subtotal (95% CI)** | **66167** | **66154** | | **62.71%** | **1.2[1.08,1.32]** |
| Total events: 804 (Screening), 672 (No screening) | | | | | |
| Heterogeneity: Tau²=0; Chi²=0.86, df=2(P=0.65); I²=0% | | | | | |
| Test for overall effect: Z=3.45(P=0) | | | | | |
| **1.15.2 Suboptimally randomised trials** | | | | | |
| Kopparberg 1977 | 475/39051 | 196/18846 | | 24.67% | 1.17[0.99,1.38] |
| Stockholm 1981 | 263/40318 | 101/19943 | | 12.61% | 1.29[1.02,1.62] |
| **Subtotal (95% CI)** | **79369** | **38789** | | **37.29%** | **1.21[1.06,1.38]** |
| Total events: 738 (Screening), 297 (No screening) | | | | | |
| Heterogeneity: Tau²=0; Chi²=0.45, df=1(P=0.5); I²=0% | | | | | |
| Test for overall effect: Z=2.78(P=0.01) | | | | | |
| **Total (95% CI)** | **145536** | **104943** | | **100%** | **1.2[1.11,1.3]** |
| Total events: 1542 (Screening), 969 (No screening) | | | | | |
| Heterogeneity: Tau²=0; Chi²=1.33, df=4(P=0.86); I²=0% | | | | | |
| Test for overall effect: Z=4.43(P<0.0001) | | | | | |
| Test for subgroup differences: Chi²=0.02, df=1 (P=0.9), I²=0% | | | | | |

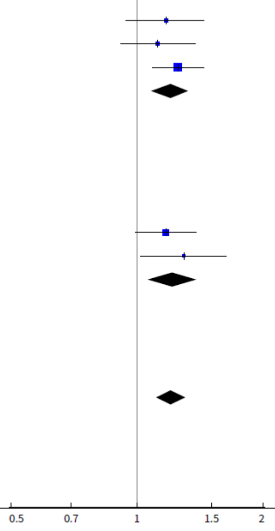

**Analysis 1.16:** Comparison 1 Screening with mammography versus no screening, Outcome 16 Number treated with radiotherapy.

| Study or subgroup | Screening n/N | No screening n/N | Risk Ratio M-H, Fixed, 95% CI | Weight | Risk Ratio M-H, Fixed, 95% CI |
|---|---|---|---|---|---|
| **1.16.1 Adequately randomised trials** | | | | | |
| Malmö 1976 | 260/21242 | 209/21244 | | 50.97% | 1.24[1.04,1.49] |
| **Subtotal (95% CI)** | **21242** | **21244** | | **50.97%** | **1.24[1.04,1.49]** |
| Total events: 260 (Screening), 209 (No screening) | | | | | |
| Heterogeneity: Not applicable | | | | | |
| Test for overall effect: Z=2.36(P=0.02) | | | | | |
| **1.16.2 Suboptimally randomised trials** | | | | | |
| Kopparberg 1977 | 433/39051 | 149/18846 | | 49.03% | 1.4[1.17,1.69] |
| **Subtotal (95% CI)** | **39051** | **18846** | | **49.03%** | **1.4[1.17,1.69]** |
| Total events: 433 (Screening), 149 (No screening) | | | | | |
| Heterogeneity: Not applicable | | | | | |
| Test for overall effect: Z=3.58(P=0) | | | | | |
| **Total (95% CI)** | **60293** | **40090** | | **100%** | **1.32[1.16,1.5]** |
| Total events: 693 (Screening), 358 (No screening) | | | | | |
| Heterogeneity: $Tau^2=0$; $Chi^2=0.82$, df=1(P=0.36); $I^2=0\%$ | | | | | |
| Test for overall effect: Z=4.22(P<0.0001) | | | | | |

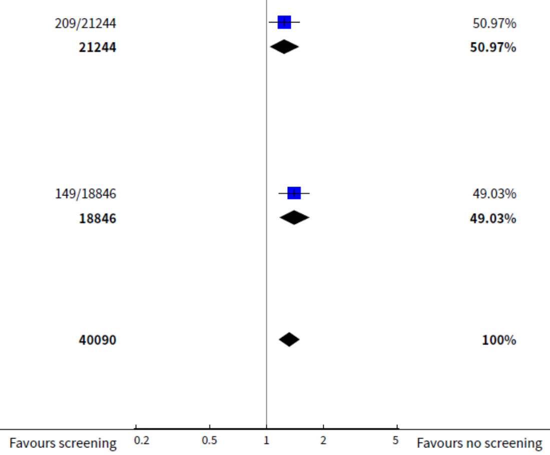

**Analysis 1.17:** Comparison 1 Screening with mammography versus no screening, Outcome 17 Number treated with chemotherapy.

| Study or subgroup | Screening n/N | No screening n/N | Risk Ratio M-H, Fixed, 95% CI | Weight | Risk Ratio M-H, Fixed, 95% CI |
|---|---|---|---|---|---|
| **1.17.1 Adequately randomised trials** | | | | | |
| Malmö 1976 | 26/21242 | 41/21244 | | 22.78% | 0.63[0.39,1.04] |
| **Subtotal (95% CI)** | **21242** | **21244** | | **22.78%** | **0.63[0.39,1.04]** |
| Total events: 26 (Screening), 41 (No screening) | | | | | |
| Heterogeneity: Not applicable | | | | | |
| Test for overall effect: Z=1.82(P=0.07) | | | | | |
| **1.17.2 Suboptimally randomised trials** | | | | | |
| Kopparberg 1977 | 226/39051 | 103/18846 | | 77.22% | 1.06[0.84,1.34] |
| **Subtotal (95% CI)** | **39051** | **18846** | | **77.22%** | **1.06[0.84,1.34]** |
| Total events: 226 (Screening), 103 (No screening) | | | | | |
| Heterogeneity: Not applicable | | | | | |
| Test for overall effect: Z=0.48(P=0.63) | | | | | |
| **Total (95% CI)** | **60293** | **40090** | | **100%** | **0.96[0.78,1.19]** |
| Total events: 252 (Screening), 144 (No screening) | | | | | |
| Heterogeneity: Tau²=0; Chi²=3.42, df=1(P=0.06); I²=70.77% | | | | | |
| Test for overall effect: Z=0.36(P=0.72) | | | | | |
| Test for subgroup differences: Chi²=3.42, df=1 (P=0.06), I²=70.77% | | | | | |

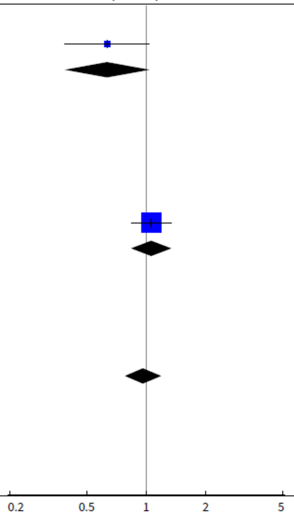

Favours screening — Favours no screening

#### Table 17: Additional imaging (no cancer)

**Table 17a:** Additional imaging with or without biopsy (no cancer) – by age subgroup

| Outcomes | Calculated Estimate (2011-2012 CPAC Data)** | Calculated Estimate (2019 British Columbia Data)** | Quality of the evidence (Based on GRADE*) | Comments |
|---|---|---|---|---|
| Additional imaging with or without biopsy (no cancer) over 10 years (40-49 years)† | **367.5 per 1000** | **477.6 per 1000** | ⊕⊕⊕◯ MODERATE [a,b,c,d,e] | Screening probably leads to at least 150 women requiring additional imaging with or without biopsy (no cancer) in 1000 women screened every 2-3 years over a 10-year period (40-49 years) |
| Additional imaging with or without biopsy (no cancer) over 10 years (50-59 years)† | **365.5 per 1000** | **410.5 per 1000** | ⊕⊕⊕◯ MODERATE [a,b,c,d,e] | Screening probably leads to at least 150 women requiring additional imaging with or without biopsy (no cancer) in 1000 women screened every 2-3 years over a 10-year period (50-59 years, started screening at age 50) |
| Additional imaging with or without biopsy (no cancer) over 10 years (50-59 years)‡ | **286.4 per 1000** | **285.2 per 1000** | ⊕⊕⊕◯ MODERATE [a,b,c,d,e] | Screening probably leads to at least 150 women requiring additional imaging with or without biopsy (no cancer) in 1000 women screened every 2-3 years over a 10-year period (50-59 years, started screening prior to age 50) |
| Additional imaging with or without biopsy (no cancer) over 10 years (60-69 years)‡ | **257.2 per 1000** | **252.4 per 1000** | ⊕⊕⊕◯ MODERATE [a,b,c,d,e] | Screening probably leads to at least 150 women requiring additional imaging with or without biopsy (no cancer) in 1000 women screened every 2-3 years over a 10-year period (60-69 years) |
| Additional imaging with or without biopsy (no cancer) over 10 years (70+ years)‡ | **220.4 per 1000** | **238.4 per 1000** | ⊕⊕⊕◯ MODERATE [a,b,c,d,e] | Screening probably leads to at least 150 women requiring additional imaging with or without biopsy (no cancer) in 1000 women screened every 2-3 years over a 10-year period (70+ years) |

*GRADE ratings are not typically applied to the context of primary evidence sets generated by analyses of quality indicator surveillance data. However, our judgements of the overall certainty of evidence have been informed by similar considerations used in the GRADE process for effectiveness data.
†Scenario 1: Assuming started biennial screening in current age decade (calculated using one initial screen and three subsequent screens over a 10-year period).
‡Scenario 2: Assuming started biennial screening in prior age decade (calculated using four subsequent screens over a 10-year period).
**Data Sources: Using data from 2011- 2012 from the 2016 CPAC report[48], we estimated the approximate rate of additional imaging with or without biopsy (no cancer) for women in each age decade over a 10 year-period (Table 7A). The BC estimates were estimated using breast screening program outcome indicators by 10-year age groups for 2019 for the "overall" risk groups (Table 9). See supplemental KQ1 GRADE Material, Appendix 6, part E for an example calculation.
Additional imaging estimates per screening cycle were calculating by subtracting cancer detection rates (invasive + DCIS) from abnormal call rates, stratified by age decade and if data were related to an "initial" or "subsequent" screen. We assumed women received four screens over a 10-year period, if the majority of women would receive a screen every 2-3 years (approximating biennial screening for the majority noting that some provinces currently offer[49] or recommend[50] annual screening in women aged 40-49 or starting at age 45 ). Scenario 1 assumes women start screening in that age decade and receive four screens over 10 years (one initial and three subsequent) (age groups: 40-49 and 50-59†). Scenario 2 assumed women started screening in prior age decades and therefore received four subsequent screens over a 10-year period (age groups: 50-59‡, 60-69, 70+).

a. The CPAC quality indicator data was used from the Canadian Breast Cancer Screening Database (CBCSD), which contained relatively complete data from participating provinces and territories for the quality indicators of interest in 2011-2012. The BC estimates were estimated using breast screening program outcome indicators by 10-year age groups for 2019, which should contain complete data. We did not downrate for risk of bias.
b. Estimates were calculated using quality indicators from screen-level data. Thus, we have no measure of imprecision in the data. All point estimates cross the minimum threshold for important effect (150 women with no cancer who require either imaging alone or imaging plus biopsy per 1000 screens).
c. We did not downrate for inconsistency. All age estimates for both the CPAC and the BC data fall above our threshold of 150 patients requiring additional imaging with or without biopsy (no cancer) per 1000 screens.
d. There appears to be an increase in recall rates over time (see Supplemental KQ1 GRADE Material, Appendix 6) depending on the data source. However, our conclusions about the rates of additional imaging with or without biopsy (no cancer) are unlikely to change, as the rates remain relatively consistent using more recent CPAC data and provincial data and above our clinical decision threshold. We did not downrate for indirectness.
e. We uprated our overall conclusion to moderate certainty of evidence as imaging recall estimates are similar across different data sources and consistently cross our threshold for clinical decision making.

**Table 17b:** – Additional imaging no biopsy (no cancer) – by age subgroup

| Outcomes | Calculated Estimate (2011-2012 CPAC Data)** | Quality of the evidence (Based on GRADE*) | Comments |
|---|---|---|---|
| Additional imaging no biopsy (no cancer) over 10 years (40-49 years)† | **312.8 per 1000** | ⊕⊕⊕◯ MODERATE [a,b,c,d] | Screening probably leads to at least 150 women requiring additional imaging no biopsy (no cancer) in 1000 women screened every 2-3 years over a 10-year period (40-49 years) |
| Additional imaging no biopsy (no cancer) over 10 years (50-59 years)† | **319.3 per 1000** | ⊕⊕⊕◯ MODERATE [a,b,c,d] | Screening probably leads to at least 150 women requiring additional imaging no biopsy (no cancer) in 1000 women screened every 2-3 years over a 10-year period (50-59 years, started screening at age 50) |
| Additional imaging no biopsy (no cancer) over 10 years (50-59 years)‡ | **252.4 per 1000** | ⊕⊕⊕◯ MODERATE [a,b,c,d] | Screening probably leads to at least 150 women requiring additional imaging no biopsy (no cancer) in 1000 women screened every 2-3 years over a 10-year period (50-59 years, started screening prior to age 50) |
| Additional imaging no biopsy (no cancer) over 10 years (60-69 years)‡ | **224.4 per 1000** | ⊕⊕⊕◯ MODERATE [a,b,c,d] | Screening probably leads to at least 150 women requiring additional imaging no biopsy (no cancer) in 1000 women screened every 2-3 years over a 10-year period (60-69 years) |
| Additional imaging no biopsy (no cancer) over 10 years (70+ years)‡ | **190 per 1000** | ⊕⊕⊕◯ MODERATE [a,b,c,d] | Screening probably leads to at least 150 women requiring additional imaging no biopsy (no cancer) in 1000 women screened every 2-3 years over a 10-year period (70+ years) |

*GRADE ratings are not typically applied to the context of primary evidence sets generated by analyses of quality indicator surveillance data. However, our judgements of the overall certainty of evidence have been informed by similar considerations used in the GRADE process for effectiveness data.
†Scenario 1: Assuming started biennial screening in current age decade (calculated using one initial screen and three subsequent screens over a 10-year period).
‡Scenario 2: Assuming started biennial screening in prior age decade (calculated using four subsequent screens over a 10-year period).

**Data Sources: Using data from 2011- 2012 from the 2016 CPAC report[48], we estimated the approximate rate of additional imaging with or without biopsy (no cancer) for women in each age decade over a 10 year-period (Table 7A).
Additional imaging estimates per screening cycle were calculating by subtracting cancer detection rates (invasive + DCIS) and the additional imaging and biopsy (no cancer) from the abnormal call rates, stratified by age decade and if data were related to an "initial" or "subsequent" screen. See supplemental KQ1 GRADE Material, Appendix 6, part E for an example calculation.
We assumed women received four screens over a 10-year period, if the majority of women would receive a screen every 2-3 years (approximating biennial screening for the majority, noting that some provinces currently offer[49] or recommend[50] annual screening in women aged 40-49 or starting at age 45. Scenario 1 assumes women start screening in that age decade and receive four screens over 10 years (one initial and three subsequent) (age groups: 40-49 and 50-59†). Scenario 2 assumed women started screening in prior age decades and therefore received four subsequent screens over a 10-year period (age groups: 50-59‡, 60-69, 70+).

a. The CPAC quality indicator data was used from the Canadian Breast Cancer Screening Database (CBCSD), which contained relatively complete data from participating provinces and territories for the quality indicators of interest in 2011-2012.
b. Estimates were calculated using quality indicators from screen-level data. Thus, we have no measure of imprecision in the data. All point estimates cross the minimum threshold for important effect (threshold used was 150 women who do not have cancer and require additional imaging and no biopsy per 1000 screens).
c. There appears to be an increase in recall rates over time (see Supplemental KQ1 GRADE Material, Appendix 6) depending on the data source. However, our conclusions about the rates of additional imaging no biopsy (no cancer) are unlikely to change, as the rates remain relatively consistent using more recent CPAC data and provincial data and above our clinical decision threshold. We did not downrate for indirectness.
d. We uprated our overall conclusion to moderate certainty of evidence as imaging recall estimates are similar across different data sources and consistently cross our threshold for clinical decision making.

**Table 17c:** – Additional imaging and biopsy (no cancer) – by age subgroup

| Outcomes | **Calculated Estimate (2011-2012 CPAC Data)** | Quality of the evidence (Based on GRADE*) | Comments |
|---|---|---|---|
| Additional imaging and biopsy (no cancer) over 10 years (40-49 years)† | **54.7 per 1000** | ⊕⊕⊕◯ MODERATE [a,b,c,d] | Screening probably leads to at least 15 women requiring additional imaging and biopsy (no cancer) in 1000 women screened every 2-3 years over a 10-year period (40-49 years) |
| Additional imaging and biopsy (no cancer) over 10 years (50-59 years)† | **46.2 per 1000** | ⊕⊕⊕◯ MODERATE [a,b,c,d] | Screening probably leads to at least 15 women requiring additional imaging and biopsy (no cancer) in 1000 women screened every 2-3 years over a 10-year period (50-59 years, started screening at age 50) |
| Additional imaging and biopsy (no cancer) over 10 years (50-59 years)‡ | **34.0 per 1000** | ⊕⊕⊕◯ MODERATE [a,b,c,d] | Screening probably leads to at least 15 women requiring additional imaging and biopsy (no cancer) in 1000 women screened every 2-3 years over a 10-year period (50-59 years, started screening prior to age 50) |
| Additional imaging and biopsy (no cancer) over 10 years (60-69 years)‡ | **32.8 per 1000** | ⊕⊕⊕◯ MODERATE [a,b,c,d] | Screening probably leads to at least 15 women requiring additional imaging and biopsy (no cancer) in 1000 women screened every 2-3 years over a 10-year period (60-69 years) |
| Additional imaging and biopsy (no cancer) over 10 years (70+ years)‡ | **30.4 per 1000** | ⊕⊕⊕◯ MODERATE [a,b,c,d] | Screening probably leads to at least 15 women requiring additional imaging and biopsy (no cancer) in 1000 women screened every 2-3 years over a 10-year period (70+ years) |

*GRADE ratings are not typically applied to the context of primary evidence sets generated by analyses of quality indicator surveillance data. However, our judgements of the overall certainty of evidence have been informed by similar considerations used in the GRADE process for effectiveness data.
†Scenario 1: Assuming started biennial screening in current age decade (calculated using one initial screen and three subsequent screens over a 10-year period).
‡Scenario 2: Assuming started biennial screening in prior age decade (calculated using four subsequent screens over a 10-year period).

**Data Sources: Using data from 2011- 2012 from the 2016 CPAC report[48], we estimated the approximate rate of additional imaging with or without biopsy (no cancer) for women in each age decade over a 10 year-period (Table 7A).
Additional imaging estimates per screening cycle were based on reported non-malignant biopsy rates, stratified by age decade and if data were related to an “initial” or “subsequent” screen. See supplemental KQ1 GRADE Material, Appendix 6, part E for an example calculation.
We assumed women received four screens over a 10-year period, if the majority of women would receive a screen every 2-3 years (approximating biennial screening for the majority, noting that some provinces currently offer[49] or recommend[50] annual screening in women aged 40-49 or starting at age 45. Scenario 1 assumes women start screening in that age decade and receive four screens over 10 years (one initial and three subsequent) (age groups: 40-49 and 50-59†). Scenario 2 assumed women started screening in prior age decades and therefore received four subsequent screens over a 10-year period (age groups: 50-59‡, 60-69, 70+)

a. The CPAC quality indicator data was used from the Canadian Breast Cancer Screening Database (CBCSD), which contained relatively complete data from participating provinces and territories for the quality indicators of interest in 2011-2012. The BC estimates were estimated using breast screening program outcome indicators by 10-year age groups for 2019, which should contain complete data. We did not downrate for risk of bias.
b. Estimates were calculated using quality indicators from screen-level data. Thus, we have no measure of imprecision in the data. All point estimates cross the minimum threshold for important effect (15 women requiring additional imaging and biopsy per 1000 screens).
c. The rates of additional imaging and biopsies (no cancer) appear to have remained consistent over time based on provincial data sources (Appendix 6, part B). We did not downrate for indirectness.
d. We uprated our overall conclusion to moderate certainty of evidence as imaging recall estimates are similar across different data sources and consistently cross our threshold for clinical decision making.

#### Appendix 9 – Cohort and RCT forest plot

| Reference | Study | Study Design | Mean Follow-up(yrs) | Screening period | log(RR) | SE | RR | 95%-CI |
|---|---|---|---|---|---|---|---|---|
| Morrel 2017 | New Zealand BSA | OBS | 12* | 2000-2011 | -0.97 | 0.13 | 0.38 | [0.29; 0.49] |
| Choi 2021 | KNCSP | OBS | 14* | 2002-2015 | -0.84 | 0.01 | 0.43 | [0.42; 0.44] |
| Duffy 2021 | National Swedish Study | OBS | 22 | 1992-2016 | -0.67 | 0.04 | 0.51 | [0.47; 0.55] |
| Coldman 2014 | Pan-Canadian Study | OBS | 19 | 1990-2009 | -0.51 | 0.06 | 0.60 | [0.53; 0.68] |
| Tabar 2011 | Swedish Two County | RCT | 29** | 1977-1984 | -0.31 | 0.10 | 0.73 | [0.60; 0.89] |
| Nystrom 2016 | Gothenburg | RCT | 24 | 1982-1989 | -0.30 | 0.14 | 0.74 | [0.56; 0.97] |
| Shapiro 1988 | HIP | RCT | 18 | 1963-1966 | -0.24 | 0.12 | 0.79 | [0.62; 1.00] |
| Nystrom 2016 | Malmo II | RCT | 22 | 1978-1990 | -0.16 | 0.22 | 0.85 | [0.55; 1.31] |
| Moss 2015 | UK Age | RCT | 17.7^ | 1991-1999 | -0.13 | 0.09 | 0.88 | [0.74; 1.05] |
| Nystrom 2016 | Malmo I | RCT | 30 | 1976-1988 | -0.13 | 0.12 | 0.88 | [0.69; 1.11] |
| Nystrom 2016 | Stockholm | RCT | 25 | 1981-1985 | -0.06 | 0.17 | 0.94 | [0.67; 1.31] |
| Miller 2014 | CNBSS II | RCT | 21.9 | 1980-1984 | 0.02 | 0.15 | 1.02 | [0.76; 1.37] |
| Miller 2014 | CNBSS I | RCT | 21.9 | 1980-1984 | 0.05 | 0.11 | 1.05 | [0.85; 1.30] |

Risk Ratio

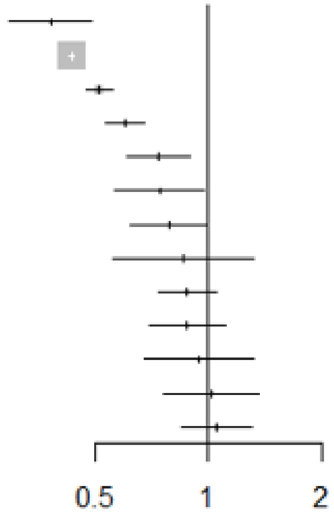

* Longest possible follow-up; ** Time since randomization; ^ Median; OBS: Observational studies; RCT: Randomized clinical trials;
KNCSP: Korean National Cancer Screening Program; BSA: BreastScreen Aotearoa; CNBSS: Canadian National Breast Screening Study;
HIP: Health Insurance Plan of Greater New York

#### Appendix 10 - Final Working Group Thresholds for GRADE

| Outcome | Threshold |
|---|---|
| Breast Cancer Mortality | Threshold One: For every 1,000 patients that undergo screening, there will be 1 less death due to breast cancer<br>Threshold Two: For every 1,000 patients that undergo screening, there will be 0.5 less deaths due to breast cancer |
| All-Cause Mortality | For every 1,000 patients that undergo screening, there will be 1 less death due to any cause |
| Breast Cancers Requiring Chemotherapy | For every 1,000 patients that undergo screening, there will be 2 less breast cancers requiring chemotherapy |
| Breast Cancers Requiring a Full Mastectomy (Versus Lumpectomy) | For every 1,000 patients that undergo screening, there will be 2 less breast cancers requiring mastectomy |
| Breast Cancers Requiring Axial Lymph Node Dissection | For every 1,000 patients that undergo screening, there will be 2 less breast cancers requiring axial lymph node dissection |
| Breast Cancers Requiring Radiotherapy | For every 1,000 patients that undergo screening, there will be 5 less breast cancers requiring radiotherapy |
| Breast Cancers Being Diagnosed at Stage III or Higher | For every 1,000 patients that undergo screening, there will be 2 less breast cancers diagnosed at stage III or higher |
| Breast Cancers Being Diagnosed at Stage II or Higher | For every 1,000 patients that undergo screening, there will be 3 less breast cancers diagnosed at stage II or higher |
| Metastatic cancer (stage IV) | For every 1,000 patients that undergo screening, there will be 1 less metastatic breast cancer diagnosed. |
| Overdiagnosed Cancers | For every 1,000 patients that undergo screening, there will be 5 more overdiagnosed breast cancers |
| Additional imaging +/- biopsy | For every 1,000 patients that undergo screening, 150 will experience one or more additional imaging +/- biopsy. |
| Additional imaging with biopsy | For every 1,000 patients that undergo screening, 15 will experience one or more additional imaging with biopsy. |
| Interval cancers | For every 1,000 patients that undergo screening, 6 will have an interval cancer. |

#### Appendix 11 - Working group survey for patient values and preferences towards screening for breast cancer

**Purpose:** To consider the working group's perspective on what effect (or risk reduction) patients would consider an important or trivial effect of a breast cancer screening program. An example of this effect could be: 1 less death per 1,000 patients screened. We want to know if you think this effect is important or trivial.

**Survey:** We will present a series of questions based on certain effects (i.e., different risk reduction scenarios for each outcome). We want to determine one threshold (e.g., 50 less per 1,000 patients) for each outcome that the working group agrees would be important to the majority of patients during decision-making (or a minimally important effect). At this point, the question is abstract because only one outcome is considered at a time (all the other benefits, harms, or burdens of interventions and their magnitude are not considered). These judgments are challenging. If possible, reflect on the question based on your knowledge of primary studies, previous focus groups, conversations with friends or family, or shared decision-making with patients.

**A few items to note:**

- *All effects are measured per 1,000 patients screened. If the risk reduction is <1 person (e.g., 0.4 less deaths per 1,000) you can think about it as per 10,000 patients screened (e.g., 4 less deaths per 10,000).*
- *Some scenarios involve a decrease (e.g., 1 less death per 1,000), while others involve an increase (e.g., 100 more false positives per 1,000) or simply state an associated rate (e.g., 2 interval cancers per 1,000).*
- *Please consider your answers in relation to a screening program consisting of 3 to 4 rounds of (annual or biannual) screening where patients are followed for 10 years after baseline.*
- *There's no right or wrong answer. All thresholds can be trivial, all can be important, or it can vary.*

1. Patients are considering the possibility of participating in a breast cancer screening program to reduce their risk of **dying from breast cancer**. Breast cancer screening lowers their risk in different scenarios in the table below over a period of 10 years. **Please choose an option that will reflect whether the majority of patients (>50%) would think this reduction in risk is an important or trivial effect.**

| | Risk reduction scenarios | Answer |
|---|---|---|
| a. | For every **1,000** patients that participate in screening, there will be **0.4** less deaths due to breast cancer over 10 years. | Choose an item. |
| b. | For every **1,000** patients that participate in screening, there will be **0.6** less deaths due to breast cancer over 10 years. | Choose an item. |
| c. | For every **1,000** patients that participate in screening, there will be **0.8** less deaths due to breast cancer over 10 years. | Choose an item. |
| d. | For every **1,000** patients that participate in screening, there will be **1** less death due to breast cancer over 10 years. | Choose an item. |
| e. | For every **1,000** patients that participate in screening, there will be **1.2** less deaths due to breast cancer over 10 years. | Choose an item. |
| f. | For every **1,000** patients that participate in screening, there will be **1.4** less deaths due to breast cancer over 10 years. | Choose an item. |

**If you would like to propose a different value in risk of dying from breast cancer, please note it here, otherwise please leave blank:**

| |
|---|
| *[Response]* |

2. Patients are considering the possibility of participating in a breast cancer screening program to reduce their risk of **dying from any cause**. Breast cancer screening lowers their risk in different scenarios in the table below over a period of 10 years. **Please choose an option that will reflect whether the majority of patients (>50%) would think this reduction in risk is an important or trivial effect.**

| | Risk reduction scenarios | Answer |
|---|---|---|
| a. | For every **1,000** patients that participate in screening, there will be **0.4** less deaths due to any cause over 10 years. | Choose an item. |
| b. | For every **1,000** patients that participate in screening, there will be **0.6** less deaths due to any cause over 10 years. | Choose an item. |
| c. | For every **1,000** patients that participate in screening, there will be **0.8** less deaths due to any cause over 10 years. | Choose an item. |
| d. | For every **1,000** patients that participate in screening, there will be **1** less death due to any cause over 10 years. | Choose an item. |
| e. | For every **1,000** patients that participate in screening, there will be **1.2** less deaths due to any cause over 10 years. | Choose an item. |
| f. | For every **1,000** patients that participate in screening, there will be **1.4** less deaths due to any cause over 10 years. | Choose an item. |

**If you would like to propose a different value in risk of dying from any cause, please note it here, otherwise please leave blank:**

| |
|---|
| *[Response]* |

3. Patients are considering the possibility of participating in a breast cancer screening program to reduce their risk of **breast cancers requiring chemotherapy (and associated complications)**. Breast cancer screening lowers their risk in different scenarios in the table below over a period of 10 years. **Please choose an option that will reflect whether the majority of patients (>50%) would think this reduction in risk is an important or trivial effect.**

| | Risk reduction scenarios | Answer |
|---|---|---|
| a. | For every **1,000** patients that participate in screening, there will be **1** less breast cancer requiring chemotherapy over 10 years. | Choose an item. |
| b. | For every **1,000** patients that participate in screening, there will be **2** less breast cancers requiring chemotherapy over 10 years. | Choose an item. |

| | | |
|---|---|---|
| c. | For every **1,000** patients that participate in screening, there will be **3** less breast cancers requiring chemotherapy over 10 years. | Choose an item. |
| d. | For every **1,000** patients that participate in screening, there will be **4** less breast cancers requiring chemotherapy over 10 years. | Choose an item. |
| e. | For every **1,000** patients that participate in screening, there will be **5** less breast cancers requiring chemotherapy over 10 years. | Choose an item. |
| f. | For every **1,000** patients that participate in screening, there will be **6** less breast cancers requiring chemotherapy over 10 years. | Choose an item. |

**If you would like to propose a different value in risk of breast cancers requiring chemotherapy, please note it here, otherwise please leave blank:**

| |
|---|
| *[Response]* |

**If you would like to propose a value in risk of breast cancers requiring a full mastectomy (vs lumpectomy, please note it here, otherwise please leave blank:**

| |
|---|
| *[Response]* |

**If you would like to propose a value in risk of breast cancers requiring axial lymph node dissection (vs sentinel lymph node biopsy), please note it here, otherwise please leave blank:**

| |
|---|
| *[Response]* |

**If you would like to propose a value in risk of breast cancers requiring radiotherapy, please note it here, otherwise please leave blank:**

| |
|---|
| *[Response]* |

4. Patients are considering the possibility of participating in a breast cancer screening program to reduce their risk of **being diagnosed with stage II or higher cancer**. Breast cancer screening lowers their risk in different scenarios in the table below over a period of 10 years. **Please choose an option that will reflect whether the majority of patients (>50%) would think this reduction in risk is an important or trivial effect.**

| | Risk reduction scenarios | Answer |
|---|---|---|
| a. | For every **1,000** patients that participate in screening, there will be **1** less ≥ stage II cancer diagnosed over 10 years. | Choose an item. |
| b. | For every **1,000** patients that participate in screening, there will be **2** less ≥ stage II cancer diagnosed over 10 years. | Choose an item. |
| c. | For every **1,000** patients that participate in screening, there will be **3** less ≥ stage II cancer diagnosed over 10 years. | Choose an item. |
| d. | For every **1,000** patients that participate in screening, there will be **4** less ≥ stage II cancer diagnosed over 10 years. | Choose an item. |
| e. | For every **1,000** patients that participate in screening, there will be **5** less ≥ stage II cancer diagnosed over 10 years. | Choose an item. |
| f. | For every **1,000** patients that participate in screening, there will be **6** less ≥ stage II cancer diagnosed over 10 years. | Choose an item. |

**If you would like to propose a different value in risk of being diagnosed with stage II or higher cancer, please note it here, otherwise please leave blank:**

| |
|---|
| *[Response]* |

5. Patients are considering the possibility of participating in a breast cancer screening program to reduce their risk of **being diagnosed with stage III or higher cancer**. Breast cancer screening lowers their risk in different scenarios in the table below over a period of 10 years. **Please choose an option that will reflect whether the majority of patients (>50%) would think this reduction in risk is an important or trivial effect.**

| | Risk reduction scenarios | Answer |
|---|---|---|
| a. | For every **1,000** patients that participate in screening, there will be **0.5** less ≥ stage III cancer diagnosed over 10 years. | Choose an item. |
| b. | For every **1,000** patients that participate in screening, there will be **1** less ≥ stage III cancer diagnosed over 10 years. | Choose an item. |
| c. | For every **1,000** patients that participate in screening, there will be **1.5** less ≥ stage III cancer diagnosed over 10 years. | Choose an item. |
| d. | For every **1,000** patients that participate in screening, there will be **2** less ≥ stage III cancer diagnosed over 10 years. | Choose an item. |
| e. | For every **1,000** patients that participate in screening, there will be **2.5** less ≥ stage III cancer diagnosed over 10 years. | Choose an item. |
| f. | For every **1,000** patients that participate in screening, there will be **3** less ≥ stage III cancer diagnosed over 10 years. | Choose an item. |

**If you would like to propose a different value in risk of being diagnosed with stage III or higher, please note it**

| |
|---|
| *[Response]* |

**here, otherwise please leave blank:**

6. Patients are considering the possibility of participating in a breast cancer screening program. Breast cancer screening may increase their risk of having an **overdiagnosed cancer** (i.e., a cancer that may never cause any harm or symptoms but leads to having a diagnosis (i.e., label) of cancer and may lead to unnecessary treatments or medications). **Please choose an option that will reflect whether the majority of patients (>50%) would think this increase in risk is an important or trivial effect.**

| | Risk reduction scenarios | Answer |
|---|---|---|
| a. | For every **1,000** patients that participate in screening, there will be **1** more overdiagnosed cancer over 10 years. | Choose an item. |
| b. | For every **1,000** patients that participate in screening, there will be **2** more overdiagnosed cancers over 10 years. | Choose an item. |
| c. | For every **1,000** patients that participate in screening, there will be **3** more overdiagnosed cancers over 10 years. | Choose an item. |
| d. | For every **1,000** patients that participate in screening, there will be **4** more overdiagnosed cancers over 10 years. | Choose an item. |
| e. | For every **1,000** patients that participate in screening, there will be **5** more overdiagnosed cancers over 10 years. | Choose an item. |
| f. | For every **1,000** patients that participate in screening, there will be **6** more overdiagnosed cancers over 10 years. | Choose an item. |

**If you would like to propose a different value in risk of having an overdiagnosed cancer, please note it here, otherwise please leave blank:**

*[Response]*

7. Patients are considering the possibility of participating in a breast cancer screening program. Breast cancer screening may lead to at least one **false positive result which could require either imaging alone or imaging plus biopsy** (i.e., a false alarm). **Please choose an option that will reflect whether the majority of patients (>50%) would think this increase in risk is an important or trivial effect.**

| | Risk reduction scenarios | Answer |
|---|---|---|
| a. | For every **1,000** patients that participate in screening, **100** will experience one or more false positives over 10 years. | Choose an item. |
| b. | For every **1,000** patients that participate in screening, **125** will experience one or more false positives over 10 years. | Choose an item. |
| c. | For every **1,000** patients that participate in screening, **150** will experience one or more false positives over 10 years. | Choose an item. |

| | | |
|---|---|---|
| d. | For every **1,000** patients that participate in screening, **175** will experience one or more false positives over 10 years. | Choose an item. |
| e. | For every **1,000** patients that participate in screening, **200** will experience one or more false positives over 10 years. | Choose an item. |
| f. | For every **1,000** patients that participate in screening, **225** will experience one or more false positives over 10 years. | Choose an item. |

**If you would like to propose a different value in having a false positive result which could require either imaging alone or imaging plus biopsy, please note it here, otherwise please leave blank:**

*[Response]*

8. Patients are considering the possibility of participating in a breast cancer screening program. Breast cancer screening may lead to at least one **false positive result that requires biopsy and imaging** (i.e., a false alarm). **Please choose an option that will reflect whether the majority of patients (>50%) would think this increase in risk is an important or trivial effect.**

| | Risk reduction scenarios | Answer |
|---|---|---|
| a. | For every **1,000** patients that participate in screening, **10** will experience one or more false positives over 10 years. | Choose an item. |
| b. | For every **1,000** patients that participate in screening, **12** will experience one or more false positives over 10 years. | Choose an item. |
| c. | For every **1,000** patients that participate in screening, **15** will experience one or more false positives over 10 years. | Choose an item. |
| d. | For every **1,000** patients that participate in screening, **18** will experience one or more false positives over 10 years. | Choose an item. |
| e. | For every **1,000** patients that participate in screening, **20** will experience one or more false positives over 10 years. | Choose an item. |
| f. | For every **1,000** patients that participate in screening, **25** will experience one or more false positives over 10 years. | Choose an item. |

**If you would like to propose a different value in risk of having a false positive result that requires biopsy and imaging, please note it here, otherwise please leave blank:**

*[Response]*

9. Patients are considering the possibility of participating in a breast cancer screening program. Breast cancer screening may lead to **interval cancers** (i.e., breast cancer that was not recognized during the screening or that develops between regular screens). These interval cancers may be more likely to occur with certain screening strategies (e.g., annual vs biannual). **Please choose an option that will reflect whether the majority of patients (>50%) would think this associated risk is an important or trivial effect.**

| | Risk reduction scenarios | Answer |
|---|---|---|
| a. | For every **1,000** patients that participate in screening, **2** will have an interval cancer over 10 years. | Choose an item. |
| b. | For every **1,000** patients that participate in screening, **6** will have an interval cancer over 10 years. | Choose an item. |
| c. | For every **1,000** patients that participate in screening, **8** will have an interval cancer over 10 years. | Choose an item. |
| d. | For every **1,000** patients that participate in screening, **10** will have an interval cancer over 10 years. | Choose an item. |
| e. | For every **1,000** patients that participate in screening, **12** will have an interval cancer over 10 years. | Choose an item. |
| f. | For every **1,000** patients that participate in screening, **15** will have an interval cancer over 10 years. | Choose an item. |

**If you would like to propose a different value in risk of having an interval cancer, please note it here, otherwise please leave blank:**

| |
|---|
| *[Response]* |

10. If you would like to leave any notes about any considerations you took when answering the survey or general comments, please note it here, otherwise please leave blank:

| |
|---|
| *[Response]* |

**Thank you!**

#### Appendix 12 – Baseline and lifetime risk calculations

**Table 1:** Calculated general population baseline risk and moderately increased risk for breast cancer mortality

| Age | Breast cancer mortality rate per 1,000 in screened group *Coldman* | SMR from screening *Coldman* | Breast cancer mortality rate per 1,000 in an unscreened group (General population risk) | Breast cancer mortality rate per 1,000 in an unscreened group (Moderately increased risk, family history) | Breast cancer mortality rate per 1,000 in an unscreened group (Moderately increased risk, dense breasts) |
|---|---|---|---|---|---|
| 40-49 | 1.02 | 0.56 | 1.8 | 2.9 | 3.5 |
| 50-59 | 1.98 | 0.60 | 3.3 | 5.3 | 6.3 |
| 60-69 | 2.50 | 0.58 | 4.3 | 6.9 | 8.2 |
| 70-79 | 3.97 | 0.65 | 6.1 | 9.7 | 11.6 |

- Breast cancer mortality rate in a Canadian population was estimated from the Pan-Canadian study (Coldman et al., 2014)
- One first degree relative increases the lifetime risk by 1.6 times (Engmann et al., 2017)
    - *E.g., For 40-49, the general population risk at 1.8 per 1,000 multiplied by 1.6 gives you 2.9 per 1,000 for family history.*
- Dense breasts increases risk by 1.9 times (Yueh-Hsia Chiu et al., 2010)
    - *E.g., For 40-49, the general population risk at 1.8 per 1,000 multiplied by 1.9 gives you 3.5 per 1,000 for dense breasts.*
- We assume the cancers diagnosed within the elevated risk population in unscreened women would have similar survival rates as those in the general risk population.

**Table 2:** Calculated baseline risk for all-cause mortality

| Age | Number of deaths in Canadian females 2021 | All-cause mortality rate per 1,000 over 10 years |
|---|---|---|
| 40-49 | 3,144 | 12.7 |
| 50-59 | 7,889 | 30.6 |
| 60-69 | 17,643 | 71.3 |
| 70-74 | 13,603 | 140.6 |

- Baseline risk has been calculated using death and age-specific mortality rate data from Statistics Canada in 2021

**Calculation to Estimate Potential Lifetime Breast Cancer Mortality Reduction for Screening Women in their 40s**

The lifetime risk of a Canadian woman dying of breast cancer is about 27.8/1000[51] . 17.5% of those deaths arise from cancers diagnosed in women between ages 40 and 49[52], so the risk of dying from those cancers is 4.86 / 1000. This rate is the result of some women who are unscreened and some who receive screening. Let p represent the fraction of women who are regularly screened.

An additional factor is that the cancer mortality statistics derive from a combination of women at general risk and those at increased risk. Let's assume that 95% are at general risk of dying from breast cancer if unscreened and 5% are at twice that risk. Further, we will make the somewhat arbitrary assumption that for these higher risk women screening with mammography does not reduce their risk of breast cancer death.

Let $R_u$ represent the risk of dying of breast cancer if unscreened for those at general risk.
From the Pan Canadian Study, the relative risk of dying of breast cancer for women who begin screening in their 40s is 0.56. We can then write for the overall mortality risk:

$$R_u [0.56 * p * .95] + R_u [(1-p) * .95] + 2 R_u * .05 = 4.86/1000$$

where the first term refers to the women at general risk who are screened, the second term to those at general risk who do not participate in screening and the third to those at elevated risk regardless of screening

So that $R_u = 4.86/1000 / \{[0.56 * p * .95] + R_u [(1-p) * .95] + 2 R_u * .05\}$

Then, if 60% of women historically participated in screening, Ru = 6.1/1000 and the absolute breast cancer mortality reduction for those who participate in screening is estimated as 0.44 * 6.1/1000 = **2.68 per 1000.**

#### Appendix 13 – List of excluded studies

**List of full-text articles excluded with reasons (n=175)**

##### Ineligible comparator (n=63)

**Autier, Philippe, Boniol, Magali, Koechlin, Alice, Pizot, Cecile, Boniol, Mathieu** (2017). Effectiveness of and overdiagnosis from mammography screening in the Netherlands: population based study *BMJ (Clinical research ed.)*, 359(#issue#), j5224

**Baines, Cornelia J., To, Teresa, Miller, Anthony B.** (2016). Revised estimates of overdiagnosis from the Canadian National Breast Screening Study *Preventive medicine*, 90(#issue#), 66-71

**Brunetti, Nicole, De Giorgis, Sara, Tosto, Simona, Garlaschi, Alessandro, Rescinito, Giuseppe, Massa, Barbara, Calabrese, Massimo, Tagliafico, Alberto Stefano** (2022). A Prospective Comparative Evaluation of Handheld Ultrasound Examination (HHUS) or Automated Ultrasound Examination (ABVS) in Women with Dense Breast *Diagnostics (Basel, Switzerland)*, 12(9), #Pages#

**Chang, Jung Min, Koo, Hye Ryoung, Moon, Woo Kyung** (2015). Radiologist-performed hand-held ultrasound screening at average risk of breast cancer: results from a single health screening center *Acta radiologica (Stockholm, Sweden : 1987)*, 56(6), 652-8

**de Munck, L., Siesling, S., Fracheboud, J., den Heeten, G. J., Broeders, M. J. M., de Bock, G. H.** (2020). Impact of mammographic screening and advanced cancer definition on the percentage of advanced-stage cancers in a steady-state breast screening programme in the Netherlands *British Journal of Cancer*, 123(7), 1191-1197

**Domingo, L., Sala, M., Louro, J., Bare, M., Barata, T., Ferrer, J., Carmona-Garcia, M. C., Comas, M., Castells, X.** (2019). Exploring the Role of Breast Density on Cancer Prognosis among Women Attending Population-Based Screening Programmes *Journal of Oncology*, 2019(#issue#), 1781762

**Duffy, Stephen W., Dibden, Amanda, Michalopoulos, Dimitrios, Offman, Judith, Parmar, Dharmishta, Jenkins, Jacquie, Collins, Beverley, Robson, Tony, Scorfield, Suzanne, Green, Kathryn, Hall, Clare, Liao, Xiao-Hui, Ryan, Michael, Johnson, Fiona, Stevens, Guy, Kearins, Olive, Sellars, Sarah, Patnick, Julietta** (2016). Screen detection of ductal carcinoma in situ and subsequent incidence of invasive interval breast cancers: a retrospective population-based study *The Lancet. Oncology*, 17(1), 109-14

**Elshof, Lotte E., Schaapveld, Michael, Rutgers, Emiel J., Schmidt, Marjanka K., de Munck, Linda, van Leeuwen, Flora E., Wesseling, Jelle** (2017). The method of detection of ductal carcinoma in situ has no therapeutic implications: results of a population-based cohort study *Breast cancer research : BCR*, 19(1), 26

**Garcia, Marilina, Redondo, Maximino, Zarcos, Irene, Louro, Javier, Rivas-Ruiz, Francisco, Tellez, Teresa, Perez, Diego, Medina Cano, Francisco, Machan, Kenza, Domingo, Laia, Del Mar Vernet, Maria, Padilla-Ruiz, Maria, Castells, Xavier, Sala, Maria** (2022). Impact of Detection Mode in a Large Cohort of Women Taking Part in a Breast Screening Program *European journal of breast health*, 18(2), 182-189

**Garcia-Fernandez, A., Barco, I., Fraile, M., Lain, J. M., Carmona, A., Gonzalez, S., Pessarrodona, A., Gimenez, N., Garcia-Font, M.** (2016). Factors predictive of mortality in a cohort of women surgically treated for breast cancer from 1997 to 2014 *International Journal of Gynecology and Obstetrics*, 134(2), 212-216

**Gram, Emma Grundtvig, Manso, Tulia Filipa Roberto, Heleno, Bruno, Siersma, Volkert, A Rogvi, Jessica, Brodersen, John Brandt** (2023). The long-term psychosocial consequences of screen-detected ductal carcinoma in situ and invasive breast cancer *Breast (Edinburgh, Scotland)*, 70(#issue#), 41-48

**Gram, Emma Grundtvig, Siersma, Volkert, Brodersen, John Brandt** (2023). Long-term psychosocial consequences of false-positive screening mammography: a cohort study with follow-up of 12-14 years in Denmark *BMJ open*, 13(4), e072188

**Greenwald, Z. R., Fregnani, J. H., Longatto-Filho, A., Watanabe, A., Mattos, J. S. C., Vazquez, F. L., Franco, E. L.** (2018). The performance of mobile screening units in a breast cancer screening program in Brazil *Cancer causes & control : CCC*, 29(2), 233-241

**Han, Hsin-Ju, Chu, Yuan-Chia, Wang, Jane, Lai, Yi-Chen, Tseng, Ling-Ming, Huang, Chi-Cheng** (2023). Characteristics of breast cancers detected by screening mammography in Taiwan: a single institute's experience *BMC women's health*, 23(1), 330

**Harding, C., Pompei, F., Burmistrov, D., Welch, H. G., Abebe, R., Wilson, R.** (2015). Breast cancer screening, incidence, and mortality across US counties *JAMA Internal Medicine*, 175(9), 1483-1489

**Harding, C., Pompei, F., Burmistrov, D., Wilson, R.** (2019). Long-term relationships between screening rates, breast cancer characteristics, and overdiagnosis in US counties, 1975-2009 *International Journal of Cancer*, 144(3), 476-488

**Heinavaara, Sirpa, Sarkeala, Tytti, Anttila, Ahti** (2016). Impact of organised mammography screening on breast cancer mortality in a case-control and cohort study *British journal of cancer*, 114(9), 1038-44

**Hellquist, Barbro Numan, Czene, Kamila, Hjalm, Anna, Nystrom, Lennarth, Jonsson, Hakan** (2015). Effectiveness of population-based service screening with mammography for women ages 40 to 49 years with a high or low risk of breast cancer: socioeconomic status, parity, and age at birth of first child *Cancer*, 121(2), 251-8

**Hersch, Jolyn, Barratt, Alexandra, McGeechan, Kevin, Jansen, Jesse, Houssami, Nehmat, Dhillon, Haryana, Jacklyn, Gemma, Irwig, Les, McCaffery, Kirsten** (2021). Informing Women About Overdetection in Breast Cancer Screening: Two-Year Outcomes From a Randomized Trial *Journal of the National Cancer Institute*, 113(11), 1523-1530

**Hofvind, Solveig, Skaane, Per, Elmore, Joann G., Sebuodegard, Sofie, Hoff, Solveig Roth, Lee, Christoph I.** (2014). Mammographic performance in a population-based screening program: before, during, and after the transition from screen-film to full-field digital mammography *Radiology*, 272(1), 52-62

**Ilenko, Anna, Sergent, Fabrice, Mercuzot, Antonin, Zitoun, Mickael, Chauffert, Bruno, Foulon, Arthur, Gondry, Jean, Chevreau, Julien** (2017). Could Patients Older than 75 Years Benefit from a Systematic Breast Cancer Screening Program? *Anticancer research*, 37(2), 903-907

**Ip, Eugenia C., Cohen-Hallaleh, Ruben B., Ng, Alexander K.** (2020). Extending Screening in "Elderly" Patients: Should We Consider a Selective Approach? *Clinical breast cancer*, 20(5), 377-381

**Jiang, Shu, Bennett, Debbie L., Rosner, Bernard A., Colditz, Graham A.** (2023). Longitudinal Analysis of Change in Mammographic Density in Each Breast and Its Association With Breast Cancer Risk *JAMA* *oncology*, 9(6), 808-814

**Johns, Louise E., Coleman, Derek A., Swerdlow, Anthony J., Moss, Susan M.** (2017). Effect of population breast screening on breast cancer mortality up to 2005 in England and Wales: an individual-level cohort study *British journal of cancer*, 116(2), 246-252

**Johnson, Kristin, Olinder, Jakob, Rosso, Aldana, Andersson, Ingvar, Lang, Kristina, Zackrisson, Sophia** (2023). False-positive recalls in the prospective Malmo Breast Tomosynthesis Screening Trial *European radiology*, #volume#(#issue#), #Pages#

**Kadhel, P., Borja De Mozota, D., Gaumond, S., Deloumeaux, J.** (2017). Characteristics of invasive breast cancer and overall survival of patients eligible for mass breast cancer screening in Guadeloupe compared to those of the preceding age group *Cancer Epidemiology*, 50(#issue#), 268-271

**Kelly, Caitriona, Fitzpatrick, Patricia, Quinn, Cecily, Flanagan, Fidelma, Connors, Alissa, Larke, Aideen, Mooney, Therese, Kennedy, Maria, Sheehan, Margaret, Bennett, Michael W., Brodie, Caroline, O'Doherty, Ann** (2022). Screen-detected ductal carcinoma in situ, 2008-2020: An observational study *Journal of medical screening*, 29(3), 172-177

**Kerlikowske, Karla, Scott, Christopher G., Mahmoudzadeh, Amir P., Ma, Lin, Winham, Stacey, Jensen, Matthew R., Wu, Fang Fang, Malkov, Serghei, Pankratz, V. Shane, Cummings, Steven R., Shepherd, John A., Brandt, Kathleen R., Miglioretti, Diana L., Vachon, Celine M.** (2018). Automated and Clinical Breast Imaging Reporting and Data System Density Measures Predict Risk for Screen-Detected and Interval Cancers: A Case-Control Study *Annals of internal medicine*, 168(11), 757-765

**Kerlikowske, Karla, Sprague, Brian L., Tosteson, Anna N. A., Wernli, Karen J., Rauscher, Garth H., Johnson, Dianne, Buist, Diana S. M., Onega, Tracy, Henderson, Louise M., O'Meara, Ellen S., Miglioretti, Diana L.** (2019). Strategies to Identify Women at High Risk of Advanced Breast Cancer During Routine Screening for Discussion of Supplemental Imaging *JAMA internal medicine*, 179(9), 1230-1239

**Kikuchi, M., Tsunoda, H., Koyama, T., Kawakita, T., Suzuki, K., Yamauchi, H., Takahashi, O., Saida, Y.** (2014). Opportunistic breast cancer screening by mammography in Japan for women in their 40s at our preventive medical center: Harm or benefit? *Breast Cancer*, 21(2), 135-139

**Kim, Soo-Yeon, Kim, Min Jung, Moon, Hee Jung, Yoon, Jung Hyun, Kim, Eun-Kyung** (2016). Application of the downgrade criteria to supplemental screening ultrasound for women with negative mammography but dense breasts *Medicine*, 95(44), e5279

**Koroukian, Siran M., Bakaki, Paul M., Htoo, Phyo Than, Han, Xiaozhen, Schluchter, Mark, Owusu, Cynthia, Cooper, Gregory S., Rose, Johnie, Flocke, Susan A.** (2017). The Breast and Cervical Cancer Early Detection Program, Medicaid, and breast cancer outcomes among Ohio's underserved women *Cancer*, 123(16), 3097-3106

**Lambeth, Chris, Burgess, Philip, Curtis, Jackie, Currow, David, Sara, Grant** (2023). Breast cancer screening participation in women using mental health services in NSW, Australia: a population study *Social psychiatry and psychiatric epidemiology*, #volume#(#issue#), #Pages#

**Lousdal, Mette Lise, Lash, Timothy L., Flanders, W. Dana, Brookhart, M. Alan, Kristiansen, Ivar Sonbo, Kalager, Mette, Stovring, Henrik** (2020). Negative controls to detect uncontrolled confounding in observational studies of mammographic screening comparing participants and non-participants *International journal of epidemiology*, 49(3), 1032-1042

**Lynge, Elsebeth, Vejborg, Ilse, Lillholm, Martin, Nielsen, Mads, Napolitano, George, von Euler-Chelpin, My** (2023). Breast density and risk of breast cancer *International journal of cancer*, 152(6), 1150-1158

**Ma, K. K., Lau, S. S. S., Cheung, P. S. Y.** (2014). Ductal carcinoma in situ in Chinese women undergoing opportunistic breast cancer screening *Surgical Practice*, 18(1), 8-14

**Masala, Giovanna, Assedi, Melania, Bendinelli, Benedetta, Pastore, Elisa, Gilio, Maria Antonietta, Mazzalupo, Vincenzo, Querci, Andrea, Fontana, Miriam, Duroni, Giacomo, Facchini, Luigi, Saieva, Calogero, Palli, Domenico, Ambrogetti, Daniela, Caini, Saverio** (2023). The FEDRA Longitudinal Study: Repeated Volumetric Breast Density Measures and Breast Cancer Risk *Cancers*, 15(6), #Pages#

**Morris, Melanie, Woods, Laura M., Rachet, Bernard** (2016). What might explain deprivation-specific differences in the excess hazard of breast cancer death amongst screen-detected women? Analysis of patients diagnosed in the West Midlands region of England from 1989 to 2011 *Oncotarget*, 7(31), 49939-49947

**Nagel, G., Oberaigner, W., Peter, R. S., Ulmer, H., Concin, H.** (2015). Evaluation of a mammography screening program within the population-based Vorarlberg Health Monitoring & Prevention Program (VHM&PP) *Cancer Epidemiology*, 39(6), 812-818

**Nelson, H. D., O'Meara, E. S., Kerlikowske, K., Balch, S., Miglioretti, D.** (2016). Factors associated with rates of false-positive and false-negative results from digital mammography screening: An analysis of registry data *Annals of Internal Medicine*, 164(4), 226-235

**Park, Hannah Lui, Chang, Jenny, Haridass, Vikram, Wang, Sophia S., Ziogas, Argyrios, Anton-Culver, Hoda** (2021). Mammography screening and mortality by risk status in the California teachers study *BMC cancer*, 21(1), 1341

**Pattacini, P., Nitrosi, A., Rossi, P. G., Duffy, S. W., Iotti, V., Ginocchi, V., Ravaioli, S., Vacondio, R., Mancuso, P., Ragazzi, M., Campari, C.** (2022). A Randomized Trial Comparing Breast Cancer Incidence and Interval Cancers after Tomosynthesis Plus Mammography versus Mammography Alone *Radiology*, 303(2), 256-266

**Roman, M., Castells, X., Hofvind, S., von Euler-Chelpin, M.** (2016). Risk of breast cancer after false-positive results in mammographic screening *Cancer Medicine*, 5(6), 1298-1306

**Roman, Marta, Hofvind, Solveig, von Euler-Chelpin, My, Castells, Xavier** (2019). Long-term risk of screen-detected and interval breast cancer after false-positive results at mammography screening: joint analysis of three national cohorts *British journal of cancer*, 120(2), 269-275

**Romero, A., Tora-Rocamora, I., Bare, M., Barata, T., Domingo, L., Ferrer, J., Tora, N., Comas, M., Merenciano, C., Macia, F., Castells, X., Sala, M.** (2016). Prevalence of persistent pain after breast cancer treatment by detection mode among participants in population-based screening programs *BMC Cancer*, 16(1), 735

**Sala, Maria, Domingo, Laia, Louro, Javier, Tora-Rocamora, Isabel, Bare, Marisa, Ferrer, Joana, Carmona-Garcia, Maria Carmen, Barata, Teresa, Roman, Marta, Macia, Francesc, Castells, Xavier**

(2018). Survival and Disease-Free Survival by Breast Density and Phenotype in Interval Breast Cancers *Cancer epidemiology, biomarkers & prevention : a publication of the American Association for Cancer Research, cosponsored by the American Society of Preventive Oncology*, 27(8), 908-916

**Schoenborn, Nancy L., Sheehan, Orla C., Roth, David L., Cidav, Tansu, Huang, Jin, Chung, Shang-En, Zhang, Talan, Lee, Sei, Xue, Qian-Li, Boyd, Cynthia M.** (2021). Association Between Receipt of Cancer Screening and All-Cause Mortality in Older Adults *JAMA network open*, 4(6), e2112062

**Seely, Jean Morag, Peddle, Susan Elizabeth, Yang, Huiming, Chiarelli, Anna M., McCallum, Megan, Narasimhan, Gopinath, Zakaria, Dianne, Earle, Craig C., Fung, Sharon, Bryant, Heather, Nicholson, Erika, Politis, Chris, Berg, Wendie A.** (2022). Breast Density and Risk of Interval Cancers: The Effect of Annual Versus Biennial Screening Mammography Policies in Canada *Canadian Association of Radiologists journal = Journal l'Association canadienne des radiologistes*, 73(1), 90-100

**Seneviratne, Sanjeewa, Campbell, Ian, Scott, Nina, Shirley, Rachel, Lawrenson, Ross** (2015). Impact of mammographic screening on ethnic and socioeconomic inequities in breast cancer stage at diagnosis and survival in New Zealand: a cohort study *BMC public health*, 15(#issue#), 46

**Sprague, Brian L., Chen, Shuai, Miglioretti, Diana L., Gard, Charlotte C., Tice, Jeffrey A., Hubbard, Rebecca A., Aiello Bowles, Erin J., Kaufman, Peter A., Kerlikowske, Karla** (2023). Cumulative 6-Year Risk of Screen-Detected Ductal Carcinoma In Situ by Screening Frequency *JAMA network open*, 6(2), e230166

**Timmermans, Lore, Bleyen, Luc, Bacher, Klaus, Van Herck, Koen, Lemmens, Kim, Van Ongeval, Chantal, Van Steen, Andre, Martens, Patrick, De Brabander, Isabel, Goossens, Mathieu, Thierens, Hubert** (2017). Screen-detected versus interval cancers: Effect of imaging modality and breast density in the Flemish Breast Cancer Screening Programme *European radiology*, 27(9), 3810-3819

**Toth, D., Varga, Z., Toth, J., Arkosy, P., Sebo, E.** (2018). Short- and Long-Term (10-year) Results of an Organized, Population-Based Breast Cancer Screening Program: Comparative, Observational Study from Hungary *World journal of surgery*, 42(5), 1396-1402

**Vacek, P. M., Skelly, J. M.** (2015). A prospective study of the use and effects of screening mammography in women aged 70 and older *Journal of the American Geriatrics Society*, 63(1), 1-7

**van Bommel, Rob M. G., Weber, Roy, Voogd, Adri C., Nederend, Joost, Louwman, Marieke W. J., Venderink, Dick, Strobbe, Luc J. A., Rutten, Matthieu J. C., Plaisier, Menno L., Lohle, Paul N., Hooijen, Marianne J. H., Tjan-Heijnen, Vivianne C. G., Duijm, Lucien E. M.** (2017). Interval breast cancer characteristics before, during and after the transition from screen-film to full-field digital screening mammography *BMC cancer*, 17(1), 315

**van der Waal, D., Verbeek, A. L. M., Broeders, M. J. M.** (2018). Breast density and breast cancer-specific survival by detection mode *BMC Cancer*, 18(1), 386

**van Luijt, P. A., Heijnsdijk, E. A. M., Fracheboud, J., Overbeek, L. I. H., Broeders, M. J. M., Wesseling, J., den Heeten, G. J., de Koning, H. J.** (2016). The distribution of ductal carcinoma in situ (DCIS) grade in 4232 women and its impact on overdiagnosis in breast cancer screening *Breast Cancer Research*, 18(1), 47

**Waheed, Naima, Hameed, Maha, Alendijani, Yaser Abdullah, Al-Juwaid, Shorouq, Alkhenizan, Abdullah H.** (2023). Breast Cancer Diagnosis and Survival among Patients Diagnosed by a Structured Community

Based Screening Program Compared to Opportunistic Diagnosis: A Case Control Study *Asian Pacific journal of cancer prevention : APJCP*, 24(3), 923-927

**Walbaum, Benjamin, Puschel, Klaus, Medina, Lidia, Merino, Tomas, Camus, Mauricio, Razmilic, Dravna, Navarro, Maria Elena, Dominguez, Francisco, Cordova-Delgado, Miguel, Pinto, Mauricio P., Acevedo, Francisco, Sanchez, Cesar** (2021). Screen-detected breast cancer is associated with better prognosis and survival compared to self-detected/symptomatic cases in a Chilean cohort of female patients *Breast cancer research and treatment*, 189(2), 561-569

**Walpole, Euan, Dunn, Nathan, Youl, Philippa, Harden, Hazel, Furnival, Colin, Moore, Julie, Taylor, Kate, Evans, Elizabeth, Philpot, Shoni** (2020). Nonbreast cancer incidence, treatment received and outcomes: Are there differences in breast screening attendees versus nonattendees? *International journal of cancer*, 147(3), 856-865

**Weigel, Stefanie, Heindel, Walter, Dietz, Caroline, Meyer-Johann, Ulrike, Graewingholt, Axel, Hense, Hans Werner** (2020). Stratification of Breast Cancer Risk in Terms of the Influence of Age and Mammographic density *Stratifizierung des Brustkrebsrisikos hinsichtlich der Einflusse von Alter und mammografischer Dichte.*, 192(7), 678-685

**Woods, L. M., Rachet, B., O'Connell, D. L., Lawrence, G., Coleman, M. P.** (2016). Are international differences in breast cancer survival between Australia and the UK present amongst both screen-detected women and non-screen-detected women? survival estimates for women diagnosed in West Midlands and New South Wales 1997-2006 *International Journal of Cancer*, 138(10), 2404-2414

**Yi, Ann, Jang, Myoung-Jin, Yim, Dahae, Kwon, Bo Ra, Shin, Sung Ui, Chang, Jung Min** (2021). Addition of Screening Breast US to Digital Mammography and Digital Breast Tomosynthesis for Breast Cancer Screening in Women at Average Risk *Radiology*, 298(3), 568-575

**Yuan, Yan, Li, Maoji, Yang, Jing, Elliot, Tracy, Dabbs, Kelly, Dickinson, James A., Fisher, Stacey, Winget, Marcy** (2016). Factors related to breast cancer detection mode and time to diagnosis in Alberta, Canada: a population-based retrospective cohort study *BMC health services research*, 16(#issue#), 65

##### Ineligible study design (n=41)

**Amorim, R. B. F., Campos, F. S. M., Silveira, L. L., Archangelo Junior, I., Novo, N. F., Ferreira, L. M., Veiga, D. F.** (2021). Effectiveness of Brazilian national health policy for mammogram screening in women aged over 50 years *Breast Journal*, 27(1), 82-83

**Anonymous** (2015). Mammographic screening for breast cancer. Overdiagnosis: an insidious adverse effect of screening *Prescrire international*, 24(162), 186-191

**Anonymous** (2015). Mammographic breast cancer screening. Part II. Non-randomised comparisons: results similar to those of randomised trials *Prescrire international*, 24(159), 99-102

**Bahl, M., Lehman, C. D.** (2019). Breast Cancer Screening Using Digital Breast Tomosynthesis: Not All Mammography Is Equal *JAMA Oncology*, 5(5), 642-643

**Bahl, Manisha** (2022). Screening MRI in Women at Intermediate Breast Cancer Risk: An Update of the Recent Literature *Journal of breast imaging*, 4(3), 231-240

**Beau, Anna-Belle, Napolitano, George M., Ewertz, Marianne, Vejborg, Ilse, Schwartz, Walter, Andersen, Per K., Lynge, Elsebeth** (2020). Impact of chronic diseases on effect of breast cancer screening *Cancer medicine*, 9(11), 3995-4003

**Bulliard, Jean-Luc, Beau, Anna-Belle, Njor, Sisse, Wu, Wendy Yi-Ying, Procopio, Pietro, Nickson, Carolyn, Lynge, Elsebeth** (2021). Breast cancer screening and overdiagnosis *International journal of cancer*, #volume#(#issue#), #Pages#

**Chang, R. W. J., Jen, G. H. H., Lin, K. C., Cheng, T. C., Chuang, S. Y., Pan, S. L., Chen, T. H. H., Yen, A. M. F.** (2022). Evaluating the effectiveness of population-based breast cancer service screening: an analysis of parsimonious patient survival information with the time-varying Cox model *International Journal of Epidemiology*, 51(6), 1910-1919

**de Gelder, Rianne, Heijnsdijk, Eveline A. M., Fracheboud, Jacques, Draisma, Gerrit, de Koning, Harry J.** (2015). The effects of population-based mammography screening starting between age 40 and 50 in the presence of adjuvant systemic therapy *International journal of cancer*, 137(1), 165-72

**Fletcher, S. W.** (2014). Annual mammography screening did not reduce long-term breast cancer mortality in women 40 to 59 years of age *ACP journal club*, 160(10), 1

**Garcia-Albeniz, X., Hernan, M. A., Logan, R. W.** (2020). Continuation of mammography screening in women older than 70 years *Annals of Internal Medicine*, 172(6), I-22

**Houssami, Nehmat** (2017). Overdiagnosis of breast cancer in population screening: does it make breast screening worthless? *Cancer biology & medicine*, 14(1), 1-8

**Huang, X., Do, K. A.** (2015). A novel case-control design to estimate the extent of over-diagnosis of breast cancer due to organised population-based mammography screening *Breast Diseases*, 26(3), 211-214

**Hubner, J., Katalinic, A., Waldmann, A., Kraywinkel, K.** (2020). Long-term Incidence and Mortality Trends for Breast Cancer in Germany *Geburtshilfe und Frauenheilkunde*, 80(6), 611-618

**Jacklyn, G., Howard, K., Irwig, L., Houssami, N., Hersch, J., Barratt, A.** (2017). Impact of extending screening mammography to older women Information to support informed choices *International Journal of Cancer*, 141(8), 1540-1550

**Jacklyn, Gemma, McGeechan, Kevin, Irwig, Les, Houssami, Nehmat, Morrell, Stephen, Bell, Katy, Barratt, Alexandra** (2017). Trends in stage-specific breast cancer incidence in New South Wales, Australia: insights into the effects of 25 years of screening mammography *Breast cancer research and treatment*, 166(3), 843-854

**Jorgensen, Karsten Juhl, Gotzsche, Peter C., Kalager, Mette, Zahl, Per-Henrik** (2017). Breast Cancer Screening in Denmark: A Cohort Study of Tumor Size and Overdiagnosis *Annals of internal medicine*, 166(5), 313-323

**Kalager, M., Loberg, M., Bretthauer, M., Adami, H. O.** (2014). Comparative analysis of breast cancer mortality following mammography screening in Denmark and Norway *Annals of oncology : official journal of the European Society for Medical Oncology*, 25(6), 1137-43

**Kopans, D. B.** (2014). Twenty five year follow-up for breast cancer incidence and mortality of the Canadian National Breast Screening Study: Randomised screening trial *Breast Diseases*, 25(3), 223-226

**Kroenke, K.** (2014). Are the harms of false-positive screening test results minimal or meaningful? *JAMA Internal Medicine*, 174(6), 961-963

**Kuhl, C. K.** (2019). Underdiagnosis is the main challenge in breast cancer screening *The Lancet Oncology*, 20(8), 1044-1046

**Lousdal, Mette L., Moller, Mette H., Kristiansen, Ivar S., Kalager, Mette, Wisloff, Torbjorn, Stovring, Henrik** (2018). The Screening Illustrator: separating the effects of lead-time and overdiagnosis in mammography screening *European journal of public health*, 28(6), 1138-1142

**Miller, A. B.** (2020). Final results of the UK Age trial on breast cancer screening age *The Lancet Oncology*, 21(9), 1125-1126

**Miller, A. B., Fletcher, S. W.** (2014). Annual mammography screening did not reduce long-term breast cancer mortality in women 40 to 59 years of age *Annals of Internal Medicine*, 160(10), JC7

**Miller, A. B., Wall, C., Baines, C. J., Sun, P., To, T., Narod, S. A.** (2014). Twenty-five-year follow-up for breast cancer incidence and mortality of the canadian national breast screening study: Randomized screening trial *Obstetrical and Gynecological Survey*, 69(6), 329-330

**Molassiotis, A., Tyrovolas, S., Gine-Vazquez, I., Yeo, W., Aapro, M., Herrstedt, J.** (2021). Organized breast cancer screening not only reduces mortality from breast cancer but also significantly decreases disability-adjusted life years: analysis of the Global Burden of Disease Study and screening programme availability in 130 countries *ESMO Open*, 6(3), 100111

**Moller, Mette H., Lousdal, Mette Lise, Kristiansen, Ivar S., Stovring, Henrik** (2019). Effect of organized mammography screening on breast cancer mortality: A population-based cohort study in Norway *International journal of cancer*, 144(4), 697-706

**Movik, Espen, Dalsbo, Therese Kristine, Fagelund, Beate Charlotte, Friberg, Eva Godske, Haheim, Lise Lund, Skar, Ase** (2017). Digital Breast Tomosynthesis with Hologic 3D Mammography Selenia Dimensions System for Use in Breast Cancer Screening: A Single Technology Assessment *#journal#*, #volume#(#issue#), #Pages#

**Plaxco, J. S., Geiser, W., Yang, W.** (2015). Effect of integrating 3D-mammography (digital breast tomosynthesis) with 2D-mammography on radiologists' true-positive and false-positive detection in a population breast screening trial *Breast Diseases*, 26(1), 45-48

**Rosenberg, Karen** (2019). Benefits Of Screening Ultrasonography For Breast Cancer May Not Outweigh Harms *The American journal of nursing*, 119(7), 62-63

**Sebuodegard, S., Botteri, E., Hofvind, S.** (2020). Breast cancer mortality after implementation of organized population-based breast cancer screening in Norway *Journal of the National Cancer Institute*, 112(8), 839-846

**Shen, S. J., Xu, Y. L., Zhou, Y. D., Ren, G. S., Jiang, J., Jiang, H. C., Zhang, J., Li, B., Jin, F., Li, Y. P., Xie, F. M., Shi, Y., Wang, Z. D., Sun, M., Yuan, S. H., Yu, J. J., Chen, Y., Sun, Q.** (2021). [A comparative study of breast cancer mass screening and opportunistic screening in Chinese women] *Zhonghua wai ke za zhi [Chinese journal of surgery]*, 59(2), 109-115

**Slomski, A.** (2020). Long-term Mortality Outcomes in Early Breast Cancer Screening Trial *JAMA*, 324(20), 2020

**Stout, N. K., Lee, S. J., Schechter, C. B., Kerlikowske, K., Alagoz, O., Berry, D., Buist, D. S. M., Cevik, M., Chisholm, G., De Koning, H. J., Huang, H., Hubbard, R. A., Miglioretti, D. L., Munsell, M. F., Trentham-Dietz, A., Van Ravesteyn, N. T., Tosteson, A. N. A., Mandelblatt, J. S.** (2014). Benefits, harms, and costs for breast cancer screening after US implementation of digital mammography *Journal of the National Cancer Institute*, 106(6), #Pages#

**Tabar, Laszlo, Yen, Amy Ming-Fang, Wu, Wendy Yi-Ying, Chen, Sam Li-Sheng, Chiu, Sherry Yueh-Hsia, Fann, Jean Ching-Yuan, Ku, May Mei-Sheng, Smith, Robert A., Duffy, Stephen W., Chen, Tony Hsiu-Hsi** (2015). Insights from the breast cancer screening trials: how screening affects the natural history of breast cancer and implications for evaluating service screening programs *The breast journal*, 21(1), 13-20

**Van Ravesteyn, N. T., Schechter, C. B., Hampton, J. M., Alagoz, O., Van Den Broek, J. J., Kerlikowske, K., Mandelblatt, J. S., Miglioretti, D. L., Sprague, B. L., Stout, N. K., De Koning, H. J., Trentham-Dietz, A., Tosteson, A. N. A.** (2021). Trade-Offs between Harms and Benefits of Different Breast Cancer Screening Intervals among Low-Risk Women *Journal of the National Cancer Institute*, 113(8), 1017-1026

**Worcester, S.** (2014). Annual mammography at age 40-59 provided no survival benefit *Oncology Report*, #volume#(MAR), 13

**Yip, C. H.** (2019). Downstaging is more important than screening for asymptomatic breast cancer *The Lancet Global Health*, 7(6), e690-e691

**Yoshida, Y., Schmaltz, C. L., Jackson-Thompson, J., Simoes, E. J.** (2018). The impact of screening on cancer incidence and mortality in Missouri, USA, 2004-2013 *Public Health*, 154(#issue#), 51-58

**Zahl, Per-Henrik, Kalager, Mette, Suhrke, Pal, Nord, Erik** (2020). Quality-of-life effects of screening mammography in Norway *International journal of cancer*, 146(8), 2104-2112

#### Systematic or scoping review (n=31)

**Autier, Philippe, Boniol, Mathieu, Smans, Michel, Sullivan, Richard, Boyle, Peter** (2016). Observed and Predicted Risk of Breast Cancer Death in Randomized Trials on Breast Cancer Screening *PloS one*, 11(4), e0154113

**Autier, Philippe, Boniol, Mathieu, Smans, Michel, Sullivan, Richard, Boyle, Peter** (2015). Statistical analyses in Swedish randomised trials on mammography screening and in other randomised trials on cancer screening: a systematic review *Journal of the Royal Society of Medicine*, 108(11), 440-50

**Braithwaite, Dejana, Walter, Louise C., Izano, Monika, Kerlikowske, Karla** (2016). Benefits and Harms of Screening Mammography by Comorbidity and Age: A Qualitative Synthesis of Observational Studies and Decision Analyses *Journal of general internal medicine*, 31(5), 561-72

**Canelo-Aybar, Carlos, Ferreira, Diogenes S., Ballesteros, Monica, Posso, Margarita, Montero, Nadia, Sola, Ivan, Saz-Parkinson, Zuleika, Lerda, Donata, Rossi, Paolo G., Duffy, Stephen W., Follmann, Markus, Grawingholt, Axel, Alonso-Coello, Pablo** (2021). Benefits and harms of breast cancer mammography screening for women at average risk of breast cancer: A systematic review for the European Commission Initiative on Breast Cancer *Journal of medical screening*, 28(4), 389-404

**Canelo-Aybar, Carlos, Posso, Margarita, Montero, Nadia, Sola, Ivan, Saz-Parkinson, Zuleika, Duffy, Stephen W., Follmann, Markus, Grawingholt, Axel, Giorgi Rossi, Paolo, Alonso-Coello, Pablo** (2022). Benefits and harms of annual, biennial, or triennial breast cancer mammography screening for women at average risk of breast cancer: a systematic review for the European Commission Initiative on Breast Cancer (ECIBC) *British journal of cancer*, 126(4), 673-688

**Dibden, Amanda, Offman, Judith, Duffy, Stephen W., Gabe, Rhian** (2020). Worldwide Review and Meta-Analysis of Cohort Studies Measuring the Effect of Mammography Screening Programmes on Incidence-Based Breast Cancer Mortality *Cancers*, 12(4), #Pages#

**Flemban, Arwa F.** (2023). Overdiagnosis Due to Screening Mammography for Breast Cancer among Women Aged 40 Years and Over: A Systematic Review and Meta-Analysis *Journal of personalized medicine*, 13(3), #Pages#

**Hamashima, Chisato, Hamashima C, Chisato, Hattori, Masakazu, Honjo, Satoshi, Kasahara, Yoshio, Katayama, Takafumi, Nakai, Masahiro, Nakayama, Tomio, Morita, Takako, Ohta, Koji, Ohnuki, Koji, Sagawa, Motoyasu, Saito, Hiroshi, Sasaki, Seiju, Shimada, Tomoyuki, Sobue, Tomotaka, Suto, Akihiko** (2016). The Japanese Guidelines for Breast Cancer Screening *Japanese journal of clinical oncology*, 46(5), 482-92

**Hill, C.** (2014). Breast cancer screening *Presse Medicale*, 43(5), 501-509

**Houssami, Nehmat, Hunter, Kylie** (2017). The epidemiology, radiology and biological characteristics of interval breast cancers in population mammography screening *NPJ breast cancer*, 3(#issue#), 12

**Irvin, Veronica L., Kaplan, Robert M.** (2014). Screening mammography & breast cancer mortality: meta-analysis of quasi-experimental studies *PloS one*, 9(6), e98105

**Jacklyn, Gemma, Glasziou, Paul, Macaskill, Petra, Barratt, Alexandra** (2016). Meta-analysis of breast cancer mortality benefit and overdiagnosis adjusted for adherence: improving information on the effects of attending screening mammography *British journal of cancer*, 114(11), 1269-76

**Khrouf, Salim, Letaief Ksontini, Feryel, Ayadi, Mouna, Belhaj Ali Rais, Henda, Mezlini, Amel** (2020). Breast cancer screening: a dividing controversy *La Tunisie medicale*, 98(1), 22-34

**Li, T., Marinovich, M. L., Houssami, N.** (2018). Digital breast tomosynthesis (3D mammography) for breast cancer screening and for assessment of screen-recalled findings: review of the evidence *Expert Review of Anticancer Therapy*, 18(8), 785-791

**Loberg, Magnus, Lousdal, Mette Lise, Bretthauer, Michael, Kalager, Mette** (2015). Benefits and harms of mammography screening *Breast cancer research : BCR*, 17(#issue#), 63

**Mandrik, Olena, Zielonke, Nadine, Meheus, Filip, Severens, J. L. Hans, Guha, Neela, Herrero Acosta,**

**Rolando, Murillo, Raul** (2019). Systematic reviews as a 'lens of evidence': Determinants of benefits and harms of breast cancer screening *International journal of cancer*, 145(4), 994-1006

**Meggetto, Olivia, Peirson, Leslea, Yakubu, Mafo, Farid-Kapadia, Mufiza, Costa-Fagbemi, Michelle, Baidoobonso, Shamara, Moffatt, Jessica, Chun, Lauren, Chiarelli, Anna M., Muradali, Derek** (2019). Breast cancer risk and breast screening for trans people: an integration of 3 systematic reviews *CMAJ open*, 7(3), E598-E609

**Melnikow, Joy, Fenton, Joshua J., Whitlock, Evelyn P., Miglioretti, Diana L., Weyrich, Meghan S., Thompson, Jamie H., Shah, Kunal** (2016). Supplemental Screening for Breast Cancer in Women With Dense Breasts: A Systematic Review for the U.S. Preventive Service Task Force *#journal#*, #volume#(#issue#), #Pages#

**Myers, Evan R., Moorman, Patricia, Gierisch, Jennifer M., Havrilesky, Laura J., Grimm, Lars J., Ghate, Sujata, Davidson, Brittany, Mongtomery, Ranee Chatterjee, Crowley, Matthew J., McCrory, Douglas C., Kendrick, Amy, Sanders, Gillian D.** (2015). Benefits and Harms of Breast Cancer Screening: A Systematic Review *JAMA*, 314(15), 1615-34

**Nelson, Heidi D., Cantor, Amy, Humphrey, Linda, Fu, Rochelle, Pappas, Miranda, Daeges, Monica, Griffin, Jessica** (2016). Screening for Breast Cancer: A Systematic Review to Update the 2009 U.S. Preventive Services Task Force Recommendation *#journal#*, #volume#(#issue#), #Pages#

**Nelson, Heidi D., Fu, Rochelle, Cantor, Amy, Pappas, Miranda, Daeges, Monica, Humphrey, Linda** (2016). Effectiveness of Breast Cancer Screening: Systematic Review and Meta-analysis to Update the 2009 U.S. Preventive Services Task Force Recommendation *Annals of internal medicine*, 164(4), 244-55

**Nelson, Heidi D., Pappas, Miranda, Cantor, Amy, Griffin, Jessica, Daeges, Monica, Humphrey, Linda** (2016). Harms of Breast Cancer Screening: Systematic Review to Update the 2009 U.S. Preventive Services Task Force Recommendation *Annals of internal medicine*, 164(4), 256-67

**Oeffinger, K. C., Fontham, E. T. H., Etzioni, R., Herzig, A., Michaelson, J. S., Shih, Y. C. T., Walter, L. C., Church, T. R., Flowers, C. R., LaMonte, S. J., Wolf, A. M. D., DeSantis, C., Lortet-Tieulent, J., Andrews, K., Manassaram-Baptiste, D., Saslow, D., Smith, R. A., Brawley, O. W., Wender, R.** (2015). Breast cancer screening for women at average risk: 2015 Guideline update from the American cancer society *JAMA - Journal of the American Medical Association*, 314(15), 1599-1614

**Pace, L. E., Keating, N. L.** (2014). A systematic assessment of benefits and risks to guide breast cancer screening decisions *JAMA*, 311(13), 1327-1335

**Raichand, S., Dunn, A. G., Ong, M. S., Bourgeois, F. T., Coiera, E., Mandl, K. D.** (2017). Conclusions in systematic reviews of mammography for breast cancer screening and associations with review design and author characteristics *Systematic Reviews*, 6(1), 105

**Saquib, Nazmus, Saquib, Juliann, Ioannidis, John P. A.** (2015). Does screening for disease save lives in asymptomatic adults? Systematic review of meta-analyses and randomized trials *International journal of epidemiology*, 44(1), 264-77

**Siu, A. L.** (2016). Screening for breast cancer: U.S. Preventive services task force recommendation statement *Annals of Internal Medicine*, 164(4), 279-296

**van den Ende, Caroline, Oordt-Speets, Anouk M., Vroling, Hilde, van Agt, Heleen M. E.** (2017). Benefits and harms of breast cancer screening with mammography in women aged 40-49 years: A systematic review *International journal of cancer*, 141(7), 1295-1306

**Velentzis, Louiza S., Freeman, Victoria, Campbell, Denise, Hughes, Suzanne, Luo, Qingwei, Steinberg, Julia, Egger, Sam, Mann, G. Bruce, Nickson, Carolyn** (2023). Breast Cancer Risk Assessment Tools for Stratifying Women into Risk Groups: A Systematic Review *Cancers*, 15(4), #Pages#

**Voss, Theis, Krag, Mikela, Martiny, Frederik, Heleno, Bruno, Jorgensen, Karsten Juhl, Brandt Brodersen, John** (2023). Quantification of overdiagnosis in randomised trials of cancer screening: an overview and re-analysis of systematic reviews *Cancer epidemiology*, 84(#issue#), 102352

**Zielonke, Nadine, Gini, Andrea, Jansen, Erik E. L., Anttila, Ahti, Segnan, Nereo, Ponti, Antonio, Veerus, Piret, de Koning, Harry J., van Ravesteyn, Nicolien T., Heijnsdijk, Eveline A. M.** (2020). Evidence for reducing cancer-specific mortality due to screening for breast cancer in Europe: A systematic review *European journal of cancer (Oxford, England : 1990)*, 127(#issue#), 191-206

##### Cohort study (all participants have breast cancer) (n=18)

**Barco, Israel, Chabrera, Carol, Garcia Font, Marc, Gimenez, Nuria, Fraile, Manel, Lain, Josep Maria, Piqueras, Merce, Vidal, M. Carmen, Torras, Merce, Gonzalez, Sonia, Pessarrodona, Antoni, Barco, Josep, Cassado, Jordi, Garcia Fernandez, Antonio** (2015). Comparison of Screened and Nonscreened Breast Cancer Patients in Relation to Age: A 2-Institution Study Clinical breast cancer, 15(6), 482-9

**Beau, Anna-Belle, Andersen, Per Kragh, Vejborg, Ilse, Lynge, Elsebeth** (2018). Limitations in the Effect of Screening on Breast Cancer Mortality Journal of clinical oncology : official journal of the American Society of Clinical Oncology, 36(30), 2988-2994

**Cedolini, Carla, Bertozzi, Serena, Londero, Ambrogio P., Bernardi, Sergio, Seriau, Luca, Concina, Serena, Cattin, Federico, Risaliti, Andrea** (2014). Type of breast cancer diagnosis, screening, and survival Clinical breast cancer, 14(4), 235-40

**Choi, Kui Son, Yoon, Minjoo, Song, Seung Hoon, Suh, Mina, Park, Boyoung, Jung, Kyu Won, Jun, Jae Kwan** (2018). Effect of mammography screening on stage at breast cancer diagnosis: results from the Korea National Cancer Screening Program Scientific reports, 8(1), 8882

**Duffy, S. W., Tabar, L., Yen, A. M. F., Dean, P. B., Smith, R. A., Jonsson, H., Tornberg, S., Chen, S. L. S., Chiu, S. Y. H., Fann, J. C. Y., Ku, M. M. S., Wu, W. Y. Y., Hsu, C. Y., Chen, Y. C., Svane, G., Azavedo, E., Grundstrom, H., Sunden, P., Leifland, K., Frodis, E., Ramos, J., Epstein, B., Akerlund, A., Sundbom, A., Bordas, P., Wallin, H., Starck, L., Bjorkgren, A., Carlson, S., Fredriksson, I., Ahlgren, J., Ohman, D., Holmberg, L., Chen, T. H. H.** (2020). Mammography screening reduces rates of advanced and fatal breast cancers: Results in 549,091 women Cancer, 126(13), 2971-2979

**Garcia Fernandez, A., Chabrera, C., Garcia Font, M., Fraile, M., Lain, J. M., Gonzalez, S., Corral, C., Torras, M., Torres, J., Teixido, M., Barco, I., Lopez, R., Gonzalez, C., Pessarrodona, A., Gimenez, N.** (2014). Mortality and recurrence patterns of breast cancer patients diagnosed under a screening programme versus comparable non-screened breast cancer patients from the same population: Analytical survey from 2002 to 2012 Tumor Biology, 35(3), 1945-1953

**Herrmann, C., Vounatsou, P., Thurlimann, B., Probst-Hensch, N., Rothermundt, C., Ess, S.** (2018). Impact of mammography screening programmes on breast cancer mortality in Switzerland, a country with different regional screening policies BMJ Open, 8(3), e017806

**Lim, Zi Lin, Ho, Peh Joo, Khng, Alexis Jiaying, Yeoh, Yen Shing, Ong, Amanda Tse Woon, Tan, Benita Kiat Tee, Tan, Ern Yu, Tan, Su-Ming, Lim, Geok Hoon, Lee, Jung Ah, Tan, Veronique Kiak-Mien, Hu, Jesse, Li, Jingmei, Hartman, Mikael** (2022). Mammography screening is associated with more favourable breast cancer tumour characteristics and better overall survival: case-only analysis of 3739 Asian breast cancer patients BMC medicine, 20(1), 239

**Luu, Xuan Quy, Lee, Kyeongmin, Jun, Jae Kwan, Suh, Mina, Jung, Kyu-Won, Choi, Kui Son** (2022). Effect of mammography screening on the long-term survival of breast cancer patients: results from the National Cancer Screening Program in Korea Epidemiology and health, 44(#issue#), e2022094

**Niraula, Saroj, Biswanger, Natalie, Hu, PingZhao, Lambert, Pascal, Decker, Kathleen** (2020). Incidence, Characteristics, and Outcomes of Interval Breast Cancers Compared With Screening-Detected Breast Cancers JAMA network open, 3(9), e2018179

**Oberaigner, W., Geiger-Gritsch, S., Edlinger, M., Daniaux, M., Knapp, R., Hubalek, M., Siebert, U., Marth, C., Buchberger, W.** (2017). Reduction in advanced breast cancer after introduction of a mammography screening program in Tyrol/Austria Breast, 33(#issue#), 178-182

**Plecha, Donna, Salem, Nelly, Kremer, Mallory, Pham, Ramya, Downs-Holmes, Catherine, Sattar, Abdus, Lyons, Janice** (2014). Neglecting to screen women between 40 and 49 years old with mammography: what is the impact on treatment morbidity and potential risk reduction? AJR. American journal of roentgenology, 202(2), 282-8

**Politi, Julieta, Sala, Maria, Domingo, Laia, Vernet-Tomas, Maria, Roman, Marta, Macia, Francesc, Castells, Xavier** (2020). Readmissions and complications in breast ductal carcinoma in situ: A retrospective study comparing screen- and non-screen-detected patients Women's health (London, England), 16(#issue#), 1745506520965899

**Roder, David, Farshid, Gelareh, Gill, Grantley, Kollias, Jim, Koczwara, Bogda, Karapetis, Chris, Adams, Jacqui, Joshi, Rohit, Keefe, Dorothy, Powell, Kate, Fusco, Kellie, Eckert, Marion, Buckley, Elizabeth, Beckmann, Kerri** (2017). Breast cancer screening-opportunistic use of registry and linked screening data for local evaluation Journal of evaluation in clinical practice, 23(3), 508-516

**Savaridas, S. L., Gierlinski, M., Warwick, V. R., Evans, A. E.** (2022). Opting into breast screening over the age of 70 years: seeking evidence to support informed choice Clinical Radiology, 77(9), 666-672

**Taylor, Richard, Gregory, Marli, Sexton, Kerry, Wharton, Jessica, Sharma, Nisha, Amoyal, Georgina, Morrell, Stephen** (2019). Breast cancer mortality and screening mammography in New Zealand: Incidence-based and aggregate analyses Journal of medical screening, 26(1), 35-43

**Tomsic, S., Zagar, T., Mihor, A., Mlakar, M., Lokar, K., Jarm, K., Zadnik, V.** (2022). Prognostic factors and outcomes in women with breast cancer in Slovenia in relation to step-wise implementation of organized screening PLoS ONE, 17(11 November), e0278384

**Wozniacki, Piotr, Skokowski, Jaroslaw, Bartoszek, Krzystof, Kosowska, Anna, Kalinowski, Leszek, Jaskiewicz, Janusz** (2017). The impact of the Polish mass breast cancer screening program on prognosis in the Pomeranian Province Archives of medical science : AMS, 13(2), 441-447

#### Screening initiation date <2000 (n=9)

**Beau, Anna-Belle, Lynge, Elsebeth, Njor, Sisse Helle, Vejborg, Ilse, Lophaven, Soren Nymand** (2017). Benefit-to-harm ratio of the Danish breast cancer screening programme *International journal of cancer*, 141(3), 512-518

**Lynge, Elsebeth, Beau, Anna-Belle, von Euler-Chelpin, My, Napolitano, George, Njor, Sisse, Olsen, Anne Helene, Schwartz, Walter, Vejborg, Ilse** (2020). Breast cancer mortality and overdiagnosis after implementation of population-based screening in Denmark *Breast cancer research and treatment*, 184(3), 891-899

**Narod, S. A., Sun, P., Wall, C., Baines, C., Miller, A. B.** (2014). Impact of screening mammography on mortality from breast cancer before age 60 in women 40 to 49 years of age *Current oncology (Toronto, Ont.)*, 21(5), 217-21

**Natal, Carmen, Caicoya, Martin, Prieto, Miguel, Tardon, Adonina** (2015). [Breast cancer incidence related with a population-based screening program] *Incidencia de cancer de mama en relacion con la participacion en un programa de cribado poblacional.*, 144(4), 156-60

**Sankatsing, Valerie D. V., van Ravesteyn, Nicolien T., Heijnsdijk, Eveline A. M., Looman, Caspar W. N., van Luijt, Paula A., Fracheboud, Jacques, den Heeten, Gerard J., Broeders, Mireille J. M., de Koning, Harry J.** (2017). The effect of population-based mammography screening in Dutch municipalities on breast cancer mortality: 20 years of follow-up *International journal of cancer*, 141(4), 671-677

**Tabar, Laszlo, Chen, Tony Hsiu-Hsi, Yen, Amy Ming-Fang, Chen, Sam Li-Sheng, Fann, Jean Ching-Yuan, Chiu, Sherry Yueh-Hsia, Ku, May M. S., Wu, Wendy Yi-Ying, Hsu, Chen-Yang, Chen, Yu-Ying, Beckmann, Kerri, Smith, Robert A., Duffy, Stephen W.** (2018). Effect of Mammography Screening on Mortality by Histological Grade *Cancer epidemiology, biomarkers & prevention : a publication of the American Association for Cancer Research, cosponsored by the American Society of Preventive Oncology*, 27(2), 154-157

**van der Waal, Danielle, Broeders, Mireille J. M., Verbeek, Andre L. M., Duffy, Stephen W., Moss, Sue M.** (2015). Case-control Studies on the Effectiveness of Breast Cancer Screening: Insights from the UK Age Trial *Epidemiology (Cambridge, Mass.)*, 26(4), 590-6

**Van Ourti, T., O'Donnell, O., Koc, H., Fracheboud, J., de Koning, H.** (2019). Effect of screening mammography on breast-cancer mortality: Quasi-experimental evidence from rollout of the Dutch population-based program with 17-year follow-up of a cohort *International journal of cancer*, #volume#(#issue#), #Pages#

**Van Ourti, Tom, O'Donnell, Owen, Koc, Hale, Fracheboud, Jacques, de Koning, Harry J.** (2020). Effect of screening mammography on breast cancer mortality: Quasi-experimental evidence from rollout of the Dutch population-based program with 17-year follow-up of a cohort *International journal of cancer*, 146(8), 2201-2208

**Yang, Lei, Wang, Shengfeng, Huang, Yubei** (2020). An exploration for quantification of overdiagnosis and its effect for breast cancer screening *Chinese journal of cancer research = Chung-kuo yen cheng yen chiu*, 32(1), 26-35

##### Previously included in 2018 review (n=4)

**Nystrom, Lennarth, Bjurstam, Nils, Jonsson, Hakan, Zackrisson, Sophia, Frisell, Jan** (2017). Reduced breast cancer mortality after 20+ years of follow-up in the Swedish randomized controlled mammography trials in Malmo, Stockholm, and Goteborg *Journal of medical screening*, 24(1), 34-42

**Bjurstam, Nils G., Bjorneld, Lena M., Duffy, Stephen W.** (2016). Updated results of the Gothenburg Trial of Mammographic Screening *Cancer*, 122(12), 1832-5

**Moss, Sue M., Wale, Christopher, Smith, Robert, Evans, Andrew, Cuckle, Howard, Duffy, Stephen W.** (2015). Effect of mammographic screening from age 40 years on breast cancer mortality in the UK Age trial at 17 years' follow-up: a randomised controlled trial *The Lancet. Oncology*, 16(9), 1123-1132

**Miller, Anthony B., Wall, Claus, Baines, Cornelia J., Sun, Ping, To, Teresa, Narod, Steven A.** (2014). Twenty-five-year follow-up for breast cancer incidence and mortality of the Canadian National Breast Screening Study: randomised screening trial *BMJ (Clinical research ed.)*, 348(#issue#), g366

##### Ineligible intervention (n=3)

**Iwamoto, T., Kumamaru, H., Miyata, H., Tomotaki, A., Niikura, N., Kawai, M., Anan, K., Hayashi, N., Masuda, S., Tsugawa, K., Aogi, K., Ishida, T., Masuoka, H., Iijima, K., Matsuoka, J., Doihara, H., Kinoshita, T., Nakamura, S., Tokuda, Y.** (2016). Distinct breast cancer characteristics between screen- and self-detected breast cancers recorded in the Japanese Breast Cancer Registry *Breast Cancer Research and Treatment*, 156(3), 485-494

**Jaffar Al Bahrani, Bassim, Mehdi, Itrat, Al Lawati, Taha Mohsin, Al Farsi, Abdulaziz M., Al Lawati, Najla A., Al Harthi, Hasina** (2022). Impact of Screening Programs on Stage Migration in Breast Cancer *The Gulf journal of oncology*, 1(38), 38-46

**Mittra, Indraneel, Mishra, Gauravi A., Dikshit, Rajesh P., Gupta, Subhadra, Kulkarni, Vasundhara Y., Shaikh, Heena Kauser A., Shastri, Surendra S., Hawaldar, Rohini, Gupta, Sudeep, Pramesh, C. S., Badwe, Rajendra A.** (2021). Effect of screening by clinical breast examination on breast cancer incidence and mortality after 20 years: prospective, cluster randomised controlled trial in Mumbai *BMJ (Clinical research ed.)*, 372(#issue#), n256

##### No outcomes of interest (n=2)

**Hemminki, K., Forsti, A.** (2022). Incidence, Mortality and Survival Trends in Breast Cancers Coincident with Introduction of Mammography in the Nordic Countries *Cancers*, 14(23), 5907

**Kang, Stella K., Jiang, Miao, Duszak, Richard, Jr., Heller, Samantha L., Hughes, Danny R., Moy, Linda** (2018). Use of Breast Cancer Screening and Its Association with Later Use of Preventive Services among Medicare Beneficiaries *Radiology*, 288(3), 660-668

##### Not English or French (n=2)

**Giudici, Fabiola, Bortul, Marina, Clagnan, Elena, Del Zotto, Stefania, Franzo, Antonella, Giordano, Livia, Gobbato, Michele, Puliti, Donella, Serraino, Diego, Zucchetto, Antonella, Zanier, Loris, Zanconati, Fabrizio, Bucchi, Lauro** (2020). [Early effects of attendance to the Friuli Venezia Giulia (Northern Italy) mammography screening programme on the incidence of advanced-stage breast cancer: a cohort study] *Effetti precoci dell'adesione al programma di screening mammografico della Regione Friuli Venezia Giulia sull'incidenza del cancro della mammella in stadio avanzato: uno studio di coorte.*, 44(2-3), 145-153

**Santos, Renata Oliveira Maciel Dos, Assis, Monica de, Dias, Maria Beatriz Kneipp, Tomazelli, Jeane Glaucia** (2023). [Risk of false-positive result in mammography screening in Brazil] *Risco de resultado falso positivo no rastreamento mamografico do Brasil.*, 39(5), e00117922

#### Not a “very high” HDI country (n=2)

**Charaka, Hafida, Khalis, Mohamed, Elfakir, Samira, Chami Khazraji, Youssef, Zidouh, Ahmed, Abousselham, Loubna, El Rhazi, Karima, Lyoussi, Badiaa, Nejjari, Chakib** (2016). Organization and Evaluation of Performance Indicators of a Breast Cancer Screening Program in Meknes-Tafilalt Region, Morocco *Asian Pacific journal of cancer prevention : APJCP*, 17(12), 5153-5157

**Su, Shih-Yung** (2022). Nationwide mammographic screening and breast cancer mortality in Taiwan: an interrupted time-series analysis *Breast cancer (Tokyo, Japan)*, 29(2), 336-342

#### Appendix 14 – Study characteristics table

| Study | Participants and Study Design | Intervention | Control | Outcomes |
|---|---|---|---|---|
| **RCTs (n=3)** | | | | |
| **Duffy 2020[53],United Kingdom and Duffy 2020[43], United Kingdom**<br><br>**Trial/Screening Program:** UK Age Trial - National Health Service Breast Screening Programme (NHSBSP) | **Participants:** Women aged 39–41 years were recruited from 23 screening units in England, Wales, and Scotland within the NHSBSP<br><br>**Study design:** RCT<br><br>**Follow-up:**<br>*For Duffy 2020[53] and Duffy 2020[43]:*<br>Mean of 23 years; median of 22·8 years (IQR 21·8–24·0) | n=53,914<br><br>Type of mammography: NR<br><br>Number of screening intervals: NR<br><br>The intervention group was invited to an annual breast screen with film mammography, two view at first screen and single view thereafter, up to and including the calendar year of their 48th birthday. All screening in the trial was completed by 2006. | n=107,007<br>The control group received usual care during the intervention periods.<br><br>At the age of 50 years, the control group became eligible for invitation to screening every 3 years as part of the NHSBSP and received their first invitation between age 50 and 52 years**.** | BC mortality, other/all-cause mortality, overdiagnosis |
| **Tarone 1995[36], Canada**<br><br>**Trial/Screening Program:** CNBSS I – Additional analysis for Swedish Two County (Kopparberg & Ostergotland), Malmo I, HIP, Stockholm | **Participants:** Women aged 40-49 years were recruited from 15 urban centres in Canada with expertise in the diagnosis and treatment of Breast Cancer<br><br>**Study design:** RCT<br><br>**Follow-up**:<br>Mean follow-up: 8.5 years | n=44,925 (CNBSS-I)<br><br>Type of mammography: Two-view film screen mammography<br><br>The screening group was invited to annual mammography and physical examination | n=44,910 (CNBSS-I)<br><br>Participants in the control group received a single physical examination at enrollment into the study and usual medical care thereafter. | Stage at Diagnosis (stage II and higher, stage III and higher) |
| **Observational studies (n=26)** | | | | |
| **Choi 2021[15], Korea**<br><br>**Screening Program/Database: Korean National Cancer Screening Program (KNCSP)** | **Participants:** Women born between 1923-1963 (aged 40-79 years) who received an invitation to the KNCSP for breast cancer screening between 2002 and 2003<br><br>**Study design:** Cohort<br><br>**Follow-up:** Maximum 14 years for mortality outcomes (2002 to 2015); Mean 8.42 years screened and 7.52 years in non-screened | n=1,099,417 (screened at first invitation);<br>n=5,026,186 (from non-screened to screened)<br><br>Type of mammography: Women were invited for screen-film mammography, computed radiography, and full-field digital mammography<br><br>Number of screening intervals: Over 70% of the screened women attended screening more than once during follow-up<br>2 rounds: 21.0% | n=2,047,569 (never screened);<br>n=5,153,696 (from non-screened to screened)<br><br>Women who never underwent BC screening during the follow-up period. Additionally, the control included the women-years of women who were eventually screened from the date of initial invitation (January 1, 2002; January 1, 2003) to the date of their first screening attendance | Breast cancer mortality, all-cause mortality, incidence of invasive BC |

| Study | Participants and Study Design | Intervention | Control | Outcomes |
|---|---|---|---|---|
| | | 3 rounds: 21.6%<br>4 rounds: 19.5%<br>5 rounds: 11.0%<br>6+ rounds: 3.0% | | |
| **Duffy 2021[17], Sweden**<br><br>**Screening Program/Database: Swedish Cause of Death Register of the Swedish** National Board of Health and Welfare; Screening data: Sectra Medical Systems, Linköping | **Participants:** Swedish women eligible for screening mammography in nine counties from 1992 to 2016 (aged 40-69 years); 37078/549091 (~7%) participants previously had BC<br><br>**Study design:** Cohort<br><br>**Follow-up:** Mean 22 years, maximum 22 years (mortality outcomes); Mean 13 years, maximum 16 years for fatal BC cases within 10 years after diagnosis | n=392,135 serial participants, defined as women who participated in both of their last two scheduled screening examinations.<br><br>Type of mammography: NR<br><br>Number of screening intervals: 2 | n=84,265 serial non-participants<br><br>Women who did not participate in either of their last two scheduled screening examinations. | BC mortality |
| **Dunn 2021[54], Australia**<br><br>**Screening Program/Database: BreastScreen Australia** | **Participants:** Women (aged 50–65 years) recorded on the Queensland Electoral Roll in the year 2000<br><br>**Study design:** Cohort<br><br>**Follow-up:** Maximum 16 years | n= 187558 screened<br><br>Women with a mammography record up to 31/12/2005. Women with a record of screening prior to 2000 were allocated to the screened group from the commencement of the accrual period.<br><br>Type of mammography: NR<br><br>Number of screening intervals: NR | n= 74792 unscreened<br><br>Women with no mammography record from 2000 up to 31/12/2005. Women with no record of screening prior to the beginning of the accrual period contributed person-years in the non-screened cohort until the date of first attendance for screening during the accrual period. | BC mortality |
| **Morrell 2017[18], New Zealand**<br><br>**Screening Program/Database:** BreastScreen Aotearoa (BSA) programme | **Participants:** A cohort comprising all New Zealand women aged 45–69 years during 1999–2011<br><br>**Study design:** Cohort<br><br>**Follow-up:** Maximum 11 years (2000-2011) | n= 542,234 ever screened<br><br>Person-years of participation in screening are calculated from the time of first screen to the beginning of each successive year, or to the year of diagnosis for women diagnosed with breast cancer.<br><br>Type of mammography: Two-view bilateral mammograms (initially), digital mammogram began in 2006 | n=327, 623 never screened<br><br>Women with no recorded screening participation in a given year, inferred by subtraction from ethnic- and age-specific census derived populations for that year, as provided by Statistics New Zealand | Breast cancer mortality |

| Study | Participants and Study Design | Intervention | Control | Outcomes |
|---|---|---|---|---|
| | | Number of screening intervals: Up to 3 | | |
| **Lund 2018[44], Norway**<br><br>**Screening Program/Database:** The Norwegian Breast Cancer Screening Program (NBCSP) | **Participants:** Women aged 49 to 79 during the first 9 years of national coverage of the NBCSP (2005-2013) were selected for the present analysis. Women living in Norway with no previous cancer diagnosis were included.<br><br>**Study design:** Cohort<br><br>**Follow-up:** Maximum 8 years (2005-2013) | n= 83,963 screened<br><br>Women who have received at a NBCSP mammogram. Additionally, person years from women were categorised as unscreened until their first NBCSP mammogram, at which they were moved to the screened category.<br><br>Type of mammography: Digital mammography<br><br>Number of screening intervals: NR | n=9,974 never screened<br><br>Women who had never taken a mammogram at time of recruitment | Overdiagnosis |
| **Puliti 2017[37], Italy**<br><br>**Screening Program/Database:** Italian National Centre for Screening Monitoring | **Participants:** Nine health care districts in central and northern Italy participated in this study. The cohort included all women 50-69 years old who were invited to the first round of their local mammography screening programme.<br><br>**Study design:** Cohort<br><br>**Follow-up:** Median duration 11 years (IQR: 9-13); Total study period 1991-2009 with follow-up truncated at 13 years | n= 276,322 attenders<br><br>Type of mammography: NR<br><br>Number of screening intervals: At least one round (two rounds of screening invitations) | n=137,125 non-attenders<br><br>Women who did not attend either of the first two screening rounds to which they received an invitation. | Stage at diagnosis |
| **Weedon-Fekjaer 2014[55], Norway**<br><br>**Screening Program/Database:** Norwegian breast cancer screening programme | **Participants:** All Norwegian women aged 50 to 79 years between 1986 and 2009 were eligible for inclusion. Data were obtained from the Norwegian Cancer Registry and women were followed until they were censored (death, reached 80 years or they had reached the end of follow-up).<br><br>**Study design:** Cohort | N= 2,407,709 person-years invited to screening<br><br>Type of mammography: NR<br><br>Number of screening intervals: NR Biennial invitations sent to women aged 50-69 years.<br><br>Two-view screening mammograms are taken in breast diagnostic centres | n=12,785,325 person-years not invited to screening<br><br>Person-time of those not invited to the Norwegian mammography screening programme, 1986-2009. | BC mortality |

| Study | Participants and Study Design | Intervention | Control | Outcomes |
|---|---|---|---|---|
| | **Follow-up:** Maximum follow-up 5-10 years after invitations to screening ended (1986-2009) | exclusively dedicated to the diagnosis and treatment of breast diseases. | | |
| **Coldman 2014[16], Canada**<br><br>**Screening Program/Database: Canadian Breast** Cancer Screening Initiative (CBCSI), including seven provincial screening programs: British Columbia, Manitoba, Ontario, Québec, New Brunswick, Nova Scotia, and Newfoundland and Labrador | **Participants:** Separate cohorts were assembled for each screening program, consisting of women who had at least one program screen between ages 40 and 79 years in the period between January 1, 1990, and December 31, 2009<br><br>**Study design:** Population-based study<br><br>**Follow-up:** Maximum 19 years (1990-2009) | Screening Cohort<br>Province:<br>British Columbia n=787815<br>Manitoba n= 132306<br>Ontario n= 797648<br>Québec n= 758912<br>New Brunswick n= 127039<br>Nova Scotia n= 162379<br>Newfoundland and Labrador n= 30373<br><br>Type of mammography: NR<br><br>Number of screening intervals: NR | Referent rates were derived from nonparticipants in each province defined to be those not in, or before entry to, the participant cohort, n=NR | Standardized mortality ratios (SMRs) by age at entry, including 40 to 49 years |
| **Richman 2023[45], United States**<br><br>**Screening Program/Database:** SEER (Surveillance, Epidemiology, and End Results)-Medicare registry | **Participants:** Study included women aged 70 years and older by January 1, 2003, had no breast cancer detected before 2002 screening mammogram and had Medicare fee-for-service insurance through 2005.<br><br>**Study design:** Cohort<br><br>**Follow-up:** Median 13.7 years (IQR, 9.2 to 14.4 years) among women aged 70 to 74 years, 10 years (IQR, 5.8 to 13.9 years) for women aged 75 to 84 years, 5.7 years (IQR, 3.1 to 9.1 years) for women 85 years and older | n=44,485<br><br>Type of mammography: Study adapted a validated algorithm that distinguishes screening mammograms from diagnostic mammograms based on Current Procedural Terminology (CPT) and Healthcare Common Procedure Coding System (HCPCS) codes.<br><br>Number of screening intervals: NR | n=10,150<br><br>Women who did not have a screening mammogram in three years after their 2002 mammogram were included in the non-screening group. | Overdiagnosis |
| **Garcia-Albeniz 2020[19], United States**<br><br>**Screening program/Database:** Medicare Data | **Participants:** Women aged 70-84 years who had a life expectancy of at least 10 years, had no previous breast cancer diagnosis, and underwent screening mammography. Women were US Medicare enrollees between 1999 and 2008 who have screening mammography the day they enter the trial. | N= 264,274<br><br>"Continue screening" strategy (Women continue annual screening mammography (with a 3-month grace period and are excused from further screening after a breast cancer diagnosis) | n= 264,274<br><br>"Stop screening" strategy (women do not have further screening after baseline) | BC mortality, treatment-related morbidity, and overdiagnosis |

| Study | Participants and Study Design | Intervention | Control | Outcomes |
|---|---|---|---|---|
| | **Study design:** Population-based cohort study<br><br>**Follow-up:** 8 years | | | |
| **Wilkinson 2023[30], Canada**<br><br>**Screening program/Database:** Canadian Community Health Survey (CCHS), Canadian Cancer Registry and population data from Statistics Canada | **Participants:** Women aged 40-49 years diagnosed with Breast Cancer between 2002 and 2007<br><br>**Study design:** Population-based study<br><br>**Follow-up:** 10 years | n=5,680 (breast cancer cases)<br><br>Number of screening intervals: NR<br><br>Five jurisdictions with organized screening programs, including self-referral and annual recall, were designated as screeners: British Columbia, Alberta, Nova Scotia, Prince Edward Island, and the Northwest Territories. | n=15,408 (breast cancer cases)<br><br>Jurisdictions with no organized screening programs and only limited opportunistic screening (Newfoundland and Labrador, New Brunswick, Quebec, Ontario, Manitoba, Saskatchewan, and Yukon Territory) | Incidence-based BC mortality |
| **Wilkinson 2022[40], Canada**<br><br>**Screening Program/Database:** Canadian Community Health Survey (CCHS), Canadian Cancer Registry and population data from Statistics Canada | **Participants:** Women aged 40-49 and 50-59 years diagnosed with invasive breast cancer between 2010 and 2017<br><br>**Study design:** Population-based study<br><br>**Follow-up:** unclear | n= 20,965 (breast cancer cases)<br><br>Number of screening intervals: NR<br><br>Five jurisdictions with organized screening programs, including self-referral and annual recall, were designated as screeners: British Columbia, Alberta, Nova Scotia, Prince Edward Island, and the Northwest Territories. | n= 34,525 (breast cancer cases)<br><br>Jurisdictions with no organized screening programs and only limited opportunistic screening (Newfoundland and Labrador, New Brunswick, Quebec, Ontario, Manitoba, Saskatchewan, and Yukon Territory) | Stage at diagnosis |
| **Blyuss 2023[56], United Kingdom**<br><br>**Screening Program/Database: UK National** Health Service Breast Screening Program | **Participants:** The population were those invited to BC screening. Cases were women who were diagnosed with primary breast cancer (invasive or in situ) in 2010 or 2011 and aged between 47 and 89 years. Controls were women sampled from the general population of those invited for screening and alive at the time of their corresponding case's date of diagnosis, matched on date of birth (within 1 month) and screening area at date of their case's diagnosis.<br><br>**Study design:** Case-control | n= 144,699 ever screened (93,020 controls and 51,679 cases)<br><br>Type of mammography: NR<br><br>Number of screening intervals: Mean number of screening rounds, excluding those never screened (range): cases 3.4 (1.0–13.0); controls 3.3 (1.0–12.0) | n=18,447 never screened (12,633 controls and 5,814 cases) | Overdiagnosis |

| Study | Participants and Study Design | Intervention | Control | Outcomes |
|---|---|---|---|---|
| | **Follow-up:** Maximum 2<br>3 years | | | |
| **De Troeyer 2023[26], Belgium**<br><br>**Screening Program/Database:** Flemish population-based mammography screening program (PMSP) | **Participants:** The study population was all women aged 50-69 years and eligible for the Flemish PMSP between 2005-2012. Cases were women from the study population who died from BC between 2005-2017 and had the opportunity to be screened within the program at least once prior to diagnosis. For each case, four referents were randomly selected among the women of the study population who were still alive at the time of death of the corresponding case and who had the opportunity to be screened prior to the date of diagnosis of the case ('pseudodiagnosis' date).<br><br>**Study design:** Case-control<br><br>**Follow-up:** Outcomes assessed between 2005-2017; screening exposure period is within 4 years prior to BC diagnosis for cases/pseudodiagnosis for controls | N= 4217 screened (620 cases and 3597 referents)<br><br>Type of mammography: Digital mammography introduced in 2005.<br><br>Number of screening intervals: At least one | n= 3638 not screened (2687 referents and 951 cases) | BC mortality |
| **Maroni 2021[25], United Kingdom**<br><br>**Screening Program/Database: UK National** Health Service Breast Screening Program | **Participants:** England-wide study of women who died of primary BC or matched controls invited to screening; women aged 47–89 years.<br><br>**Study design:** Case-control<br><br>**Follow-up:** Maximum 21 years (1990-2011) | n= 19946 (6547 cases and 13399 controls) screened<br><br>Type of mammography: NR<br><br>Number of screening intervals: 62.3% controls had 2+ rounds; 53.5% cases had 2+ rounds | n= 3544 never screened (1803 controls and 1741 cases) | BC mortality |
| **van der Waal 2017[24], Netherlands** | **Participants:** Case subjects were women who died of BC in Nijmegen between 1975 and 2008. Each case subject was | n= 1460 screened (220 cases and 1240 controls) | n= 538 not screened (425 controls and 113 cases) | BC mortality |

| Study | Participants and Study Design | Intervention | Control | Outcomes |
|---|---|---|---|---|
| **Screening Program/Database:** Nijmegen (Dutch) screening program | matched to five control subjects, alive at the time of death of the matched case. All subjects had been invited to screening in the index round. The proportion of women with dense breasts among the interval cases ranged from 38.7% to 54.5%, and these proportions were always greater than for the screen-detected cases (ranged from 20.7% to 30.5%).<br><br>**Study design:** Case-control<br><br>**Follow-up:** Maximum 33 years (1975 – 2008) | Screening exposure was defined as attending the index screening round and/or the screening examination preceding the index round (pre-index round). This reflects the screening participation within the 4 years before (pseudo-)diagnosis.<br><br>Type of mammography: Only screen-film mammograms<br><br>Number of screening intervals: 1-2 | | |
| **Massat 2016[22], United Kingdom**<br><br>**Screening Program/Database: UK National Health Service Breast Screening Program** | **Participants:** Women residing in the London region, who had been invited to participate in the NHS BSP from 1988 onward. Cases were women who died of primary BC aged 47 to 89 years between 2008-2009, and who were diagnosed with primary BC (invasive) between the ages of 47 to 89 years after 1990. Each case was matched based on age and geographical area to 1-2 general population controls.<br><br>**Study design:** Case-control<br><br>**Follow-up:** Maximum 11 years (1998-2009; over 80% of women in our dataset selected were diagnosed from the year 2000 onward) | n=1991 screened at least once and invited at least once (649 cases and 1342 controls)<br><br>n= 1286 screened >1 and invited at least twice (406 cases and 880 controls)<br><br>Type of mammography: NR<br><br>Number of screening intervals: Median (range) 2.0 (0-7) controls, 1.0 (0-8) cases | n=520 never screened and invited at least once (220 cases and 300 controls)<br><br>n=299 never screened and invited at least twice (121 cases and 178 controls) | BC mortality |
| **Beckmann 2015[57], Australia**<br><br>**Screening Program/Database:** BreastScreen South Australia (BSSA) | **Participants:** Cases were selected from the South Australian Cancer Registry (SACR) and consisted of all women diagnosed with breast cancer, between 2006 and 2010, who were aged | n= 17664 (3370 cases and 14294 controls) screened<br><br>Type of mammography: Analogue screen-film technology | n= 7709 no screening records (1,213 cases and 6,496 controls) | Overdiagnosis |

| Study | Participants and Study Design | Intervention | Control | Outcomes |
|---|---|---|---|---|
| | between 45 and 85 years and resident in SA at the time of diagnosis. Five age-matched controls per case with the same month and year of birth were randomly sampled from the South Australian electoral rolls (ER) using an incidence density sampling approach.<br><br>**Study design:** Case-control<br><br>**Follow-up:** Maximum 21 years (1989-2010) | Number of screening intervals: At least 1 BreastScreen SA; Mean rounds of screening excluding never screened: 5.1 (1–18) cases; 4.9 (1–16) controls | | |
| **Pocobelli 2015[21], Canada**<br><br>**Screening Program/Database:** Screening Program for Breast Cancer (SPBC) | **Participants:** Cases were women who died of BC at 50–79 years of age during 1990–2008 and who had continuous Saskatchewan healthcare coverage for at least 5 years prior to their first primary BC diagnosis (index date). For each case, 15 potential controls were randomly sampled, with the same birth year and the same duration of continuous health coverage.<br><br>**Study design:** Case-control<br><br>**Follow-up:** Maximum 13 years (1995–2008) | n=2833 (186 cases + 2647 controls) screened within the two years prior to and including the index date.<br><br>SPBC began in selected regions of Saskatchewan. Women 50–69 years of age are identified from the population registry and are mailed a letter of invitation to receive a screening mammogram (women >70 years of age may attend, but they are not mailed a letter of invitation).<br><br>Type of mammography: NR<br><br>Number of screening intervals: NR | n=2677 not screened (315 cases and 2362 controls)<br><br>No screening within the two years prior to and including the index date. | BC mortality |
| **Paap 2014[20], Netherlands**<br><br>**Screening Program/Database: Dutch breast cancer screening program** | **Participants:** All women aged 50-75 years who received at least one invitation to the service screening program in the five participating screening regions. Cases originated from the source population and were defined as women who died from breast cancer in 2004 or 2005. Cases were individually matched to referents from the population invited to screening.<br><br>**Study design:** Case-control | n= NR (overall 80.8% participation rate) screened<br><br>Participated in the screening examination following their index invitation and/or the invitation in the screening round before the index invitation<br><br>Type of mammography: NR<br><br>Number of screening intervals: Maximum 2 | n=NR (overall 19.2 non-participation rate) non-screened<br><br>No participation in either the screening examination following their index invitation or the invitation in the screening round before the index invitation | BC mortality |

| Study | Participants and Study Design | Intervention | Control | Outcomes |
|---|---|---|---|---|
| | **Follow-up:** Maximum 15 years (1990-2005) | | | |
| **Ripping 2016[23], Netherlands**<br><br>**Screening Program/Database:** Nijmegen (Dutch) screening program | **Participants:** The case-control study was conducted within the population invited to the biennial mammographic screening program in Nijmegen, the Netherlands.<br>Cases were defined as women who were aged 50 to 75 years at diagnosis and died from BC before 2013. For each case, five controls were matched based on age and invitation date to screening program.<br><br>**Study design:** Case-control<br><br>**Follow-up:** Maximum 38 years (1975-2013); analytic screening period 4 years before case diagnosis | N= 10,264 screened (1,541 cases and 8,723 controls)<br><br>Type of mammography: Film before 2007, digital after 2007<br><br>Number of screening intervals: NR – Analytic period for screening was 4 years before the diagnosis of the case, covering 2 consecutive screening invitations. | n=3,328 unscreened (724 cases and 2,604 controls) | BC mortality |
| **Katalinic 2020[58], Germany**<br><br>**Screening Program/Database:** German National mammography BC screening program | **Participants:** Study used anonymized individual data of female patients with first (incident) registered ductal carcinoma in situ (DCIS, ICD10: D05.1, representing about 95% of all in situ BCs) or invasive BC diagnosis (ICD-10:C50)<br><br>**Study design:** Time-trend analysis<br><br>**Follow-up:** NA | Active-screening period (2015-2016); n=NR<br><br>Type of mammography: NR<br><br>Number of screening intervals: NR<br><br>The German screening program follows the European guidelines on BC screening and includes an invitation every two years. | Pre-screening period (non-screened; 2003-2004); n=NR | BC mortality (age-specific rate/100,000) by age group) |
| **de Glas 2014[38], Netherlands**<br><br>**Screening Program/Database:** National Cancer Registry Netherlands | **Participants:** From the Netherlands cancer registry, we selected all patients aged 70-75 with a diagnosis of invasive and ductal carcinoma in situ BC between 1995 and 2011<br><br>**Study design:** Time-trend analysis | Active screening (2003-2011) time period; n= 3,394,055 person-years<br><br>Type of mammography: NR<br><br>Number of screening intervals: NR<br><br>Study assessed the incidence of early stage and advanced stage breast | Pre-screening (1995-1997) time period; n=1,115,508 person-years | Stage at diagnosis for 70–75-year age group |

| Study | Participants and Study Design | Intervention | Control | Outcomes |
|---|---|---|---|---|
| | **Follow-up:** NA | cancer before and after implementation of the mass screening programme in women aged 70-75 years in the Netherlands. | | |
| **Parvinen 2015[28], Finland**<br><br>**Screening Program/Database:** Finish Cancer Registry | **Participants:** Since 1992, all Finnish women aged 50–59 years receive screening invitations every second year. The screening in Turku targeted female inhabitants aged 40–74 years.<br><br>**Study design:** Time-trend analysis<br><br>**Follow-up:** NA | Active-screening period (1987-2009); n= 468,195 person-years (1987-1997) and 541,096 person-years (1997-2009)<br><br>Type of mammography: NR<br><br>Number of screening intervals: NR<br><br>The aim of this study was to evaluate the effectiveness of a large-scale screening programme for breast cancer (BC) in Turku, Finland. In Turku, women aged 40–49 years were invited at modified invitation intervals from 1987 to 2009. Women born in even years were invited annually, and those born in odd years, triennially. | Pre-screening time period (1976-1986); n= 430,462 person-years | BC mortality (for screening ages of interest 40-49 years) |
| **Helvie 2014[39], United States**<br><br>**Screening Program/Database:** Surveillance, Epidemiology and End Results (SEER) database | **Participants:** Women aged 40 or greater in the U.S. that participated in the National Cancer Institute's SEER program<br><br>**Study design:** Time-trend analysis<br><br>**Follow-up:** NA | Active screening period (2007-2009); n=NR<br><br>Type of mammography: NR<br><br>Number of screening intervals: NR | Pre-screening time period (1977 to 1979); n=NR | Stage specific breast cancer (early stage, late stage) |
| **Tabar 2019[29], Sweden**<br><br>**Screening program/Database: the National Death Registry of the Swedish** National Board of Health and Welfare | **Participants:** All women aged 40 to 69 years in the county of Dalarna, Sweden, during 39 years of the screening era (1977-2015).<br><br>**Study design:** Time-trend analysis<br><br>**Follow-up:** 10 years (for those diagnosed with BC through 2005) and 20 years (those diagnosed with BC through 1995) | n=NR<br><br>Type of mammography: NR<br><br>Number of screening intervals: NR<br><br>In the service screening program, women aged 40 to 54 years are invited to mammography screening every 18 months, and those aged 55 to 69 years are invited to mammography screening every 24 months. | n=NR (Unscreened and Pre-screening control group)<br><br>Unscreened population included women who did not participate in mammography screening.<br><br>Pre-screening control means doing historical comparisons of the rate of death from breast cancer among women before | Breast cancer mortality |

| Study | Participants and Study Design | Intervention | Control | Outcomes |
|---|---|---|---|---|
| | | The screening protocol is 2-view mammography. | the onset of the screening programs (1958-1976) | |

#### Appendix 15 – Summary characteristics of RCTs (taken from previous 2017 review)

| | Malmo I | Malmo II | Stockholm | Gothenburg | CNBSS 1 | CNBSS2 | AGE | HIP | Swedish Two County (Ostergotland) | Swedish Two County (Kopparberg) |
|---|---|---|---|---|---|---|---|---|---|---|
| **Year of study** | 1976 | 1978 | 1981 | 1982 | 1980 | 1980 | 1991 | 1963 | 1977 | 1978 |
| **Country (Rural/Urban)** | Sweden (Urban) | Sweden (Urban) | Sweden (Urban) | Sweden (Urban) | Canada (Urban) | Canada (Urban) | UK (Urban) | USA (Urban) | Sweden (Urban) | Sweden (Urban) |
| **Study Design** | RCT | RCT | Quasi-RCT | Quasi-RCT | RCT | RCT | RCT | RCT | Cluster-RCT | Cluster-RCT |
| **Age at Entry** | 45-70 | 43-49 | 39-65 | 39-59 | 40-49 | 50-59 | 39-41 | 40-64 | 40-74 | 40-74 |
| **Total Randomized (n)** | N=42,283 | N=17,793 | N=60,117 | N=50,200 | N=50,489 | N=39,459 | N=160,921 | N=61,004^{B} | N=75,894 | N=57,171 |
| **Ethnicity** | NR | NR | NR | NR | NR | NR | NR | NR | NR | NR |
| **SES** | NR | NR | NR | NR | Level of Education, Occupation | Level of Education, Occupation | NR | NR | NR | NR |
| **% Breast density** | NR | NR | NR | NR | NR | NR | NR | NR | NR | NR |
| **Longest follow-up reported** | 30 yrs (mean)* | 22 yrs (mean)* | 25 yrs (mean)* | 24 yrs (mean)* | 21.9 yrs (mean)* | 21.9 yrs (mean)* | 17.7 yrs (median)* | 18 yrs (mean) | 25.7 yrs (mean) | 25.7 yrs (mean) |
| **Intervention type** | M (Film) | M (Film) | M (Film) | M (Film) | M (Film) + | M (Film) + CBE | M (NR) | M (Film) + CBE | M (NR) | M (NR) |
| **Intervention n randomized** | (n=21,088) | (n=9,581) | (n=39,139) | (n=21,000) | (n=25,246) | (n=19,735) | (n=53,914) | (n=30,239) | (n=38,491) | (n=38,589) |
| **Comparator type** | UC | UC | UC | UC | UC | CBE | UC | UC | UC | UC |
| **Comparator n randomized** | (n=21,195) | (n=8,212) | (n=20,978) | (n=29,200) | (n=25,243) | (n=19,724) | (n=107,007) | (n=30,765) | (n=37,403) | (n=18,582) |
| **Received screening at end of study period?** | No | Yes | Yes | Yes | No | No | Yes | NR | Yes | Yes |
| **# of views** | 2 | 2 | 1 | 2 | 2 | 2 | 2 | 2 | 1 | 1 |
| **# of readers** | 2 | 2 | 1 | 1 | NR | NR | NR | NR | 1 | 1 |
| **Screening interval** | 18-24 mo. | 18-24 mo. | 28 mo. | 18 mo. | 12 mo. | 12 mo. | 12 mo. | 12 mo. | 24-33 mo. | 24-33 mo. |
| **Duration of screening** | 12 yrs | 12 yrs | 4 yrs | 7 yrs | 4 yrs | 4 yrs | 8 yrs | 3 yrs | 7 yrs | 7 yrs |
| **Attendance rate** | 74% | 74% | 82% | 84% | 88% | 88% | 81% | 65% | 85% | 85% |

#### Appendix 16 – Risk of bias summary tables

**Table 1**. Risk of bias ratings for observational studies using JBI critical appraisal tools

| JBI critical appraisal tool for cohort studies | | | | | | | | | | | | |
|---|---|---|---|---|---|---|---|---|---|---|---|---|
| **Author, Year** | Were the two groups similar and recruited from the same population? | Were the exposures measured similarly to assign people to both exposed and unexposed | Was the exposure measured in a valid and reliable way? | Were confounding factors identified? | Were strategies to deal with confounding factors stated? | Were the groups/participants free of the outcome at the start of the study (or at the moment | Were the outcomes measured in a valid and reliable way? | Was the follow up time reported and sufficient to be long enough for outcomes to occur? | Was follow up complete, and if not, were the reasons to loss to follow up described and | Were strategies to address incomplete follow up utilized? | Was appropriate statistical analysis used? | Overall Assessment of Risk of Bias |
| **Choi, 2021** | Green | Green | Red | Red | Green | Green | Green | Green | Green | Green | Green | Green |
| **Duffy, 2020** | Green | Green | Green | Red | Green | Red | Yellow | Green | Red | Green | Green | Yellow |
| **Dunn, 2021** | Green | Green | Yellow | Red | Green | Green | Green | Green | Green | Green | Green | Green |
| **Morrell, 2017** | Red | Red | Green | Red | Green | Yellow | Yellow | Yellow | Red | Green | Green | Red |
| **Lund, 2018** | Green | Green | Green | Red | Green | Yellow | Green | Red | Red | Green | Red | Red |
| **Puliti, 2018** | Green | Green | Green | Red | Green | Yellow | Yellow | Green | Red | Green | Green | Yellow |
| **Weedon-Fekjaer, 2014** | Green | Green | Green | Red | Green | Yellow | Green | Red | Red | Green | Red | Red |
| **Coldman, 2014** | Green | Green | Green | Green | Green | Yellow | Green | Green | Red | Green | Green | Green |
| **Richman, 2023** | Green | Green | Green | Green | Green | Green | Green | Green | Red | Green | Green | Green |
| **Garcia-Albeniz, 2020** | Green | Green | Green | Green | Green | Green | Green | Green | Red | Red | Green | Green |
| **JBI critical appraisal tool for case-control studies** | | | | | | | | | | | | |

| | Were the groups comparable other than the presence of disease in cases or the [illegible] | Were cases and controls matched appropriately? | Were the same criteria used for identification of cases and controls? | Was exposure measured in a standard, valid and reliable way? | Was exposure measured in the same way for cases and controls? | Were confounding factors identified? | Were strategies to deal with confounding factors stated? | Were outcomes assessed in a standard, valid and reliable way for cases and controls? | Was the exposure period of interest long enough to be meaningful? | Was appropriate statistical analysis used? | | Overall Assessment of Risk of Bias |
|---|---|---|---|---|---|---|---|---|---|---|---|---|
| **Blyuss, 2023** | Green | Green | Green | Green | Green | Green | Green | Green | Yellow | Green | | Green |
| **De Troeyer, 2023** | Green | Red | Green | Red | Green | Red | Green | Green | Yellow | Green | | Yellow |
| **Maroni, 2021** | Green | Green | Green | Green | Green | Red | Green | Green | Yellow | Green | | Yellow |
| **van der Waal, 2017** | Green | Red | Green | Green | Green | Red | Red | Green | Yellow | Green | | Yellow |
| **Massat, 2016** | Green | Green | Green | Red | Green | Red | Green | Green | Yellow | Green | | Yellow |
| **Beckmann, 2015** | Green | Green | Green | Red | Green | Red | Green | Green | Yellow | Green | | Yellow |
| **Pocobelli, 2015** | Green | Green | Green | Red | Green | Green | Green | Green | Yellow | Red | | Yellow |
| **Paap, 2014** | Green | Green | Green | Red | Green | Red | Green | Green | Yellow | Green | | Yellow |
| **Ripping, 2016** | Green | Green | Green | Red | Green | Red | Red | Green | Yellow | Red | | Red |

**JBI critical appraisal tool for Quasi Experimental studies**

| | Is it clear in the study what is the cause ‘ and what is the ‘ effect’ (i.e. there is no | Were the participants included in any comparisons similar? | Were the participants included in any comparisons receiving similar treatment/care, other | Was there a control group? | Were there multiple measurements of the outcome both pre and post the | Was follow up complete and if not, were differences between groups in terms of their follow | Were the outcomes of participants included in any comparisons measured in the | Were outcomes measured in a reliable way? | Was appropriate statistical analysis used? | | | Overall Assessment of Risk of Bias |
|---|---|---|---|---|---|---|---|---|---|---|---|---|
| **Katalinic, 2020** | Green | Green | Green | Green | Green | Yellow | Green | Yellow | Green | | | Yellow |
| **de Glas, 2014** | Green | Green | Green | Green | Green | Yellow | Green | Yellow | Green | | | Yellow |
| **Parvinen, 2015** | Green | Red | Green | Yellow | Green | Yellow | Green | Yellow | Green | | | Red |
| **Helvie, 2014** | Green | Green | Green | Yellow | Green | Yellow | Green | Yellow | Green | | | Yellow |
| **Tabar, 2019** | Green | Green | Green | Yellow | Green | Yellow | Green | Yellow | Green | | | Yellow |
| **Wilkinson, 2022** | Green | Green | Red | Yellow | Grey | Grey | Green | Yellow | Green | | | Yellow |
| **Wilkinson, 2023** | Green | Red | Red | Yellow | Grey | Grey | Green | Yellow | Green | | | Yellow |

Colour coding: Green: Yes/Low; Yellow: Unclear/Moderate; Red: No/High. For the overall score, observational studies were colour-coded as “high”, “moderate” or “low” based on tallied scores of 75-100%, 50-75% of items, or below 50% of checklist items sufficiently met (Yes).

**Table 2.** Risk of bias ratings for Stage at Diagnosis Outcomes (Randomized Controlled Trials)

| Study | Randomization | Allocation Concealment | Blinding (patients/ personnel) | Blinding (outcomes) | Incomplete Outcome Assessments | Selective Outcome Reporting | Other | Overall Judgement |
|---|---|---|---|---|---|---|---|---|
| **Swedish Two County (Kopparberg & Ostergotland) (Tabar 1995)** | yes | no | unclear | yes | *unclear* | *unclear* | *unclear* | high |
| **Malmo I (Nystrom 2016)** | unclear | unclear | unclear | yes | *unclear* | *unclear* | yes | *moderate* |
| **CNBSS 1 (Miller 2014)** | no | unclear | unclear | yes | *unclear* | *unclear* | yes | high |
| **HIP (Shapiro 1988)** | unclear | unclear | unclear | no | *unclear* | *unclear* | unclear | high |
| **Stockholm (Nystrom 2016)** | no | no | unclear | yes | *unclear* | *unclear* | yes | high |

Assessments performed using the Cochrane RoB-2 tool. The study is judged to be at overall high risk of bias if at least one domain had this result, moderate risk of bias if there were some concerns observed, and low risk of bias if all domains had this rating.

**Table 3.** Risk of bias ratings for Treatment-related Morbidity Outcomes (Randomized Controlled Trials)

| Study | Randomization | Allocation Concealment | Blinding (patients/ personnel) | Blinding (outcomes) | Incomplete Outcome Assessments | Selective Outcome Reporting | Other | Overall Judgement |
|---|---|---|---|---|---|---|---|---|
| **CNBSS 1 (Miller 2014)** | no | unclear | unclear | yes | *unclear* | *unclear* | yes | high |
| **CNBSS 2 (Miller 2014)** | no | unclear | unclear | yes | *unclear* | *unclear* | yes | high |

| | | | | | | | | |
|---|---|---|---|---|---|---|---|---|
| **Malmo I (Nystrom 2016)** | unclear | unclear | unclear | yes | *unclear* | *unclear* | yes | *moderate* |
| **Swedish Two County (Kopparberg) (Tabar 1995)** | yes | no | unclear | yes | unclear | *unclear* | *unclear* | high |
| **Stockholm (Nystrom 2016)** | no | no | unclear | yes | *unclear* | *unclear* | yes | high |

Assessments performed using the Cochrane RoB-2 tool. The study is judged to be at overall high risk of bias if at least one domain had this result, moderate risk of bias if there were some concerns observed, and low risk of bias if all domains had this rating.

#### Appendix 17 – Breast cancer mortality findings summary for exploratory analyses

**Moderately increased risk (for family history)**

The following data are extrapolations. We did not extract this data, however, used data from previously reported literature[2] and extrapolated.

a) **40 to 49 years**

From RCT data, we extrapolated 0.44 fewer breast cancer deaths per 1,000 and from observational studies values ranged from 1.28 to 1.51 fewer deaths per 1,000, depending on the study design.

b) **50 to 59 years**

From RCT data, we extrapolated 0.79 fewer breast cancer deaths per 1,000 and from observational studies values ranged from 2.33 to 2.76 fewer deaths per 1,000.

c) **60 to 69 years**

From RCT data, we extrapolated 1.04 fewer breast cancer deaths per 1,000 and from observational studies values ranged from 3.04 to 3.59 fewer deaths per 1,000.

d) **70 to 74 years**

From RCT data, we extrapolated 1.47 fewer breast cancer deaths per 1,000 and from observational studies values ranged from 4.31 to 5.10 fewer deaths per 1,000.

**Moderately increased risk (for dense breasts)**

The following data are extrapolations. We did not extract this data, however used data from previously reported literature[3] and extrapolated.

a) **40 to 49 years**

From RCT data, we extrapolated 0.53 fewer breast cancer deaths per 1,000 and from observational studies, values ranged from 1.54 to 1.82 fewer deaths per 1,000, depending on the study design.

b) **50 to 59 years**

From RCT data, we extrapolated 0.95 fewer breast cancer deaths per 1,000 and from observational studies values ranged from 2.77 to 3.28 fewer deaths per 1,000.

c) **60 to 69 years**

From RCT data, we extrapolated 1.23 fewer breast cancer deaths per 1,000 and from observational studies values ranged from 3.61 to 4.26 fewer deaths per 1,000.

d) **70 to 74 years**

From RCT data we extrapolated 1.74 fewer breast cancer deaths per 1,000 and from observational studies values ranged from 5.10 to 6.03 fewer deaths per 1,000.

#### Appendix 18 - Sensitivity analysis by RoB - Breast cancer mortality RCTs

1. **New analysis:** Breast cancer mortality (RCTs, short-case accrual, stratified by age)
   Rated as moderate: Malmo I & II, AGE

| Study or Subgroup | log[] | SE | Weight | IV, Random, 95% CI |
|---|---|---|---|---|
| **1.23.1 Moderate risk of bias (40-49 years)** | | | | |
| Moss 2015- UK Age | -0.12783337 | 0.08681802 | 80.5% | 0.88 [0.74, 1.04] |
| Nystrom 2002- Malmo I | -0.30110509 | 0.28626091 | 7.4% | 0.74 [0.42, 1.30] |
| Nystrom 2016- Malmo II | -0.16251893 | 0.22429226 | 12.1% | 0.85 [0.55, 1.32] |
| **Subtotal (95% CI)** | | | **100.0%** | **0.87 [0.74, 1.01]** |
| Heterogeneity: Tau² = 0.00; Chi² = 0.34, df = 2 (P = 0.84); I² = 0% | | | | |
| Test for overall effect: Z = 1.86 (P = 0.06) | | | | |
| **1.23.2 High risk of bias (40-49 years)** | | | | |
| Miller 2014- CNBSS-1 | 0.0861777 | 0.15865298 | 26.1% | 1.09 [0.80, 1.49] |
| Nystrom 2016- Gothenburg | -0.52763274 | 0.21995498 | 20.1% | 0.59 [0.38, 0.91] |
| Nystrom 2016- Stockholm | 0.40546511 | 0.35026764 | 11.5% | 1.50 [0.75, 2.98] |
| Shapiro 1988- HIP | -0.32850407 | 0.19684153 | 22.2% | 0.72 [0.49, 1.06] |
| Tabar 2011- Two-County | -0.05129329 | 0.21848782 | 20.2% | 0.95 [0.62, 1.46] |
| **Subtotal (95% CI)** | | | **100.0%** | **0.89 [0.67, 1.17]** |
| Heterogeneity: Tau² = 0.05; Chi² = 8.59, df = 4 (P = 0.07); I² = 53% | | | | |
| Test for overall effect: Z = 0.84 (P = 0.40) | | | | |
| **1.23.3 Moderate risk of bias (50-59 years)** | | | | |
| Nystrom 2016- Malmo I | 0 | 0.16611811 | 100.0% | 1.00 [0.72, 1.38] |
| **Subtotal (95% CI)** | | | **100.0%** | **1.00 [0.72, 1.38]** |
| Heterogeneity: Not applicable | | | | |
| Test for overall effect: Z = 0.00 (P = 1.00) | | | | |
| **1.23.4 High risk of bias (50-59 years)** | | | | |
| Miller 2014- CNBSS-2 | 0.01980263 | 0.14511466 | 28.1% | 1.02 [0.77, 1.36] |
| Nystrom 2016- Gothenburg | -0.11653382 | 0.19724232 | 18.6% | 0.89 [0.60, 1.31] |
| Nystrom 2016- Stockholm | -0.49429632 | 0.26816583 | 11.4% | 0.61 [0.36, 1.03] |
| Shapiro 1988- HIP | -0.22314355 | 0.18356644 | 20.6% | 0.80 [0.56, 1.15] |
| Tabar 2011- Two-County | -0.4462871 | 0.1796421 | 21.3% | 0.64 [0.45, 0.91] |
| **Subtotal (95% CI)** | | | **100.0%** | **0.81 [0.67, 0.98]** |
| Heterogeneity: Tau² = 0.01; Chi² = 5.57, df = 4 (P = 0.23); I² = 28% | | | | |
| Test for overall effect: Z = 2.16 (P = 0.03) | | | | |
| **1.23.5 Moderate risk of bias (60-69 years)** | | | | |
| Nystrom 2016- Malmo I | -0.31471074 | 0.25594441 | 100.0% | 0.73 [0.44, 1.21] |
| **Subtotal (95% CI)** | | | **100.0%** | **0.73 [0.44, 1.21]** |
| Heterogeneity: Not applicable | | | | |
| Test for overall effect: Z = 1.23 (P = 0.22) | | | | |
| **1.23.6 High risk of bias (60-69 years)** | | | | |
| Nystrom 2002- Stockholm | -0.0618754 | 0.37745569 | 15.8% | 0.94 [0.45, 1.97] |
| Shapiro 1988- HIP | -0.03045921 | 0.32593381 | 20.5% | 0.97 [0.51, 1.84] |
| Tabar 2011- Two-County | -0.49429632 | 0.1530757 | 63.7% | 0.61 [0.45, 0.82] |
| **Subtotal (95% CI)** | | | **100.0%** | **0.72 [0.53, 0.98]** |
| Heterogeneity: Tau² = 0.02; Chi² = 2.41, df = 2 (P = 0.30); I² = 17% | | | | |
| Test for overall effect: Z = 2.09 (P = 0.04) | | | | |
| **1.23.7 Moderate risk of bias (70-74 years)** | | | | |
| Nystrom 2002- Malmo I | -0.02020271 | 0.9653545 | 100.0% | 0.98 [0.15, 6.50] |
| **Subtotal (95% CI)** | | | **100.0%** | **0.98 [0.15, 6.50]** |
| Heterogeneity: Not applicable | | | | |
| Test for overall effect: Z = 0.02 (P = 0.98) | | | | |
| **1.23.8 High risk of bias (70-74 years)** | | | | |
| Tabar 2011- Two-County | -0.11653382 | 0.22740374 | 100.0% | 0.89 [0.57, 1.39] |
| **Subtotal (95% CI)** | | | **100.0%** | **0.89 [0.57, 1.39]** |
| Heterogeneity: Not applicable | | | | |
| Test for overall effect: Z = 0.51 (P = 0.61) | | | | |

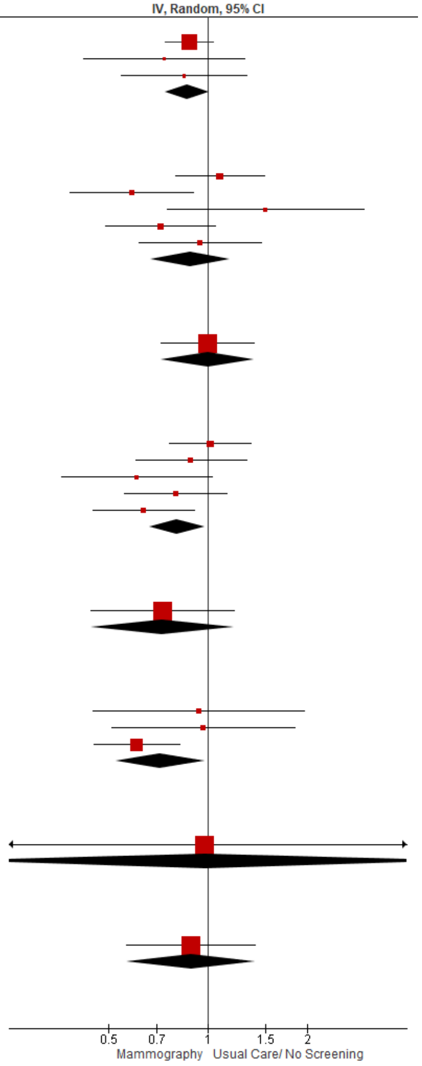

2. **Original analysis 2018 systematic review:** Breast cancer mortality (RCTs, short-case accrual, stratified by age)

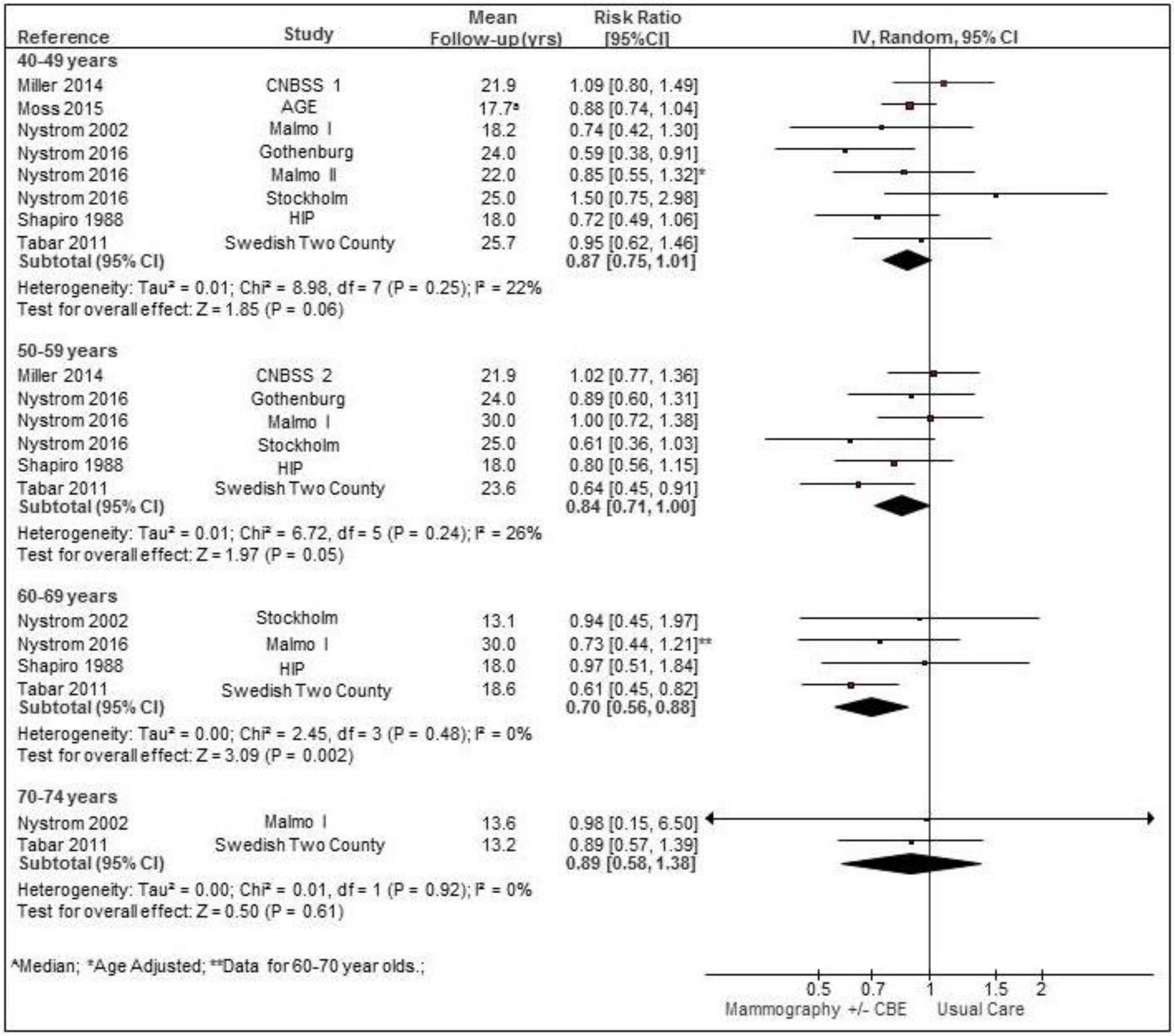

3. **New analysis:** Breast cancer mortality sensitivity analysis (RCTs, short-case accrual, all ages)

| Study or Subgroup | log[Risk Ratio] | SE | Weight | Risk Ratio IV, Random, 95% CI |
|---|---|---|---|---|
| **1.25.1 Moderate risk of bias** | | | | |
| Moss 2015- UK Age | -0.12783337 | 0.08681781 | 58.2% | 0.88 [0.74, 1.04] |
| Nystrom 2016- Malmo I | -0.12783337 | 0.11530233 | 33.0% | 0.88 [0.70, 1.10] |
| Nystrom 2016- Malmo II | -0.16251893 | 0.22412 | 8.7% | 0.85 [0.55, 1.32] |
| **Subtotal (95% CI)** | | | **100.0%** | **0.88 [0.77, 1.00]** |
| Heterogeneity: Tau² = 0.00; Chi² = 0.02, df = 2 (P = 0.99); I² = 0% | | | | |
| Test for overall effect: Z = 1.98 (P = 0.05) | | | | |
| **1.25.2 High risk of bias** | | | | |
| Miller 2014- CNBSS 1 & 2 | 0.04879016 | 0.10838857 | 23.4% | 1.05 [0.85, 1.30] |
| Nystrom 2016- Gothenburg | -0.30110509 | 0.14275913 | 17.5% | 0.74 [0.56, 0.98] |
| Nystrom 2016- Stockholm | -0.0618754 | 0.1729285 | 13.7% | 0.94 [0.67, 1.32] |
| Shapiro 1988- HIP | -0.23572233 | 0.11938405 | 21.3% | 0.79 [0.63, 1.00] |
| Tabar 2011- Two-County | -0.31471074 | 0.10487217 | 24.1% | 0.73 [0.59, 0.90] |
| **Subtotal (95% CI)** | | | **100.0%** | **0.84 [0.72, 0.98]** |
| Heterogeneity: Tau² = 0.01; Chi² = 7.50, df = 4 (P = 0.11); I² = 47% | | | | |
| Test for overall effect: Z = 2.28 (P = 0.02) | | | | |

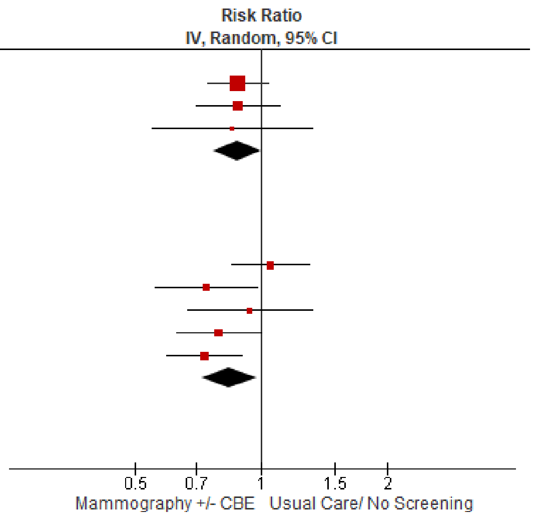

Test for subgroup differences: Chi² = 0.20, df = 1 (P = 0.66), I² = 0%

4. **Original analysis 2018 systematic review:** Breast cancer mortality (RCTs, short-case accrual, all ages)

| Reference | Study | Age (at entry) | Mean Follow-up (yrs) | log [RR] | SE | Weight | Risk Ratio [95%CI] |
|---|---|---|---|---|---|---|---|
| Miller 2014 | CNBSS 1 & 2 | 40-69 | 21.9 | 0.05 | 0.11 | 15.3% | 1.05 [0.85, 1.30] |
| Moss 2015 | AGE | 39-41 | 17.7[a] | -0.13 | 0.09 | 22.2% | 0.88 [0.74, 1.04] |
| Nystrom 2016 | Gothenburg | 40-59 | 24.0 | -0.30 | 0.14 | 9.3% | 0.74 [0.56, 0.98] |
| Nystrom 2016 | Malmo I | 45-70 | 30.0 | -0.13 | 0.12 | 13.7% | 0.88 [0.70, 1.10] |
| Nystrom 2016 | Malmo II | 43-49 | 22.0 | -0.16 | 0.22 | 4.0% | 0.85 [0.55, 1.32]* |
| Nystrom 2016 | Stockholm | 40-65 | 25.0 | -0.06 | 0.17 | 6.5% | 0.94 [0.67, 1.32] |
| Shapiro 1988 | HIP | 40-64 | 18.0 | -0.24 | 0.12 | 12.9% | 0.79 [0.63, 1.00] |
| Tabar 2011 | Swedish Two County | 40-74 | 29.0[b] | -0.31 | 0.10 | 16.2% | 0.73 [0.59, 0.90]** |
| **Total (95% CI)** | | | | | | **100.0%** | **0.85 [0.78, 0.93]** |

Heterogeneity: Tau² = 0.00; Chi² = 7.80, df = 7 (P = 0.35); I² = 10%
Test for overall effect: Z = 3.49 (P = 0.0005)

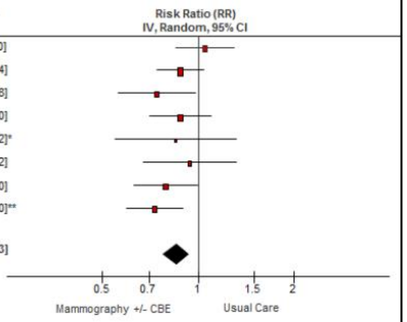

[a]Median; [b]Time since randomization; *Adjusted for Age; ** Adjusted for clustering

5. **New analysis:** All-cause mortality (RCTs, stratified by age)

| Study or Subgroup | log[Risk Ratio] | SE | Weight | Risk Ratio IV, Random, 95% CI |
|---|---|---|---|---|
| **1.24.1 Moderate risk of bias (40-49 years)** | | | | |
| Moss 2015- UK Age | -0.02020271 | 0.02605344 | 89.5% | 0.98 [0.93, 1.03] |
| Nystrom 2002- Malmo II | 0.0295588 | 0.07623862 | 10.5% | 1.03 [0.89, 1.20] |
| **Subtotal (95% CI)** | | | **100.0%** | **0.99 [0.94, 1.03]** |
| Heterogeneity: Tau² = 0.00; Chi² = 0.38, df = 1 (P = 0.54); I² = 0% | | | | |
| Test for overall effect: Z = 0.61 (P = 0.54) | | | | |
| **1.24.2 High risk of bias (40-49 years)** | | | | |
| Bjurstam 1997- Gothenburg | -0.02020271 | 0.06738561 | 36.9% | 0.98 [0.86, 1.12] |
| Frisell 1997- Stockholm | 0.11332869 | 0.37690402 | 1.4% | 1.12 [0.54, 2.34] |
| Miller 2002- CNBSS-1 | 0 | 0.07117994 | 33.7% | 1.00 [0.87, 1.15] |
| Tabar 1989- Kopparberg | 0.28517894 | 0.1481964 | 9.0% | 1.33 [0.99, 1.78] |
| Tabar 1989- Ostergotland | -0.07257069 | 0.09891978 | 19.0% | 0.93 [0.77, 1.13] |
| **Subtotal (95% CI)** | | | **100.0%** | **1.01 [0.92, 1.10]** |
| Heterogeneity: Tau² = 0.00; Chi² = 4.42, df = 4 (P = 0.35); I² = 9% | | | | |
| Test for overall effect: Z = 0.13 (P = 0.90) | | | | |
| **1.24.4 High risk of bias (50-50 years)** | | | | |
| Miller 2000- CNBSS-2 | 0.05826891 | 0.05263685 | 46.2% | 1.06 [0.96, 1.18] |
| Tabar 1989- Kopparberg | 0 | 0.0785272 | 20.7% | 1.00 [0.86, 1.17] |
| Tabar 1989- Ostergotland | -0.02020271 | 0.06214849 | 33.1% | 0.98 [0.87, 1.11] |
| **Subtotal (95% CI)** | | | **100.0%** | **1.02 [0.95, 1.09]** |
| Heterogeneity: Tau² = 0.00; Chi² = 1.01, df = 2 (P = 0.60); I² = 0% | | | | |
| Test for overall effect: Z = 0.57 (P = 0.57) | | | | |
| **1.24.6 High risk of bias (60-69 years)** | | | | |
| Tabar 1989- Kopparberg | -0.05129329 | 0.04553132 | 39.1% | 0.95 [0.87, 1.04] |
| Tabar 1989- Ostergotland | -0.02020271 | 0.03650532 | 60.9% | 0.98 [0.91, 1.05] |
| **Subtotal (95% CI)** | | | **100.0%** | **0.97 [0.92, 1.02]** |
| Heterogeneity: Tau² = 0.00; Chi² = 0.28, df = 1 (P = 0.59); I² = 0% | | | | |
| Test for overall effect: Z = 1.14 (P = 0.26) | | | | |
| **1.24.8 High risk of bias (70-74 years)** | | | | |
| Tabar 1989- Kopparberg | 0.04879016 | 0.04873858 | 46.9% | 1.05 [0.95, 1.16] |
| Tabar 1989- Ostergotland | -0.07257069 | 0.03806439 | 53.1% | 0.93 [0.86, 1.00] |
| **Subtotal (95% CI)** | | | **100.0%** | **0.98 [0.87, 1.11]** |
| Heterogeneity: Tau² = 0.01; Chi² = 3.85, df = 1 (P = 0.05); I² = 74% | | | | |
| Test for overall effect: Z = 0.26 (P = 0.80) | | | | |

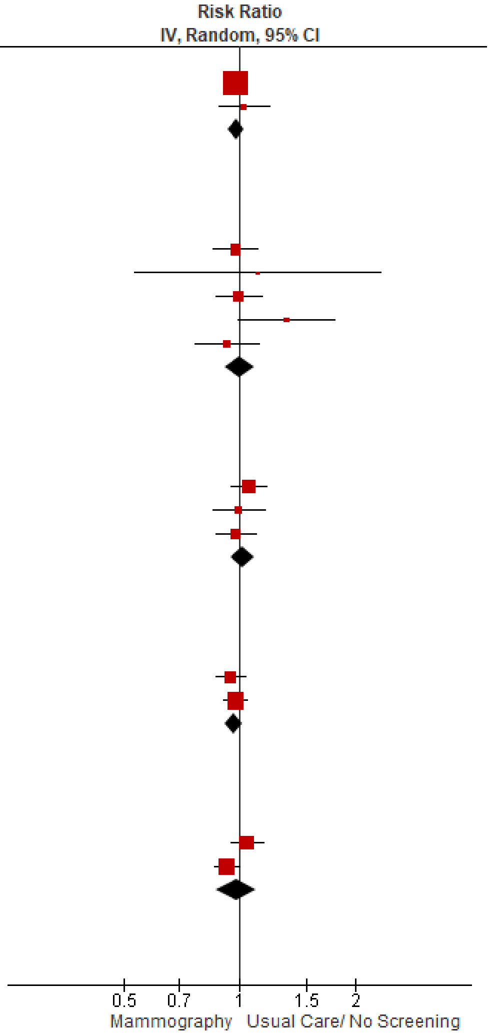

Test for subgroup differences: Chi² = 1.49, df = 4 (P = 0.83), I² = 0%

6. **Original analysis 2018 systematic review:** All-cause mortality (RCTs, stratified by age)

| Study or Subgroup | log[Risk Ratio] | SE | Weight | Risk Ratio IV, Random, 95% CI |
|---|---|---|---|---|
| **1.6.1 40-49 years** | | | | |
| Bjurstam 1997- Gothenburg | -0.02020271 | 0.06738561 | 9.9% | 0.98 [0.86, 1.12] |
| Frisell 1997- Stockholm | 0.11332869 | 0.37690402 | 0.3% | 1.12 [0.54, 2.34] |
| Miller 2002- CNBSS-1 | 0 | 0.07117994 | 8.9% | 1.00 [0.87, 1.15] |
| Moss 2015- UK Age | -0.02020271 | 0.02605344 | 66.4% | 0.98 [0.93, 1.03] |
| Nystrom 2002- Malmo II | 0.0295588 | 0.07623862 | 7.8% | 1.03 [0.89, 1.20] |
| Tabar 1989- Kopparberg | 0.28517894 | 0.1481964 | 2.1% | 1.33 [0.99, 1.78] |
| Tabar 1989- Ostergotland | -0.07257069 | 0.09891978 | 4.6% | 0.93 [0.77, 1.13] |
| **Subtotal (95% CI)** | | | **100.0%** | **0.99 [0.95, 1.03]** |
| Heterogeneity: Tau² = 0.00; Chi² = 4.94, df = 6 (P = 0.55); I² = 0% | | | | |
| Test for overall effect: Z = 0.48 (P = 0.63) | | | | |
| **1.6.2 50-59 years** | | | | |
| Miller 2000- CNBSS-2 | 0.05826891 | 0.05263685 | 46.2% | 1.06 [0.96, 1.18] |
| Tabar 1989- Kopparberg | 0 | 0.0785272 | 20.7% | 1.00 [0.86, 1.17] |
| Tabar 1989- Ostergotland | -0.02020271 | 0.06214849 | 33.1% | 0.98 [0.87, 1.11] |
| **Subtotal (95% CI)** | | | **100.0%** | **1.02 [0.95, 1.09]** |
| Heterogeneity: Tau² = 0.00; Chi² = 1.01, df = 2 (P = 0.60); I² = 0% | | | | |
| Test for overall effect: Z = 0.57 (P = 0.57) | | | | |
| **1.6.3 60-69 years** | | | | |
| Tabar 1989- Kopparberg | -0.05129329 | 0.04553132 | 39.1% | 0.95 [0.87, 1.04] |
| Tabar 1989- Ostergotland | -0.02020271 | 0.03650532 | 60.9% | 0.98 [0.91, 1.05] |
| **Subtotal (95% CI)** | | | **100.0%** | **0.97 [0.92, 1.02]** |
| Heterogeneity: Tau² = 0.00; Chi² = 0.28, df = 1 (P = 0.59); I² = 0% | | | | |
| Test for overall effect: Z = 1.14 (P = 0.26) | | | | |
| **1.6.4 70-74 years** | | | | |
| Tabar 1989- Kopparberg | 0.04879016 | 0.04873858 | 46.9% | 1.05 [0.95, 1.16] |
| Tabar 1989- Ostergotland | -0.07257069 | 0.03806439 | 53.1% | 0.93 [0.86, 1.00] |
| **Subtotal (95% CI)** | | | **100.0%** | **0.98 [0.87, 1.11]** |
| Heterogeneity: Tau² = 0.01; Chi² = 3.85, df = 1 (P = 0.05); I² = 74% | | | | |
| Test for overall effect: Z = 0.26 (P = 0.80) | | | | |

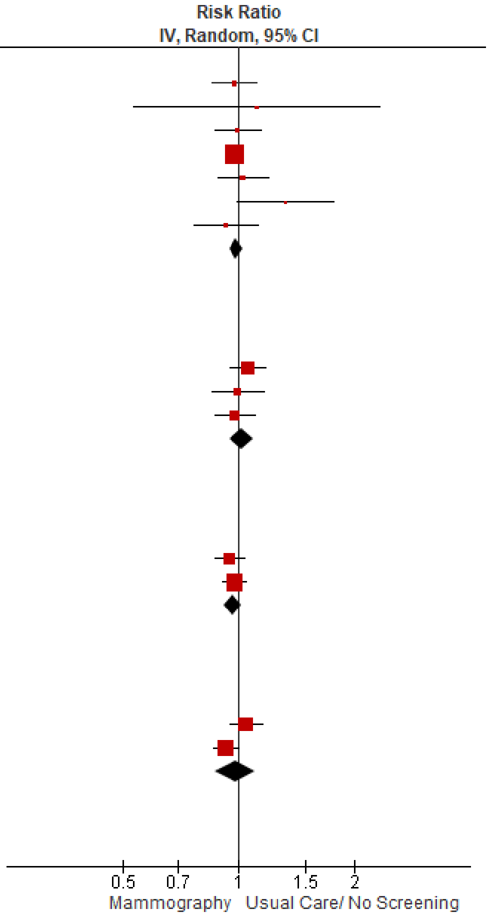

Test for subgroup differences: Chi² = 1.33, df = 3 (P = 0.72), I² = 0%

#### Appendix 19 – Sensitivity analysis: Overdiagnosis RCTs removing high risk of bias trials

##### Mammography +/- CBE compared to Usual Care

| Outcomes | Absolute effects | | Relative effect (95% CI) | № of participants (studies) | Quality of the evidence (GRADE) | Comments |
|---|---|---|---|---|---|---|
| | **Incident rates with usual care (Assumed rate) ‡** | **Absolute risk (95% CI)** | | | | |
| Main analysis: Overdiagnosis invasive + in situ cancers (40-49 years)<br><br>Range of follow-up (yrs): 9 to 15 years | 17.4 per 1,000 | **1.57 more per 1,000** (from 0.35 more to 2.78 more) | **RR 1.09** (1.02 to 1.16) | 293,152 (2 [1,2] RCTs) | ⊕⊕◯◯ LOW [a,b,c,d,e] | Using a threshold of 5, screening may lead to little to no difference in overdiagnosed cancers in individuals aged 40 to 49. |
| Other analysis: Overdiagnosis invasive cancers only (40-49 years)<br><br>Range of follow-up (yrs): 9 to 15 years | 16.2 per 1,000 | **0.49 more per 1,000** (from 0.65 fewer to 1.78 more) | **RR 1.03** (0.96 to 1.11) | 293,152 (2 [1,2] RCTs) | ⊕⊕◯◯ LOW [a,b,c,d,e] | Using a threshold of 5, screening may lead to little to no difference in overdiagnosed invasive cancers in individuals aged 40 to 49 years |
| Main analysis: Overdiagnosis invasive + in situ cancers (50-59 years)<br><br>Range of follow-up (yrs): 10 to 15 years | 32.9 per 1,000 | **3.95 more per 1,000** (from 0.66 more to 7.9 more) | **RR 1.12** (1.02 to 1.24) | 132,231 (1[1] RCTs) | ⊕⊕◯◯ LOW [a,c,d,e,f] | Using a threshold of 5, screening may lead to little to no difference in overdiagnosed cancers in individuals aged 50 to 59 years. |
| Other analysis: Overdiagnosis invasive cancers only (50-59 years)<br><br>Range of follow-up (yrs): 10 to 15 years | 31.2 per 1,000 | **2.81 more per 1,000** (from 0.62 fewer to 6.55 more) | **RR 1.09** (0.98 to 1.21) | 132,231 (1[1] RCTs) | ⊕⊕◯◯ LOW [a,c,d,e,f] | Using a threshold of 5, screening may lead to little to no difference in overdiagnosed invasive cancers in individuals aged 50 to 59 years. |

‡The assumed rate was calculated using the control event rates across included studies.
**CI:** Confidence interval

**Bibliography:** 1: Malmo I (Zackrisson 2006[41]); 2: AGE (Duffy 2020[43])

**Mammography +/- CBE compared to Usual Care**

| Outcomes | Absolute effects | | Relative effect (95% CI) | № of participants (studies) | Quality of the evidence (GRADE) | Comments |
|---|---|---|---|---|---|---|
| | Incident rates with usual care (Assumed rate) ‡ | Absolute risk (95% CI) | | | | |

**GRADE Working Group grades of evidence**
**High quality:** We are very confident that the true effect lies close to that of the estimate of the effect
**Moderate quality:** We are moderately confident in the effect estimate: The true effect is likely to be close to the estimate of the effect, but there is a possibility that it is substantially different
**Low quality:** Our confidence in the effect estimate is limited: The true effect may be substantially different from the estimate of the effect
**Very low quality:** We have very little confidence in the effect estimate: The true effect is likely to be substantially different from the estimate of effect

#### Explanations

a. We downrated once for risk of bias. Randomisation and allocation concealment were not reported or unclear.
b. Not downrated for inconsistency; all point estimates in our pooled analysis lie to one side of our threshold.
c. Downrated once for indirectness. Data are from trials initiated in the 1960s-1990s and some trial estimates included participants outside the previously defined age decades (e.g., in the 40-49 age decade, one study included some individuals in their 50s).
d. Not rated down for imprecision. Clinical decision threshold set at 5.
e. According to Egger et al.[11], 10 trials are needed to asses publication bias. We cannot assess publication bias due to insufficient number of trials, therefore, we did not rate down for publication bias.
f. Only one trial evaluated, we did not downrate for inconsistency.

#### Forest plots

##### Age 40-49 (invasive + in situ)

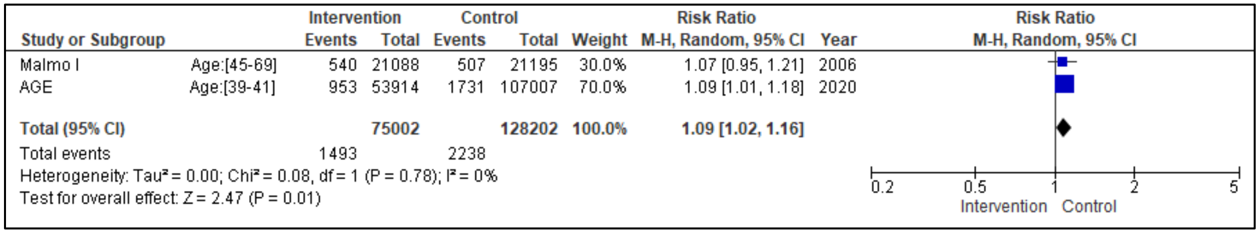

##### Age 40-49 (invasive only)

| Study or Subgroup | | Intervention Events | Intervention Total | Control Events | Control Total | Weight | Risk Ratio M-H, Random, 95% CI | Year |
|---|---|---|---|---|---|---|---|---|
| Malmo I | Age:[45-54] | 481 | 21088 | 454 | 21195 | 29.9% | 1.06 [0.94, 1.21] | 2006 |
| AGE | Age:[39-41] | 835 | 53914 | 1628 | 107007 | 70.1% | 1.02 [0.94, 1.11] | 2020 |
| **Total (95% CI)** | | | **75002** | | **128202** | **100.0%** | **1.03 [0.96, 1.11]** | |
| Total events | | 1316 | | 2082 | | | | |

Heterogeneity: Tau² = 0.00; Chi² = 0.34, df = 1 (P = 0.56); I² = 0%

Test for overall effect: Z = 0.88 (P = 0.38)

Risk Ratio
M-H, Random, 95% CI

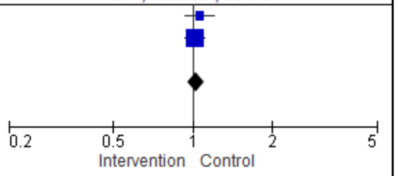

#### Appendix 20 – Supplementary data on additional imaging (no cancers)

##### Part A: Additional imaging (no cancer) resolved by imaging or biopsy per 1,000 screens from provincial sources

1. **Canada (excluding YT and NU), aged 50-69 by year (2004-2016), initial vs subsequent screen**

| Year | Per 1,000 screens | |
|---|---|---|
| | Initial screen | Subsequent screen |
| 2004 | 120.5 | 60.5 |
| 2005 | 120.6 | 58.4 |
| 2006 | 120.2 | 56.4 |
| 2007 | 102.7 | 56.5 |
| 2008 | 111.5 | 58.3 |
| 2009 | 121.6 | 58.4 |
| 2010 | 131.4 | 60.3 |
| 2011 | 142 | 67.7 |
| 2012 | 150.4 | 70.3 |
| 2013 | 159.9 | 72 |
| 2014 | 160.6 | 71.7 |
| 2015 | 161.9 | 73.8 |
| 2016 | 166.6 | 75.2 |

Source: {FP=number of abnormal calls that were not confirmed as cancers per 1000 screens} from the CPAC 2020 report
Includes all provinces and territories, except YT and NU. Data prior to 2007 excludes AB.
Abnormal call rate: Data after 2014 do not include NS.
Invasive cancer detection rate: Data from 2013 to 2016 do not include NS and NB. MB data in 2016 might be underestimated.

2. **Quebec, aged 50-69 by year (2011-2021), initial vs subsequent screen**

| Year | Per 1,000 screens | | |
|---|---|---|---|
| | Initial screen | Subsequent screen | Total |
| 2011 | 176.2 | 80.6 | 97.5 |
| 2012 | 197 | 91.5 | 110.3 |
| 2013 | 204.2 | 92.8 | 112.7 |
| 2014 | 204.3 | 90.8 | 109.6 |
| 2015 | 205.5 | 87.7 | 106.5 |
| 2016 | 220 | 88.7 | 109.5 |
| 2017 | 211.4 | 82.7 | 101.4 |
| 2018 | 216.7 | 81.9 | 100.6 |
| 2019 | 219.3 | 81.9 | 101.5 |
| 2020 | 210 | 83.7 | 97.4 |
| 2021 | 227.5 | 86.8 | 107.5 |
| **5 years average** | 217.4 | 83.8 | 102.5 |
| **10 years average** | 211.1 | 86.9 | 105.6 |

Source: Programme Québécois de Dépistage du Cancer du Sein, released October 2023, available from :
https://www.inspq.qc.ca/sites/default/files/documents/pqdcs/tableaubordpqdcs.pdf

Notes: Number of abnormal calls that were not confirmed as cancers per 1000 screens calculated using total reference rate minus total cancer detection rate

3. **British Columbia, by age group (40-49, 50-59, 50-69, 70-79) during 2015-2019**

| Additional imaging (no cancer) rates | Per 1,000 screens | | | |
|---|---|---|---|---|
| Age group | 40-49 yrs | 50-59 yrs | 60-69 yrs | 70-79 yrs |
| **British Columbia (2015-2019)** {calculated based on Specificity = (1 - false positive rate)} | 123 | 84 | 71 | 65 |

Source : BC Cancer Breast Screening 2019 Program Results, September 2020, available from: http://www.bccancer.bc.ca/screening/Documents/Breast-Screening-Program-Report-2019.pdf (Table 13)

4. **BC, ON, QC (different age groups) screened in 2017**

| Calculated additional imaging (no cancer) rates rate per 1000 screens (2017) | |
|---|---|
| **British Columbia** | **84 (40-74 yrs)** |
| **Ontario** | **83 (50-74 yrs)** |
| **Quebec** | **101.4 (50-69 yrs)** |

Sources:

- BC Source : BC Cancer Breast Screening 2019 Program Results, September 2020, available from: http://www.bccancer.bc.ca/screening/Documents/Breast-Screening-Program-Report-2019.pdf (Table 12)
- BC additional imaging (no cancer) rate (2017) = 1- specificity = 1- 91.6% = 8.4%= 84 per 1000
- Quebec Source: Source: Programme Québécois de Dépistage du Cancer du Sein, released October 2023, available from : https://www.inspq.qc.ca/sites/default/files/documents/pqdcs/tableaubordpqdcs.pdf
- Quebec additional imaging (no cancer) rate per 1000 (Total) (2017)= reference rate - cancer diagnosis rate = 108 – 6.6 = 101.4 per 1000
- Ontario Source: The Ontario Cancer Screening Performance Report 2020, available from: https://www.cancercareontario.ca/sites/ccocancercare/files/assets/OntarioCancerScreeningReport2020.pdf (Tables 15-17)
- Ontario additional imaging (no cancer) rate (2017) = FP/(FP+TN) {reconstructed a 2x2 table}= (53,535 /643,564 ) x 100 = 8.3% = 83 per 1000

5. **Nova Scotia (different age groups) followed from 2000-2011**

| Additional imaging (no cancer) rates per 1000 women screened | 40-49 yrs | 50-69 yrs | 70-74 yrs |
|---|---|---|---|
| **Nova Scotia (2000-2011)** (based on the Nova Scotia breast screening program) {as reported} | **210** | **143** | **76** |

Source: Nova Scotia Annual 2017 Report, available from: https://breastscreening.nshealth.ca/sites/default/files/sites/default/files/annual_report2017.pdf (Table 11.3)
*Note*: This data is not directly comparable to the CPAC data as it is measured per 1000 women over 11 years

#### Part B: Additional imaging and biopsy (no cancer), per 1,000 screens

1. **British Columbia, aged 40-74, by biopsy type in 2019**

| **FPs per 1000 screens** | | Age group (40-74 years) |
|---|---|---|
| British Columbia (2019) | Needle Core | 7.3 |
| | Surgical | 1.0 |
| | Biopsy (core or surgical) | 8.3 |

- Source: BC Cancer Breast Screening 2019 Program Results, September 2020, available from: http://www.bccancer.bc.ca/screening/Documents/Breast-Screening-Program-Report-2019.pdf (Figure 11)
- Calculations:
    - Total additional imaging and biopsy (no cancer) rate for core/FNA biopsies = 1944/266405 = 0.73% or 7.3/1000
    - Total additional imaging and biopsy (no cancer) rate for open (surgical) biopsies = 269/266405 = 0.10% or 1.0/1000
    - Total additional imaging and biopsy (no cancer) rate for biopsies = 2213/266405 = 0.83% or 8.3/1000

2. **British Columbia, aged 50-69, by biopsy type and initial vs rescreen, in 2020**

| **Age 50-69** | **Biopsy type** | **Screen type** | **Per 1000 screens (2020)** |
|---|---|---|---|
| British Columbia (2020) | Core | Initial screen | 27.6 |
| | | Re-screen | 5.4 |
| | Open (surgical) | Initial screen | 3.0 |
| | | Re-screen | 0.8 |

- Source: BC Cancer Breast Screening 2019 Program Results, September 2020, available from: http://www.bccancer.bc.ca/screening/Documents/Breast-Screening-Program-Report-2019.pdf (Table 16)

3. **Quebec, aged 50-69, resolved by open surgical biopsy, by initial vs rescreen and year (2011-2021)**

| **Year** | **Per 1,000 screens** | | |
|---|---|---|---|
| | **Initial screen** | **Subsequent screen** | **Total** |
| 2011 | 2.2 | 1 | 1.3 |
| 2012 | 2.7 | 1.1 | 1.4 |
| 2013 | 2.3 | 1.1 | 1.3 |
| 2014 | 2.3 | 1.1 | 1.3 |
| 2015 | 2.7 | 1.2 | 1.4 |
| 2016 | 2.7 | 1 | 1.3 |
| 2017 | 2.2 | 1 | 1.2 |
| 2018 | 2.9 | 0.9 | 1.2 |
| 2019 | 2.2 | 0.9 | 1 |
| 2020 | 2.3 | 1 | 1.1 |
| 2021 | 2.6 | 0.8 | 1 |
| **5 years average** | 2.4 | 0.9 | 1.1 |
| **10 years average** | 2.5 | 1 | 1.2 |

- Quebec Source: Source: Programme Québécois de Dépistage du Cancer du Sein, released October 2023, available from : https://www.inspq.qc.ca/sites/default/files/documents/pqdcs/tableaubordpqdcs.pdf

4. **Nova Scotia, aged 50-69, resolved by core biopsy, by initial vs rescreen and year (2013-2016)**

| Age 50-69 | Screen type | Per 1,000 women screened (by year) | | | |
|---|---|---|---|---|---|
| | | 2013 | 2014 | 2015 | 2016 |
| Nova Scotia | Initial screen | 49.8 | 31 | 29.5 | 39.4 |
| | Re-screen | 10.2 | 10.2 | 9.5 | 9.7 |

Source: Nova Scotia Annual 2017 Report, available from: https://breastscreening.nshealth.ca/sites/default/files/sites/default/files/annual_report2017.pdf (Table 10.2)

#### Part C: Additional imaging with or without biopsy (No Cancer) rate using 2019-2020 CPAC data, ages 40-49

| | BC 2019 | BC 2020 | PE 2019 |
|---|---|---|---|
| Rate per 1000 screens[1] | 129.4 | 127.5 | 108.4 |
| Rate per 1000 screens estimated over 10 years (four rounds of screening)[2] | 517.6 | 510 | 433.6 |

*Data source: Provincial Breast Cancer Screening Program*

1. Additional imaging rate per screening round calculated by subtracting the cancer detection rates from the abnormal call rate. Abnormal calls were among individuals aged 40-49, who were screened within screening mammography program, by jurisdiction. The screening cancer detection rate (per 1,000) was used among individuals aged 40-49, who were screened within screening mammography program.
2. Additional imaging rate estimated using four rounds of screening over a 10-year period, assuming that the majority of women would receive a screen every 2-3 years.

#### Part D: Additional imaging with or without biopsy (No Cancer) using 2019-2020 CPAC data, ages 50-74

| | All ages | 50-59 | 60-69 | 70-74 |
|---|---|---|---|---|
| Rate per 1000 screens[1] | 77.1 | 90.6 | 67.6 | 63.3 |
| Rate per 1000 screens estimated over 10 years (four rounds of screening)[2] | 308.4 | 362.4 | 270.4 | 253.2 |

*Data source: Provincial Breast Cancer Screening Program*

1. Abnormal calls were among individuals aged 50-74, who were screened within screening mammography program. Data include BC, AB, ON, NB, PE and NL. In PE, if a woman was screened a second time within 9 months of the previous screen, these screens were dropped because they were likely follow-ups to the first screens. Cancer detection rate (per 1,000) among individuals aged 50-74, who were screened within screening mammography program. Jurisdictions combined: Data include BC, AB, ON, NB, PE and NL. In NB and NL, screens that were lost to follow-up within 6 months of screening were not excluded.
2. Additional imaging rate estimated using four rounds of screening over a 10-year period, assuming that the majority of women would receive a screen every 2-3 years.

#### Part E: Additional imaging example calculations

Women aged 50-59 years using 2011-2012 CPAC data

1. **Additional imaging with or without biopsy (no cancer):**

Abnormal call rate per 1000 screens

- Initial = 156 per 1000
- Subsequent = 75 per 1000

Cancer detection rate (CDR) per 1000 screens

- Initial = invasive CDR + in situ CDR = 4.1 per 1000 + 1.2 per 1000 = 5.3 per 1000
- Subsequent = invasive CDR + in situ CDR = 2.7 per 1000 + 0.7 per 1000 = 3.4 per 1000

Calculated additional imaging rates (no cancer):

- Initial = 156 – 5.3 = 150.7 per 1000
- Subsequent = 75 – 3.4 = 71.6 per 1000

Rates over 10 years:

- Assuming started screening in this age decade (initial screen and three subsequent): 150.7 + (71.6*3) = 365.5 per 1000 in a 10-year period
- Assuming started screening in 40s (four subsequent screens): 71.6*4 = 286.4 per 1000 in a 10-year period

2. **Additional imaging and biopsy (no cancer):**

Non-malignant biopsy rate, initial screen (per 1,000 screens) = 20.7

Non-malignant biopsy rate, subsequent screen (per 1,000 screens) = 8.5

Rates over 10 years:

- Assuming started screening in this age decade (initial screen and three subsequent): 20.7 + (8.5*3) = 46.2 per 1000 in a 10-year period
- Assuming started screening in 40s (four subsequent screens): 8.5*4 = 34 per 1000 in a 10-year period

3. **Additional imaging no biopsy (no cancer):**

Additional imaging no biopsy (no cancer) = recall rate – (cancer detection rate + non-malignant biopsy rate)

Initial Screening:

- Additional imaging no biopsy (no cancer) = 156 per 1000 – (5.3 per 100 + 20.7 per 1000) = 130 per 1000

Subsequent Screening:

- Additional imaging no biopsy (no cancer) = 75 per 1000 – (3.4 per 100 + 8.5 per 1000) = 63.1 per 1000

Rates over 10 years:

- Assuming started screening in this age decade (initial screen and three subsequent): 130 + (63.1*3) = 319.3 per 1000
- Assuming started screening in 40s (four subsequent screens): 63.1*4 = 252.4 per 1000

#### Appendix 21 – Sensitivity analysis: Observational studies

The following sensitivity analysis includes our adherence to screen analysis for breast cancer mortality. There were notable differences between studies, and we teased out how removing certain studies impacted the overall estimate. A few items to note:

- Coldman reported standardized mortality, different from others that reported RR
- Morrel reported 3 totals based on screening types (ever screened, not screened regularly, and screened regularly)

A1) All adherence to screen papers: Coldman, Morrel, Duffy, Choi

| Study or Subgroup | log[Risk Ratio] | SE | Weight | Risk Ratio IV, Random, 95% CI |
|---|---|---|---|---|
| Choi 2021 | -0.844 | 0.0117 | 28.3% | 0.43 [0.42, 0.44] |
| Coldman 2014 | -0.5108 | 0.0563 | 26.3% | 0.60 [0.54, 0.67] |
| Duffy 2021 | -0.5798 | 0.0681 | 25.4% | 0.56 [0.49, 0.64] |
| Morrell 2017 | -0.9676 | 0.1297 | 20.0% | 0.38 [0.29, 0.49] |
| **Total (95% CI)** | | | **100.0%** | **0.49 [0.40, 0.60]** |

Heterogeneity: Tau² = 0.04; Chi² = 47.95, df = 3 (P < 0.00001); I² = 94%
Test for overall effect: Z = 6.68 (P < 0.00001)

Risk Ratio IV, Random, 95% CI

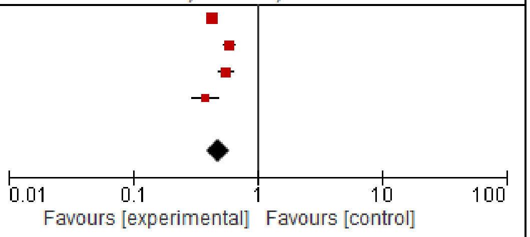

The cohort adherence to screen: Morrel reported BC mortality for ever screened cohort (control group is never screened).

A2) All adherence to screen papers: Coldman, Morrel, Duffy, Choi

| Study or Subgroup | log[Risk Ratio] | SE | Weight | Risk Ratio IV, Random, 95% CI |
|---|---|---|---|---|
| Choi 2021 | -0.844 | 0.0117 | 27.9% | 0.43 [0.42, 0.44] |
| Coldman 2014 | -0.5108 | 0.0563 | 25.8% | 0.60 [0.54, 0.67] |
| Duffy 2021 | -0.5798 | 0.0681 | 24.9% | 0.56 [0.49, 0.64] |
| Morrell 2017 | -0.8675 | 0.109 | 21.3% | 0.42 [0.34, 0.52] |
| **Total (95% CI)** | | | **100.0%** | **0.50 [0.41, 0.61]** |

Heterogeneity: Tau² = 0.04; Chi² = 46.89, df = 3 (P < 0.00001); I² = 94%
Test for overall effect: Z = 6.78 (P < 0.00001)

Risk Ratio IV, Random, 95% CI

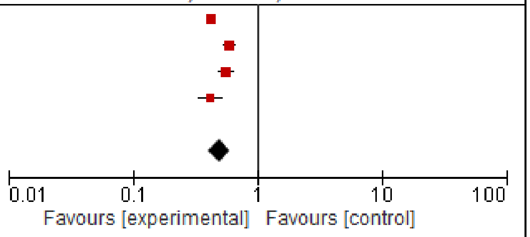

The cohort adherence to screen: Morrel reported BC mortality for not regularly screened cohort (control group is never screened).

A3) All adherence to screen papers: Coldman, Morrel, Duffy, Choi

| Study or Subgroup | log[Risk Ratio] | SE | Weight | Risk Ratio IV, Random, 95% CI |
|---|---|---|---|---|
| Choi 2021 | -0.844 | 0.0117 | 30.8% | 0.43 [0.42, 0.44] |
| Coldman 2014 | -0.5108 | 0.0563 | 28.8% | 0.60 [0.54, 0.67] |
| Duffy 2021 | -0.5798 | 0.0681 | 27.9% | 0.56 [0.49, 0.64] |
| Morrell 2017 | -1.1087 | 0.2513 | 12.6% | 0.33 [0.20, 0.54] |
| **Total (95% CI)** | | | **100.0%** | **0.49 [0.39, 0.62]** |

Heterogeneity: Tau² = 0.04; Chi² = 48.01, df = 3 (P < 0.00001); I² = 94%
Test for overall effect: Z = 6.11 (P < 0.00001)

Risk Ratio IV, Random, 95% CI

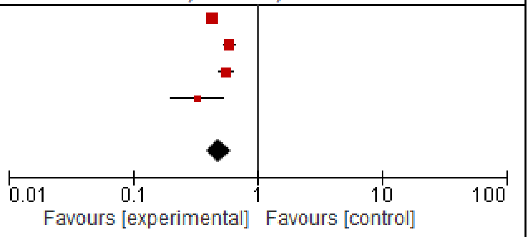

The cohort adherence to screen: Morrel reported BC mortality for regularly screened cohort (control group is never screened).

B) Low risk of bias papers: Coldman, Duffy, Choi

| Study or Subgroup | log[Risk Ratio] | SE | Weight | Risk Ratio IV, Random, 95% CI | Year |
|---|---|---|---|---|---|
| Coldman 2014 | -0.5108 | 0.0563 | 32.9% | 0.60 [0.54, 0.67] | 2014 |
| Choi 2021 | -0.844 | 0.0117 | 35.2% | 0.43 [0.42, 0.44] | 2021 |
| Duffy 2021 | -0.5798 | 0.0681 | 31.9% | 0.56 [0.49, 0.64] | 2021 |
| **Total (95% CI)** | | | **100.0%** | **0.52 [0.41, 0.67]** | |

Heterogeneity: Tau² = 0.04; Chi² = 46.72, df = 2 (P < 0.00001); I² = 96%
Test for overall effect: Z = 5.20 (P < 0.00001)

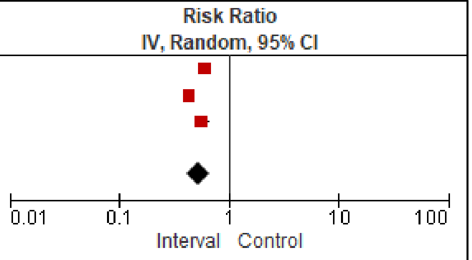

C) Low risk of bias papers: Duffy, Choi (presenting relative risks only)

| Study or Subgroup | log[Risk Ratio] | SE | Weight | Risk Ratio IV, Random, 95% CI | Year |
|---|---|---|---|---|---|
| Choi 2021 | -0.844 | 0.0117 | 53.2% | 0.43 [0.42, 0.44] | 2021 |
| Duffy 2021 | -0.5798 | 0.0681 | 46.8% | 0.56 [0.49, 0.64] | 2021 |
| **Total (95% CI)** | | | **100.0%** | **0.49 [0.38, 0.63]** | |

Heterogeneity: Tau² = 0.03; Chi² = 14.62, df = 1 (P = 0.0001); I² = 93%
Test for overall effect: Z = 5.46 (P < 0.00001)

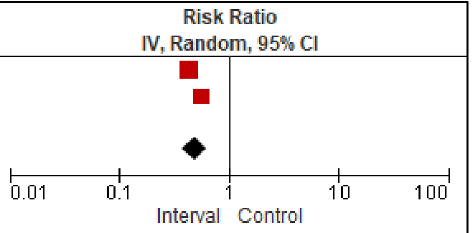

#### Appendices References

1. Coldman A, Phillips N, Wilson C, Decker K, Chiarelli AM, Brisson J, et al. Pan-Canadian study of mammography screening and mortality from breast cancer. 2014;106(11).

2. Engmann NJ, Golmakani MK, Miglioretti DL, Sprague BL, Kerlikowske K. Population-Attributable Risk Proportion of Clinical Risk Factors for Breast Cancer. JAMA Oncol. 2017 Sep 1;3(9):1228–36.

3. Chiu SYH, Duffy S, Yen AMF, Tabár L, Smith RA, Chen HH. Effect of Baseline Breast Density on Breast Cancer Incidence, Stage, Mortality, and Screening Parameters: 25-Year Follow-up of a Swedish Mammographic Screening. Cancer Epidemiology, Biomarkers & Prevention. 2010 May 1;19(5):1219–28.

4. Klarenbach S, Sims-Jones N, Lewin G, Singh H, Thériault G, Tonelli M, et al. Recommendations on screening for breast cancer in women aged 40–74 years who are not at increased risk for breast cancer. CMAJ. 2018 Dec 10;190(49):E1441–51.

5. Nyström L, Bjurstam N, Jonsson H, Zackrisson S, Frisell J. Reduced breast cancer mortality after 20+ years of follow-up in the Swedish randomized controlled mammography trials in Malmö, Stockholm, and Göteborg. J Med Screen. 2017 Mar;24(1):34–42.

6. Moss SM, Wale C, Smith R, Evans A, Cuckle H, Duffy SW. Effect of mammographic screening from age 40 years on breast cancer mortality in the UK Age trial at 17 years' follow-up: a randomised controlled trial. The Lancet Oncology. 2015;16(9):1123–32.

7. Tabár L, Vitak B, Chen THH, Yen AMF, Cohen A, Tot T, et al. Swedish Two-County Trial: Impact of Mammographic Screening on Breast Cancer Mortality during 3 Decades. Radiology. 2011 Sep;260(3):658–63.

8. Nyström L, Andersson I, Bjurstam N, Frisell J, Nordenskjöld B, Rutqvist LE. Long-term effects of mammography screening: updated overview of the Swedish randomised trials. The Lancet. 2002 Mar;359(9310):909–19.

9. Miller AB, Wall C, Baines CJ, Sun P, To T, Narod SA. Twenty five year follow-up for breast cancer incidence and mortality of the Canadian National Breast Screening Study: randomised screening trial. BMJ. 2014 Feb 11;348(feb11 9):g366–g366.

10. Shapiro S, Venet W, Strax P. Current results of the breast cancer screening randomized trial: the health insurance plan (HIP) of greater New York study. Hans Huber. 1988;3–15.

11. Egger M, Davey Smith G, Schneider M, Minder C. Bias in meta-analysis detected by a simple, graphical test. BMJ. 1997 Sep;315(7109):629–34.

12. Bjurstam N, Björneld L, Warwick J, Sala E, Duffy SW, Nyström L, et al. The Gothenburg Breast Screening Trial: The Gothenburg Breast Screening Trial. Cancer. 2003 May 15;97(10):2387–96.

13. Tabar L, Fagerberg G, Chen H, Duffy S, Smart C, Gad A. Efficacy of breast cancer screening by age. Cancer. 1995;75(10):2507–17.

14. Habbema J, Oortmarssen G, Putten D van, Lubbe J, Maas P. Age-specific reduction in breast cancer mortality by screening: an analysis of the results of the Health Insurance Plan of Greater New York study. JNCI: Journal of the National Cancer Institute. 1986;77(2):317–20.

15. Choi E, Jun JK, Suh M, Jung KW, Park B, Lee K, et al. Effectiveness of the Korean National Cancer Screening Program in reducing breast cancer mortality. NPJ breast cancer. 2021;7(1):83.

16. Coldman A, Phillips N, Wilson C, Decker K, Chiarelli AM, Brisson J, et al. Pan-Canadian Study of Mammography Screening and Mortality from Breast Cancer. JNCI: Journal of the National Cancer Institute [Internet]. 2014 Nov [cited 2023 Sep 11];106(11). Available from: https://academic.oup.com/jnci/article-lookup/doi/10.1093/jnci/dju261

17. Duffy SW, Tabar L, Yen AMF, Dean PB, Smith RA, Jonsson H, et al. Beneficial Effect of Consecutive Screening Mammography Examinations on Mortality from Breast Cancer: A Prospective Study. Radiology. 2021;299(3):541–7.

18. Morrell S, Taylor R, Roder D, Robson B, Gregory M, Craig K. Mammography service screening and breast cancer mortality in New Zealand: a National Cohort Study 1999–2011. Br J Cancer. 2017 Mar;116(6):828–39.

19. Garcia-Albeniz X, Hernan MA, Logan RW, Price M, Armstrong K, Hsu J. Continuation of Annual Screening Mammography and Breast Cancer Mortality in Women Older Than 70 Years. Annals of internal medicine. 2020;172(6):381–9.

20. Paap E, Verbeek ALM, Botterweck AAM, van Doorne-Nagtegaal HJ, Imhof-Tas M, e Koning HJ, et al. Breast cancer screening halves the risk of breast cancer death: a case-referent study. Breast (Edinburgh, Scotland). 2014;23(4):439–44.

21. Pocobelli G, Weiss NS. Breast cancer mortality in relation to receipt of screening mammography: a case–control study in Saskatchewan, Canada. Cancer Causes Control. 2015 Feb;26(2):231–7.

22. Massat NJ, Dibden A, Parmar D, Cuzick J, Sasieni PD, Duffy SW. Impact of Screening on Breast Cancer Mortality: The UK Program 20 Years On. Cancer epidemiology, biomarkers & prevention : a publication of the American Association for Cancer Research, cosponsored by the American Society of Preventive Oncology. 2016;25(3):455–62.

23. Ripping TM, van der Waal D, Verbeek ALM, Broeders MJM. The relative effect of mammographic screening on breast cancer mortality by socioeconomic status. Medicine. 2016 Aug;95(31):e4335.

24. van der Waal D, Ripping TM, Verbeek ALM, Broeders MJM. Breast cancer screening effect across breast density strata: A case-control study: Screening effect across breast density strata. Int J Cancer. 2017 Jan 1;140(1):41–9.

25. Maroni R, Massat NJ, Parmar D, Dibden A, Cuzick J, Sasieni PD, et al. A case-control study to evaluate the impact of the breast screening programme on mortality in England. Br J Cancer. 2021 Feb 16;124(4):736–43.

26. De Troeyer K, Silversmit G, Rosskamp M, Truyen I, Van Herck K, Goossens MM, et al. The effect of the Flemish breast cancer screening program on breast cancer-specific mortality: A case-referent study. Cancer epidemiology. 2023;82:102320.

27. Katalinic A, Eisemann N, Kraywinkel K, Noftz MR, Hübner J. Breast cancer incidence and mortality before and after implementation of the German mammography screening program. Intl Journal of Cancer. 2020 Aug;147(3):709–18.

28. Parvinen I, Heinavaara S, Anttila A, Helenius H, Klemi P, Pylkkanen L. Mammography screening in three Finnish residential areas: Comprehensive population-based study of breast cancer incidence and incidence-based mortality 1976-2009. British Journal of Cancer. 2015;112(5):918–24.

29. Tabár L, Dean PB, Chen TH, Yen AM, Chen SL, Fann JC, et al. The incidence of fatal breast cancer measures the increased effectiveness of therapy in women participating in mammography screening. Cancer. 2019 Feb 15;125(4):515–23.

30. Wilkinson AN, Ellison LF, Billette JM, Seely JM. Impact of Breast Cancer Screening on 10-Year Net Survival in Canadian Women Age 40-49 Years. J Clin Oncol. 2023 Oct 10;41(29):4669–77.

31. Tabar L, Fagerberg G, Duffy SW, Day NE. The Swedish two county trial of mammographic screening for breast cancer: recent results and calculation of benefit. J Epidemiol Community Health. 1989 Jun;43(2):107–14.

32. Frisell J, Lidbrink E, Hellström L, Rutqvist LE. Followup after 11 years – update of mortality results in the Stockholm mammographic screening trial. Breast Cancer Res Treat. 1997 Sep;45(3):263–70.

33. Miller AB, To T, Baines CJ, Wall C. The Canadian National Breast Screening Study-1: Breast Cancer Mortality after 11 to 16 Years of Follow-up: A Randomized Screening Trial of Mammography in Women Age 40 to 49 Years. Annals of Internal Medicine. 2002;137(5):305–12.

34. Miller AB, To T, Baines CJ, Wall C, For the Canadian National Breast Screening Study-2. Canadian National Breast Screening Study-2: 13-Year Results of a Randomized Trial in Women Aged 50–59 Years. JNCI: Journal of the National Cancer Institute. 2000 Sep 20;92(18):1490–9.

35. Bjurstam N, Björneld L, Duffy SW, Smith TC, Cahlin E, Eriksson O, et al. The Gothenburg breast screening trial: First results on mortality, incidence, and mode of detection for women ages 39-49 years at randomization. Cancer. 1997 Dec;80(11):2091–9.

36. Tarone RE. The excess of patients with advanced breast cancer in young women screened with mammography in the Canadian National Breast Screening Study. Cancer. 1995;75(4):997–1003.

37. Puliti D, Bucchi L, Mancini S, Paci E, Baracco S, Campari C, et al. Advanced breast cancer rates in the epoch of service screening: The 400,000 women cohort study from Italy. European Journal of Cancer. 2017 Apr;75:109–16.

38. de Glas NA, e Craen AJM, Bastiaannet E, Op 't Land EG, Kiderlen M, van de Water W, et al. Effect of implementation of the mass breast cancer screening programme in older women in the Netherlands: population based study. BMJ (Clinical research ed). 2014;349:g5410.

39. Helvie MA, Chang JT, Hendrick RE, Banerjee M. Reduction in late-stage breast cancer incidence in the mammography era: Implications for overdiagnosis of invasive cancer. Cancer. 2014;120(17):2649–56.

40. Wilkinson AN, Billette JM, Ellison LF, Killip MA, Islam N, Seely JM. The Impact of Organised Screening Programs on Breast Cancer Stage at Diagnosis for Canadian Women Aged 40–49 and 50–59. Current Oncology. 2022;29(8):5627–43.

41. Zackrisson S, Andersson I, Janzon L, Manjer J, Garne JP. Rate of over-diagnosis of breast cancer 15 years after end of Malmö mammographic screening trial: follow-up study. BMJ. 2006 Mar 25;332(7543):689–92.

42. Baines CJ, To T, Miller AB. Revised estimates of overdiagnosis from the Canadian National Breast Screening Study. Preventive Medicine. 2016 Sep;90:66–71.

43. Duffy SW, Vulkan D, Cuckle H, Parmar D, Sheikh S, Smith RA, et al. Effect of mammographic screening from age 40 years on breast cancer mortality (UK Age trial): final results of a randomised, controlled trial. The Lancet Oncology. 2020 Sep;21(9):1165–72.

44. Lund E, Nakamura A, Thalabard JC. No overdiagnosis in the Norwegian Breast Cancer Screening Program estimated by combining record linkage and questionnaire information in the Norwegian Women and Cancer study. European Journal of Cancer. 2018 Jan;89:102–12.

45. Richman IB, Long JB, Soulos PR, Wang SY, Gross CP. Estimating Breast Cancer Overdiagnosis After Screening Mammography Among Older Women in the United States. Ann Intern Med. 2023 Sep 19;176(9):1172–80.

46. Bjurstam NG, Björneld LM, Duffy SW. Updated results of the Gothenburg Trial of Mammographic Screening. Cancer. 2016 Jun 15;122(12):1832–5.

47. Gøtzsche PC, Jørgensen KJ. Screening for breast cancer with mammography. Cochrane Database Syst Rev. 2013 Jun 4;2013(6):CD001877.

48. Canadian Partnership Against Cancer (CPAC). Breast Cancer Screening in Canada: Monitoring and Evaluation of Quality Indicators - Results Report, January 2011 to December 2012. Toronto, ON: Canadian Partnership Against Cancer; 2016.

49. Nova Scotia Breast Screening Program. Imaging Guidelines [Internet]. Nova Scotia Breast Cancer Screening Program. Available from: https://nsbreastscreening.ca/program-information/imaging-guidelines

50. Health PEI. PEI Breast Screening Program [Internet]. Government of Prince Edward Island. 2023. Available from: https://www.princeedwardisland.ca/en/information/health-pei/pei-breast-screening-program

51. Brenner DR, Gillis J, Demers AA, Ellison LF, Billette JM, Zhang SX, et al. Projected estimates of cancer in Canada in 2024. CMAJ. 2024;196(18):E615–23.

52. Oeffinger KC, Fontham ET, Etzioni R, Herzig A, Michaelson JS, Shih YCT, et al. Breast cancer screening for women at average risk: 2015 guideline update from the American Cancer Society. Jama. 2015;314(15):1599–614.

53. Duffy S, Vulkan D, Cuckle H, Parmar D, Sheikh S, Smith R, et al. Annual mammographic screening to reduce breast cancer mortality in women from age 40 years: long-term follow-up of the UK Age RCT. Health Technol Assess. 2020 Oct;24(55):1–24.

54. Dunn N, Youl P, Moore J, Harden H, Walpole E, Evans E, et al. Breast-cancer mortality in screened versus unscreened women: Long-term results from a population-based study in Queensland, Australia. Journal of medical screening. 2021;28(2):193–9.

55. Weedon-Fekjaer H, Romundstad PR, Vatten LJ. Modern mammography screening and breast cancer mortality: population study. BMJ (Clinical research ed). 2014;348:g3701.

56. Blyuss O, Dibden A, Massat NJ, Parmar D, Cuzick J, Duffy SW, et al. A case-control study to evaluate the impact of the breast screening programme on breast cancer incidence in England. Cancer medicine. 2023;12(2):1878–87.

57. Beckmann KR, Lynch JW, Hiller JE, Farshid G, Houssami N, Duffy SW, et al. A novel case-control design to estimate the extent of over-diagnosis of breast cancer due to organised population-based mammography screening. International journal of cancer. 2015;136(6):1411–21.

58. Katalinic A, Eisemann N, Kraywinkel K, Noftz MR, Hubner J. Breast cancer incidence and mortality before and after implementation of the German mammography screening program. International journal of cancer. 2020;147(3):709–18.
